# Metabolic signature of the ganglion cell–inner plexiform layer thickness and the risks of mortality and morbidity: a population-based study in UK Biobank

**DOI:** 10.1101/2022.09.26.22280334

**Authors:** Shaopeng Yang, Yixiong Yuan, Yanping Chen, Shiran Zhang, Yujie Wang, Xianwen Shang, Gabriella Bulloch, Huan Liao, Yifan Chen, Lei Zhang, Zhuoting Zhu, Mingguang He, Wei Wang

**Affiliations:** State Key Laboratory of Ophthalmology, Zhongshan Ophthalmic Center, Sun Yat-sen University, Guangdong Provincial Key Laboratory of Ophthalmology and Visual Science, Guangdong Provincial Clinical Research Center for Ocular Diseases, Guangzhou, China; Department of Ophthalmology, Shanghai General Hospital, Shanghai Jiao Tong University School of Medicine, Shanghai, China; Centre for Eye Research Australia, Royal Victorian Eye and Ear Hospital, Melbourne, Australia; Department of Ophthalmology, Guangdong Academy of Medical Sciences, Guangdong Provincial People’s Hospital, Guangzhou, China; Epigenetics and Neural Plasticity Laboratory, Florey Institute of Neuroscience and Mental Health, University of Melbourne; John Radcliffe Hospital, Oxford University Hospitals NHS Foundation Trust, Oxford, UK

**Keywords:** retina, GCIPL, metabolomics, mortality, type 2 diabetes, obstructive sleep apnea/hypopnea syndrome, cardiovascular disease, dementia, ganglion cell–inner plexiform layer thickness, myocardial infarction, heart failure, stroke

## Abstract

**Background:** The retina is considered a unique window to systemic health, but their biological link remains unknown.

**Methods:** A total of 93,838 UK Biobank participants with metabolomics data were included in the study. Plasma metabolites associated with GCIPLT were identified in 7,824 participants who also underwent retinal optical coherence tomography; prospective associations of GCIPLT-associated metabolites with 12-year risk of mortality and major age-related diseases were assessed in 86,014 participants. The primary outcomes included all- and specific-cause mortality. The secondary outcomes included incident type 2 diabetes mellitus (T2DM), obstructive sleep apnea/hypopnea syndrome (OSAHS), myocardial infarction (MI), heart failure, ischemic stroke, and dementia. C-statistics and net reclassification indexes (NRIs) were calculated to evaluate the added predictive value of GCIPLT metabolites. Calibration was assessed using calibration plots.

**Findings:** Sixteen metabolomic signatures were associated with GCIPLT (P< 0.009 [Bonferroni-corrected threshold]), and most were associated with the future risk of mortality and age-related diseases. The constructed meta-GCIPLT scores distinguished well between patients with high and low risks of mortality and morbidity, showing predictive values higher than or comparable to those of traditional risk factors (C-statistics: 0.780[0.771-0.788], T2DM; 0.725[0.707-0.743], OSAHS; 0.711[0.695-0.726], MI; 0.685[0.662-0.707], cardiovascular mortality; 0.657[0.640-0.674], heart failure; 0.638[0.636-0.660], other mortality; 0.630[0.618-0.642], all-cause mortality; 0.620[0.598-0.643], dementia; 0.614[0.593-0.634], stroke; and 0.601[0.585-0.617], cancer mortality). The NRIs confirmed the inclusion of GCIPLT metabolomic signatures to the models based on traditional risk factors resulted in significant improvements in model performance (5.18%, T2DM [P=3.86E-11]; 4.43%, dementia [P=0.003]; 4.20%, cardiovascular mortality [P=6.04E-04]; 3.73%, MI [P=1.72E-07]; 2.93%, OSAHS [P=3.13E-05]; 2.39%, all-cause mortality [P=3.89E-05]; 2.33%, stroke [P=0.049]; 2.09%, cancer mortality [P=0.039]; and 1.59%, heart failure [P=2.72E-083.07E-04]). Calibration plots showed excellent calibration between predicted risk and actual incidence in the new models.

**Interpretation:** GCIPLT-associated plasma metabolites captured the residual risk for mortality and major systemic diseases not quantified by traditional risk factors in the general population. Incorporating GCIPLT metabolomic signatures into prediction models may assist in screening for future risks of these health outcomes.

**Funding:** National Natural Science Foundation (China).

**Research in context:** *Evidence before this study:* Recent studies have recognized that retinal measurements can indicate an accelerated risk of aging and multiple systemic diseases preceding clinical symptoms and signs. Despite these insights, it remains unknown how retinal alterations are biologically linked to systemic health.

*Added value of this study:* Using the UK Biobank, we identified ganglion cell–inner plexiform layer thickness (GCIPLT) metabolomic signatures, and revealed their association with the risk of all- and specific-cause mortality and six age related diseases: type 2 diabetes, dementia, stroke, myocardial infarction, heart failure, and obstructive sleep apnea/hypopnea syndrome. The meta-GCIPLT score significantly improved the discriminative power of the predictive models for theses health outcomes based on conventional risk factors.

*Implications of all the available evidence:* GCIPLT-associated plasma metabolites have the potential to capture the residual risk of systemic diseases and mortality not quantified by traditional risk factors. Incorporating GCIPLT metabolomic signatures into prediction models may assist in screening for future risks of these health outcomes. Since metabolism is a modifiable risk factor that can be treated medically, the future holds promise for the development of new strategies that reverse or interrupt the onset of these diseases by modifying metabolic factors.

## Introduction

The retina is an extension of the central nervous system and a unique window to systemic health since its microstructures can be non-invasively imaged.^1-3^ Optical coherence tomography (OCT) of the retina has identified retinal nerve fiber layers (RNFL) and ganglion cell–inner plexiform layer (GCIPL) as biomarkers of aging and related diseases preceding clinical symptoms and signs.^4-13^ With the booming OCT deployment in primary care settings and people’s concerns for eye health, the easily accessible, risk-free, and high-resolution retinal scans are becoming an attractive alternative for screening systemic health in routine community scenarios.^14-17^

The biological link between alterations in retinal layers and systemic health remains unknown. Metabolomics offers a novel opportunity to study the biological signatures behind these complex features,^18^ especially considering that metabolic risk factors contribute substantially to various age-related diseases.^19-21^ Conversely, previous studies have reported associations between circulating metabolites and alterations in retinal layers,^22-24^ but these are limited to single-biomarker approaches. Additionally, these studies majorly focused on the RNFL (representing axons of retinal ganglion cells [RGCs]), while studies analyzing metabolic factors with GCIPL (representing cytosol and dendrites of RGCs) are rare, despite growing evidence suggesting that GCIPL is more sensitive and a reproducible mirror representing retinal damage and systemic diseases.^6, 25-27^

We hypothesized that metabolites may underlie the links between retinal layer changes and systemic health. Since RGCs are extremely susceptible to systemic injury,^27^ studying GCIPL-mediated biological changes *in vivo* may predict the future risk and course of systemic diseases earlier in their pathogenesis. Therefore, the objective of this study was to investigate the association between the GCIPLT metabolomic signature and the risk of mortality and major age-related diseases in a large population-based cohort.

## Methods

### Study population

This study utilized participants from the UK Biobank, a large population-based cohort study including over half a million participants aged 40–69 years from England, Scotland, and Wales registered with the National Health Service (NHS). The study design is described previously.^28^ This study was conducted in accordance with the principles of the Declaration of Helsinki and was approved by the North West Multi-center Research Ethics Committee. All the participants signed an informed consent form.

The overall design of this study is shown in **Figure 1** and consists of two phases, in which the UK biobank was divided into three parts: with both metabolomic and OCT data (population I), with metabolomic data only (population II), and without metabolomic data (population III). Population I was included in the phase I analysis; 7,824 participants who underwent a qualifying macular OCT scan and had available nuclear magnetic resonance (NMR) metabolomic data were included after stringent exclusion criteria. A total of 110,730 participants who underwent NMR metabolomics were included in phase II analysis. After excluding participants with missing metabolomic data (population III), missing hospitalization records (n = 18,032), and data used in phase I analysis (n = 6,684), 86,014 participants were finally included in phase II analysis. The detailed inclusion and exclusion criteria for this study are shown in **Supplementary Figure S1**.

**Figure 1.**
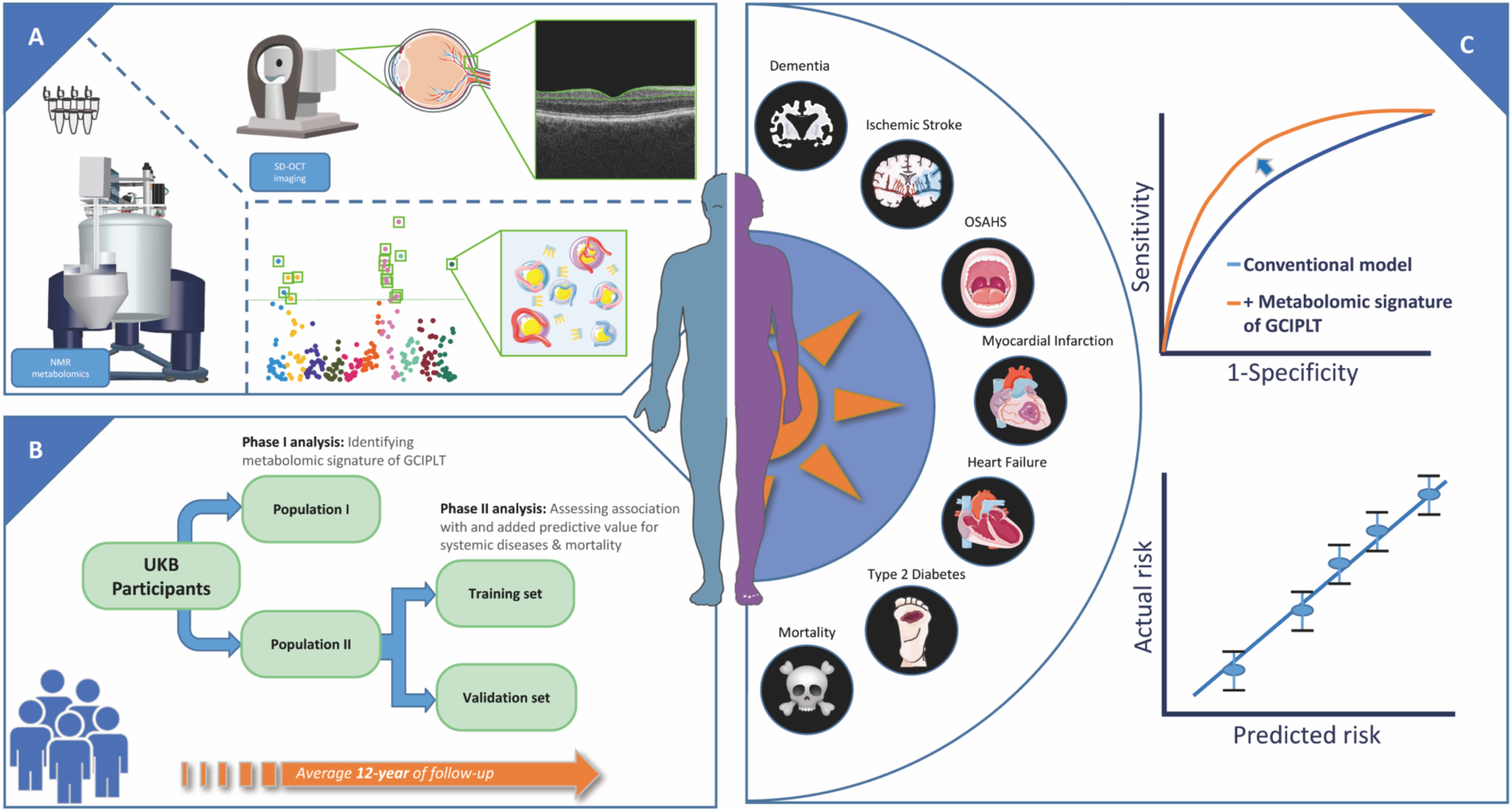
Schematic diagram summarizing the study design. (A) demonstrates the quantification of metabolic biomarkers and macula GCIPLT. (B) summarizes the study design and population. (C) shows the study endpoints. SD-OCT =spectral-domain optical coherence tomography; NMR =nuclear magnetic resonance; GCIPLT =ganglion cell-inner plexiform layer thickness; OSAHS =obstructive sleep apnea/hypopnea syndrome.

**Figure 2.**
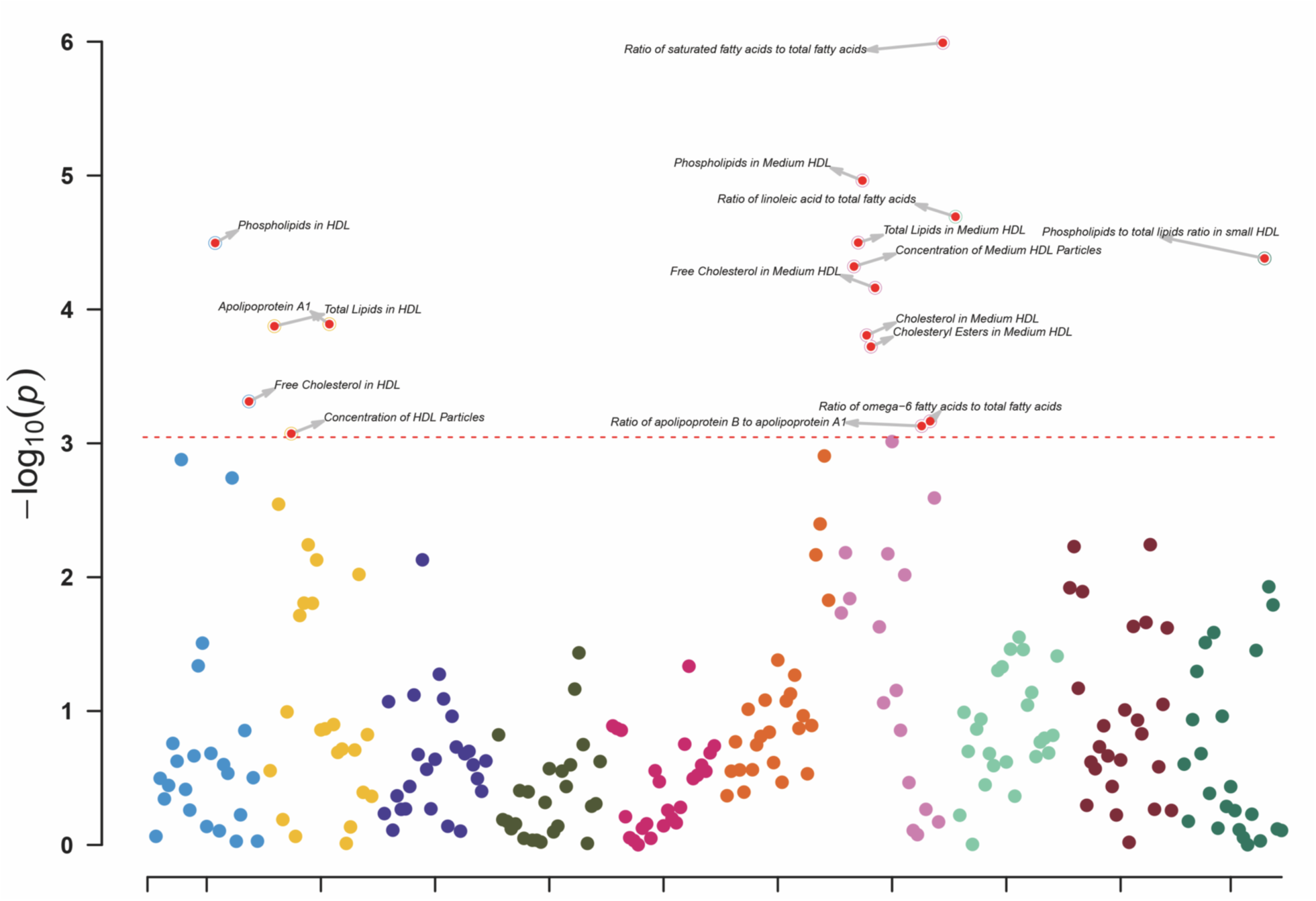
Metabolic metrics that reached the Bonferroni-corrected significance threshold for multiple comparisons in phase I analysis. GCIPLT =ganglion cell-inner plexiform layer thickness; HDL =high-density lipoprotein.

### Participants’ demographic, systemic, and ocular characteristics

At baseline (2006–2010), physical measurements, face-to-face interviews, and detailed self-administered touchscreen questionnaires were conducted on all participants. The questionnaires included demographic and socioeconomic factors (age, sex, race, education level, Townsend deprivation index, and income), lifestyle factors (smoking and drinking status), family history, and medical history, including the use of insulin, lipid-lowering medications, and antihypertensive drugs. Baseline diseases were defined using questionnaires, interviews, and inpatient data based on the ICD-10 codes. Physical measurements including baseline body mass index (BMI), waist-to-hip ratio (WHR), blood pressure, visual acuity, refractive error, spherical equivalent, intraocular pressure (IOP), total cholesterol, low-density lipoprotein cholesterol (LDL-c), and high-density lipoprotein cholesterol (HDL-c) levels, glycosylated hemoglobin A1c, urine microalbumin and creatinine levels were obtained. (**Supplementary Methods**) The presence of apolipoprotein E (apoE) ε4 allele was defined using the apoE ε4+ dominant model of ε3/ε4 and ε4/ε4. The field codes are listed in **Supplementary Table S1**.

### Proton nuclear magnetic resonance metabolomics

A total of 249 metabolic metrics were quantified in plasma samples from participants using high-throughput NMR platform (Nightingale Health, Finland). Details about the metabolic-profiling protocol are described elsewhere,^29-31^ but in brief, cryopreserved plasma samples were thawed and centrifuged, and the supernatant was mixed with phosphate buffer. The samples were then loaded onto a cooled sample changer, and two NMR spectra of each plasma sample were recorded using a 500 MHz NMR spectrometer (Bruker AVANCE IIIHD). One spectrum characterized resonances produced mainly by proteins and lipid lipoprotein particles, whereas the other detected low-molecular-weight metabolites. After quality control, the metabolic metrics were quantified using the Nightingale Health biomarker quantification library 2020, including 168 metrics presented at absolute levels and 81 metrics presented as ratio values. For the lipoprotein subgroups, lipid concentrations and compositions were measured based on the triglyceride (TG), phospholipid, total cholesterol, cholesteryl ester, free cholesterol, and total lipid concentrations in each subclass. Metabolic indicators were measured in absolute concentration units (mmol/L) or ratios.

### Spectral-domain optical coherence tomography imaging

Spectral-domain OCT was performed in a closed darkroom using a Topcon 3D OCT-1000 Mk II (Topcon, Inc., Oakland, NJ, USA). The system had an axial resolution of 6 μm and an image acquisition rate of 18,000 A-scans/s. Using a three-dimensional 6 × 6 mm macular volume scan mode, the retina was imaged at a scan density of 512 A-scans × 128 B-scans in 3.6 s. The Topcon Advanced Boundary Segmentation algorithm (version 1.6.1.1) automatically segmented the retinal layers and determined the macular GCIPL thickness. The image quality score, internal limiting membrane indicator, validity count, and motion indicators were used to detect and ensure quality control, whereby images with low signal strength (Q<45) or poor segmentation or centration (poorest 20% of each indicator) were excluded. If both eyes were eligible for the analysis, one eye was randomly selected for further analysis.

### Mortality and morbidity outcomes

The Hospital Episode Statistics database, Scottish Morbidity Record, and Patient Episode Database were used to record inpatient hospital records for England, Scotland, and Wales. Mortality data were obtained from national datasets with the NHS Digital (England and Wales) and NHS Central Register (Scotland), and the primary cause of mortality was recorded using the ICD-10 (**Supplementary Table S1**). The follow-up period was from March 16, 2006, to March 31, 2021. Person-days for each participant were calculated from the date of baseline assessment to the date of disease onset, mortality, or the end of follow-up, whichever came first.

### Statistical analysis

R software (version 4.1.2) was used for all data analyses and for the presentation of the results. Student’s t-test and the chi-square test were used to compare continuous and categorical variables, respectively. The z-score was first standardized for all metabolite measurements to ensure comparability across metabolites.

In the phase I analysis, the association between 249 metabolite measures and macular GCIPLT was assessed using a multifactorial linear regression model after adjusting for age, sex, race, education, Townsend deprivation coefficient, household income, BMI, smoking status, alcohol consumption status, use of lipid-lowering medications, spherical equivalent, and IOP. After performing principal component analysis on 249 circulating metabolites, P values for multiple comparisons were set using the Bonferroni method to reduce the false-positive rate.

In phase II analysis, participants were randomly categorized by 1:1 ratio into training and validation sets. Participants diagnosed with the corresponding disease at baseline were excluded from the corresponding analysis (e.g., in T2DM analysis, participants diagnosed with T2DM at baseline were excluded). The metabolites that reached significant levels in the phase I analysis were analyzed using multifactorial Cox regression for the risk of six age-related diseases (T2DM, obstructive sleep apnea/hypopnea syndrome [OSAHS], myocardial infarction [MI], heart failure, ischemic stroke, and dementia) and four mortality types (all-cause, cardiovascular, cancer, and other mortality), adjusting for covariates similar to those in the phase I analysis, excluding spherical equivalents and IOP.

Subsequently, the metabolic risk scores were calculated for each disease. First, all metabolites that reached significance in the phase I analysis were selected as candidate variables. Metabolic markers included in the calculation were screened based on Bayesian information criteria using a forward– backward method. The metabolic score was calculated using the following formula: 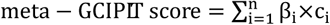 where β_i_ represents the coefficient of the i^th^ metabolite [ln (hazar ratio)] and c_i_ represents the concentration of the i^th^ metabolite (after z-score normalization). The participants were divided into the top 10% and bottom 10% groups based on their metabolic GCIPLT scores, and Kaplan–Meier survival analysis was used to compare the risk of morbidity and mortality of participants within the top 10% metabolic scores with those in the bottom 10%. The predictive value of the metabolic scores for the occurrence of these systemic diseases and mortality types was compared with those of the traditional risk factors. Next, the metabolic GCIPLT score was analyzed for additional predictive value compared with the conventional model. NRIs were calculated to quantify the net benefit of adding the metabolic score to the classical model. Finally, calibration plots were created to compare the predicted and actual risk.

## Results

### Characteristics of the study population

A total of 3,913 right eyes and 3,911 left eyes of 7,824 participants (population I) were eligible for phase I analysis (**Supplementary Figure S1**). For the phase II analysis, 86,014 participants were eligible (population II). Participants who underwent OCT measurements were younger (P<0.001), male (P=0.002), more educated (P<0.001), had a higher income (P<0.001), had a lower BMI (P=0.002), smoked less (P=0.029), and were less likely to be on lipid-lowering (P=0.004) or antihypertensive medications (P<0.001) than those who did not. The distributions of participant characteristics in the training and validation sets were similar (all P>0.05). The baseline characteristics of the study population are summarized in **Table 1**.

**Table 1.** Baseline characteristics of the study population.

| Characteristic | Total | Population I | Population II |  | P value † § | P value ‡ § |
| --- | --- | --- | --- | --- | --- | --- |
|  |  |  | Training set | Validation set |  |  |
| Number of subjects | 93838 | 7824 | 43007 | 43007 | - | - |
| Age at recruitment |  |  |  |  |  |  |
| ≤49 | 21438 (22.8) | 2248 (28.7) | 9626 (22.4) | 9564 (22.2) | **<0.001** | 0.776 |
| 50-54 | 13843 (14.8) | 1191 (15.2) | 6334 (14.7) | 6318 (14.7) |  |  |
| 55-59 | 16856 (18.0) | 1319 (16.9) | 7694 (17.9) | 7843 (18.2) |  |  |
| 60-64 | 23246 (24.8) | 1800 (23.0) | 10742 (25.0) | 10704 (24.9) |  |  |
| ≥65 | 18455 (19.7) | 1266 (16.2) | 8611 (20.0) | 8578 (19.9) |  |  |
| Gender |  |  |  |  |  |  |
| Female | 51182 (54.5) | 4122 (52.7) | 23596 (54.9) | 23464 (54.6) | **0.002** | 0.370 |
| Male | 42656 (45.5) | 3702 (47.3) | 19411 (45.1) | 19543 (45.4) |  |  |
| Race |  |  |  |  |  |  |
| White | 88754 (94.6) | 7188 (91.9) | 40771 (94.8) | 40795 (94.9) | **<0.001** | 0.900 |
| Others | 4652 (5.0) | 589 (7.5) | 2045 (4.8) | 2018 (4.7) |  |  |
| Missing | 432 (0.5) | 47 (0.6) | 191 (0.4) | 194 (0.5) |  |  |
| Townsend Deprivation Index |  |  |  |  |  |  |
| Quantile 1 | 23598 (25.1) | 1749 (22.4) | 10871 (25.3) | 10978 (25.5) | **<0.001** | 0.567 |
| Quantile 2 | 23394 (24.9) | 1861 (23.8) | 10713 (24.9) | 10820 (25.2) |  |  |
| Quantile 3 | 23324 (24.9) | 2145 (27.4) | 10689 (24.9) | 10490 (24.4) |  |  |
| Quantile 4 | 23397 (24.9) | 2059 (26.3) | 10676 (24.8) | 10662 (24.8) |  |  |
| Missing | 125 (0.1) | 10 (0.1) | 58 (0.1) | 57 (0.1) |  |  |
| Average total household income before tax (£) |  |  |  |  |  |  |
| < 18k | 18820 (20.1) | 1214 (15.5) | 8742 (20.3) | 8864 (20.6) | **<0.001** | 0.714 |
| 18k~30k | 20770 (22.1) | 1627 (20.8) | 9593 (22.3) | 9550 (22.2) |  |  |
| 31k~51k | 20594 (21.9) | 1846 (23.6) | 9425 (21.9) | 9323 (21.7) |  |  |
| 52k~100k | 15728 (16.8) | 1610 (20.6) | 7058 (16.4) | 7060 (16.4) |  |  |
| > 100k | 3982 (4.2) | 516 (6.6) | 1700 (4.0) | 1766 (4.1) |  |  |
| Missing | 13944 (14.9) | 1011 (12.9) | 6489 (15.1) | 6444 (15.0) |  |  |
| Education achievement |  |  |  |  |  |  |
| Level O | 32411 (34.5) | 2316 (29.6) | 14983 (34.8) | 15112 (35.1) | **<0.001** | 0.517 |
| Level A | 4934 (5.3) | 499 (6.4) | 2186 (5.1) | 2249 (5.2) |  |  |
| University | 55406 (59.0) | 5009 (64.0) | 25287 (58.8) | 25110 (58.4) |  |  |
| Missing | 1087 (1.2) | 0 (0.0) | 551 (1.3) | 536 (1.2) |  |  |
| Body mass index, kg/m<sup>2</sup> |  |  |  |  |  |  |
| Normal | 30120 (32.1) | 2650 (33.9) | 13751 (32.0) | 13719 (31.9) | **0.002** | 0.518 |
| Overweight | 39832 (42.4) | 3311 (42.3) | 18290 (42.5) | 18231 (42.4) |  |  |
| Obesity | 23514 (25.1) | 1839 (23.5) | 10805 (25.1) | 10870 (25.3) |  |  |
| Missing | 372 (0.4) | 24 (0.3) | 161 (0.4) | 187 (0.4) |  |  |
| Smoking |  |  |  |  |  |  |
| Never | 32963 (35.1) | 2711 (34.6) | 15085 (35.1) | 15167 (35.3) | **0.029** | 0.793 |
| Ever/Current | 10110 (10.8) | 771 (9.9) | 4661 (10.8) | 4678 (10.9) |  |  |
| Missing | 50765 (54.1) | 4342 (55.5) | 23261 (54.1) | 23162 (53.9) |  |  |
| Drinking |  |  |  |  |  |  |
| Never | 3510 (3.7) | 261 (3.3) | 1615 (3.8) | 1634 (3.8) | 0.077 | 0.846 |
| Ever/Current | 85974 (91.6) | 7235 (92.5) | 39393 (91.6) | 39346 (91.5) |  |  |
| Missing | 4354 (4.6) | 328 (4.2) | 1999 (4.6) | 2027 (4.7) |  |  |
| Spherical equivalent, diopter | -0.05 ± 1.88 | -0.05 ± 1.88 | - | - | - | - |
| Intraocular pressure, mmHg | 15.21 ± 2.90 | 15.21 ± 2.90 | - | - | - | - |
| Lipid-lowering medication |  |  |  |  |  |  |
| No | 76961 (82.0) | 6525 (83.4) | 35231 (81.9) | 35205 (81.9) | **0.004** | 0.825 |
| Yes | 16877 (18.0) | 1299 (16.6) | 7776 (18.1) | 7802 (18.1) |  |  |
| Antihypertensive medication |  |  |  |  |  |  |
| No | 73674 (78.5) | 6424 (82.1) | 33630 (78.2) | 33620 (78.2) | **<0.001** | 0.941 |
| Yes | 20164 (21.5) | 1400 (17.9) | 9377 (21.8) | 9387 (21.8) |  |  |
| Insulin |  |  |  |  |  |  |
| No | 93056 (99.2) | 7772 (99.3) | 42631 (99.1) | 42653 (99.2) | 0.164 | 0.435 |
| Yes | 782 (0.8) | 52 (0.7) | 376 (0.9) | 354 (0.8) |  |  |

§ Bold indicates statistically significant.
† Comparison of characteristics between population I and population II.
‡ Comparison of characteristics between training set and validation set in population II.

### Metabolite associated with GCIPLT

**Supplementary Figure S2** shows the association between the metabolites and macular GCIPL thickness. Principal component analysis of the 249 circulating metabolites showed that 55 principal components accounted for >95% of the total variation. Thus, a Bonferroni-corrected P-value of <0.009 (0.05/55) was considered statistically significant. Of these, 16 metabolic indicators reached significance and were considered GCIPLT metabolomic signatures (**Supplementary Figure S3)**.

Higher levels of phospholipids in medium high-density lipoprotein (HDL) and total HDL, total lipids in medium HDL and total HDL, free cholesterol in medium HDL and total HDL, and cholesterol and cholesteryl esters in medium HDL were significantly associated with reduced macular GCIPLT. In addition, apolipoprotein A1 (apoA1), medium HDL particle concentration, total HDL particles, ratios of saturated fatty acids (FAs) to total FAs, and ratios of phospholipids to total lipids in small HDL were significantly associated with reduced macular GCIPLT. In contrast, the ratios of linoleic acid to total FA, omega-6 FA to total FA, and apolipoprotein B (apoB) to apoA1 were positively correlated with GCIPLT (**Supplementary Results, Supplementary Table S2**).

### Metabolomic signature and risk of incident morbidity and mortality

In the phase II analysis, after follow-up (median=12.0 years), 6,524 participants died. Of these, 1,544 died of cardiovascular disease; 3,151, of cancer, and 1,559 of other causes (**Supplementary Table S3**). A total of 5,714 participants developed T2DM, 1,366 developed OSAHS, 1,219 developed dementia, 1,578 developed ischemic stroke, 2,537 developed heart failure, and 2,866 had MI. These participants were associated with older age, male sex, higher Townsend deprivation indices, lower income, higher BMI, were smokers, or drank alcohol more frequently (all P<0.001). The baseline characteristics of the participants stratified by outcomes are summarized in **Supplementary Tables S4–S10**.

**Figures 3 and 4** show the correlation of the GCIPLT metabolic signatures with the risk of mortality and systemic diseases. Of the 16 aforementioned metabolic markers, numerous were independently associated with each health outcome. Consistent correlations were obtained in the validation set (**Supplementary Results, Supplementary Tables S14–S23**).

**Figure 3.**
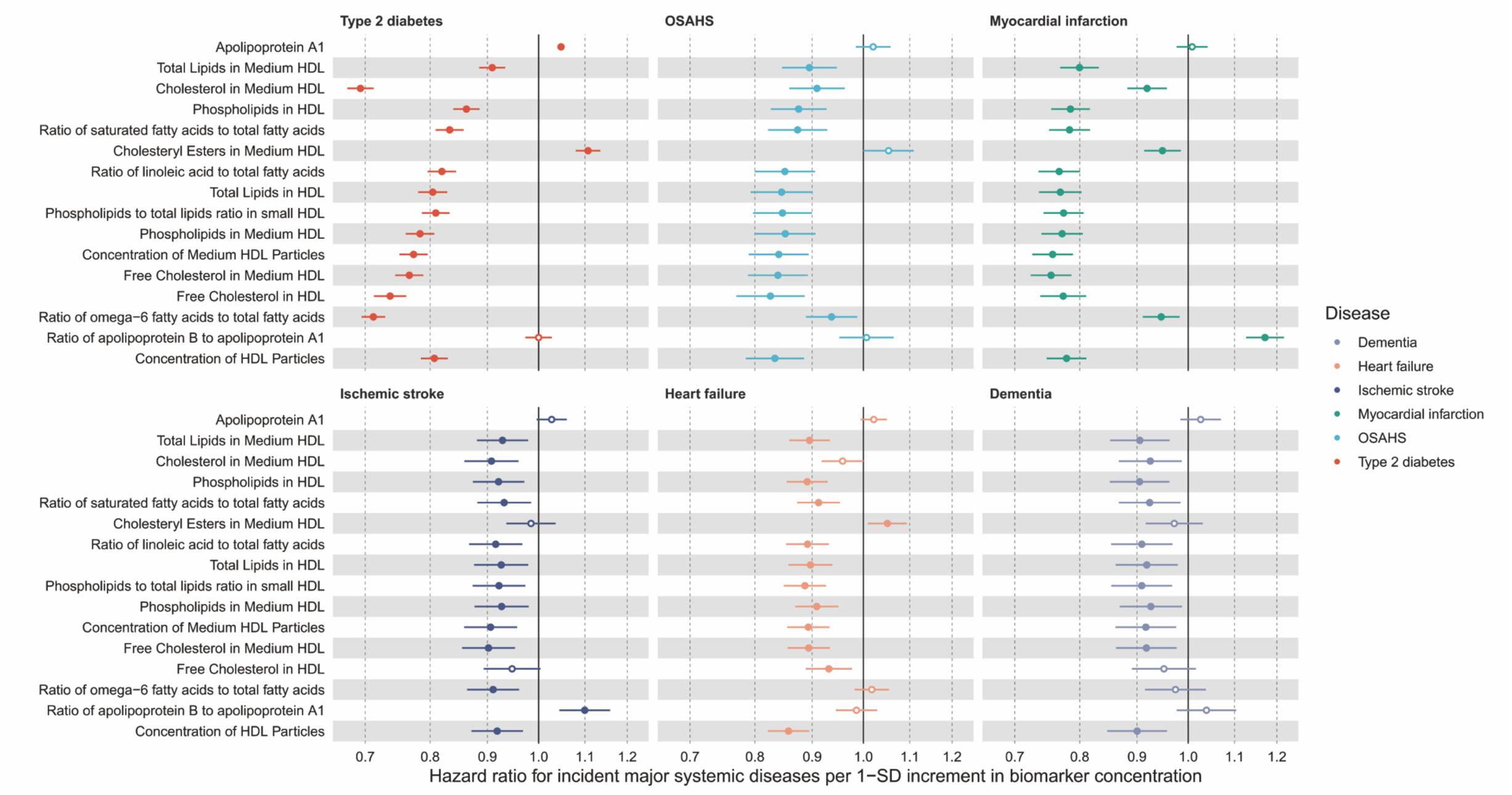
Associations of GCIPLT metabolomic signature and risk of type 2 diabetes, dementia, OSAHS, heat failure, myocardial infarction, and ischemic stroke. GCIPLT =ganglion cell-inner plexiform layer thickness; OSAHS =obstructive sleep apnea/hypopnea syndrome; HDL =high-density lipoprotein.

**Figure 4.**
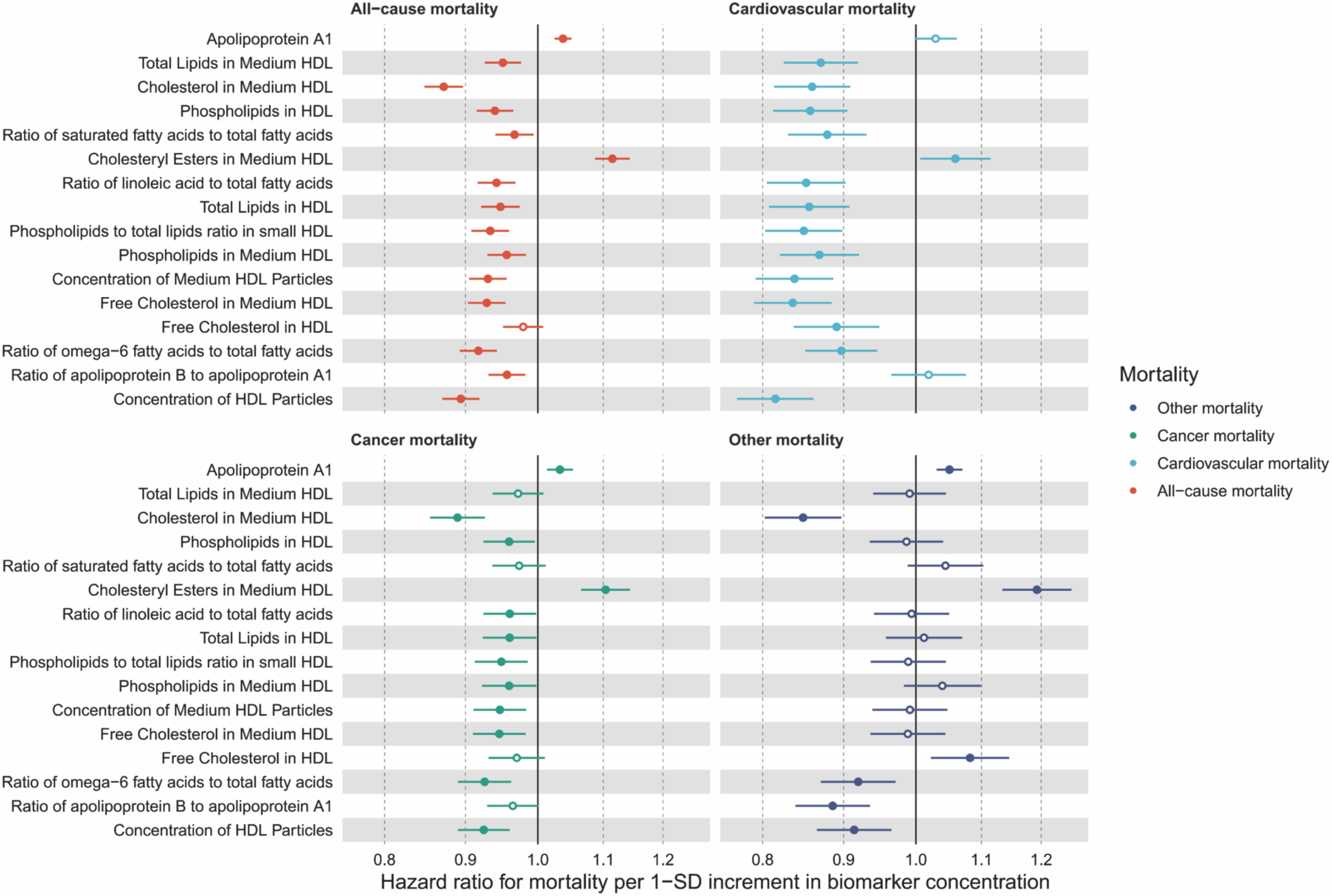
Associations of GCIPLT metabolomic signature and risk of all-cause mortality, cardiovascular mortality, cancer mortality, and other mortality. GCIPLT =ganglion cell-inner plexiform layer thickness; HDL =high-density lipoprotein.

### Meta-GCIPLT score between high- and low-risk participants

Participants with metabolic scores in the top 10% percentile had significantly higher risks for all- and specific-cause mortality as well as systemic diseases than those in the bottom 10% percentile (all P <1.48E-11) (**Supplementary Figures S4–S5**). Similar results were obtained for the validation set (**Supplementary Figures S6–S7**). This suggests that the GCIPLT metabolic score distinguishes between individuals at low and high risk of mortality and major systemic diseases.

### Predictive power of Meta-GCIPLT score

The GCIPLT metabolomic signature showed a predictive value higher than or comparable to that of traditional risk factors for all endpoints (**Figures 5–6**). For mortality prediction, GCIPLT metabolomic signatures had the best prediction capacity aside from age in all models, with an C-statistic of 0.630 (95% confidence interval [CI]:0.618, 0.642) for all-cause mortality (age, 0.687; 95% CI:0.677, 0.697; P=2.38E-15), 0.685 (0.662–0.707) for cardiovascular mortality (age, 0.688; 95% CI:0.670, 0.707, P=0.363), 0.638 (0.636–0.660) for other mortality types (age, 0.700; 95% CI:0.675, 0.713; P=8.05E-06), and 0.601 (0.585–0.617) for cancer mortality (age, 0.660; 95% CI, 0.646, 0.674; P=1.82E-11), exceeding all other risk factors (all P<1.33E-05).

**Figure 5.**
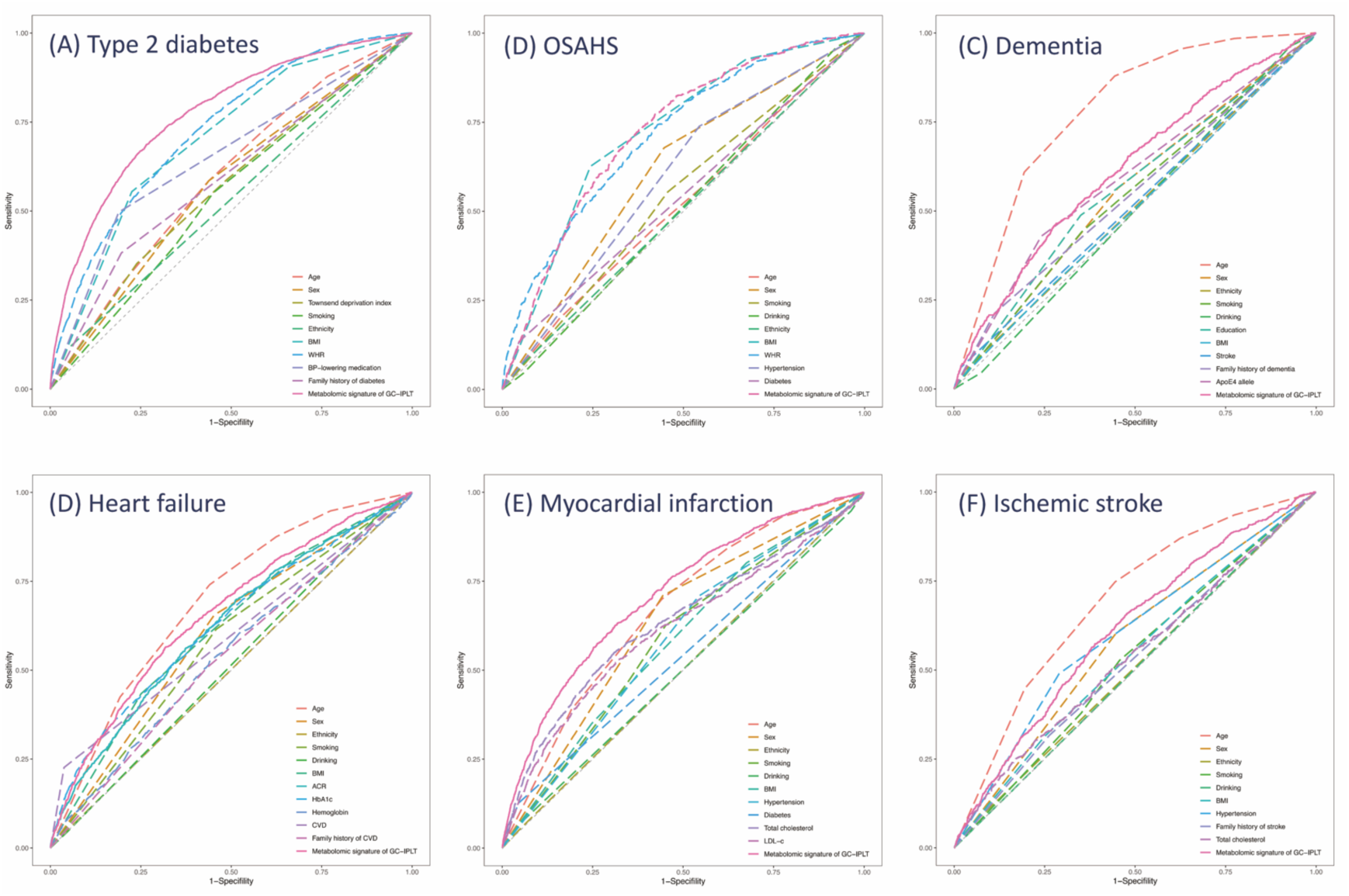
Receiver operating characteristic curves of established risk factors and meta-GCIPLT score for predicting (A) type 2 diabetes, (B) OSAHS, (C) dementia, (D) heat failure, (E) myocardial infarction, and (F) ischemic stroke. GCIPLT =ganglion cell-inner plexiform layer thickness; OSAHS =obstructive sleep apnea/hypopnea syndrome; BMI =body mass index; WHR =waist-to-hip ratio; BP =blood pressure; apoE = apolipoprotein E; ACR =microalbumin/creatinine ratio; HbA1c =glycosylated hemoglobin A1c; CVD =cardiovascular disease; LDL-c =low-density lipoprotein cholesterol.

**Figure 6.**
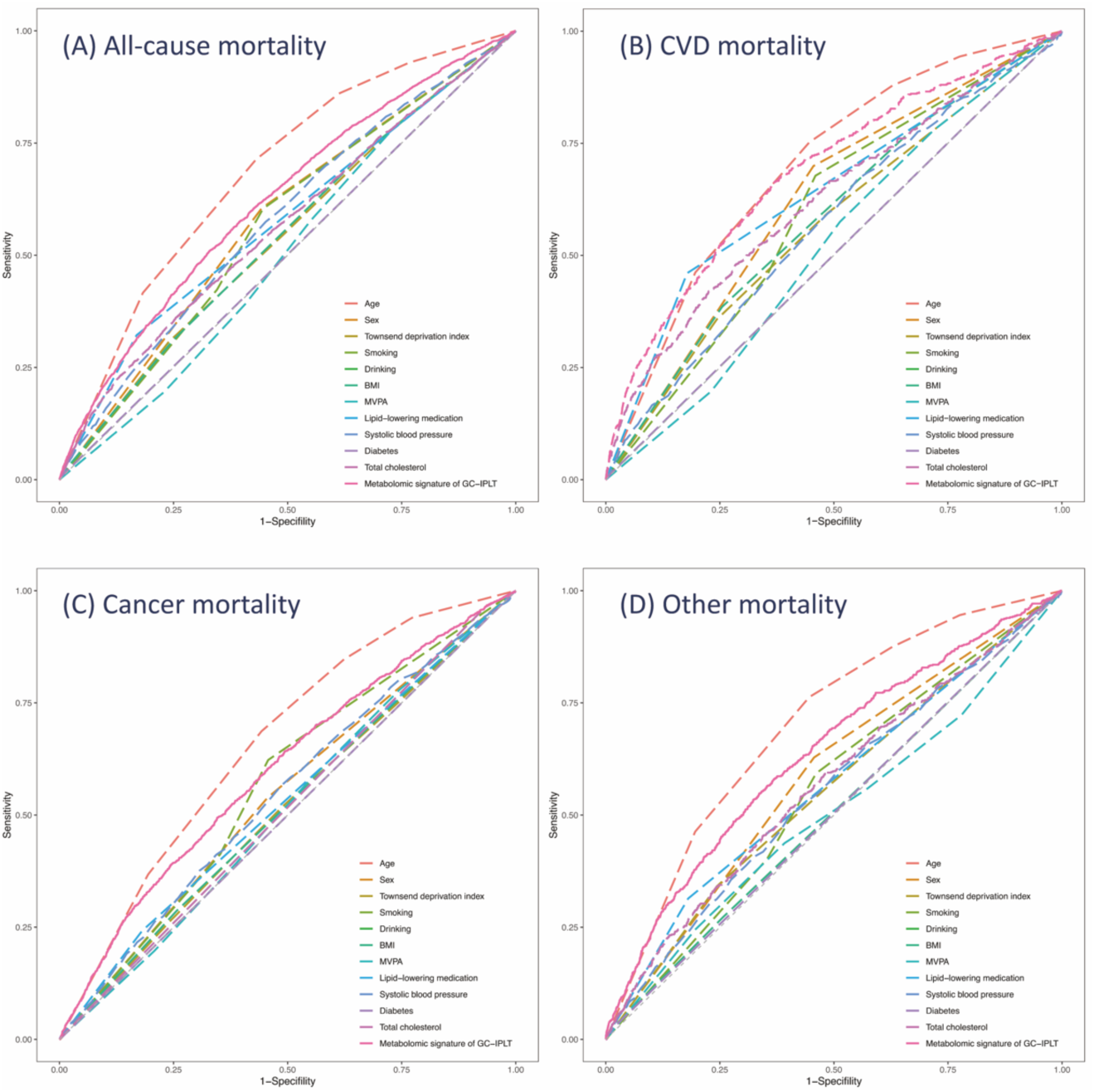
Receiver operating characteristic curves of established risk factors and meta-GCIPLT score for predicting (A) all-cause mortality, (B) CVD mortality, (C) cancer mortality, and (D) other mortality types. GCIPLT =ganglion cell-inner plexiform layer thickness; CVD =cardiovascular disease; BMI =body mass index; MVPA =moderate to vigorous physical activity.

In the models predicting the risk of T2DM, GCIPLT metabolomic signatures had the largest C-statistic (0.780; 95% CI:0.771–0.788), which exceeded all traditional risk factors (all P<2.20E-16). In the models predicting the development of OSAHS, GCIPLT metabolomic signatures and BMI had the largest C-statistic (0.725; 95% CI:0.707, 0.743, and 0.724; 95% CI:0.708, 0.741, respectively, P=0.856) compared with other risk factors (all P<0.005). For the prediction of dementia, age had the highest C-statistic (0.768; 95% CI:0.753, 0.784), followed by GCIPLT metabolomic signatures and the apoE ε4 allele (0.620; 95% CI:0.598, 0.643, and 0.594; 95% CI:0.574, 0.614, respectively, P=0.128), followed by other traditional risk factors (all P<0.001).

When comparing the C-statistics of the models predicting the risk of MI, GCIPLT metabolomic signatures had the largest C-statistic (0.711; 95% CI:0.695, 0.726) compared with traditional risk factors (all P<2.52E-05). For the prediction of ischemic stroke, age had the highest C-statistic (0.692; 95% CI:0.674–0.710), followed by the GCIPLT metabolomic signature and hypertension (0.614; 95% CI:0.593, 0.634, and 0.600; 95% CI:0.582, 0.618; P=0.259), followed by other risk factors (all P<0.005). In the models predicting the risk of developing heart failure, age had the highest C-statistic (0.690; 95% CI:0.676, 0.705), followed by GCIPLT metabolomic signatures (0.657; 95% CI:0.640, 0.674), and other risk factors (all P<0.005). Consistent results were obtained in the validation set (**Supplementary Figures S8–S9**).

### Incremental value of GCIPLT metabolomic signatures

After adding the GCIPLT metabolomic signatures to the conventional model, an increase in C-statistic was observed for all endpoints. (**Supplementary Results, Table 2, Supplementary Figures S10-S11**). Similar results were obtained for the validation set (**Supplementary Figures S12–S13**).

**Table 2.**
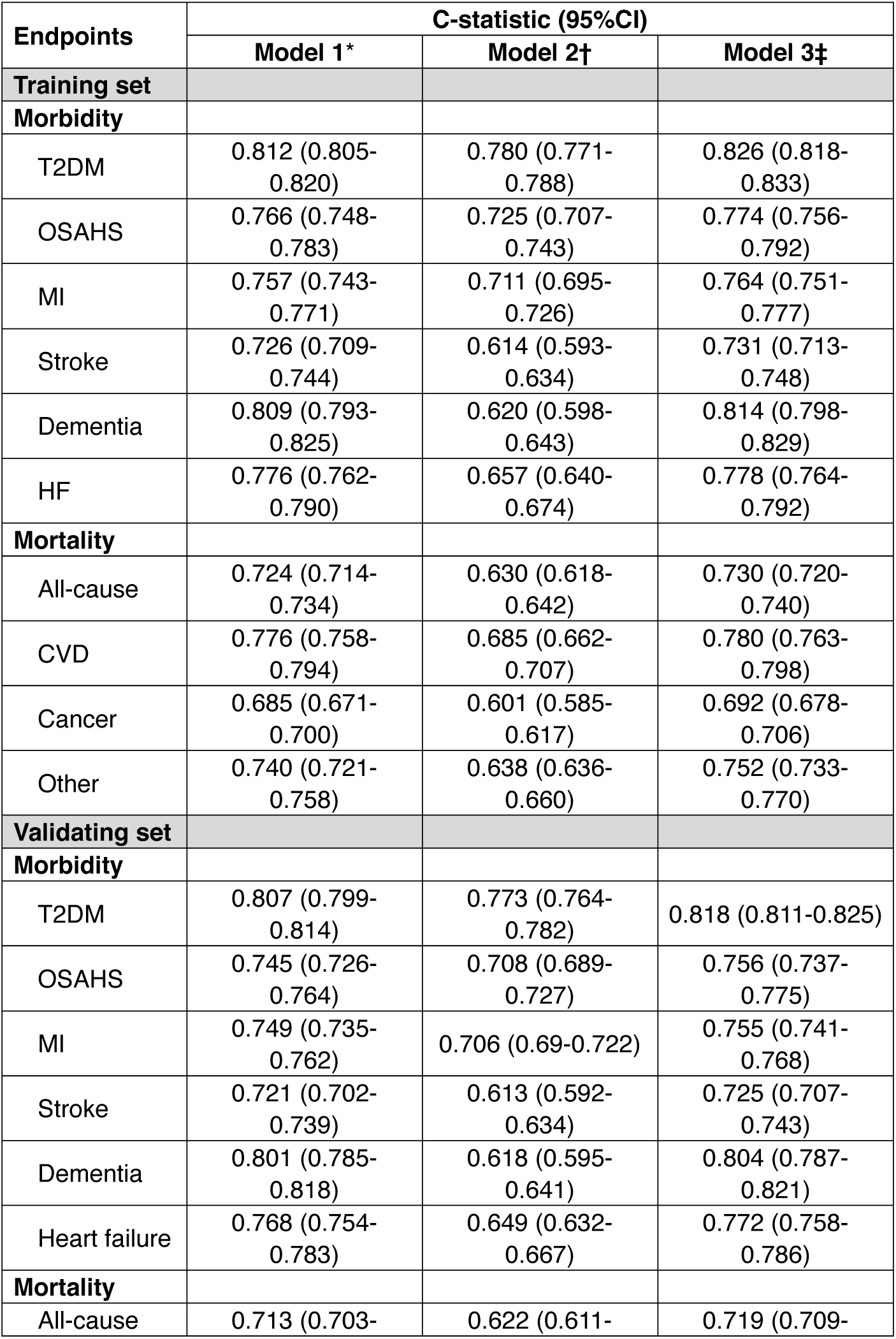
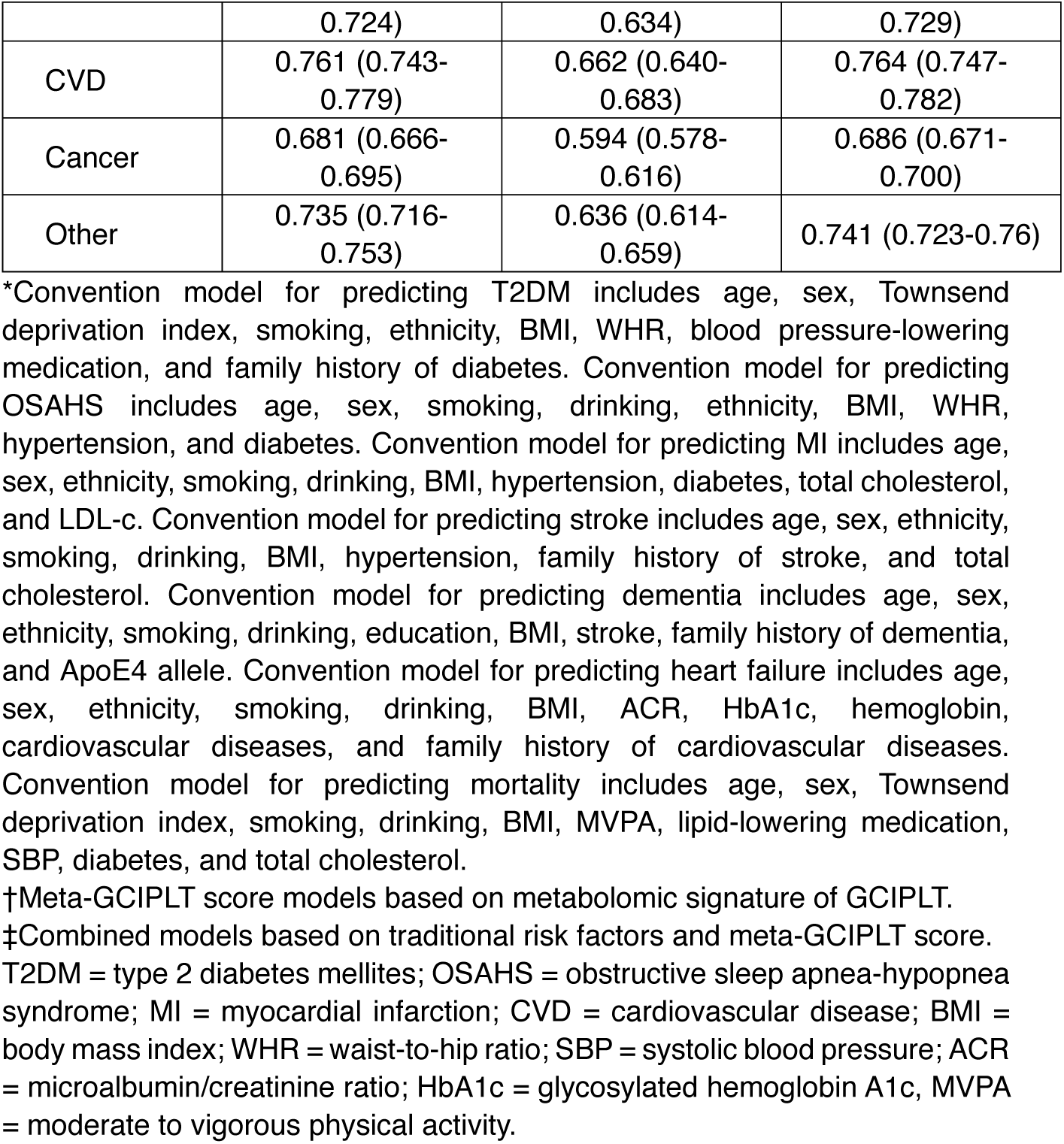
Discriminative power of traditional factors and meta-GCIPLT score for predicting mortality and major systemic diseases.

NRIs quantified the net benefits in the reclassification ability of adding metabolic scores to conventional models for each disease. After the addition of GCIPLT metabolomic signatures, NRI values were 2.39% (standard error [SE]=0.006, P=3.89E-05) for predicting all-cause mortality, 4.20% (SE=0.012, P=6.04E-04) for predicting cardiovascular mortality, 2.09% (SE=0.010, P=0.039) for predicting cancer mortality, and 5.17% (SE=0.021, P=0.042) for predicting other mortality types. Similarly, the NRI values were 5.18% (SE=0.026, P=3.86E-11) for predicting T2DM, 2.94% (SE=0.007, P=3.13E-05) for predicting OSAHS, 3.73% (SE=0.007, P=1.72E-07) for predicting MI, 1.59% (SE=0.004, P=2.72E-08) for predicting heart failure, 2.33% (SE=0.009, P=0.049) for predicting stroke, and 4.43% (SE=0.006, P=3.01E-12) for predicting dementia (**Supplementary Figures S14–S15**). Similar results were obtained for the validation set (**Supplementary Results, Supplementary Figures S16–S17**).

### Calibration of the combined model

Good calibration between the predicted risk and actual onset was shown for all six systemic diseases and four mortality risks in both the training and validation sets (**Supplementary Figures S18–S21**).

## Discussion

In the present study, 249 plasma metabolites were tested for their association with GCIPLT and 16 were identified as metabolic signatures. Further analyses revealed an independent association of these metabolites with the risk of developing future mortality and major age-related diseases. After a metabolic score was constructed, GCIPLT signatures could differentiate between high- and low-risk patients for all health outcomes, and its predictive power was higher than or comparable to that of traditional risk factors. Their addition to conventional predictive models significantly improved model discrimination, suggesting that these metabolites have the potential to capture the residual risk for systemic diseases not quantified by traditional risk factors in the general population.

Among the 16 signatures identified, numerous were HDL-associated metabolites that were protective against all-cause, cardiovascular, and cancer-related mortality, reinforcing their powerful roles in systemic health and disease processes in the human body. In addition, these signatures had stronger predictive power than that of other risk factors, notably comparable to that of age, for the prediction of cardiovascular mortality. This is plausible since these signatures were associated with various major cardiovascular diseases in our analysis, including MI, heart failure, and stroke. Consistently, the improvement in the reclassification ability of the model predicting cardiovascular mortality was the greatest among models predicting mortality with the inclusion of GCIPLT signatures. While HDL-associated metabolites reduce the risk of cardiovascular diseases, our analysis concluded that it was also associated with future cancer mortality, which is currently a debated topic.^32^ A possible explanation is that apoA1 and HDL components in tumors promote cholesterol efflux and inhibit the growth and proliferation of tumor cells with high cholesterol demands.^33^ On analysis of other mortality types, it was puzzling that HDL was deemed a risk factor for unspecified mortality, while apoB/apoA1 were protective. We hypothesize that these unexpected associations are likely due to heterogeneity within mortality types, where metabolites played no significant role in death (for e.g., death by accidents). These populations may not necessarily have any systemic metabolic alterations, which partly explains this confusion.

T2DM is a metabolic disease characterized by insulin insensitivity, which causes impaired glucose uptake and systemic fat mobilization.^34^ This study implicates decreased HDL cholesterol, and changes in its phospholipids, FAs, and other related metabolites as factors that confer an increased risk of T2DM. Our findings likely reflect the complex shifts in lipid metabolism that occur in T2DM. Precisely, the synthesis and release of TG-rich very low-density lipoprotein (VLDL) in the liver drives the exchange of TG and cholesterol esters between VLDL and HDL, leading to a decrease in HDL cholesterol in T2DM.^35^ In addition, unstable TG-rich HDL particles are considered more susceptible to clearance,^36^ explaining the negative association between HDL and apoA1 levels and the risk of developing T2DM in this study. Meanwhile, such component modifications have been reported to alter the functional domains of apoA1 structures, also preventing interactions with HDL that limit their physiological function.^37^ Since HDL particles are primarily responsible for reverse transport of cellular cholesterol, their reduction or inactivity can cause lipid accumulation in pancreatic β-cells, causing inflammation and impaired β-cell function.^37-40^ This cholesterol transport activity and antioxidant capacity of HDL were also reported to depend on its surface lipid components, which determines the protein’s mobility^41-43^; hence, decreased ratios of unsaturated FAs to saturated FAs, phospholipids, and free cholesterol observed in the present study impair antioxidant capacity, which predispose an individual to T2DM. Notably, the predictive value of this group of metabolites for diabetes exceeded that of BMI and WHR, suggesting their exceptional importance in T2DM risk.

OSAHS is a common sleep disorder and airway disease characterized by sleep apnea, causing subsequent chronic hypoxia.^44^ Elevated levels of free FAs (FFAs) are common in mice exposed to intermittent hypoxia (IH) and in OSAHS patients,^45-49^ likely from IH-associated activation of the sympathetic system that triggers the release of FFAs from adipose tissue.^50^ Similar to T2DM, FFAs are sent to the liver to synthesize TG-rich VLDL, which ultimately decreases HDL cholesterol and clearance of unstable HDL particles,^36^ as supported by our results where similar lipid metabolite profiles were implicated in OSAHS. IH is also thought to impair the sensitivity of adipose tissue to insulin, which further leads to increased FFAs release and decreased HDL cholesterol and HDL particles.^51^ In addition, patients with OSAHS have been reported to undergo lipid peroxidation more frequently,^52^ hence higher levels of HDL particles with higher phospholipid and free cholesterol content that are associated with good antioxidant capacity were deemed protective against OSAHS in the present study.^42-43^

The present study found that apoA1, HDL particle concentration and multiple components within HDL were independently associated with decreased risk of dementia, a neurodegenerative disease affecting over 50 million people worldwide.^53^ Debate about the prospective relationship between plasma HDL levels and dementia is ongoing,^54^ and although it was believed that apoE-based HDL in the brain differs from apoA1-based HDL in plasma, it was recently demonstrated that plasma apoA1-HDL crossed the blood–brain barrier through scavenger receptors, inferring it could participate in brain lipid metabolism.^55^ Extracellular deposition of amyloid β (Aβ) is thought to be the initiating event for dementia,^56^ and *in vitro* studies show that apoA1 binds to Aβ to interfere with Aβ monomer assembly, preventing neurotoxicity.^57-58^ In addition, Aβ-bound HDL was reported to promote *in situ* degradation of Aβ by binding to scavenger receptors on glial cells,^59^ explaining why plasma apoA1 and HDL were protective against dementia in this study. In addition, apoA1 or apoA1-HDL has been observed to promote non-amyloidogenic cleavage of the Aβ protein rather than amyloidogenic cleavage, thereby promoting cellular cholesterol efflux and increasing cell membrane fluidity.^60^ Considering phospholipids and cholesterol in HDL were associated with reduced dementia risk in the present study, these consolidate the current literature and provide further insight into the pathophysiology of dementia.

Previous studies have identified low plasma HDL concentrations are independent risk factors for cardiovascular diseases, since HDL mediates reverse transport of cholesterol in atherosclerotic plaques^61-62^. In the present study, we observed that apoA1, HDL particle concentration, cholesteryl esters, free cholesterol, and phospholipids in HDL particles protect against heart failure, MI, and stroke. An increase in these components contributes to the efflux of cellular cholesterol via HDL and prevents the deposition of oxidized lipids, which otherwise leads to vascular inflammation.^41-42^ In addition, linoleic and omega-6 FAs in total FAs were also protective against MI and stroke, which is important considering that the role of omega-6 FAs in cardiovascular disease remains uncertain. Some studies suggest that linoleic acid, a major omega-6 FA in the Western diet, may reduce the risk of cardiovascular disease, while others have concerns that it can elongate to form arachidonic acid, which has potential pro-inflammatory and thrombogenic properties that may be harmful to the heart.^63-64^ Recent randomized controlled trials show that elevated dietary linoleic acid has no significant effect on inflammation, immune activation, or platelet function, presumably due to its limited conversion to arachidonic acid in humans.^65^ These results alongside the present findings suggest that linoleic acid is protective against cardiovascular diseases, and considering it was also protective for T2DM, OSAHS, dementia, and all-and specific-cause mortality in our results, its benefits should be considered in dietary recommendations.

Despite the breadth of new information, this study had certain limitations. First, some patients did not have diagnostic data in their initial hospital records; therefore, their ages at diagnosis were based on self-reported questionnaires. As self-reported data are corruptible to memory, this may have introduced a bias for some associations. Second, participants with retinal OCT measurements in this study were younger, male, more educated, earned a higher income, had lower BMIs, and were non-smokers compared to other participants (**Table 1**). Therefore, caution should be exercised before generalizing the metabolomic signatures to the general population. Finally, although a comprehensive range of confounders was adjusted for in this analysis, potential residual confounders that could not be excluded may still exist.

## Conclusion

In summary, this study identified GCIPLT metabolomic signatures that had higher predictive power than traditional risk factors for major systemic diseases and causes of mortality. GCIPLT-associated plasma metabolites have the potential to capture the residual risk of systemic diseases and mortality not quantified by traditional risk factors. This study contributes new knowledge that deepens our understanding of the retina as a window to systemic health, and paves the way for developing strategies targeting metabolites that may reverse or interrupt health outcomes.

## Data Availability

All data produced in the present study are available upon reasonable request to the authors.

## Abbreviations and Acronyms

OCT: optical coherence tomography
GCIPL: ganglion cell-inner plexiform layer
RNFL: retinal nerve fiber layer
GCIPLT: GCIPL thickness
RGC: retinal ganglion cell
OSAHS: obstructive sleep apnea/hypopnea syndrome
MI: myocardial infarction
NHS: National Health Service
NMR: nuclear magnetic resonance
BMI: body mass index
WHR: waist-to-hip ratio
IOP: intraocular pressure
CI: confidence interval
HDL: high-density lipoprotein
apoE: apolipoprotein E
apoA1: apolipoprotein A1
FA: fatty acid,
SE: standard error
apoB: apolipoprotein
B; TG: triglyceride
VLDL: very low-density lipoprotein
IH: intermittent hypoxia
Aβ: amyloid β

## Author Contributions

Study concept and design: WW, ZZ, MH; Acquisition, analyses, or interpretation: All authors; Drafting of the manuscript: SY, WW YY; Critical revision of the manuscript for important intellectual content: All authors; Statistical analyses: WW, SY, YY; Obtained funding: WW, ZZ; Administrative, technical, or material support: SZ, WH; Study supervision: WW, ZZ, MH.

## Data Sharing Statement

All data used in this study are available from UK Biobank via data access procedures (http://www.ukbiobank.ac.uk). Permission to use the UK Biobank Resource was obtained via material transfer agreement as part of Application 62443, 62489, 62491 and 62525.

## Acknowledgements

This study was funded by the National Natural Science Foundation of China (82000901), the Guangzhou Science & Technology Plan of Guangdong Pearl River Talents Program (202102010162), the Fundamental Research Funds of the State Key Laboratory of Ophthalmology (303060202400362). The authors thank all the participants and staff in the UKB.

