## Supplementary materials for "Metabolic signature of the ganglion cell–inner plexiform layer thickness and the risks of mortality and morbidity: a population-based study in UK Biobank"

### SUPPLEMENTARY TEXT

#### Supplementary Methods

In the present study, age was divided into five categories: ≤ 49, 50–54, 55–59, 60–64, and ≥ 65 years; education into four categories: level O, level A, university, and missing; and ethnicity into three categories: white, other, and missing. The Townsend deprivation index was divided into four quartiles. The average household income was divided into five categories: <18k, 18k-30k, 31k-51k, 52k-100k, >100k, and missing. Baseline diseases were defined using questionnaires, interviews, and inpatient data based on the ICD-10 codes.

Body mass index (BMI) was calculated as weight divided by height squared and waist-to-hip ratio (WHR) as waist circumference divided by hip circumference. The blood pressure was measured using a digital blood pressure monitor (Omron, Japan). Visual acuity was assessed using the traditional LogMAR charts. The refractive error was measured using an autorefractor (Tomey, Japan), and spherical equivalent values were calculated based on the autorefraction results. Intraocular pressure (IOP) was measured using an ocular response analyzer (Rerchert, USA). Total cholesterol, low-density lipoprotein cholesterol (LDL-c), and high-density lipoprotein cholesterol (HDL-c) levels were measured using Konelab (Thermo Fisher, USA). Glycosylated haemoglobin A1c was measured using liquid chromatography (Bio-Rad, USA). Urine microalbumin and creatinine levels were measured using Beckman Coulter (USA), and the albumin-to-creatinine ratio was calculated. All field codes are summerized in **Supplementary Table S1**.

#### Supplementary Results

**Metabolite associated with GCIPLT**

Higher levels of phospholipids in medium high-density lipoprotein (HDL) (β=-0.284; 95% CI:-0.410, -0.157; P=1.02E-06) and total HDL (β=-0.279; 95% CI:-0.411, -0.148; P=3.19E-05), total lipids in medium HDL (β=-0.271; 95% CI:-0.399, -0.143; P=3.17E-05) and total HDL (β=-0.260; 95% CI:-0.393, -0.127; P=1.34E-04), free cholesterol in medium HDL (β=-0.264; 95% CI:-0.394, -0.134; P=6.90E-05) and total HDL (β=-0.240; 95% CI:-0.375, -0.105; P=4.84E-04), and cholesterol (β=-0.251; 95% CI:-0.381, -0.121; P=1.56E-04) and cholesteryl esters (β=-0.247; 95% CI:-0.377, -0.117; P=1.90E-04) in medium HDL were significantly associated with reduced macular GCIPLT. In addition, apolipoprotein A1 (apoA1) (β=-0.253; 95% CI:-0.383, -0.124; P=1.29E-04), medium HDL particle concentration (β=-0.266; 95% CI:-0.394, -0.138; P=4.78E-05), total HDL particles (β=-0.213; 95% CI:-0.338, -0.088; P=8.47E-04), ratios of saturated fatty acids (FAs) to total FAs (β=-0.411; 95% CI:-0.576, -0.246; P=1.02E-06), and ratios of phospholipids to total lipids in small HDL (β=-0.257; 95% CI:-0.380, -0.134; P=4.16E-05) were significantly associated with reduced macular GCIPLT. In contrast, the ratios of linoleic acid to total FA (β=0.327; 95% CI:0.176, 0.477; P=2.03E-05), omega-6 FA to total FA (β=0.259; 95% CI:0.110, 0.408; P=6.85E-04), and apolipoprotein B (apoB) to apoA1 (β=0.222; 95% CI:0.093, 0.351; P=7.43E-04) were positively correlated with GCIPLT. See **Supplementary Table S2** for details.

**Metabolomic signature and risk of incident morbidity and mortality**

Of the 16 aforementioned metabolic markers, 15 were independently associated with all-cause mortality (13 negative, 2 positive), 14 with cardiovascular disease mortality (12 negative, 2 positive), 12 with cancer mortality (10 negative, 2 positive), and seven with other mortality types (4 negative, 3 positive). Among systemic diseases, 15 were independently associated with the risk of T2DM (13 negative, 2 positive), 13 were independently associated with the risk of OSAHS (all negative), 11 were independently associated with the risk of dementia (all negative), 13 were independently associated with the risk of stroke (12 negative, 1 positive), 12 were independently associated with the risk of heart failure (11 negative, 1 positive), and 15 were independently associated with the risk of heart attack (14 negative, 1 positive). Consistent correlations were obtained in the validation set. See **Supplementary Table S14-S23** for details.

**Incremental value of GCIPLT metabolomic signatures**

**C-statistics increased in new models**

The C-statistic increased from 0.724 (0.714–0.734) to 0.730 (0.720–0.740) (P=1.61E-10) for all-cause mortality; from 0.776 (0.758–0.794) to 0.780 (0.763–0.798) (P=0.006) for cardiovascular mortality; from 0.740 (0.721–0.758) to 0.752 (0.733–0.770) (P=3.27E-05) for other-cause mortality; and from 0.685 (0.671–0.700) to 0.692 (0.678–0.706) (P=5.66E-05) for cancer mortality (**Supplementary Figure S11**).

The meta-GCIPLT score increased the C-statistic from 0.812 (0.805–0.820) to 0.826 (0.818–0.833) (P < 2.20E-16) for predicting T2DM, from 0.766 (0.748–0.783) to 0.774 (0.756–0.792) (P=1.17E-05) for predicting OSAHS, from 0.757 (0.743–0.771) to 0.764 (0.751–0.777) (P=7.29E-13) for predicting myocardial infarction, from 0.776 (0.762–0.790) to 0.778 (0.764–0.792) (P=0.035) for predicting heart failure, from 0.726 (0.709–0.744) to 0.731 (0.713–0.748) (P=0.017) for predicting stroke, and from 0.809 (0.793–0.825) to 0.814 (0.798–0.829) (P=0.006) for predicting dementia (**Supplementary Figure S10**). Similar results were obtained for the validation set (**Supplementary Figures S12–S13**).

**NRI values in new models for the validation set**

The NRI values were 2.01% (SE=0.006, P=3.07E-04) for predicting all-cause mortality, 3.27% (SE=0.008, P=2.87E-05) for predicting cardiovascular mortality, 1.84% (SE=0.005, P=4.48E-04) for predicting cancer mortality, 4.08% (SE=0.006, P=5.06E-11) for predicting T2DM, 3.02% (SE=0.014, P=0.031) for predicting OSAHS, 3.09% (SE=0.009, P=4.45E-04) for predicting myocardial infarction, 1.27% (SE=0.003, P=3.07E-04) for predicting heart failure, and 3.54% (SE=0.012, P=0.004). The NRI values for the models predicting stroke, cardiovascular disease mortality, and other mortality types were not statistically significant (**Supplementary Figures S16–S17**).

### SUPPLEMENTARY FIGURES

#### Supplementary Figure S1

**Flowchart demonstrating the eligibility criteria.**

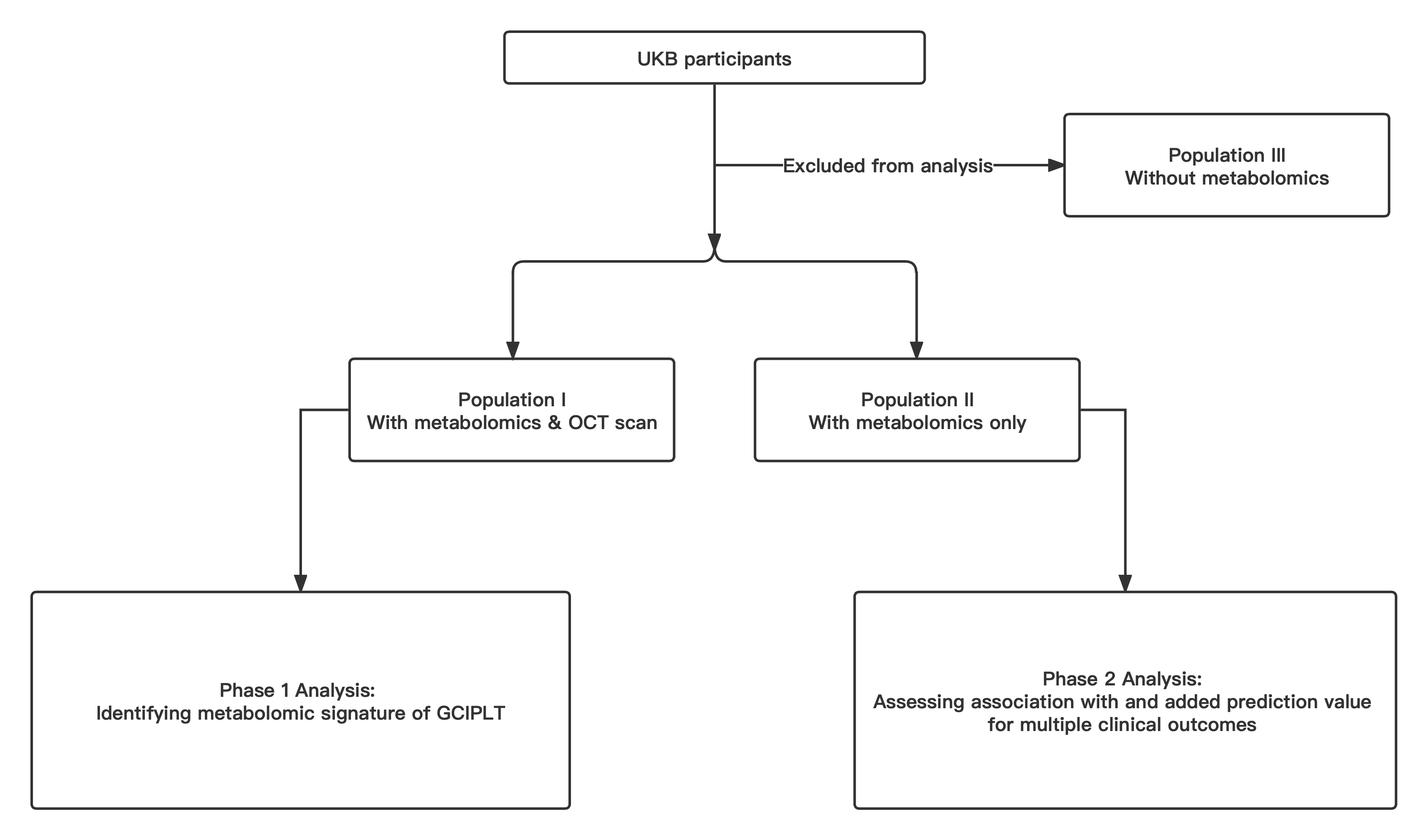

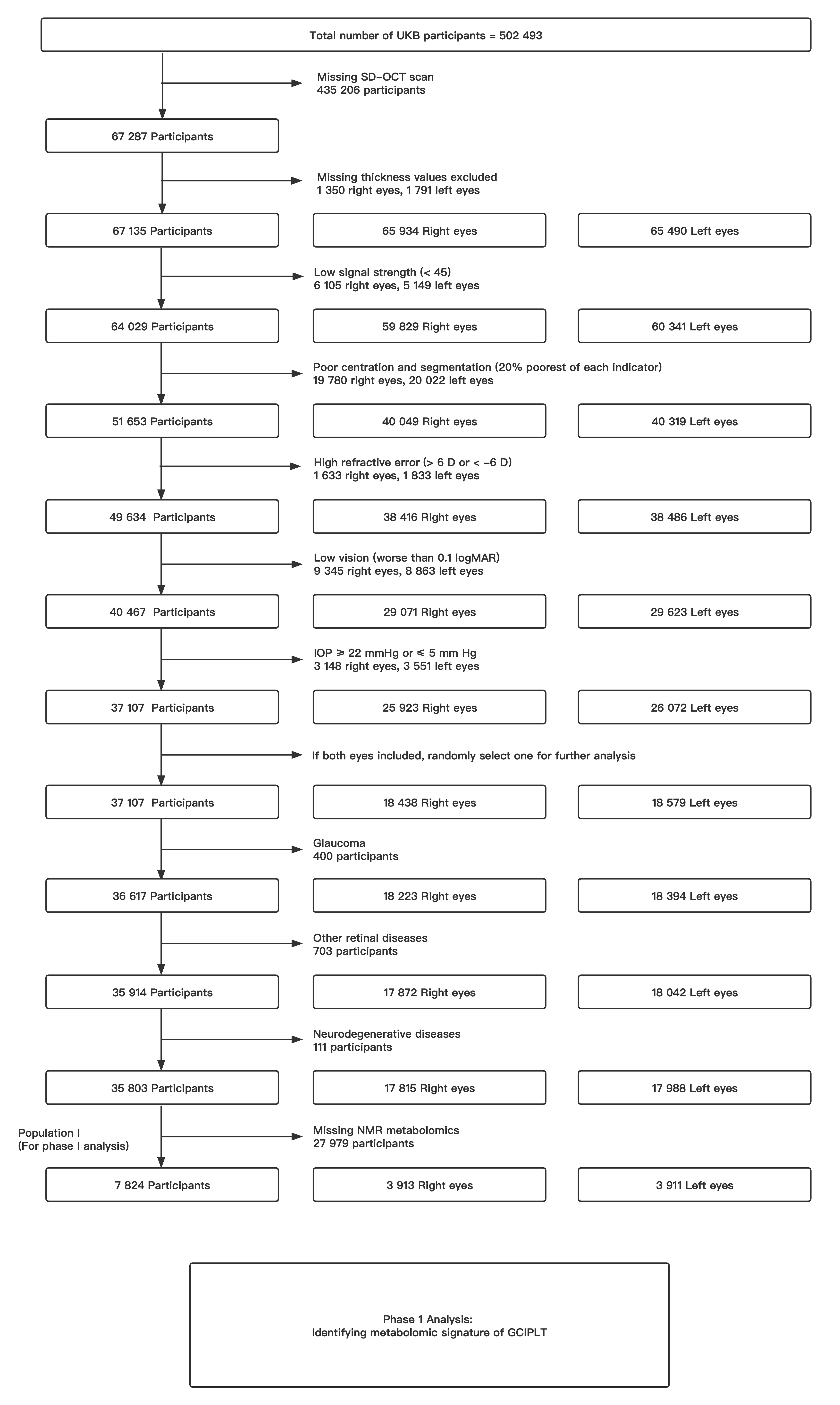

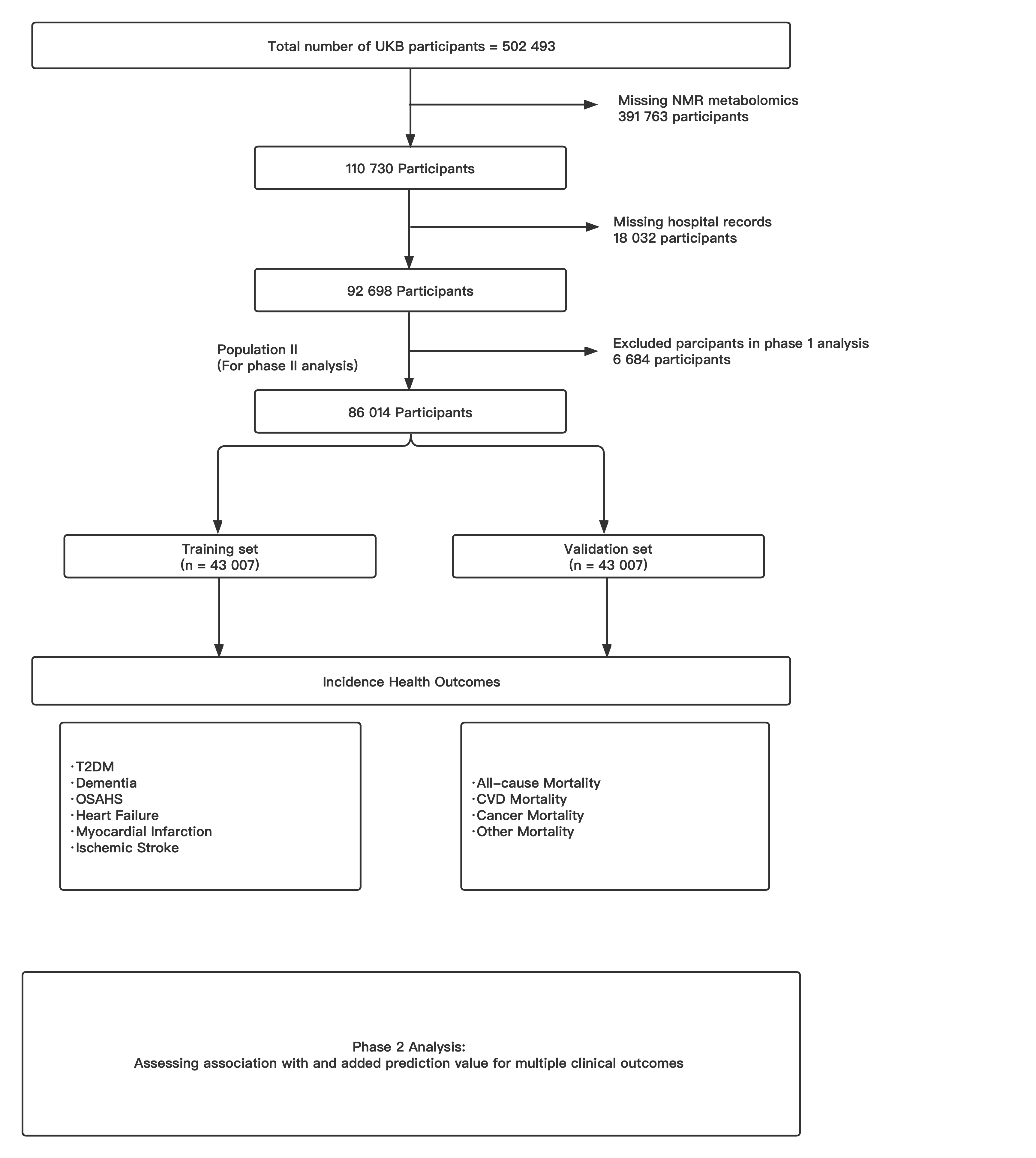

#### Supplementary Figure S2

**Heatmaps demonstrating the overall correlations of GCIPLT and metabolites.**

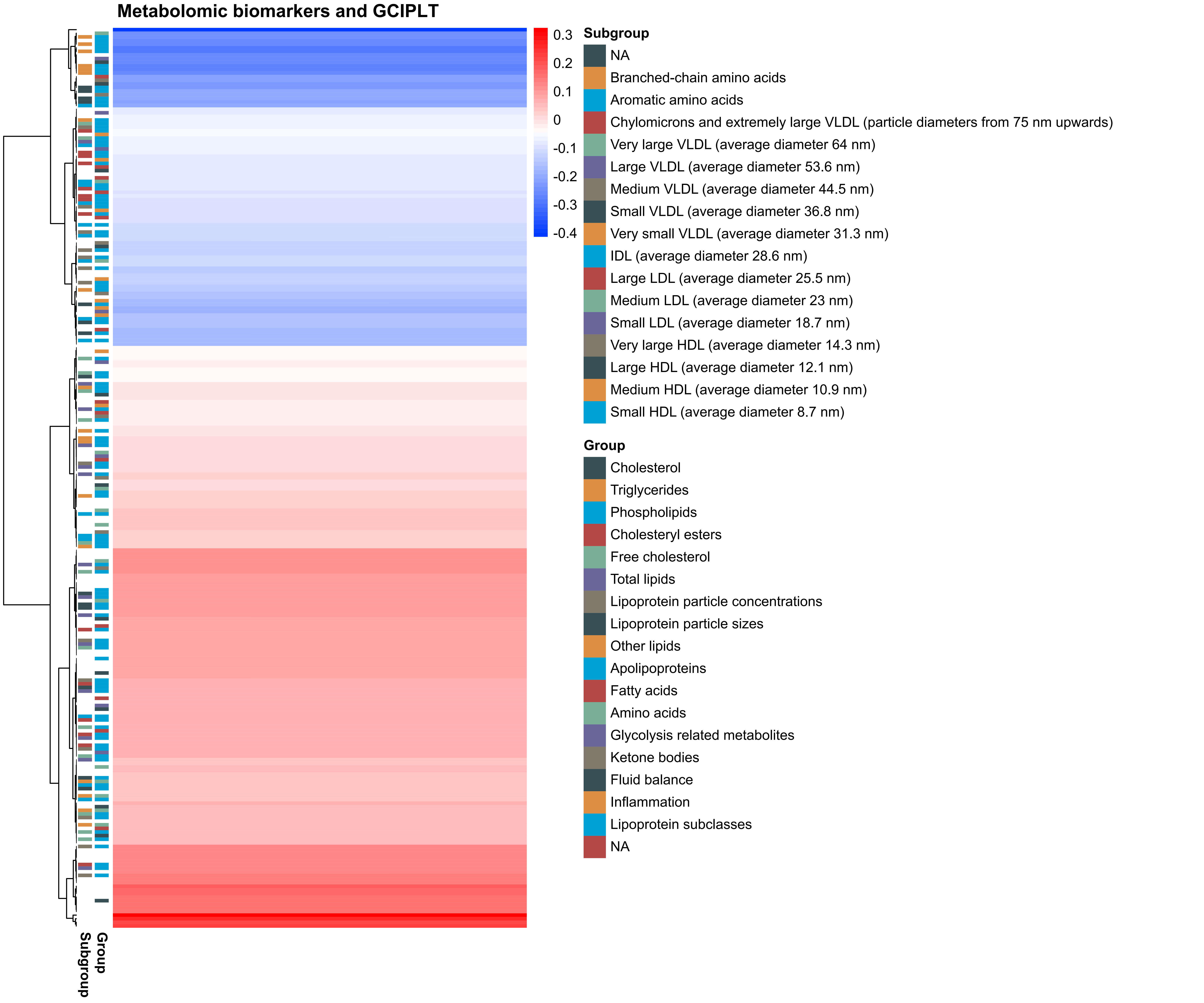

#### Supplementary Figure S3

**Forest plot demonstrating correlation between GCIPLT and metabolic biomarkers that reached the set significance threshold of P value.**

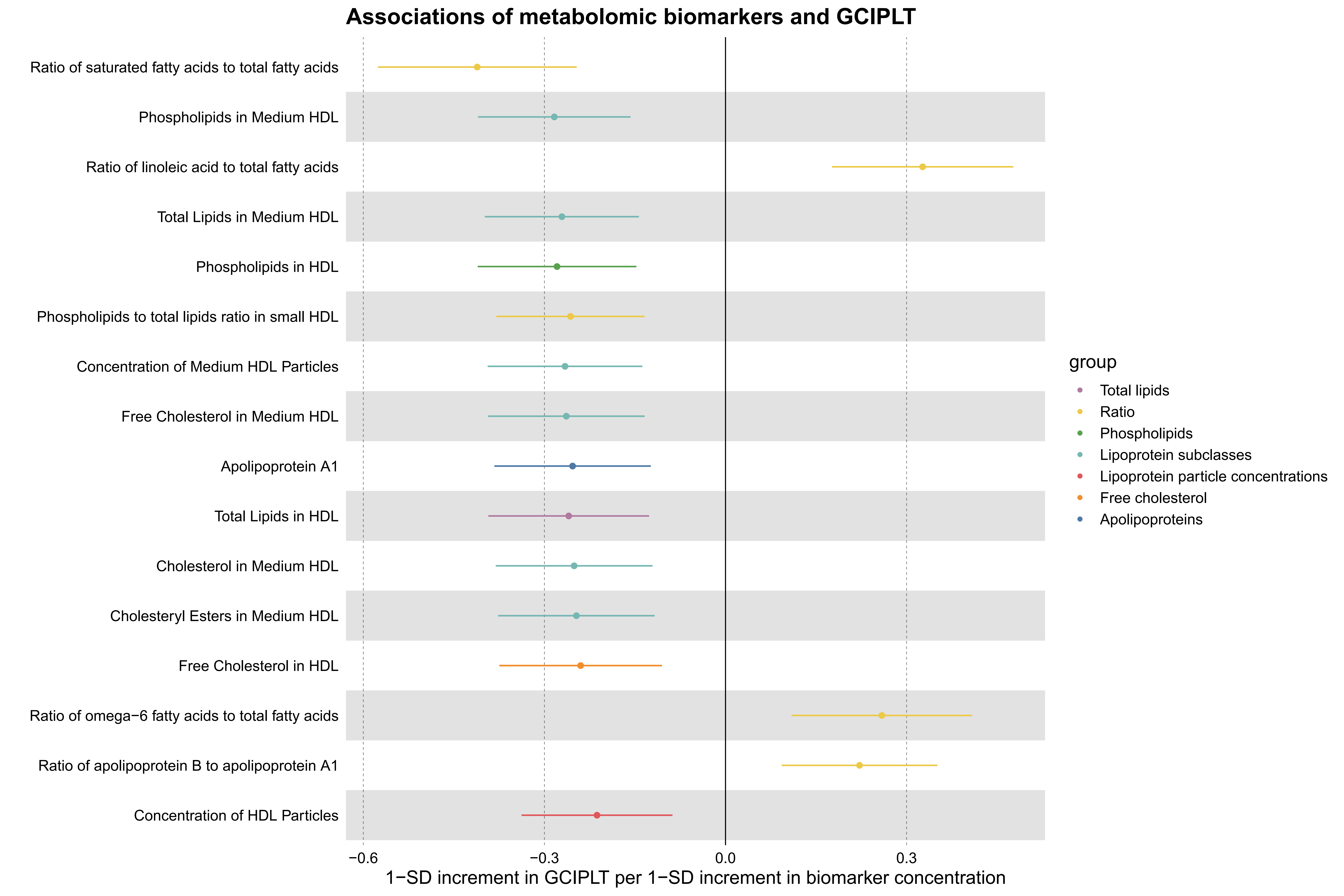

#### Supplementary Figure S4

**Comparisons of morbidity of systemic diseases in the top and bottom 10% of the meta-GCIPL score in training set.**

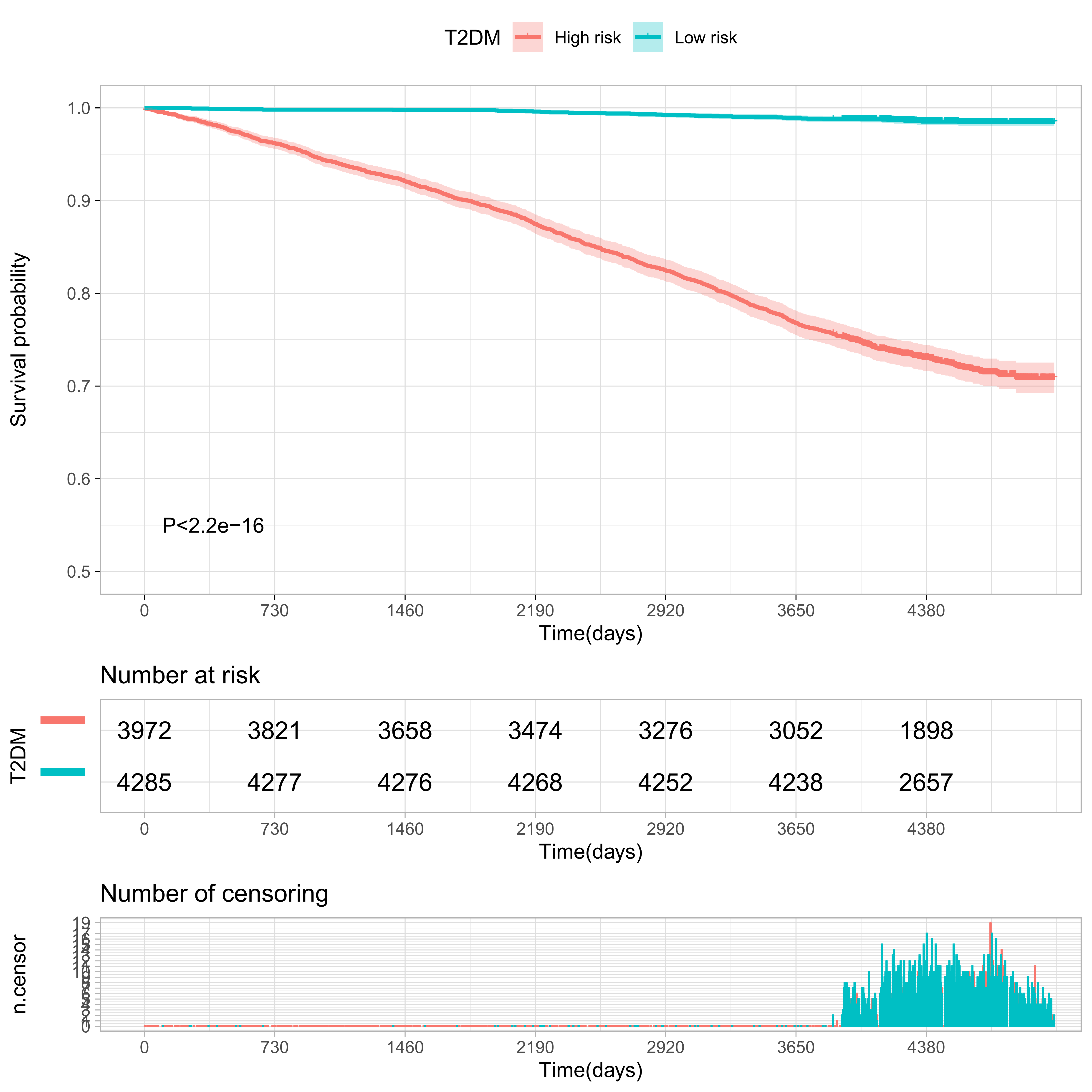

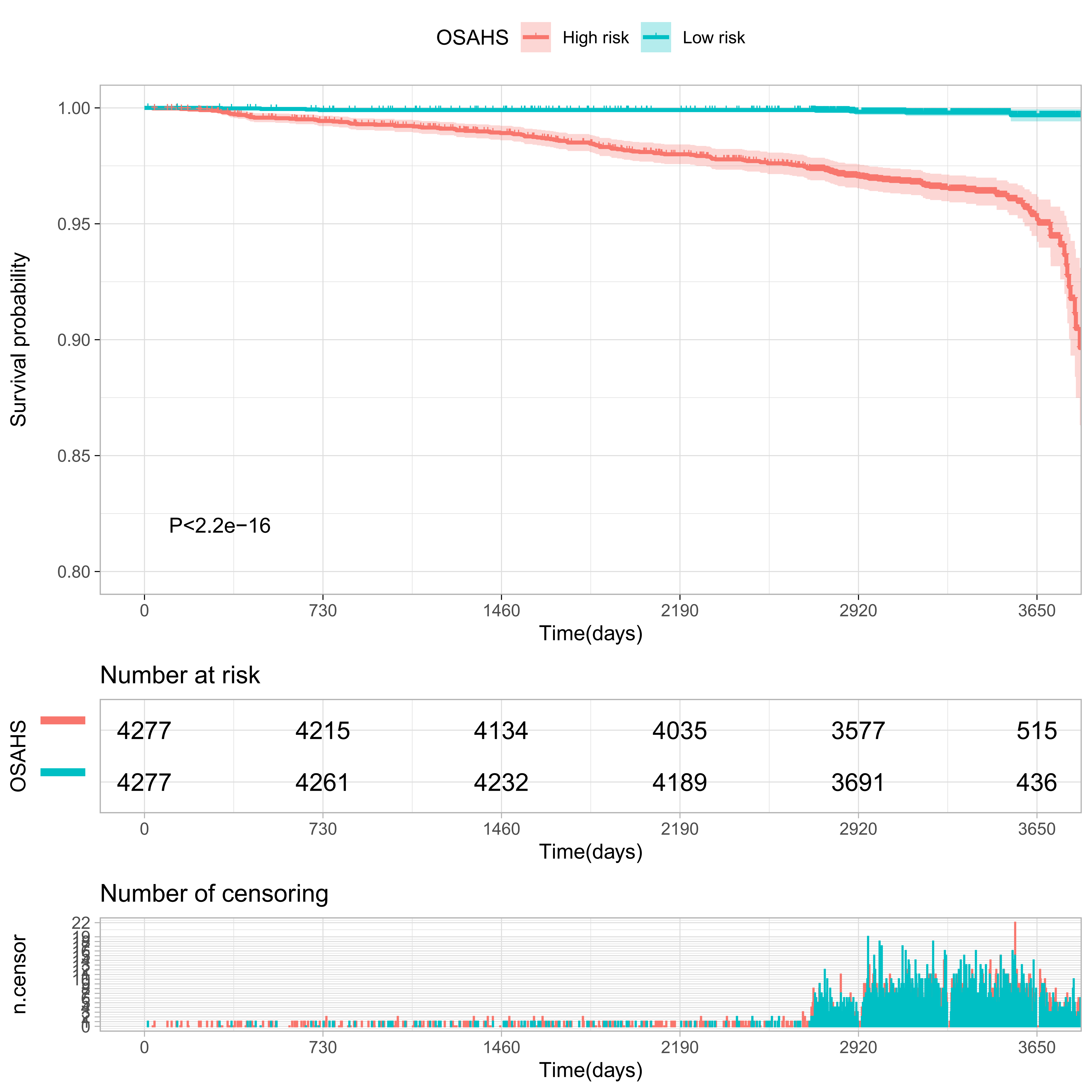

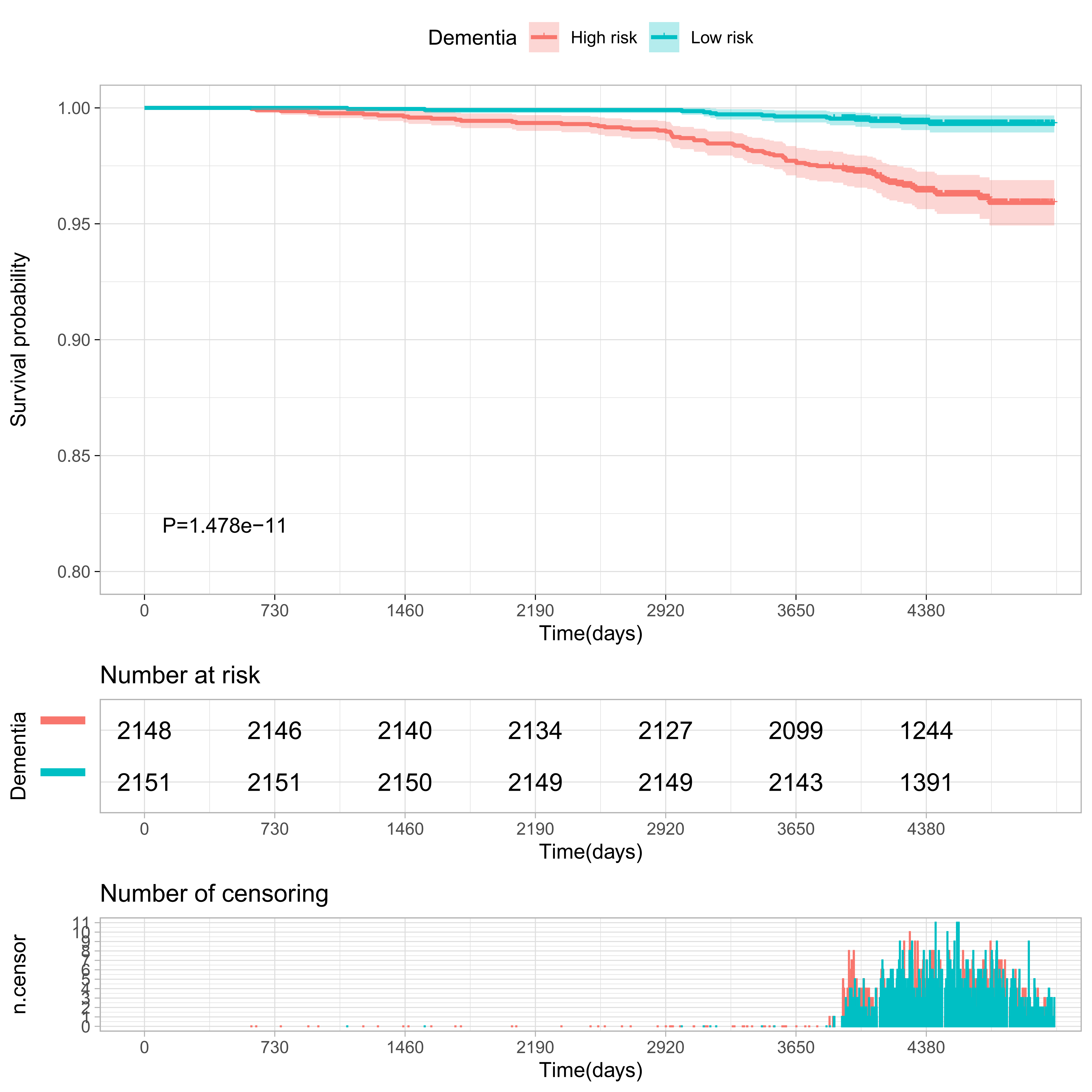

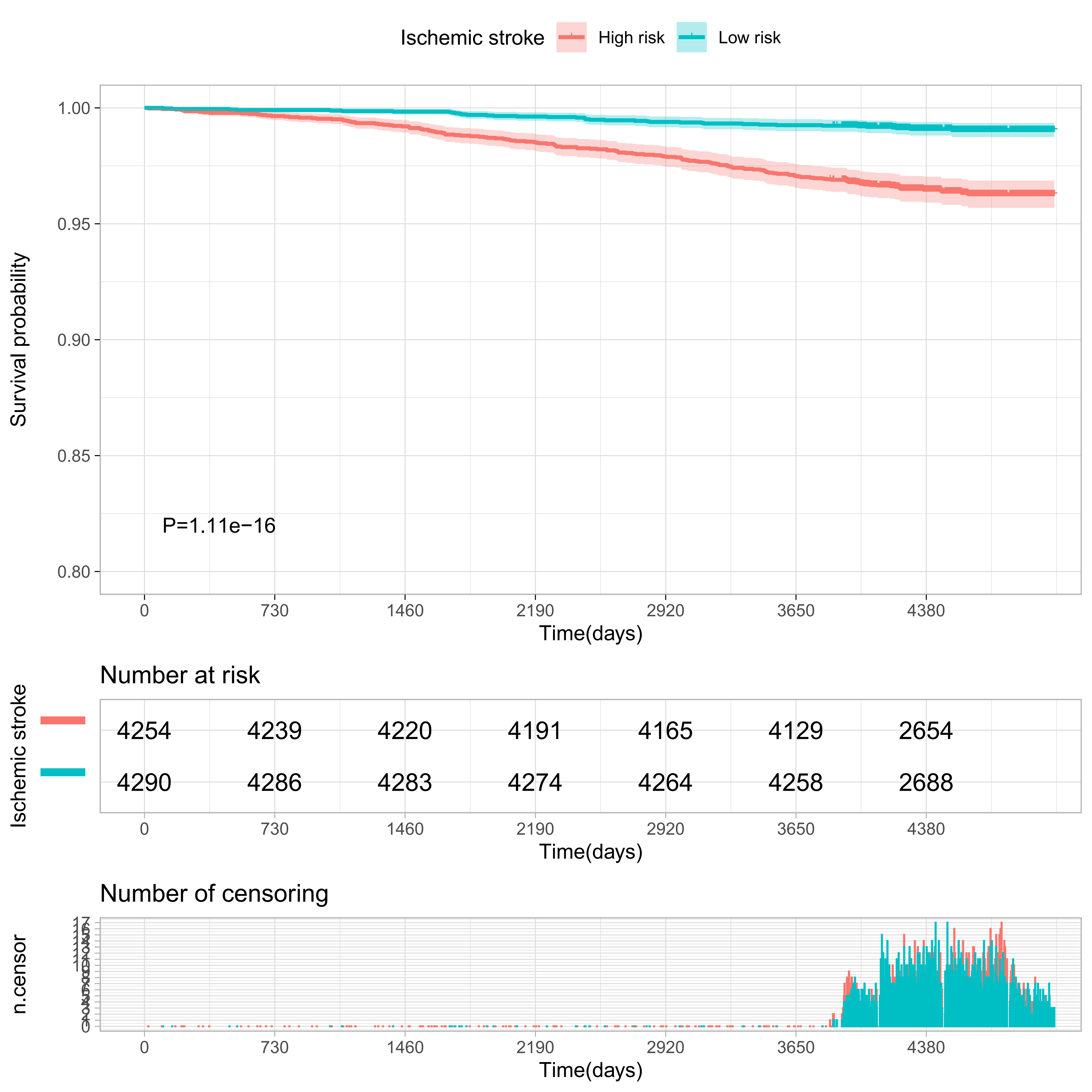

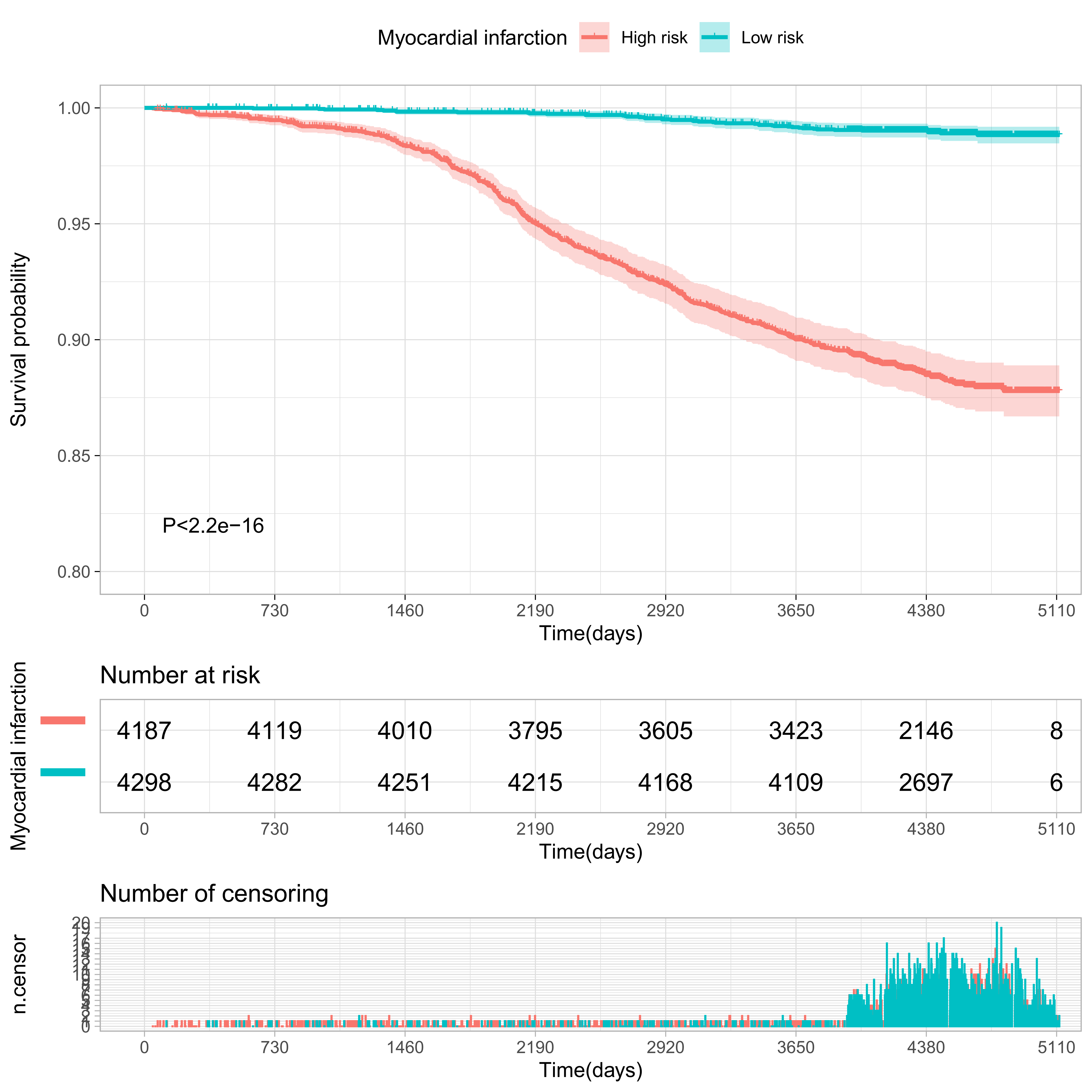

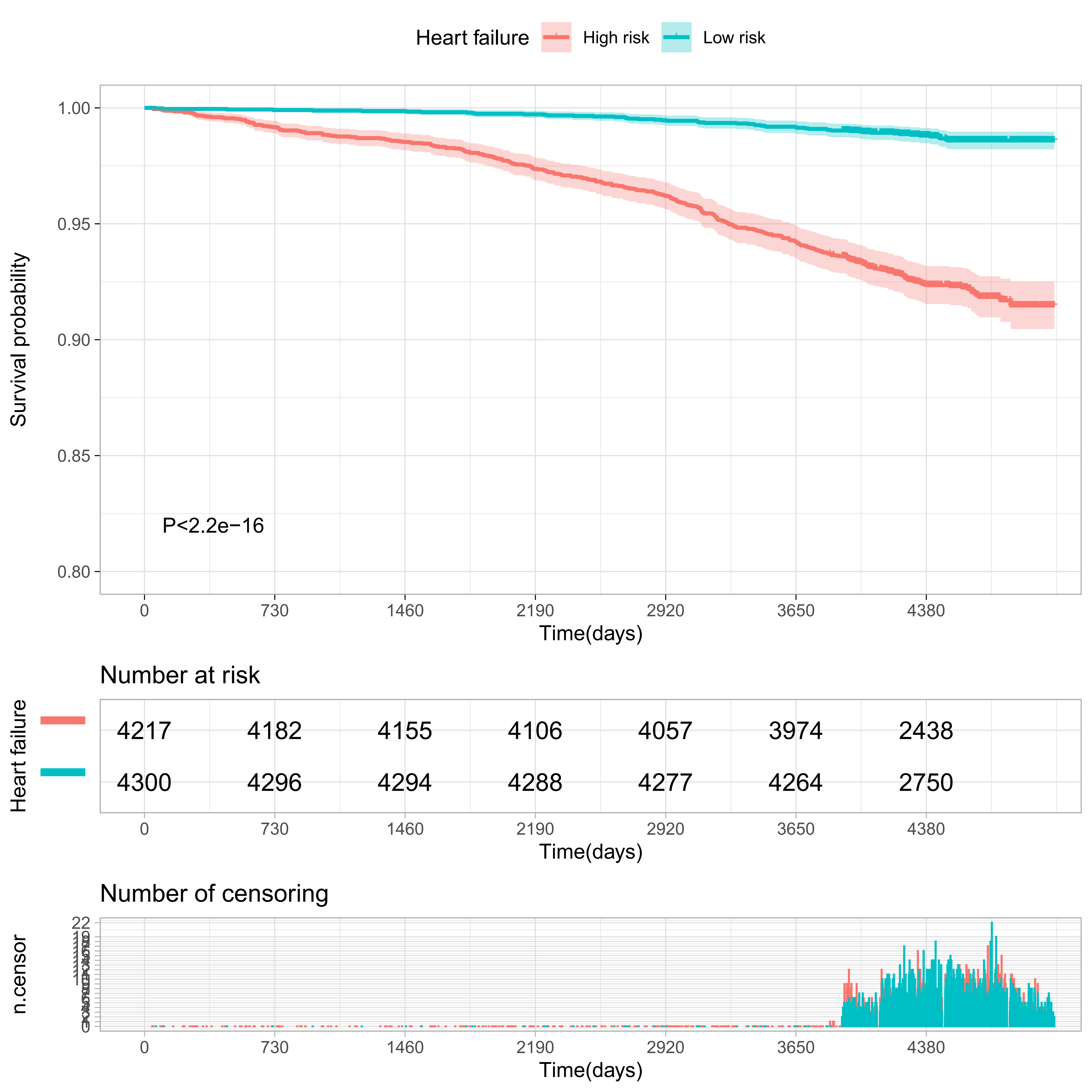

#### Supplementary Figure S5

**Comparisons of mortality in the top and bottom 10% of the meta-GCIPL score in training set.**

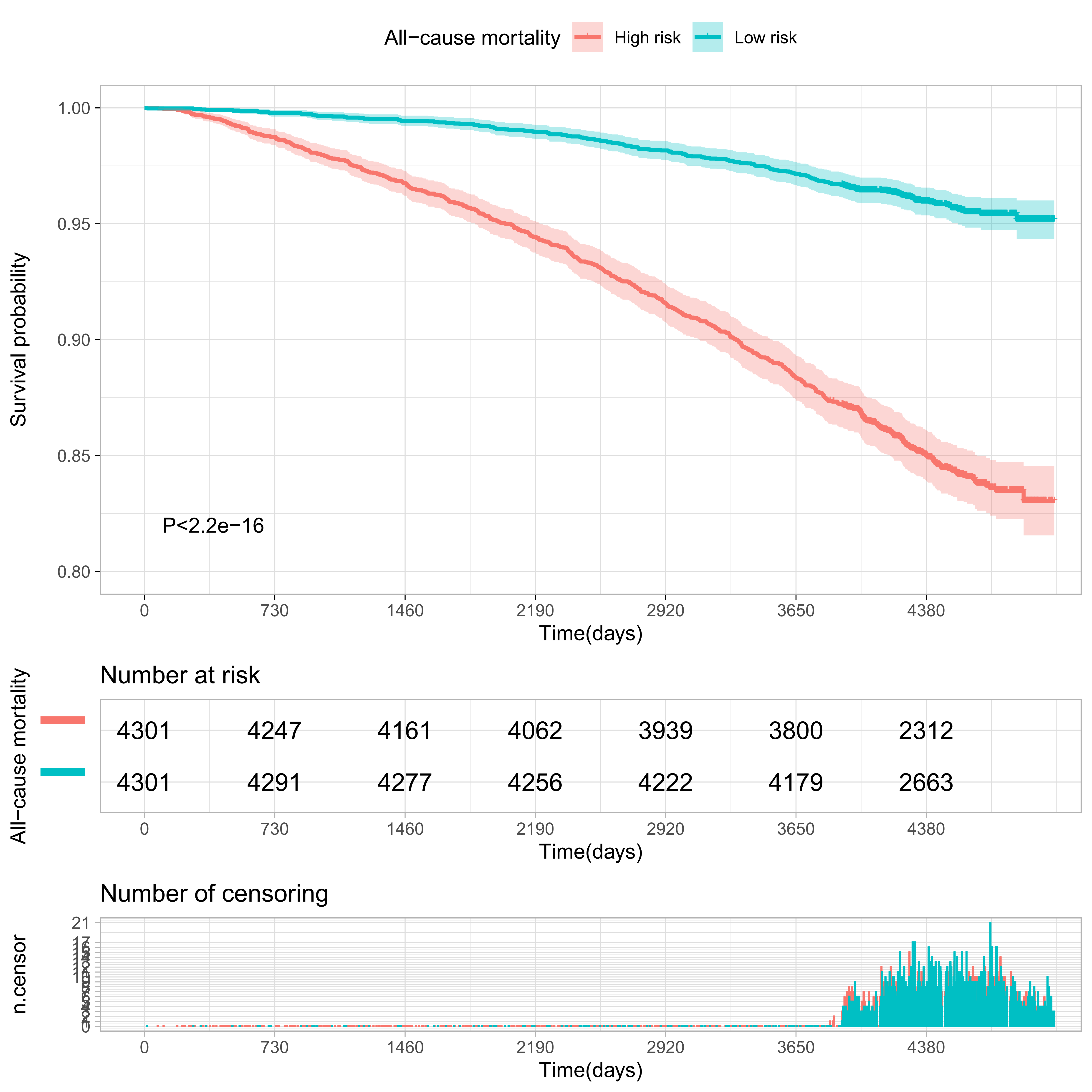

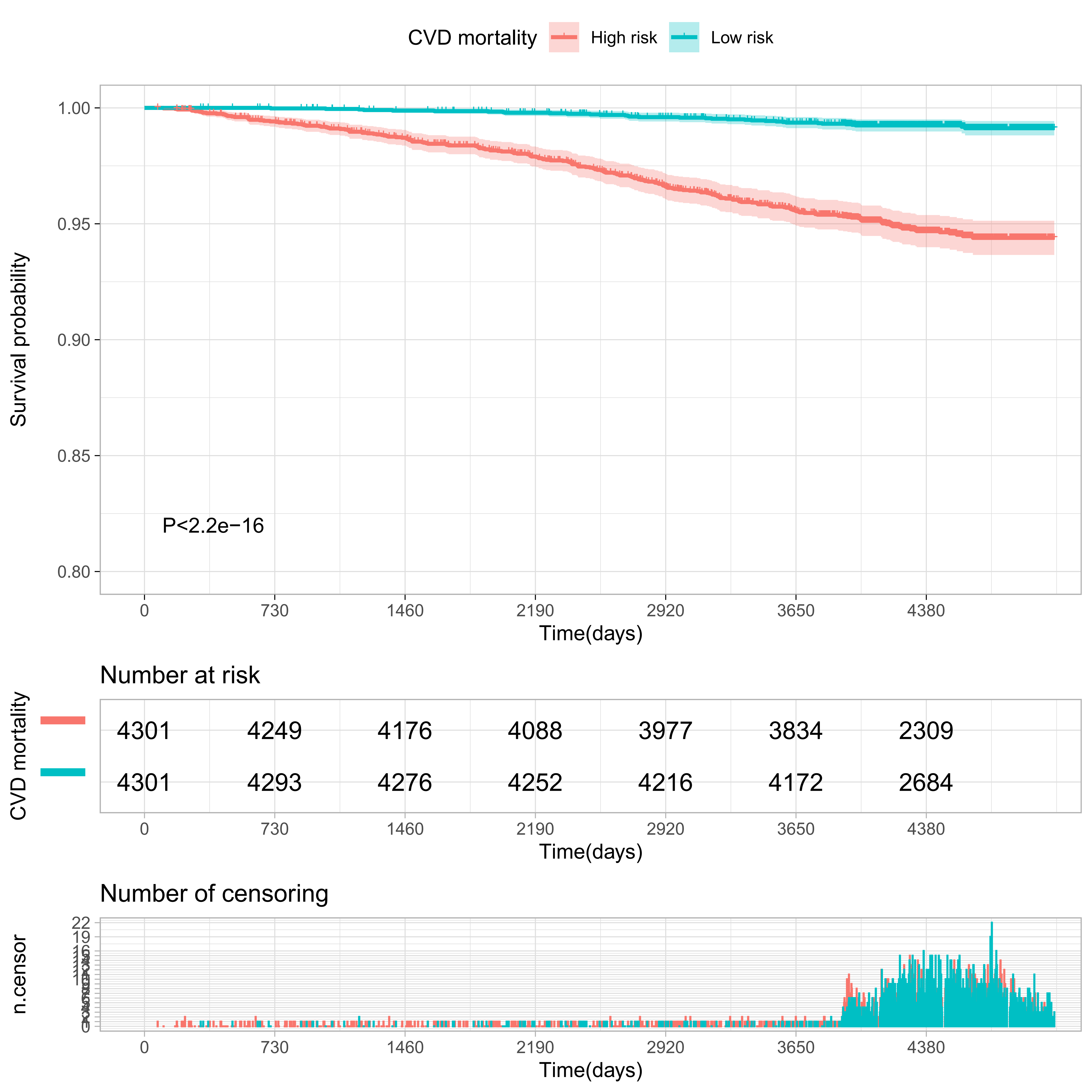

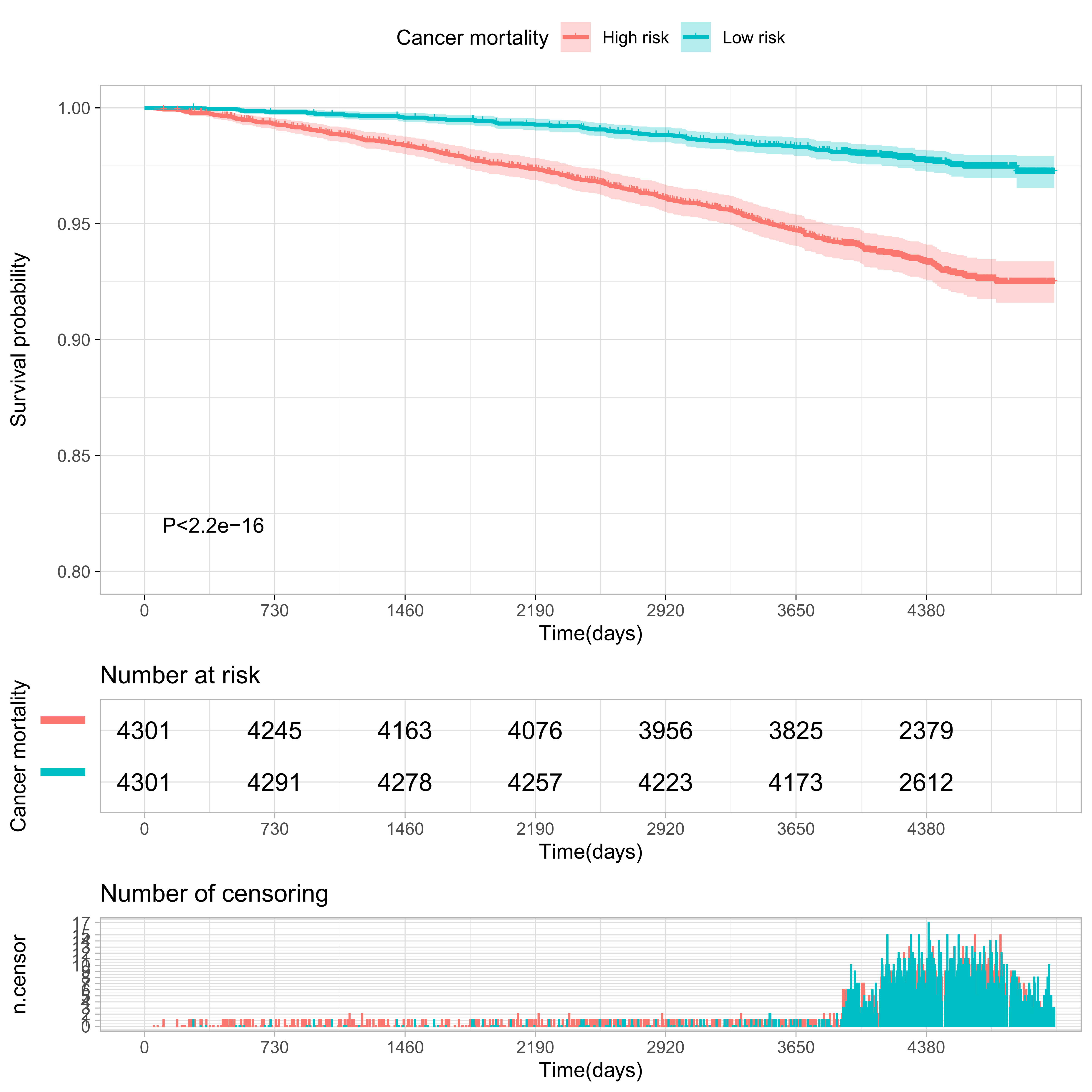

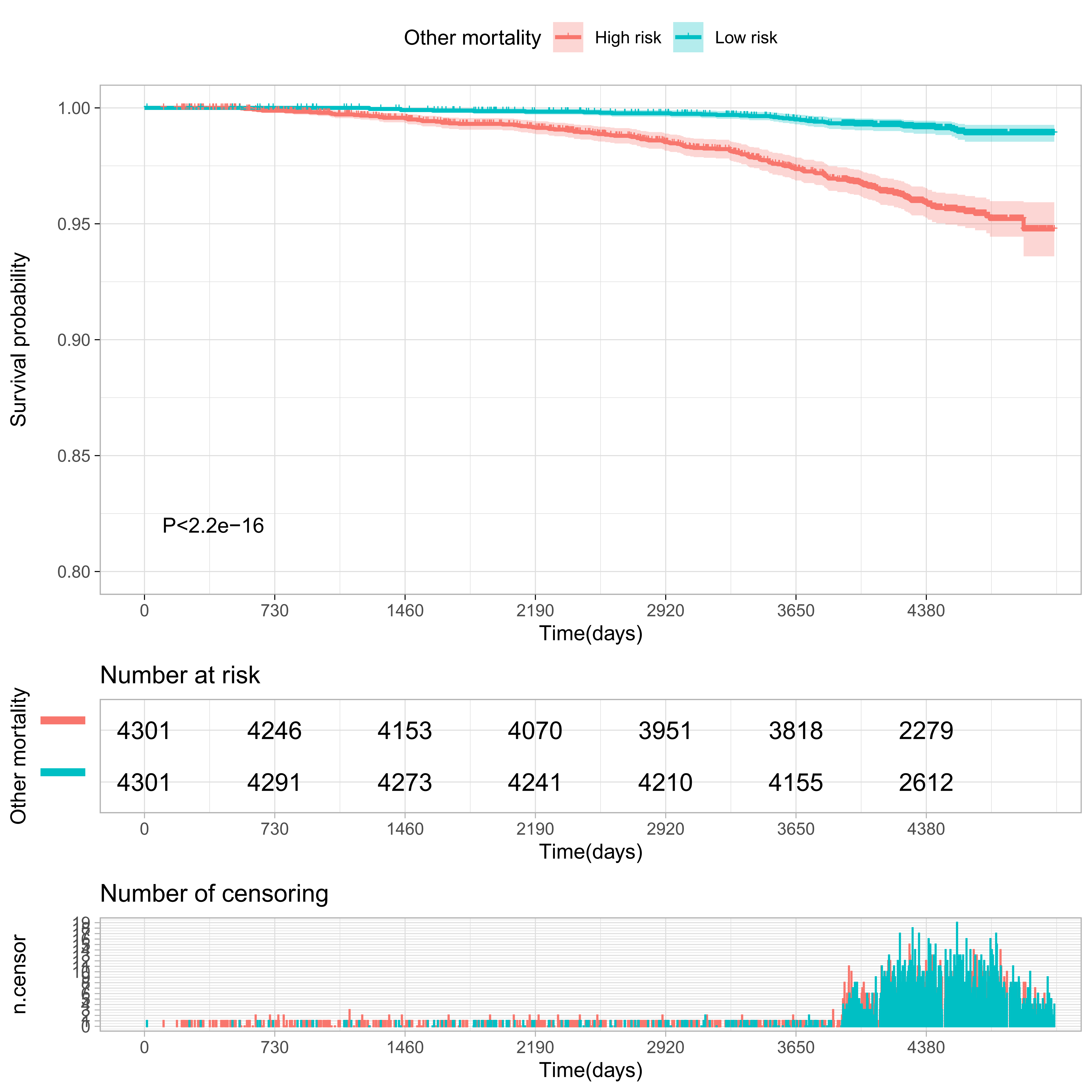

#### Supplementary Figure S6

**Comparisons of morbidity of systemic diseases in the top and bottom 10% of the meta-GCIPL score in validation set.**

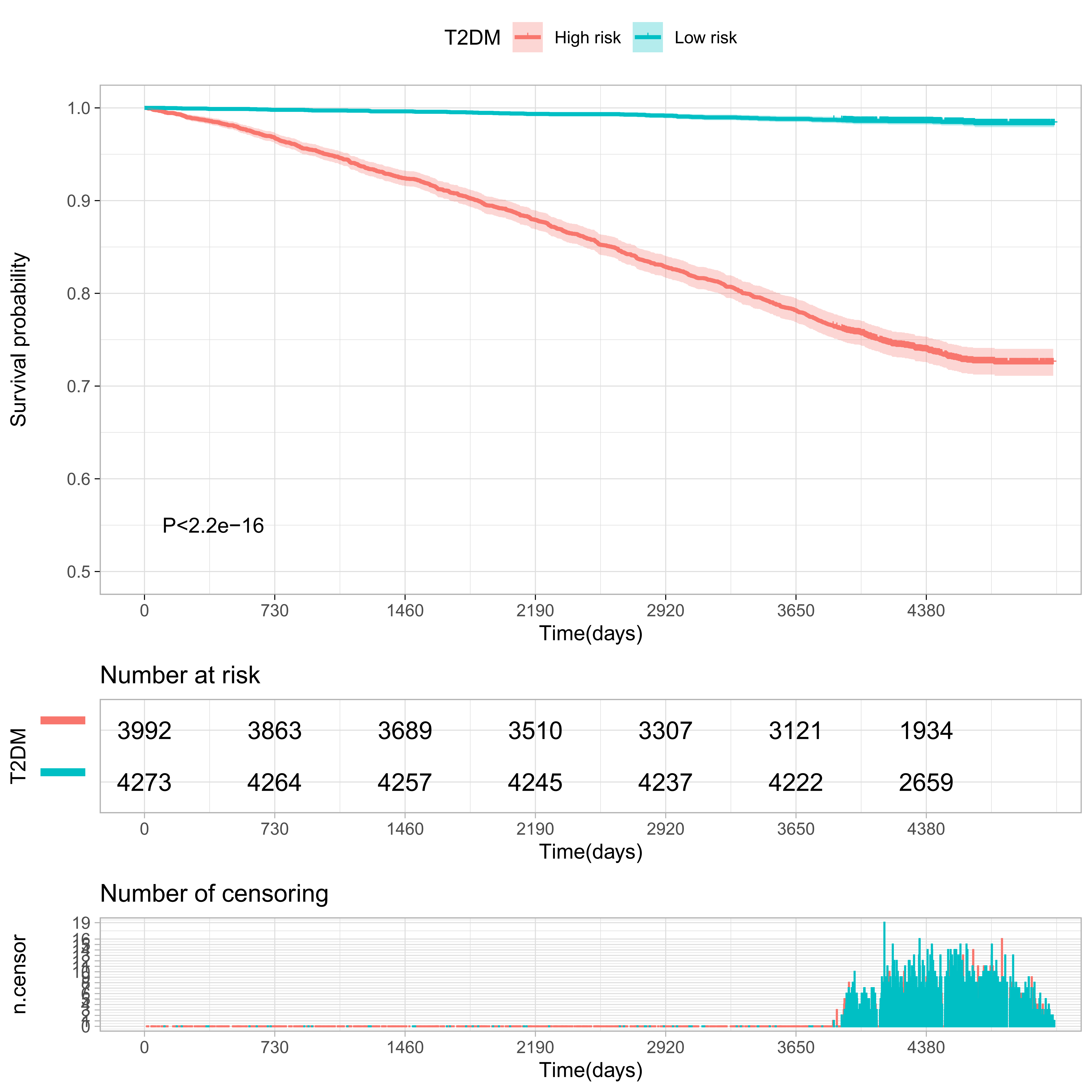

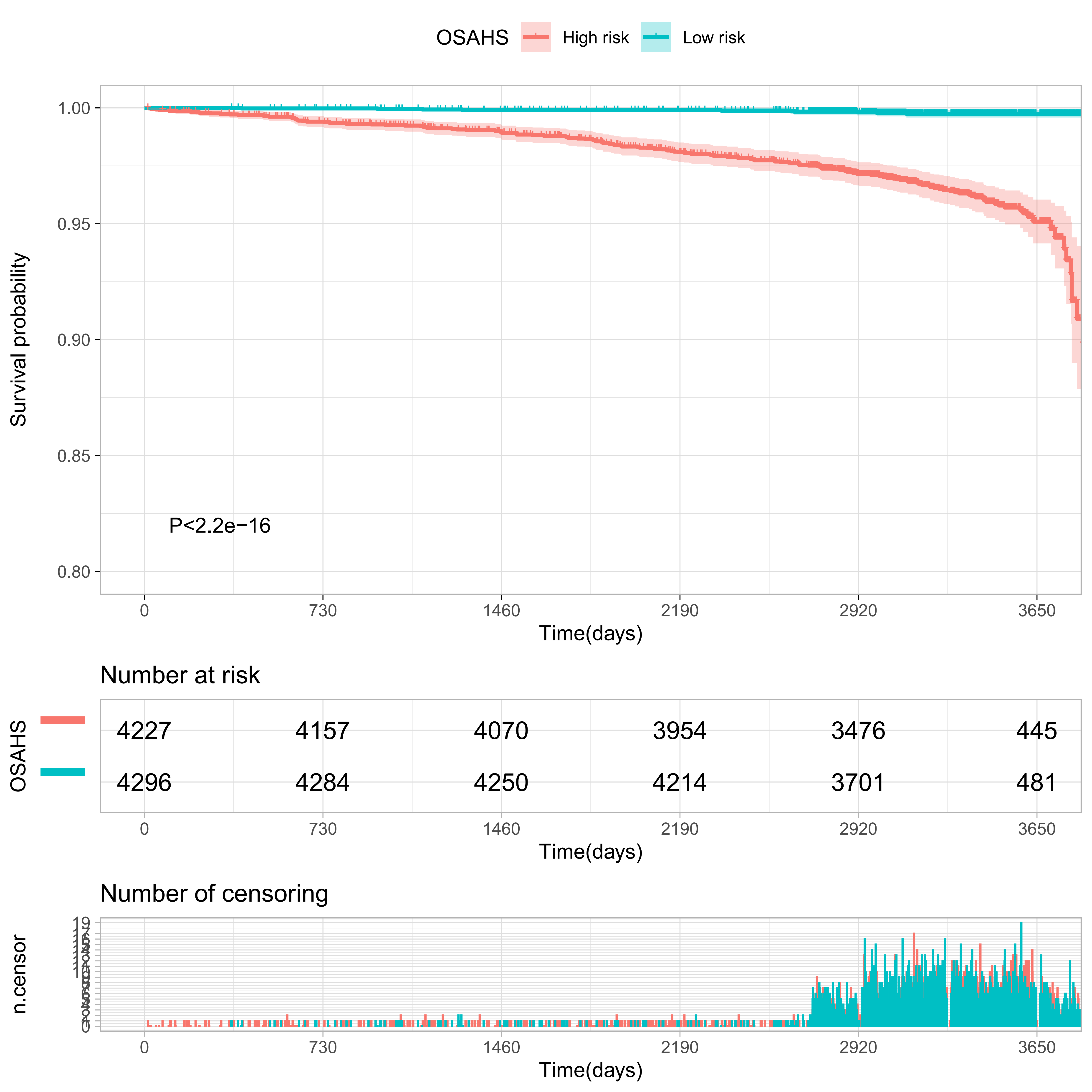

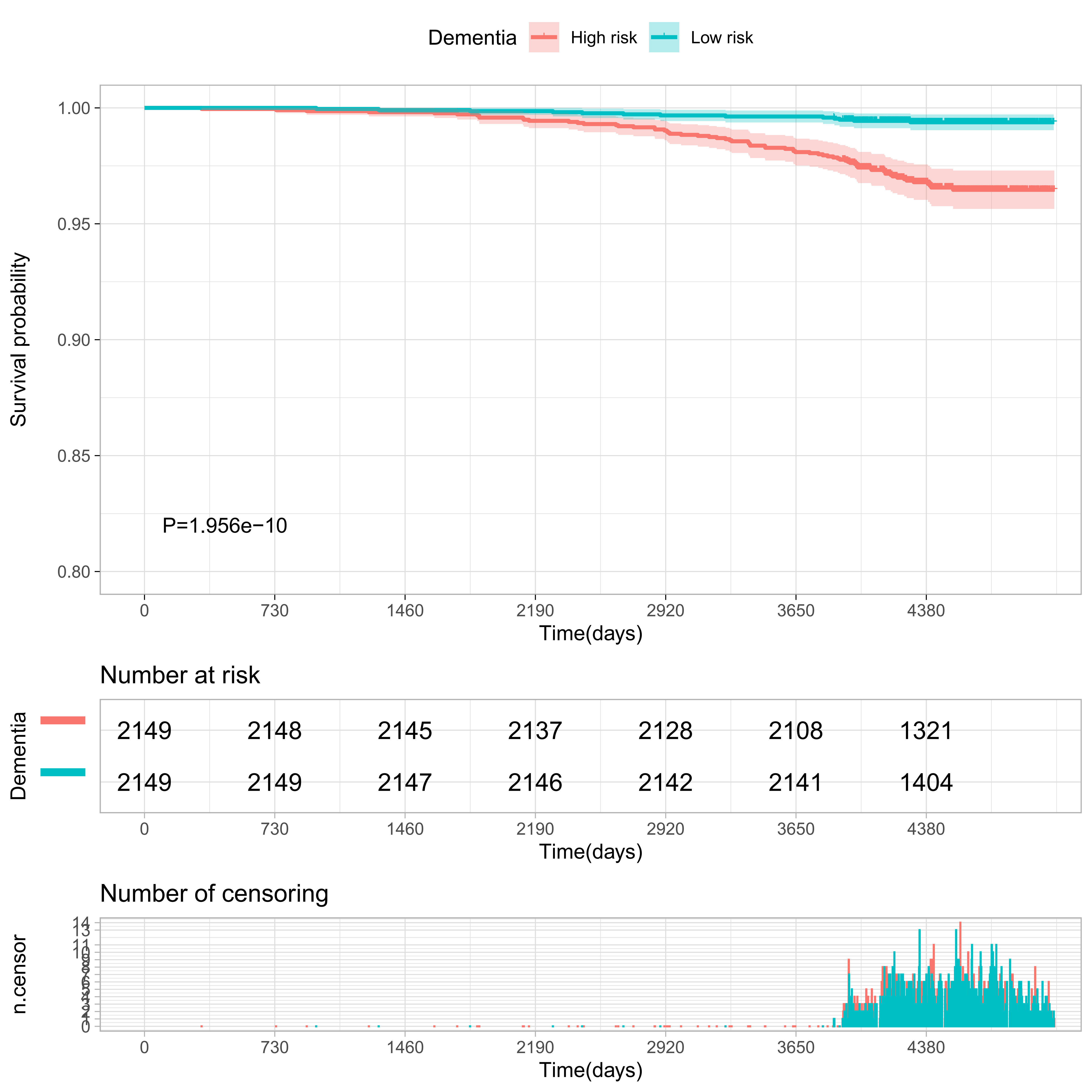

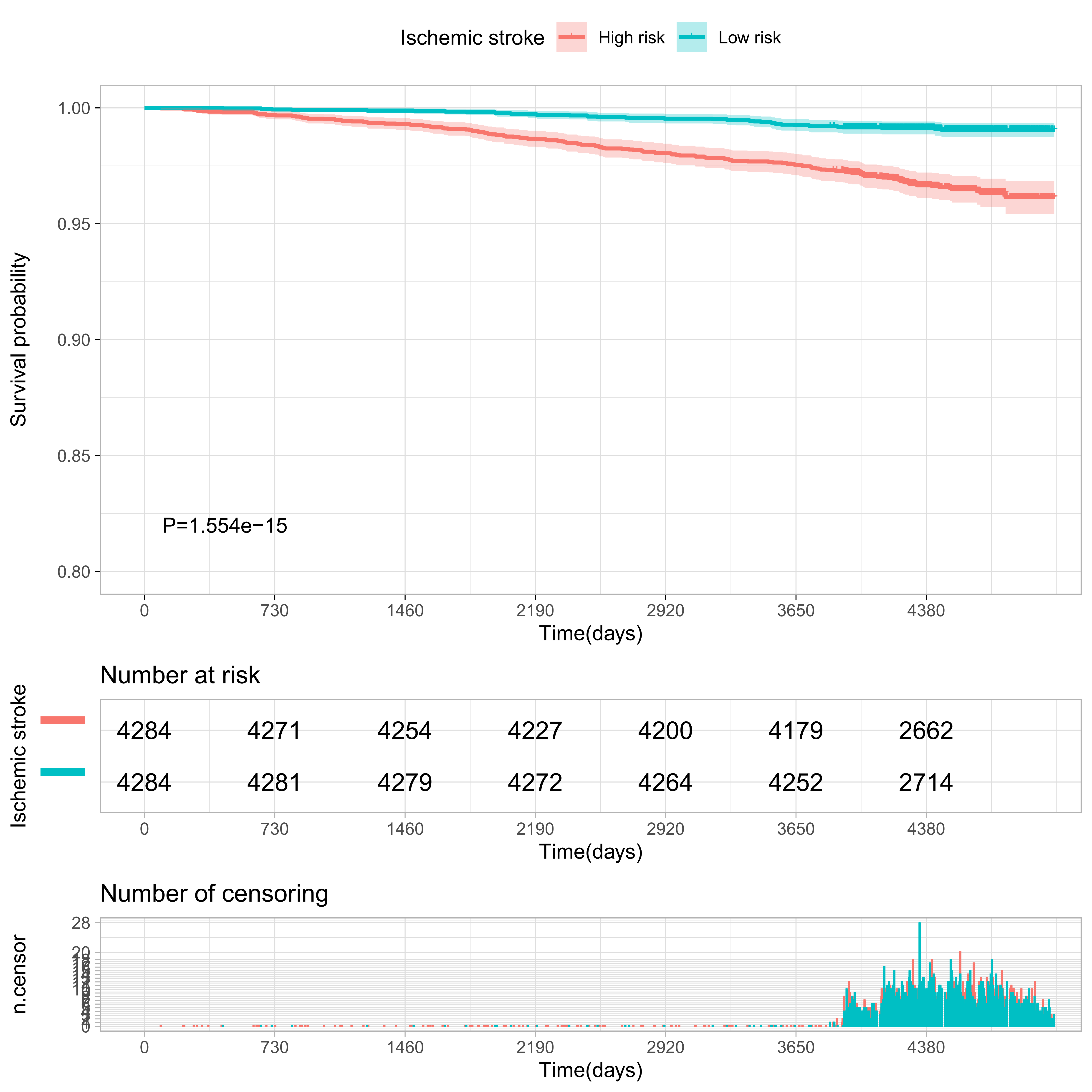

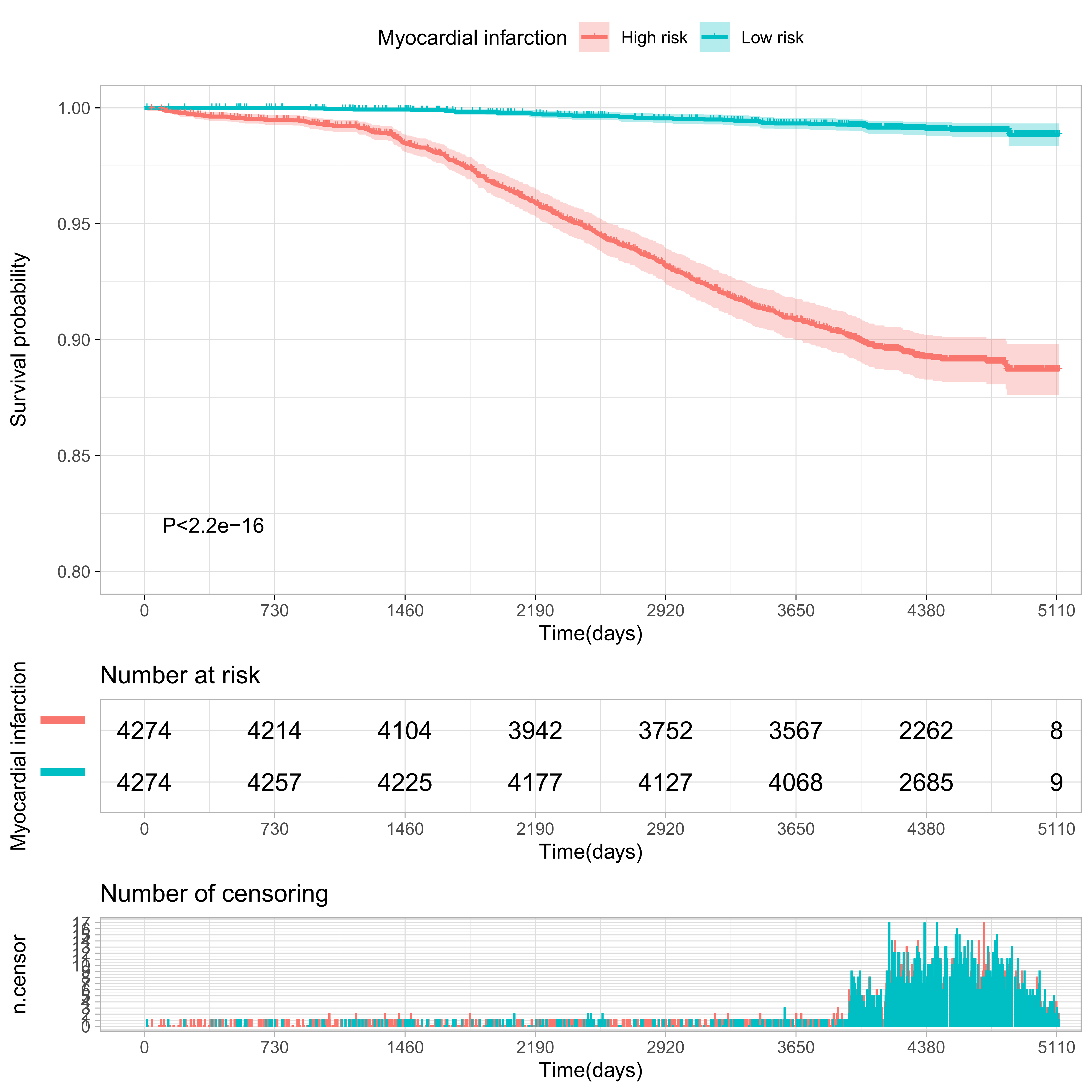

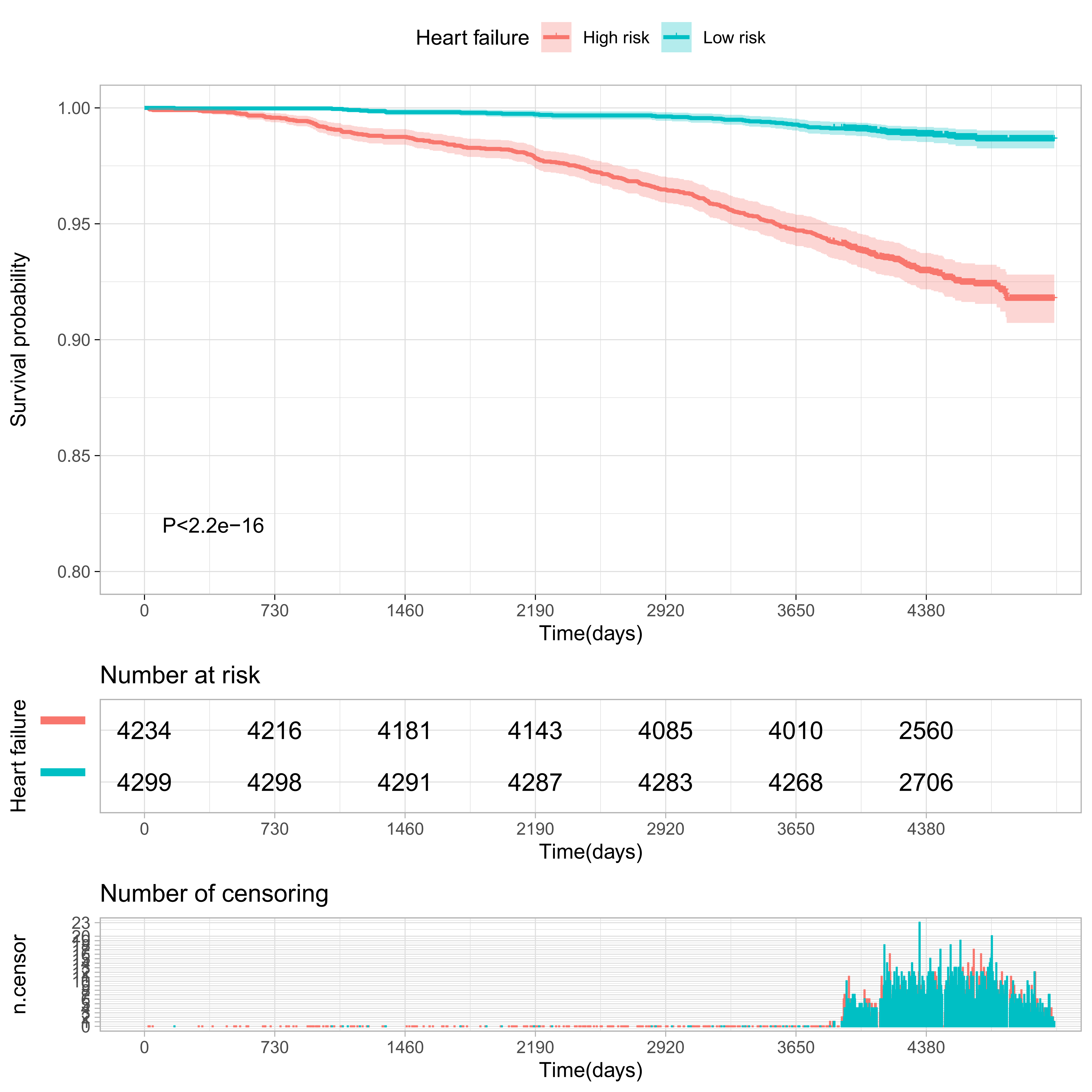

#### Supplementary Figure S7

**Comparisons of mortality in the top and bottom 10% of the meta-GCIPL score in validation set.**

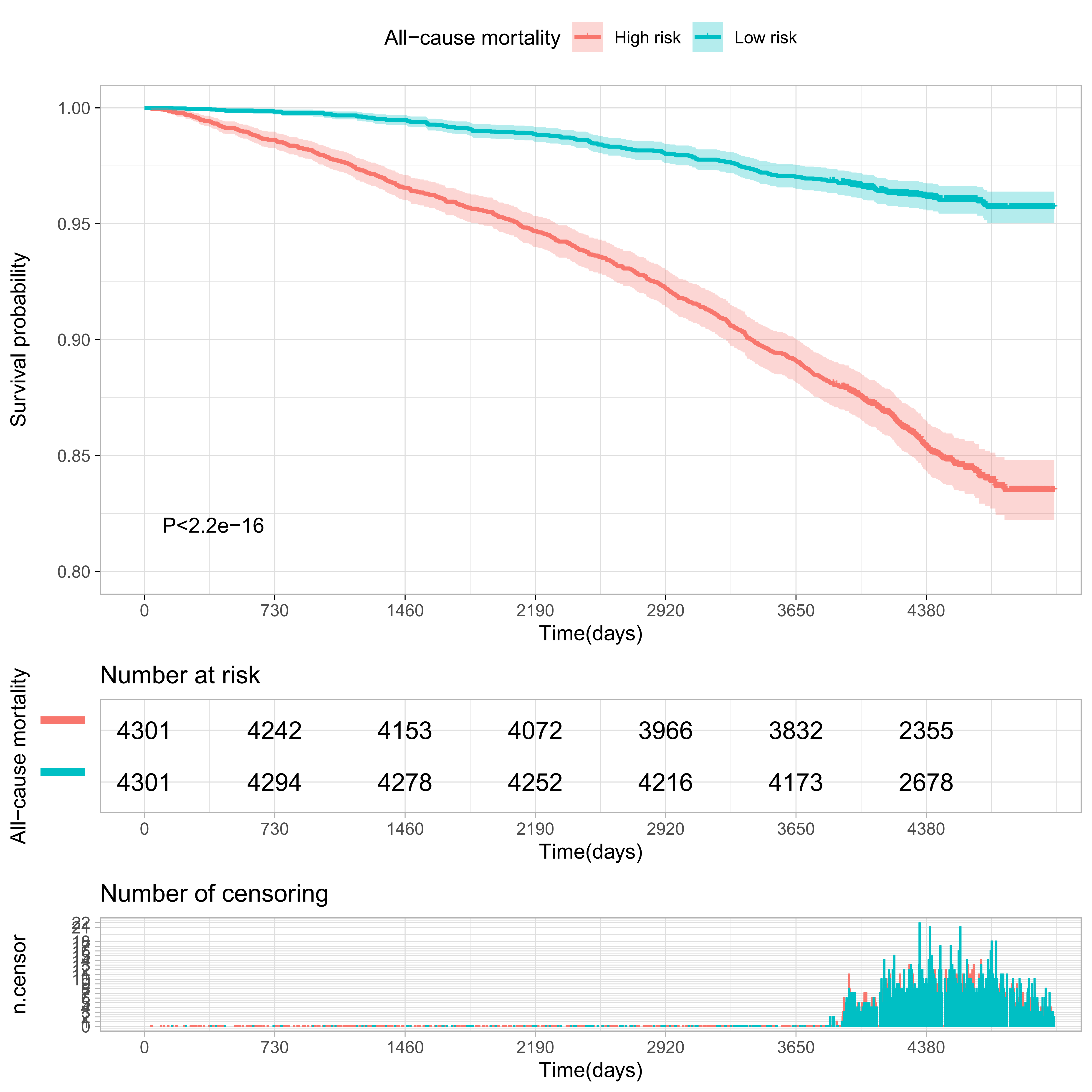

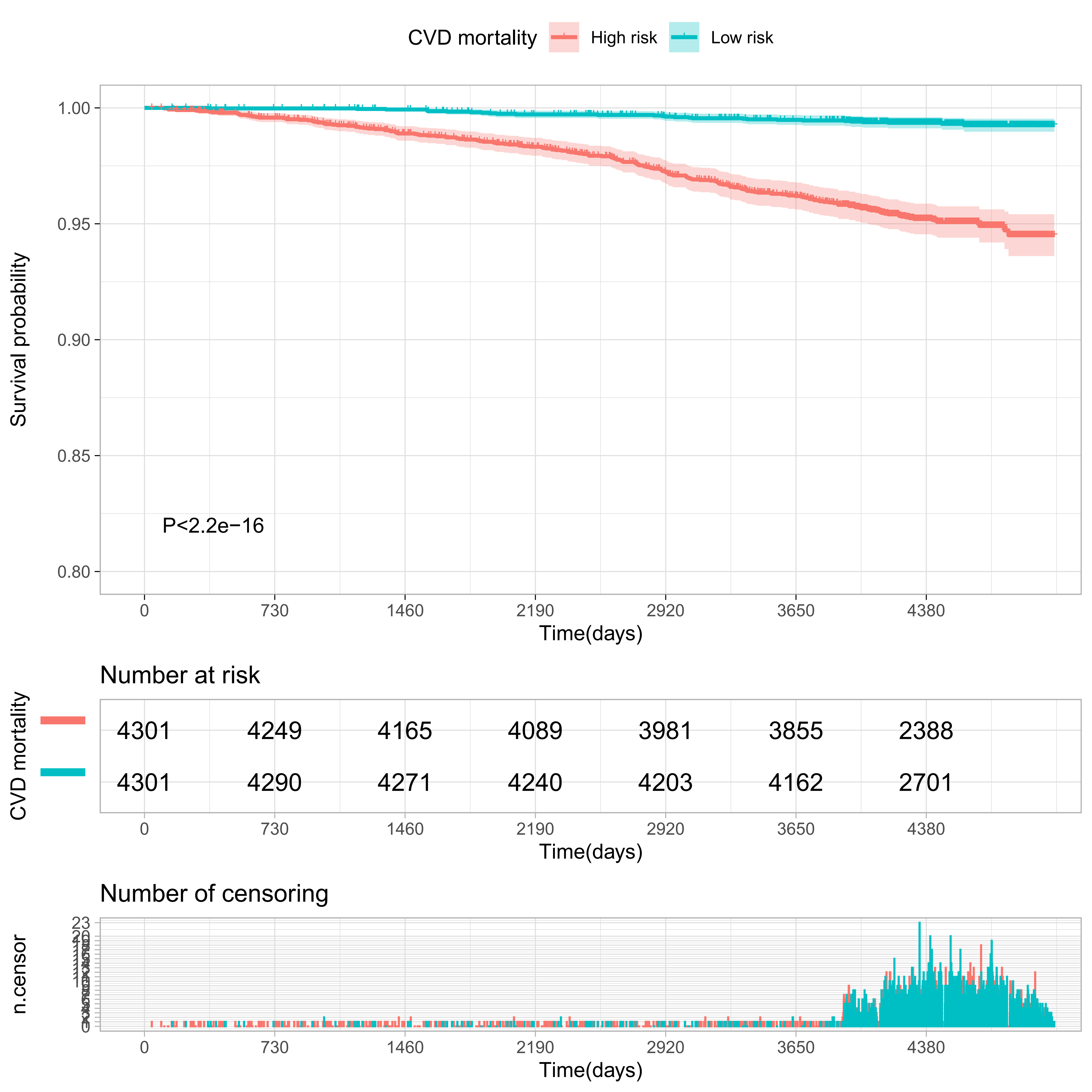

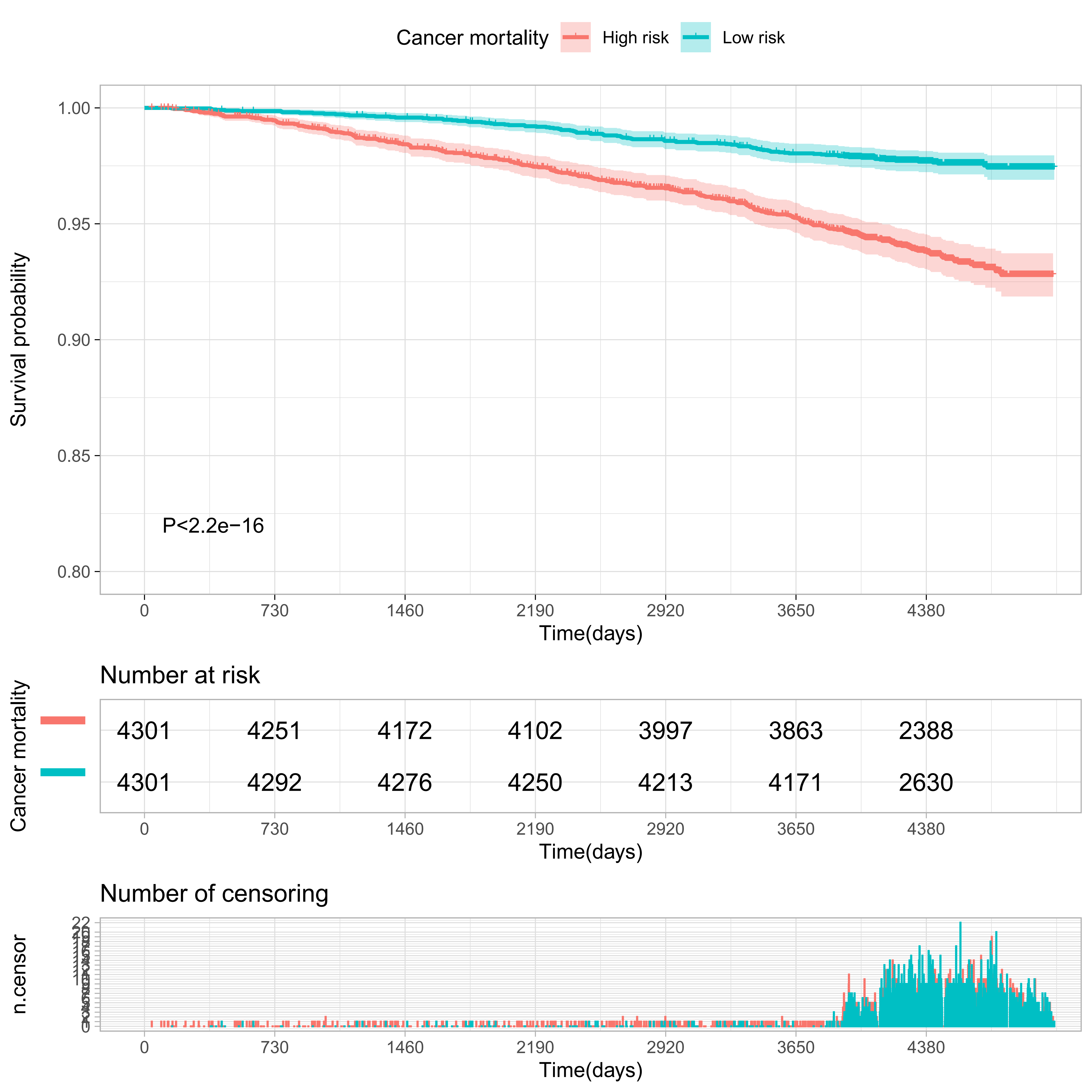

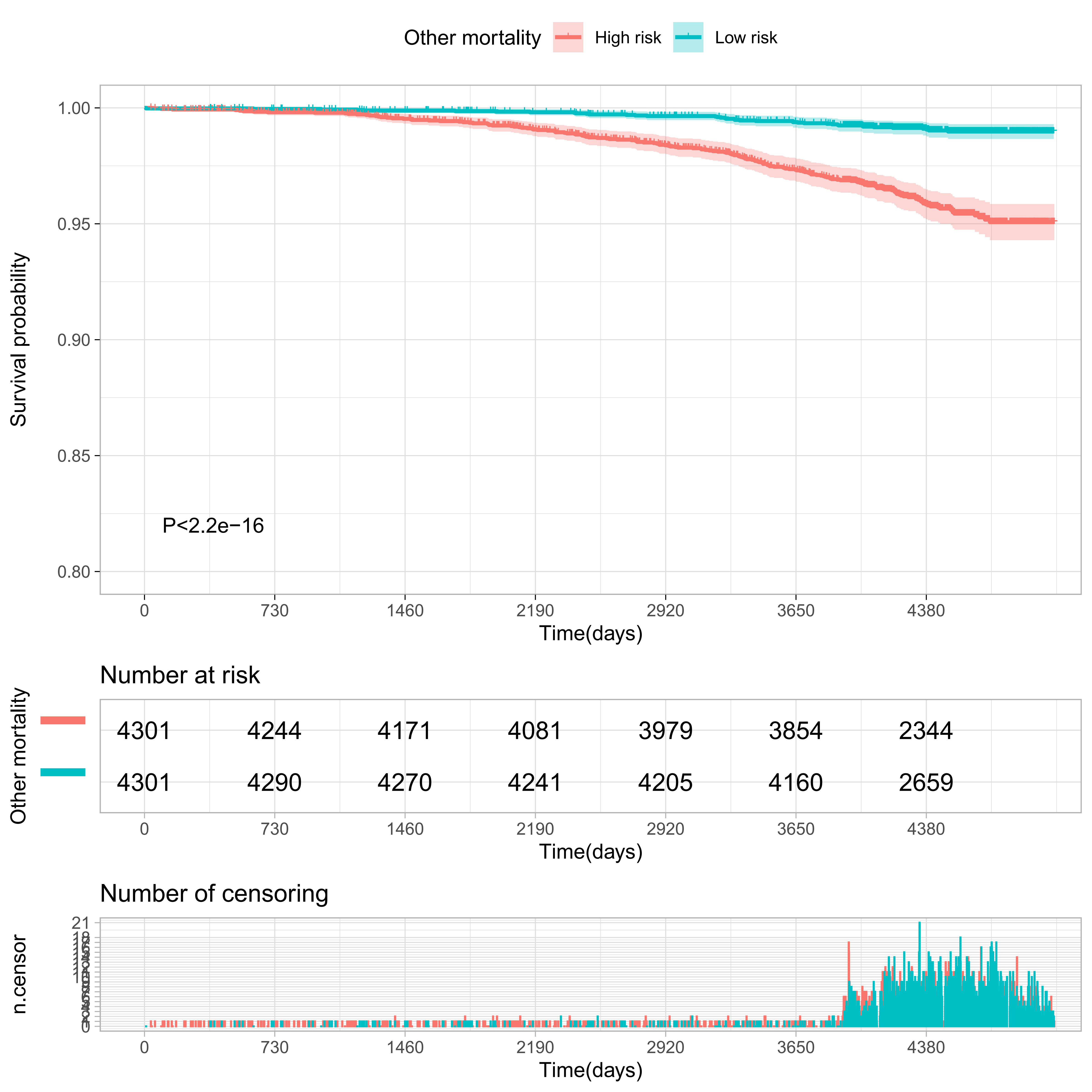

#### Supplementary Figure S8

**Receiver operating characteristic (ROC) curves of established risk factors and meta-GCIPLT score for predicting systemic diseases in validation set.**

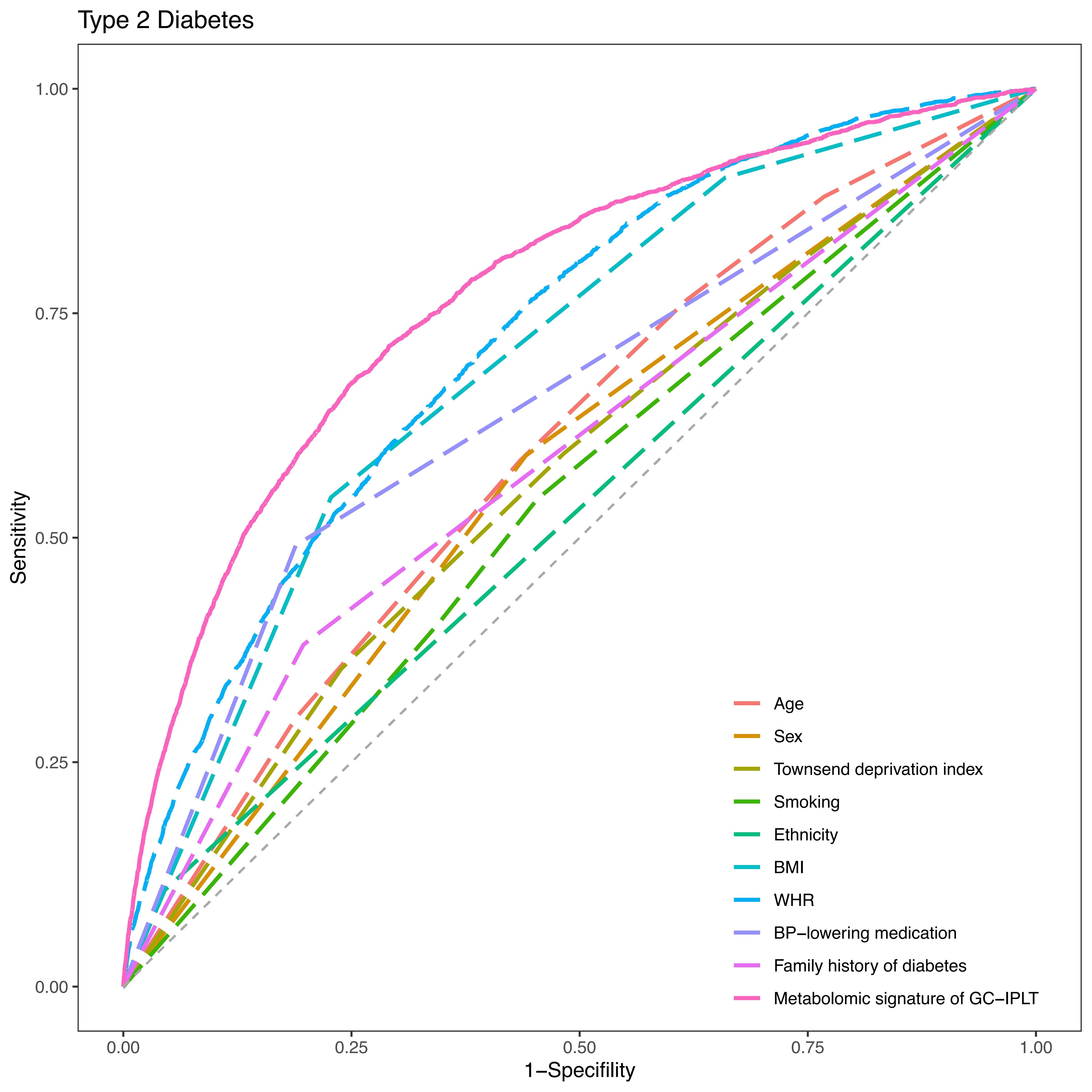

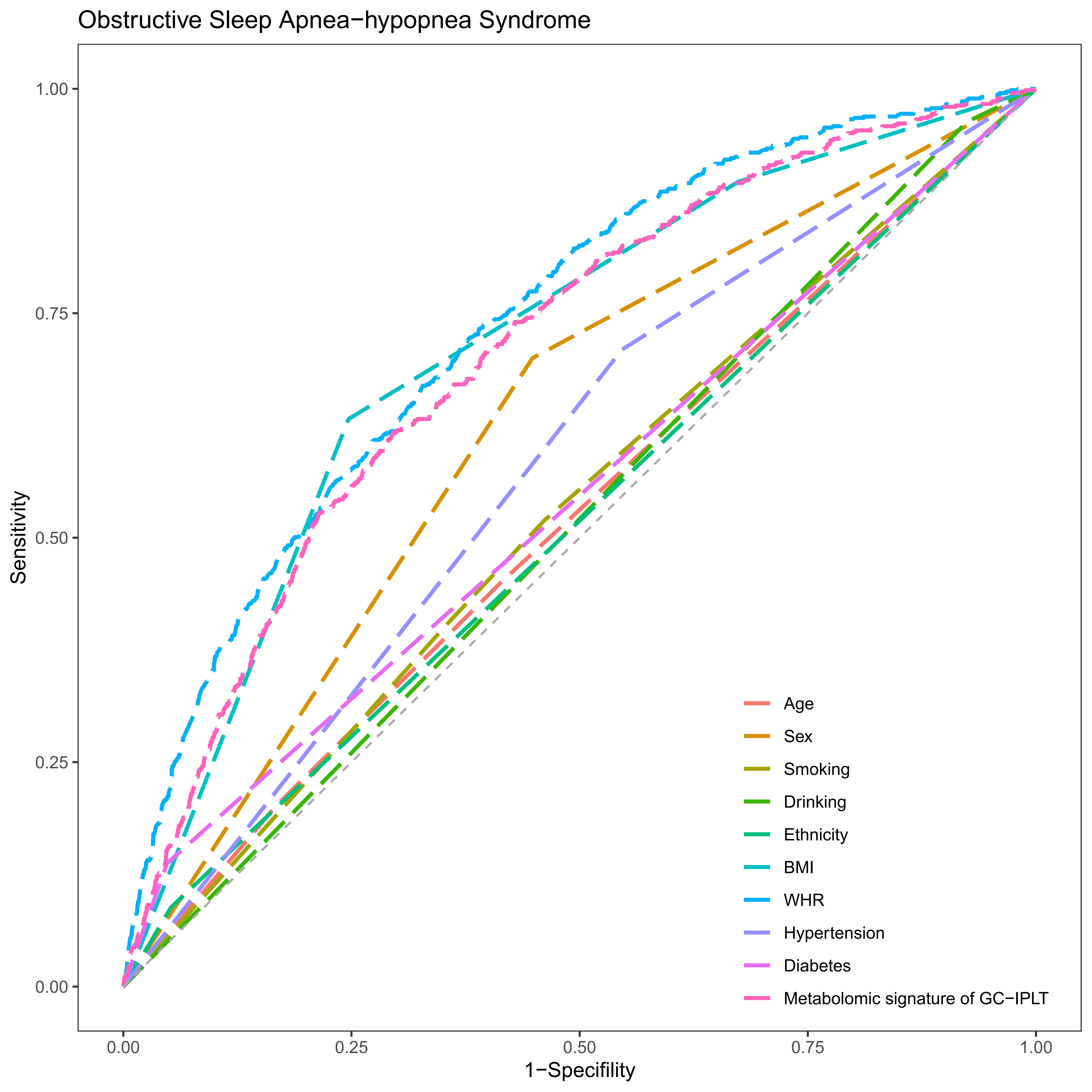

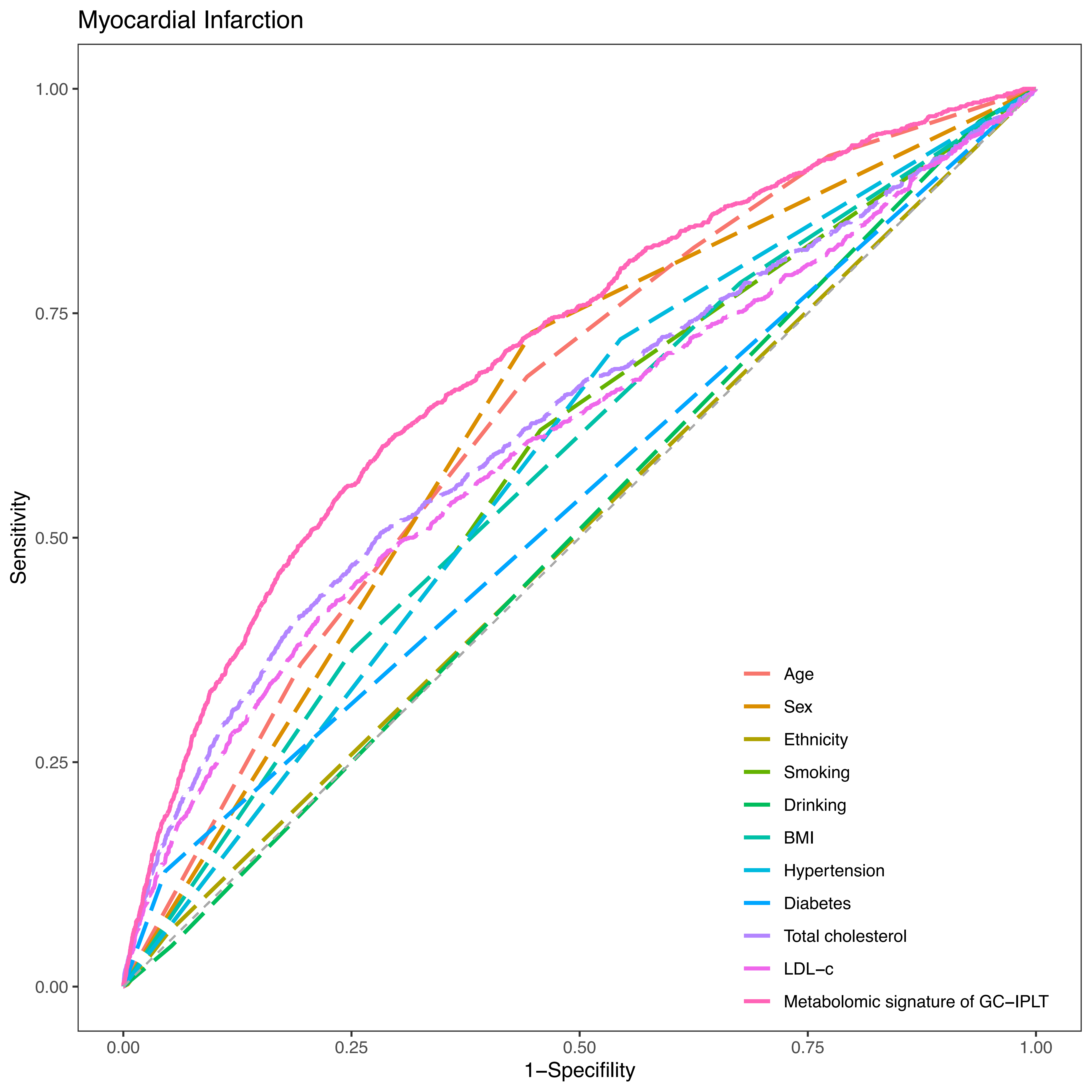

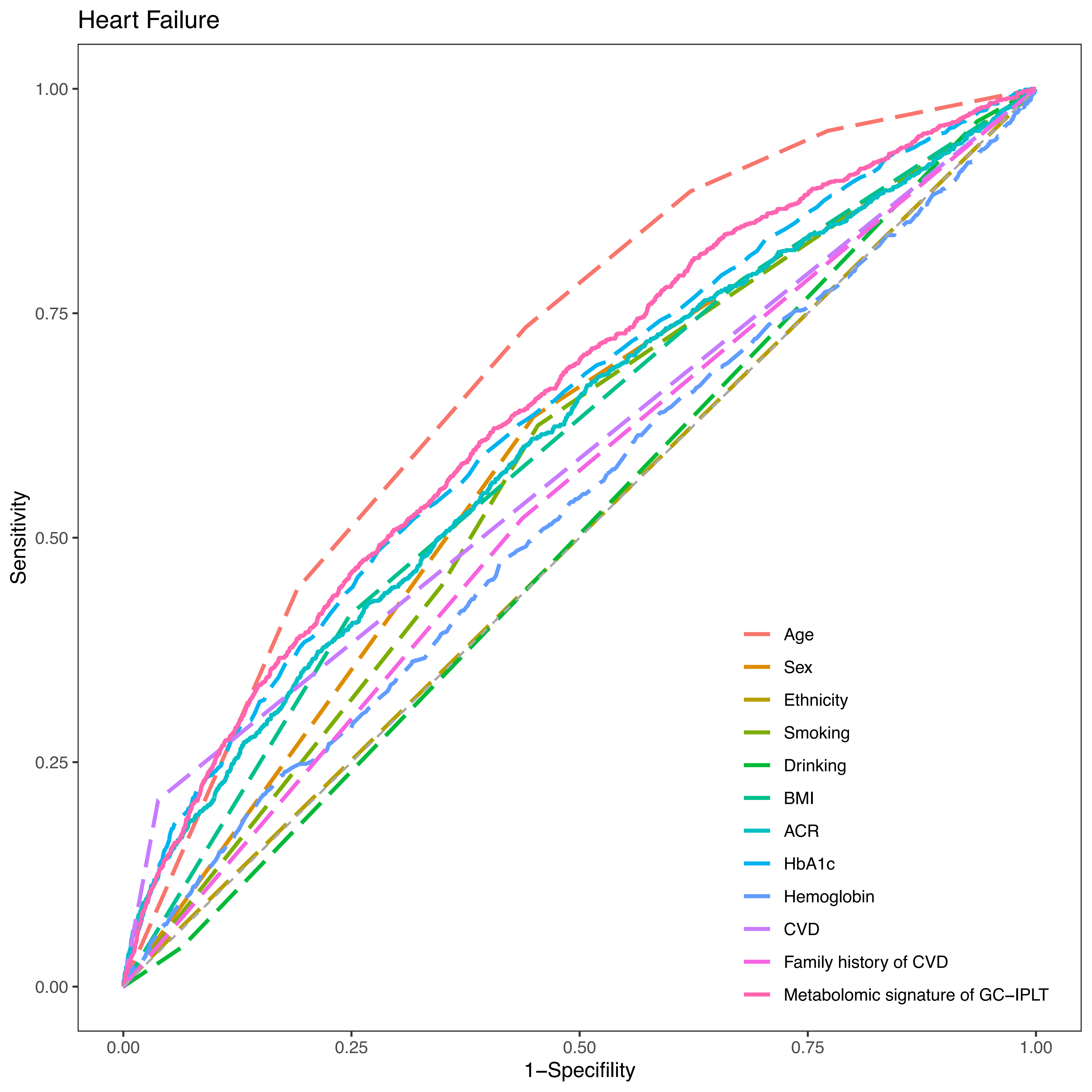

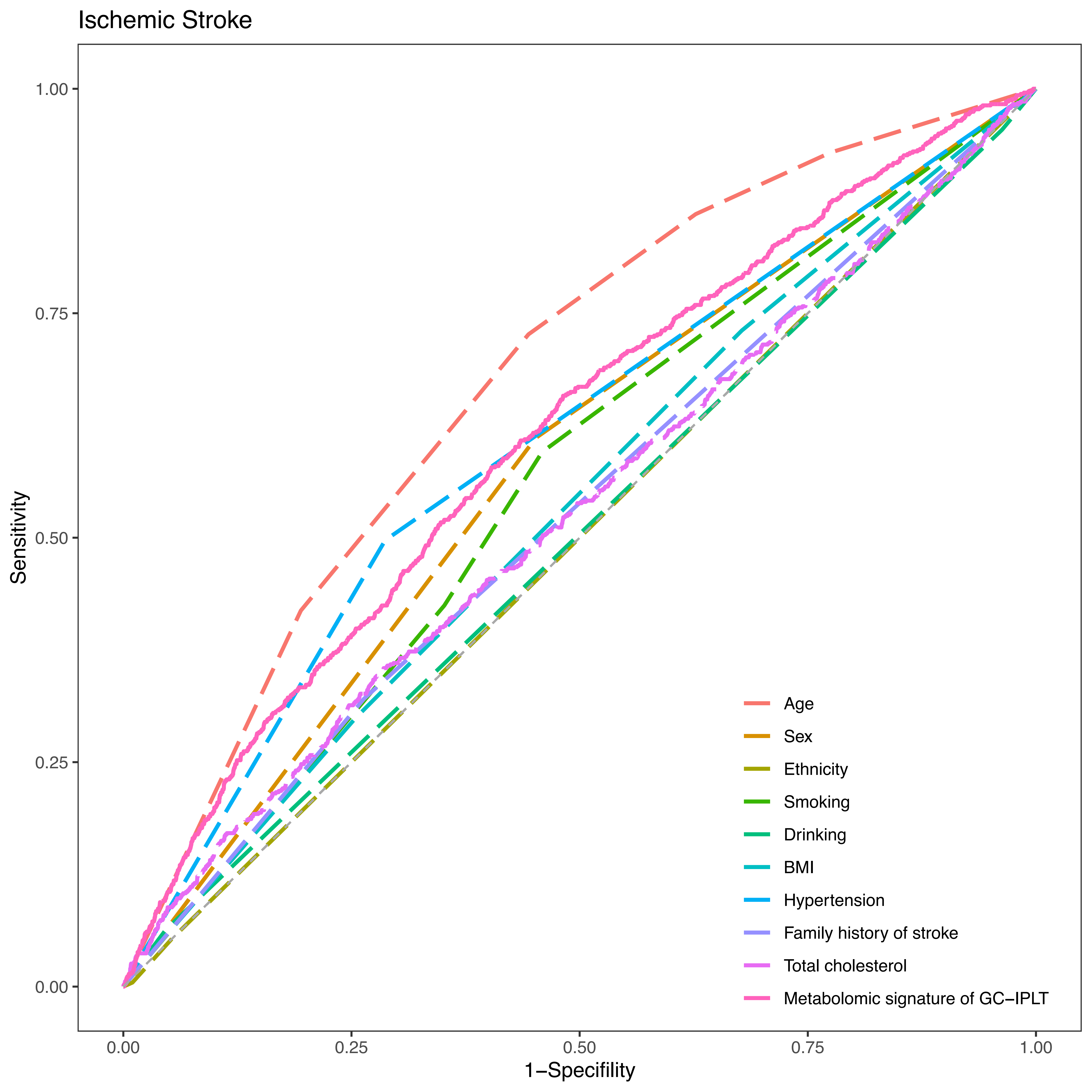

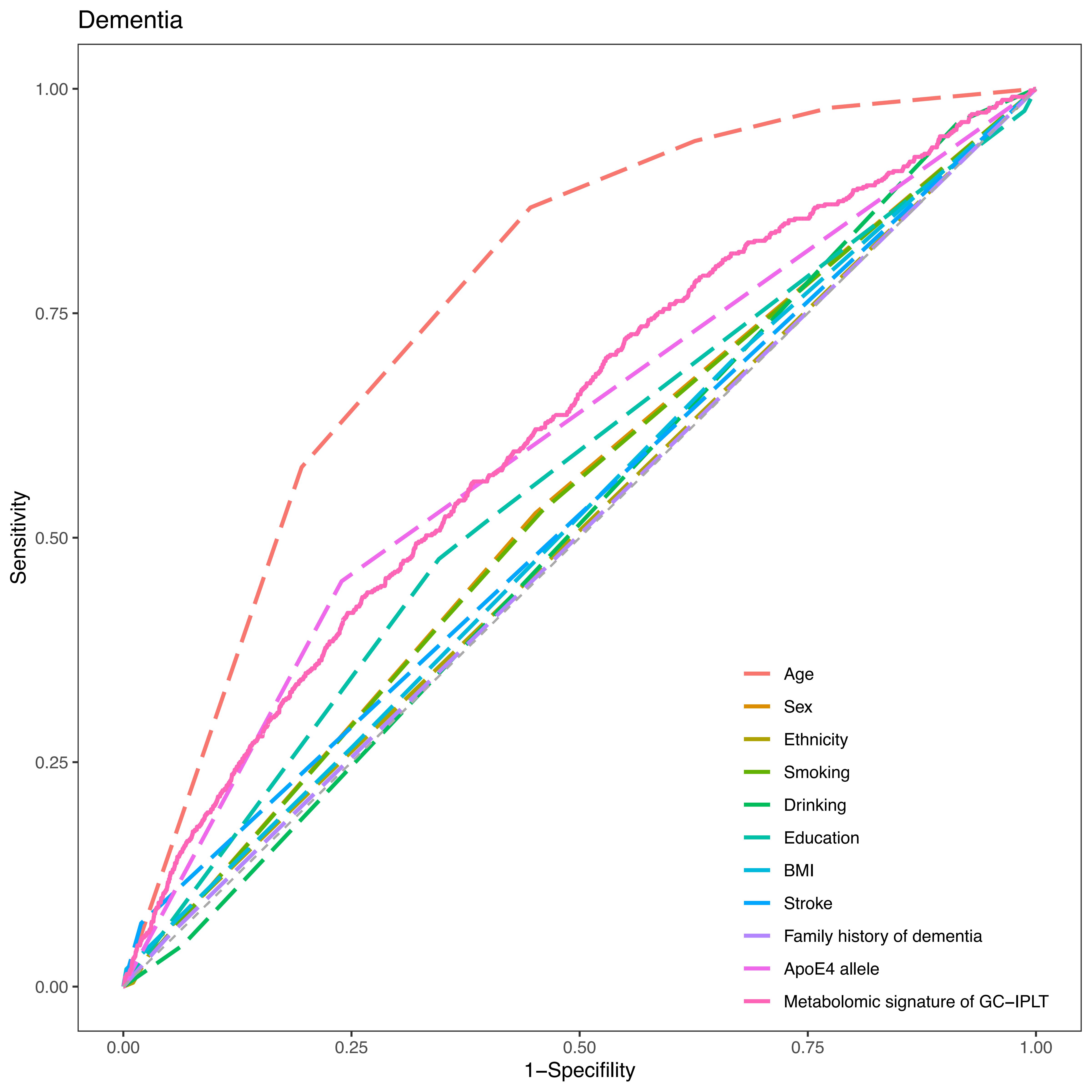

#### Supplementary Figure S9

**Receiver operating characteristic (ROC) curves of established risk factors and meta-GCIPLT score for predicting mortality in validation set.**

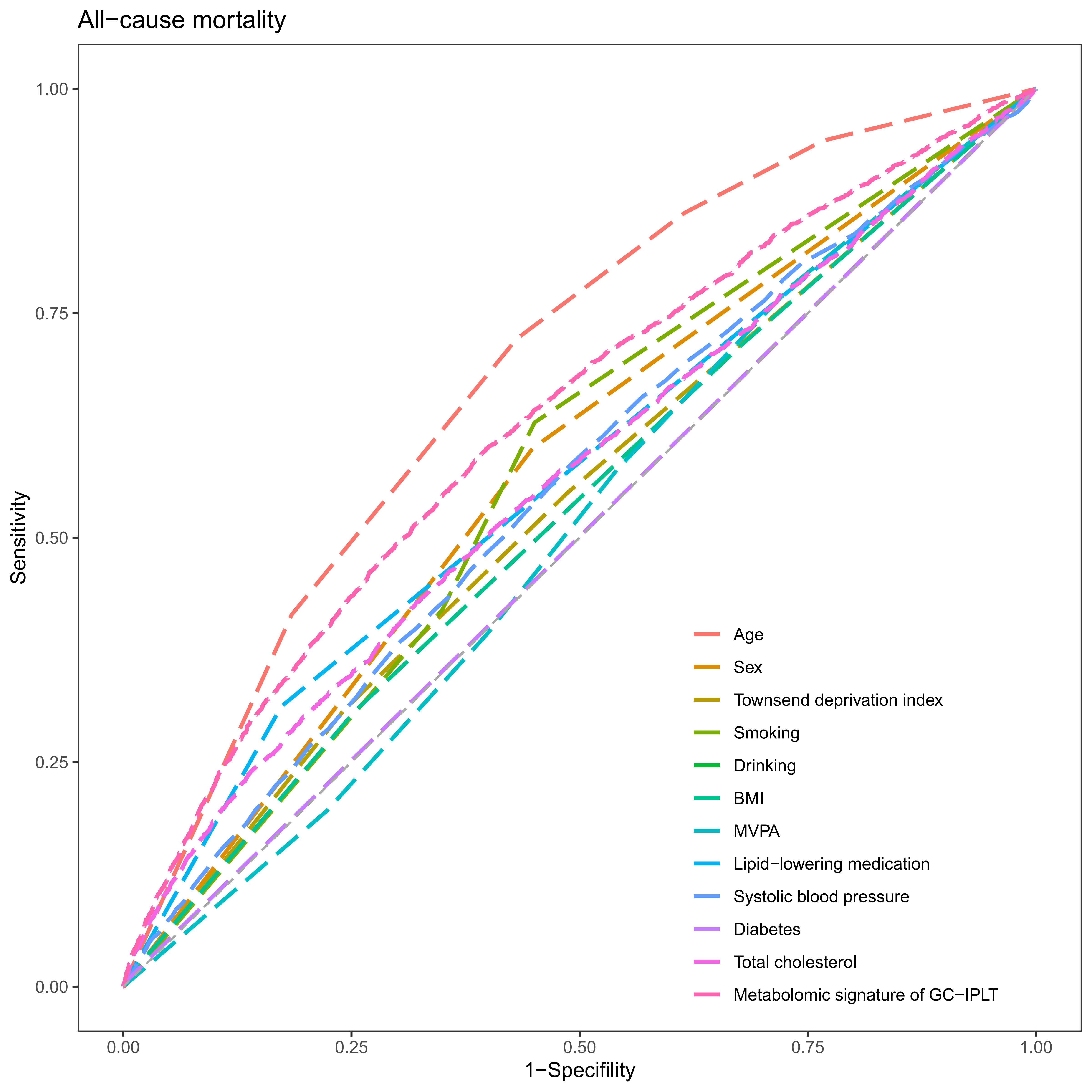

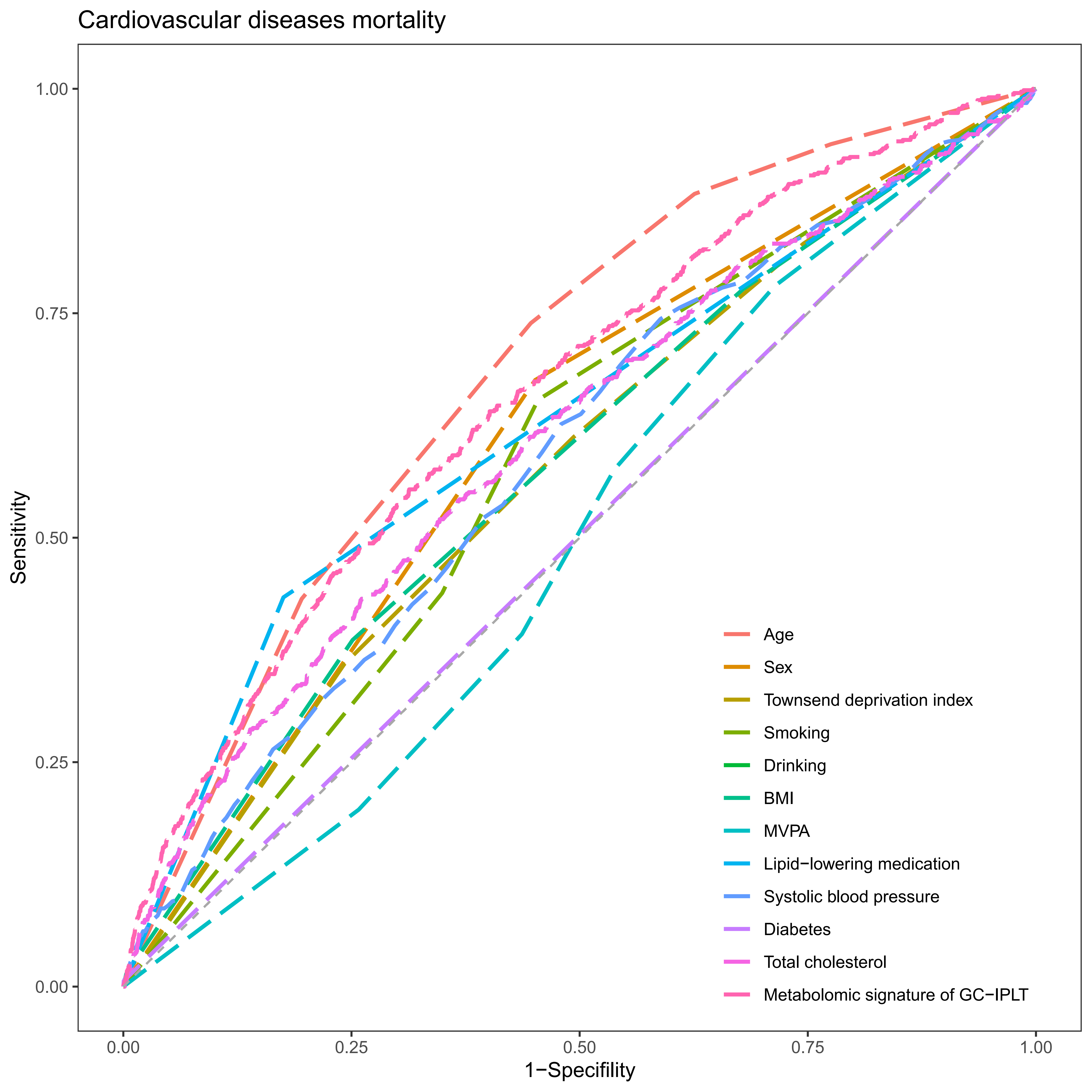

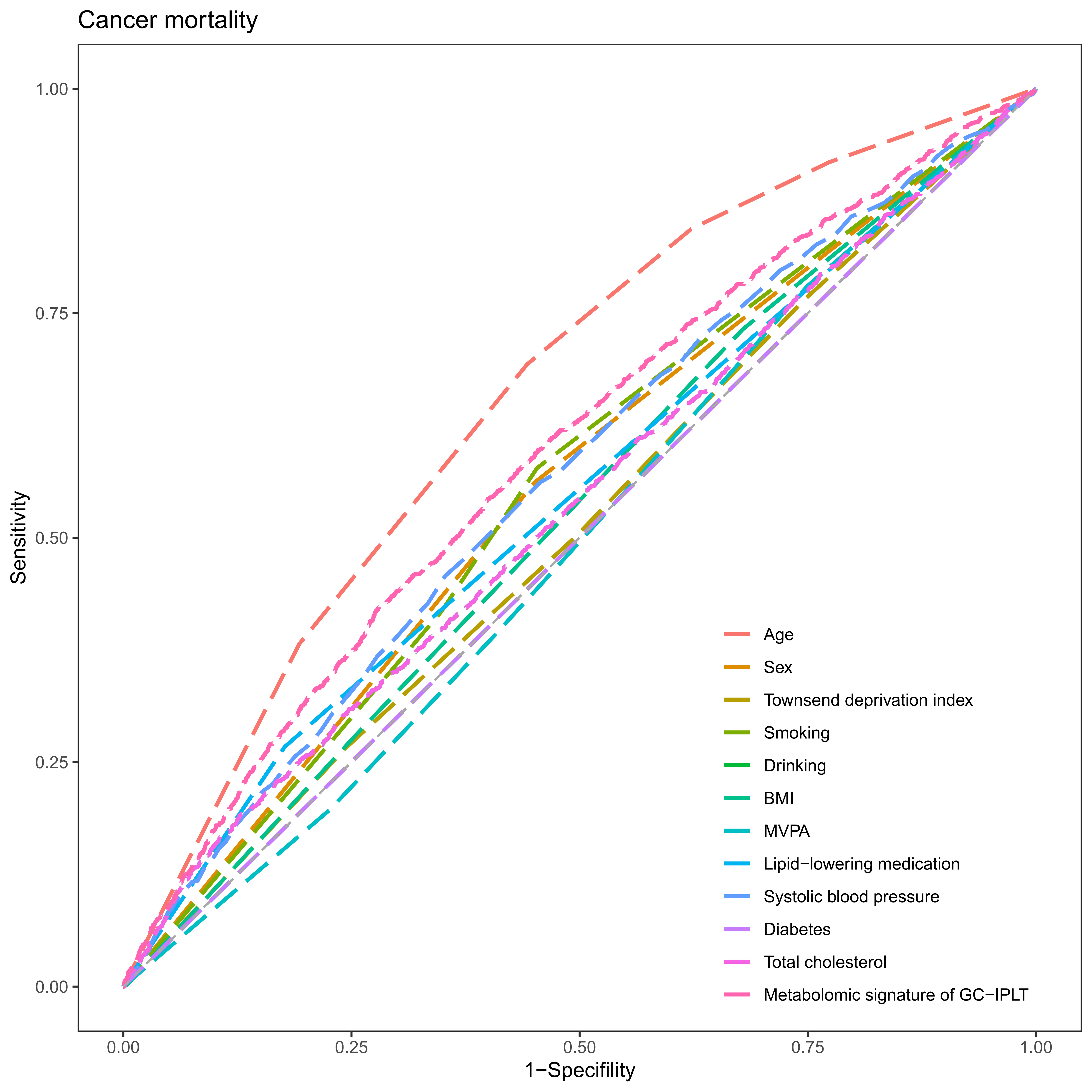

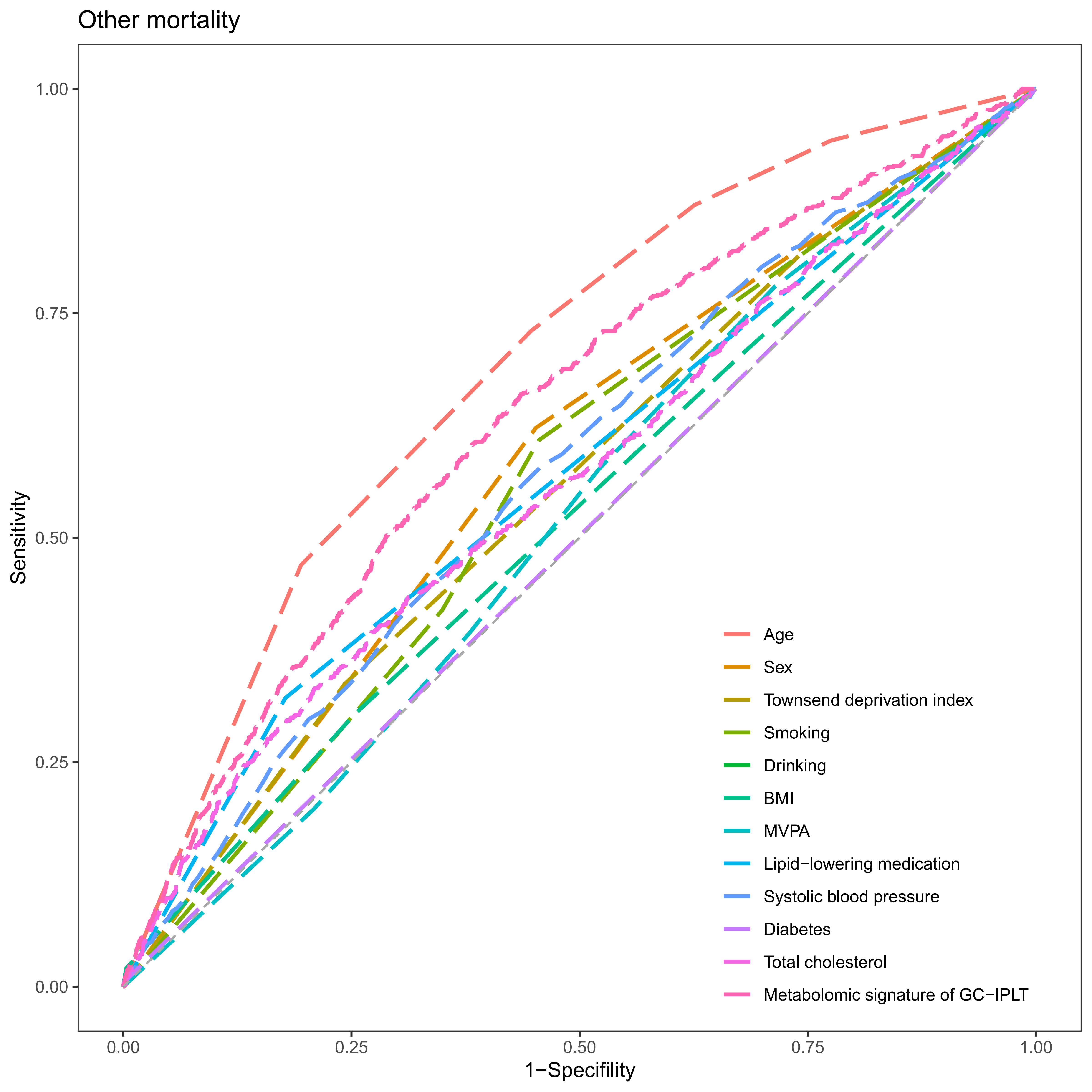

#### Supplementary Figure S10

**Receiver operating characteristic (ROC) curves of conventional model, meta-GCIPLT score, and combined model predicting systemic diseases in training set.**

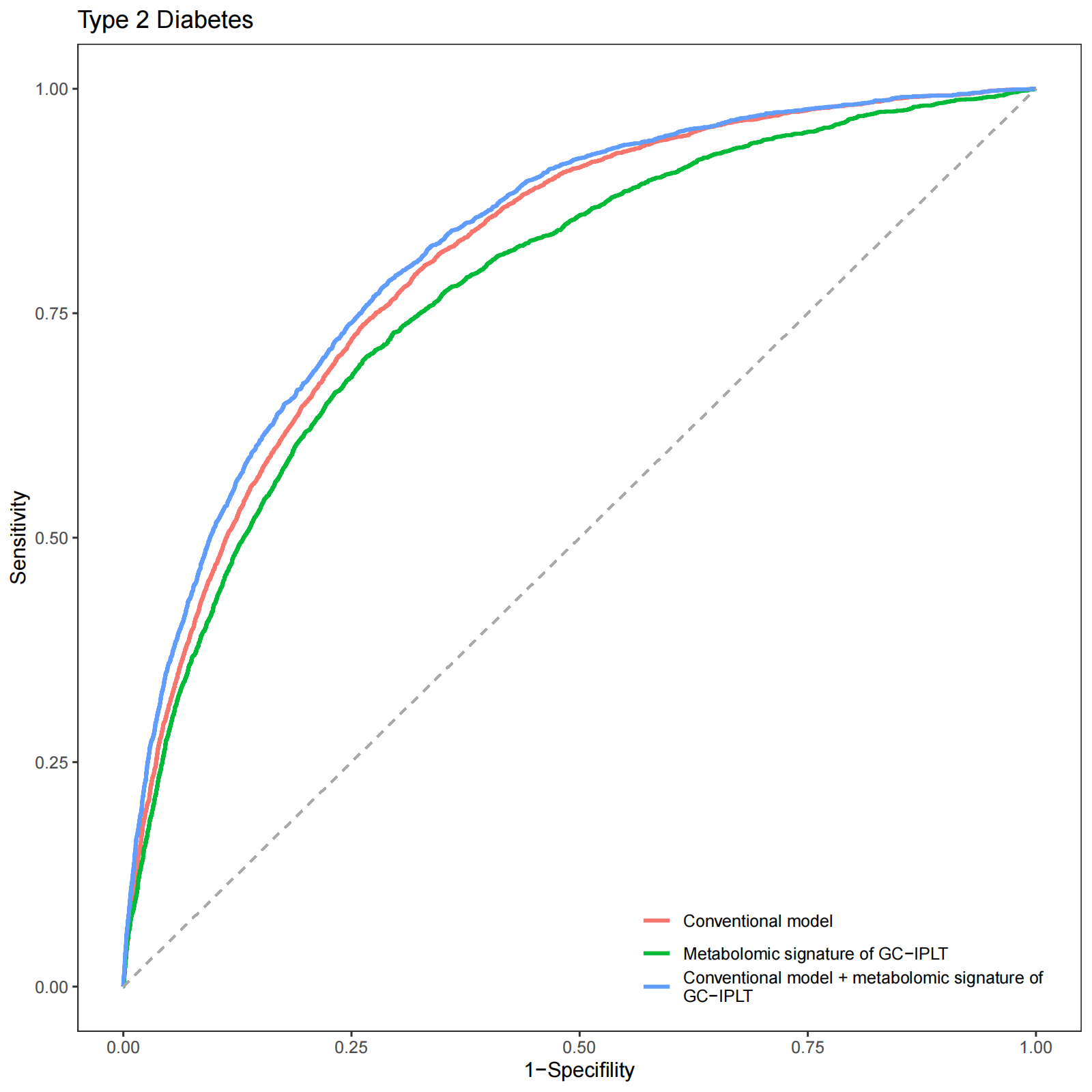

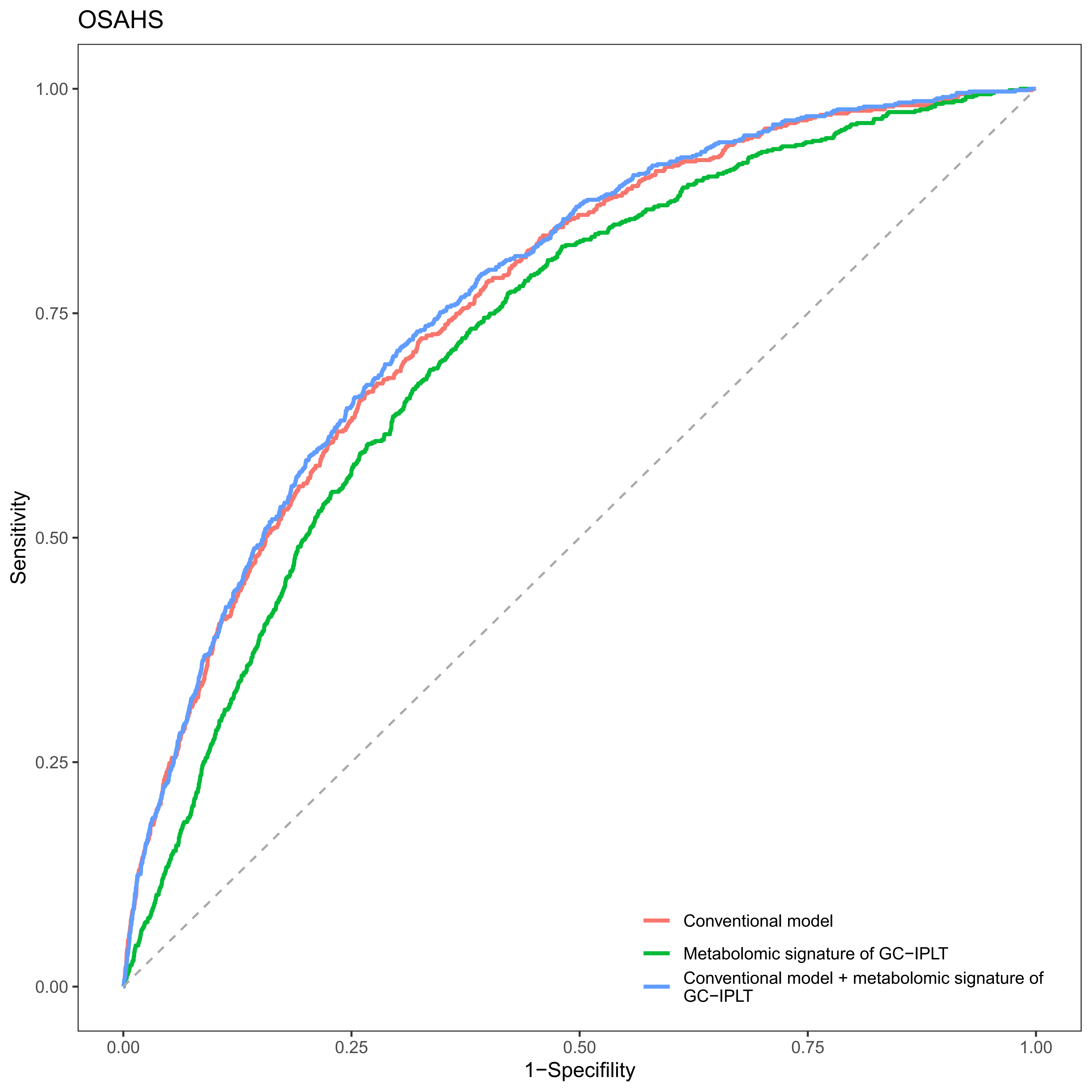

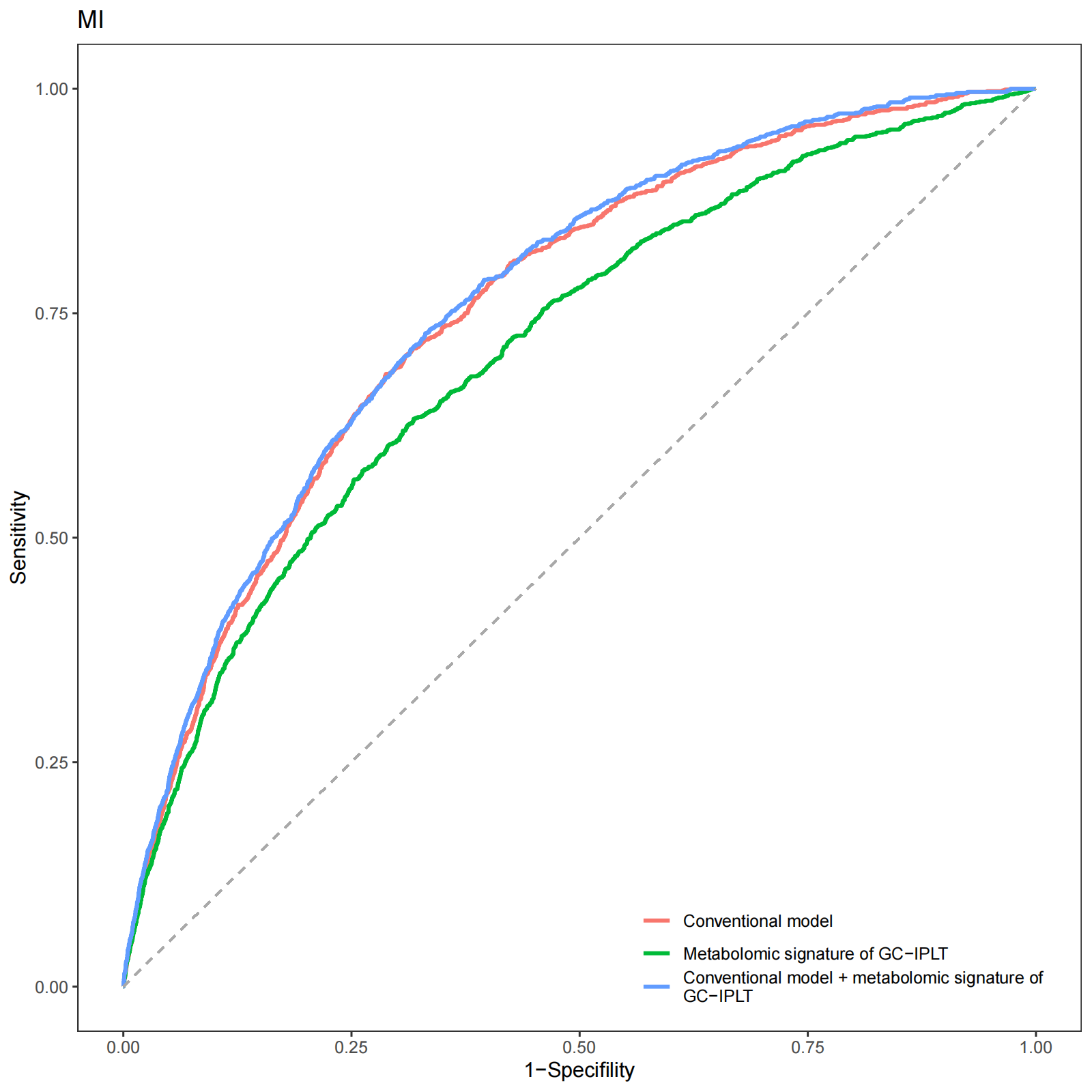

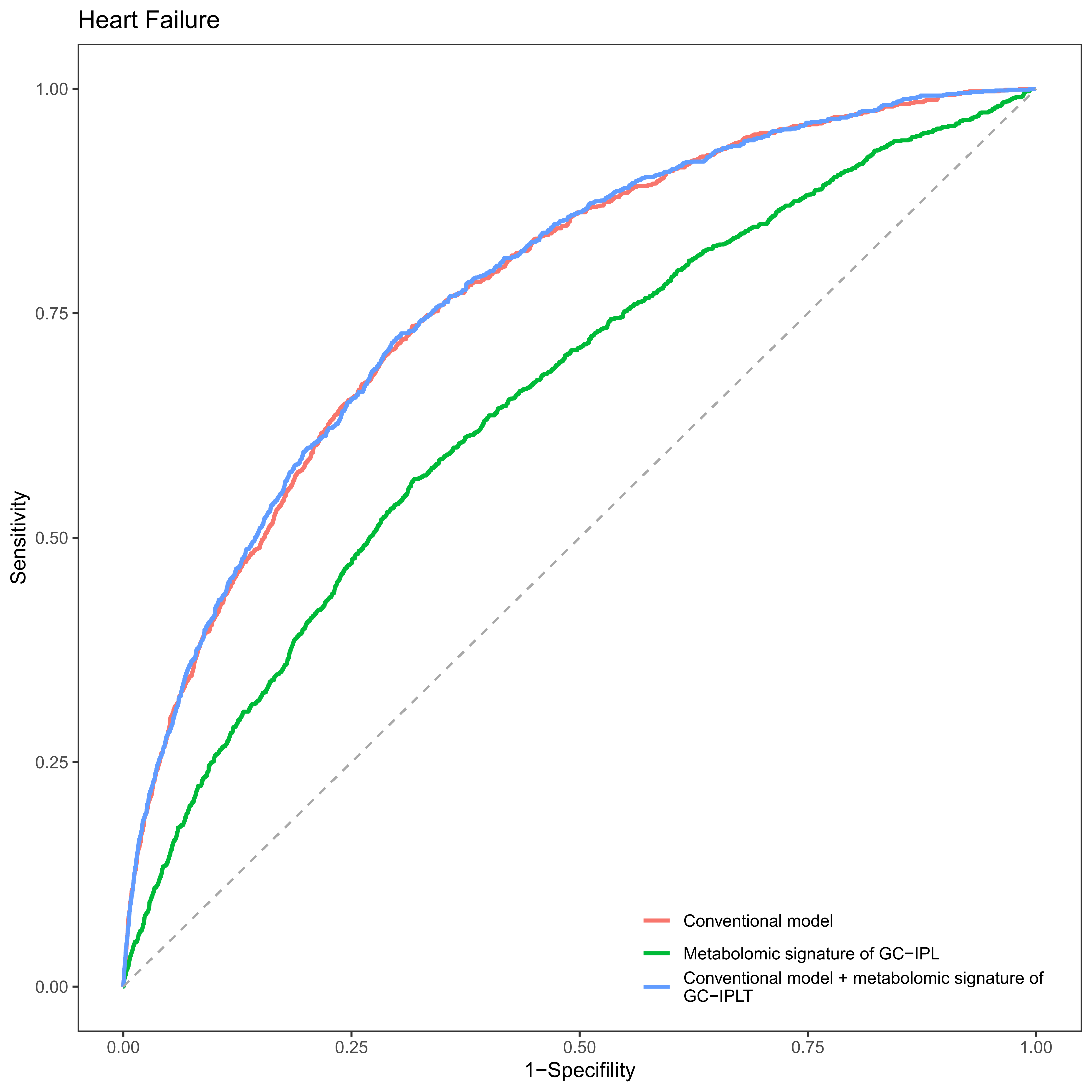

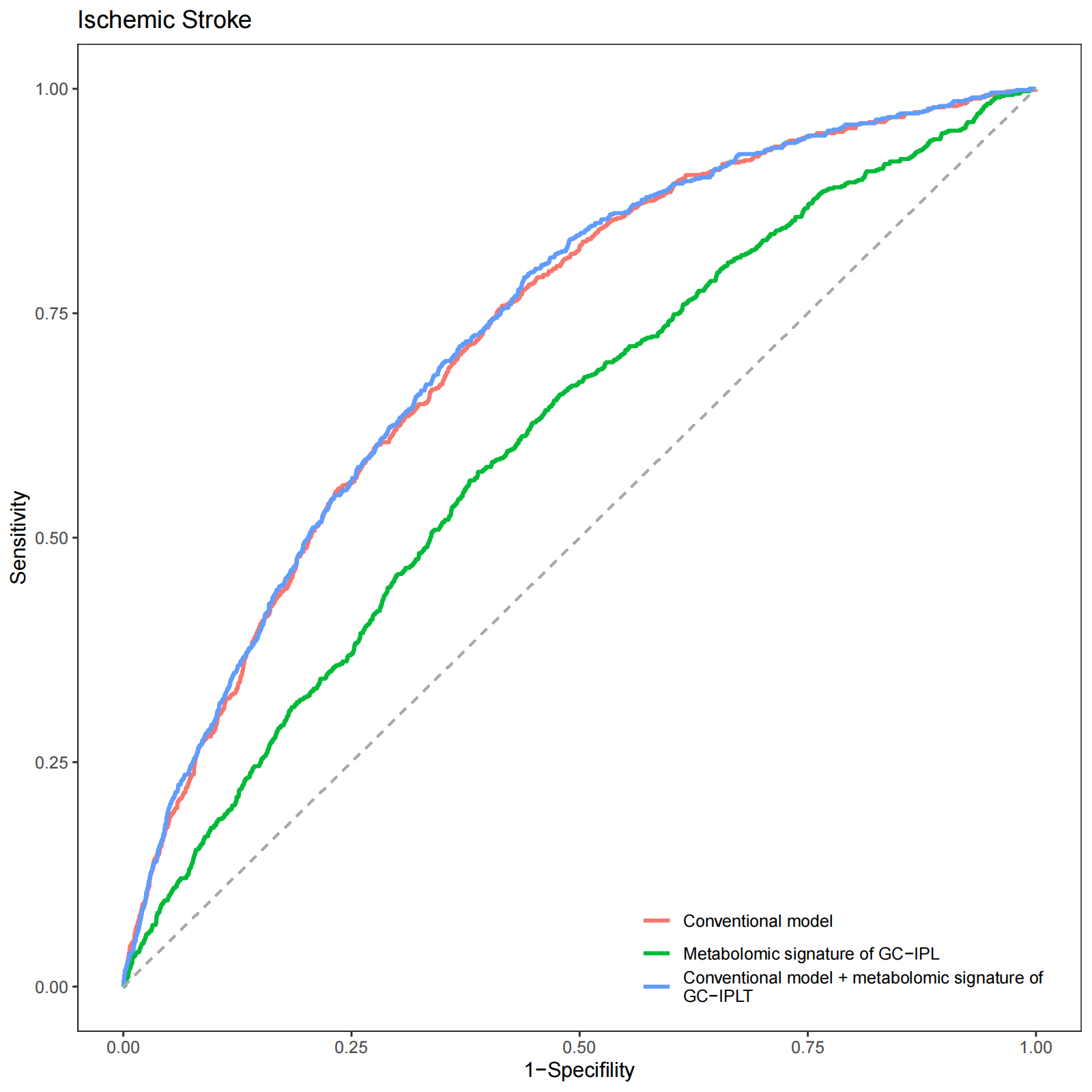

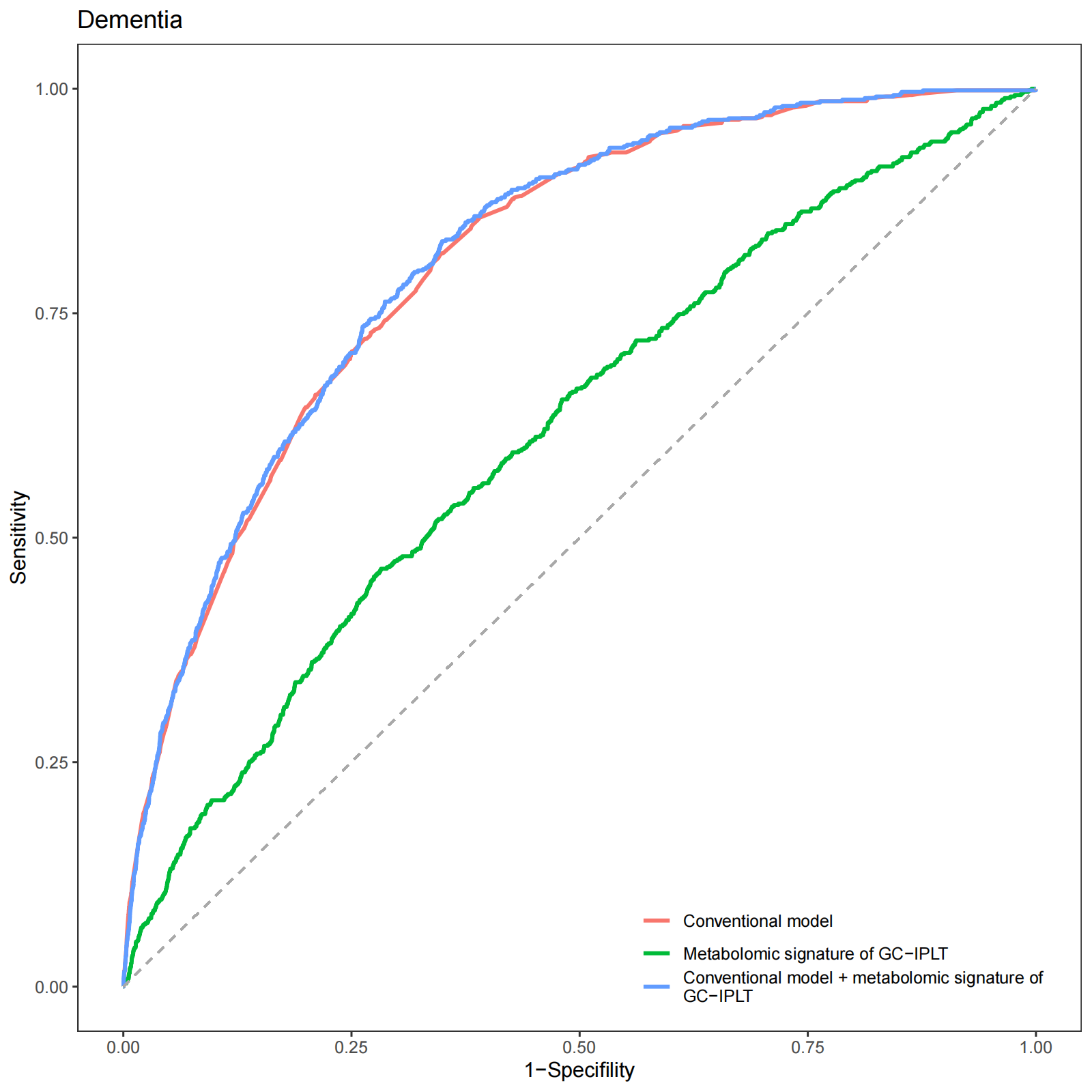

#### Supplementary Figure S11

**Receiver operating characteristic (ROC) curves of conventional model, meta-GCIPLT score, and combined model predicting mortality in training set.**

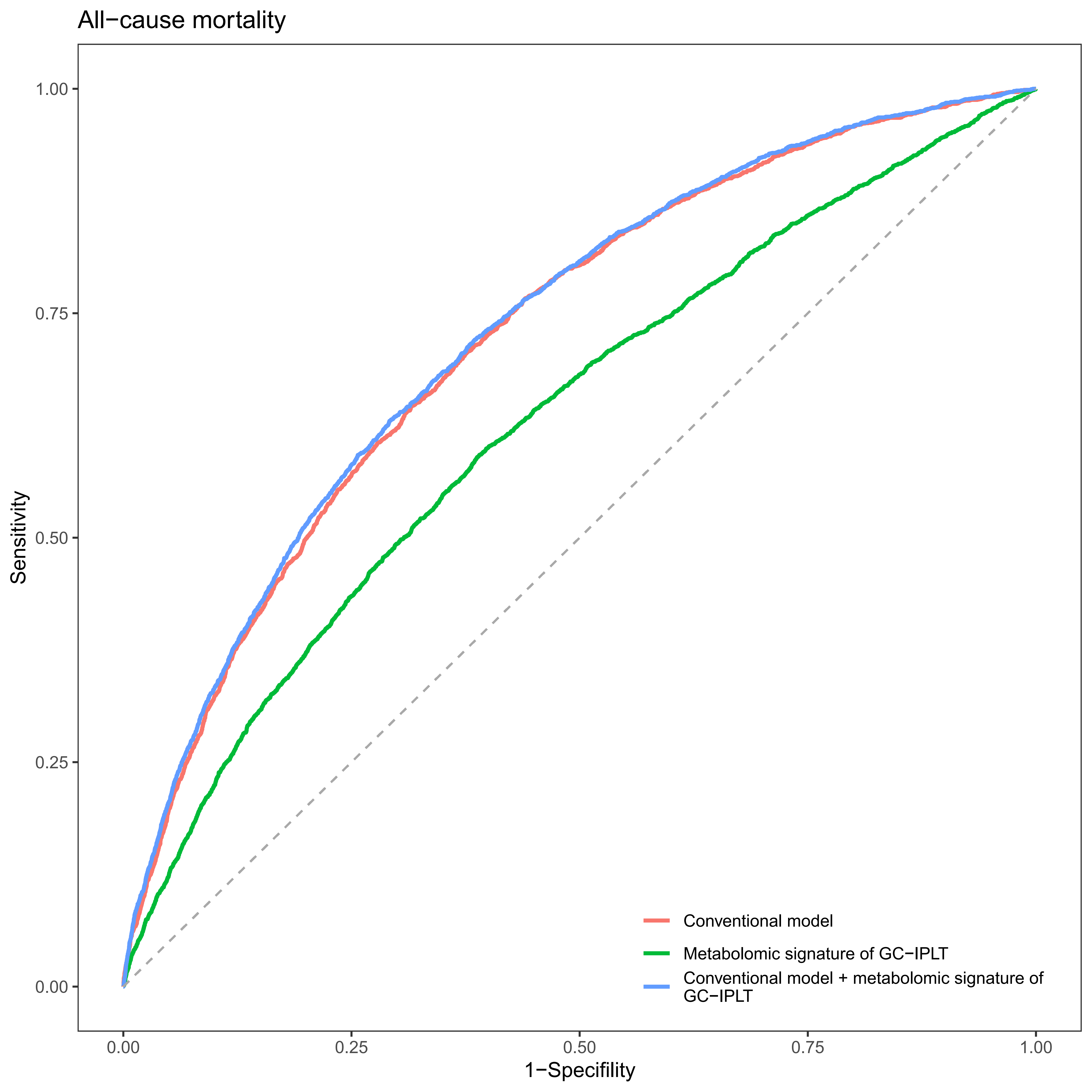

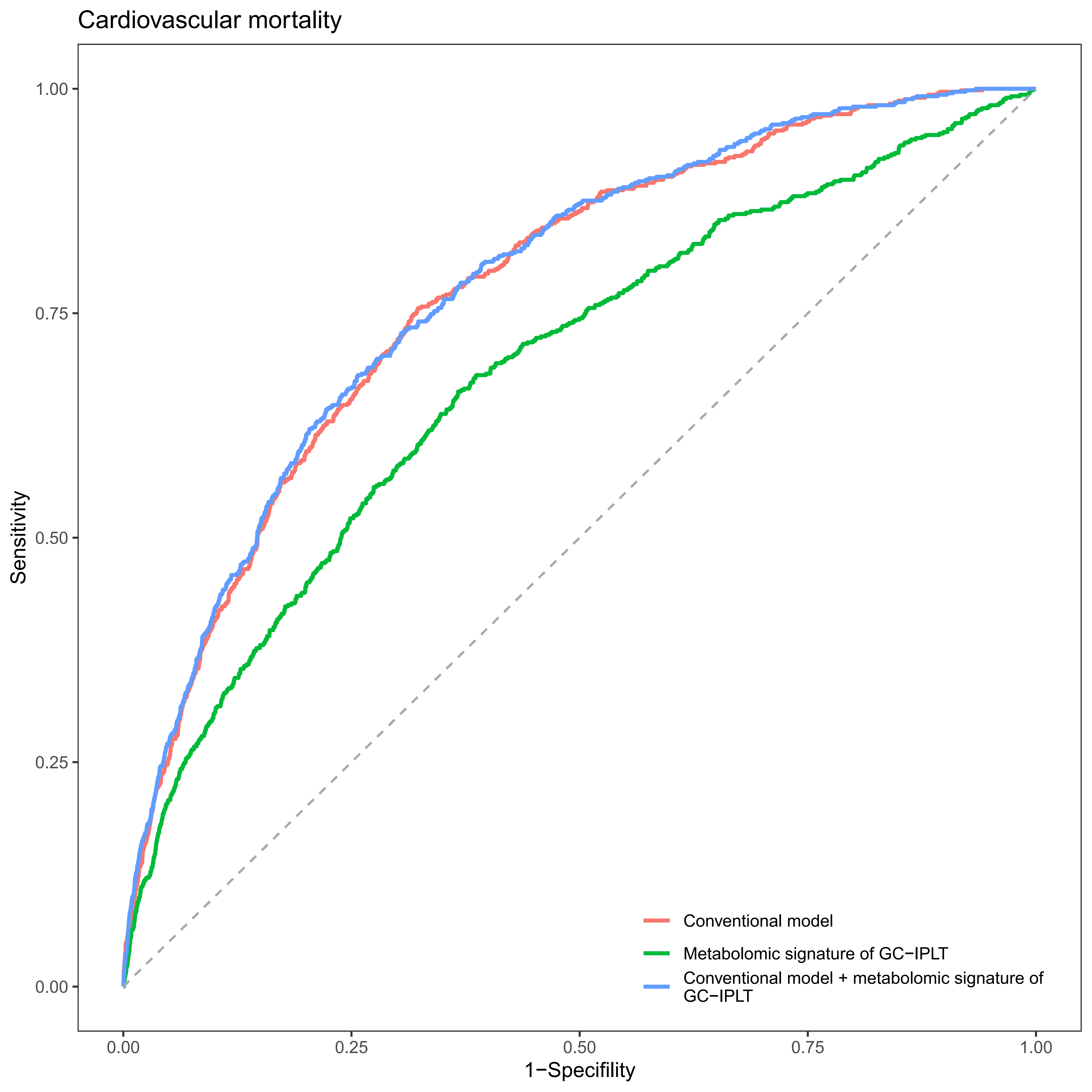

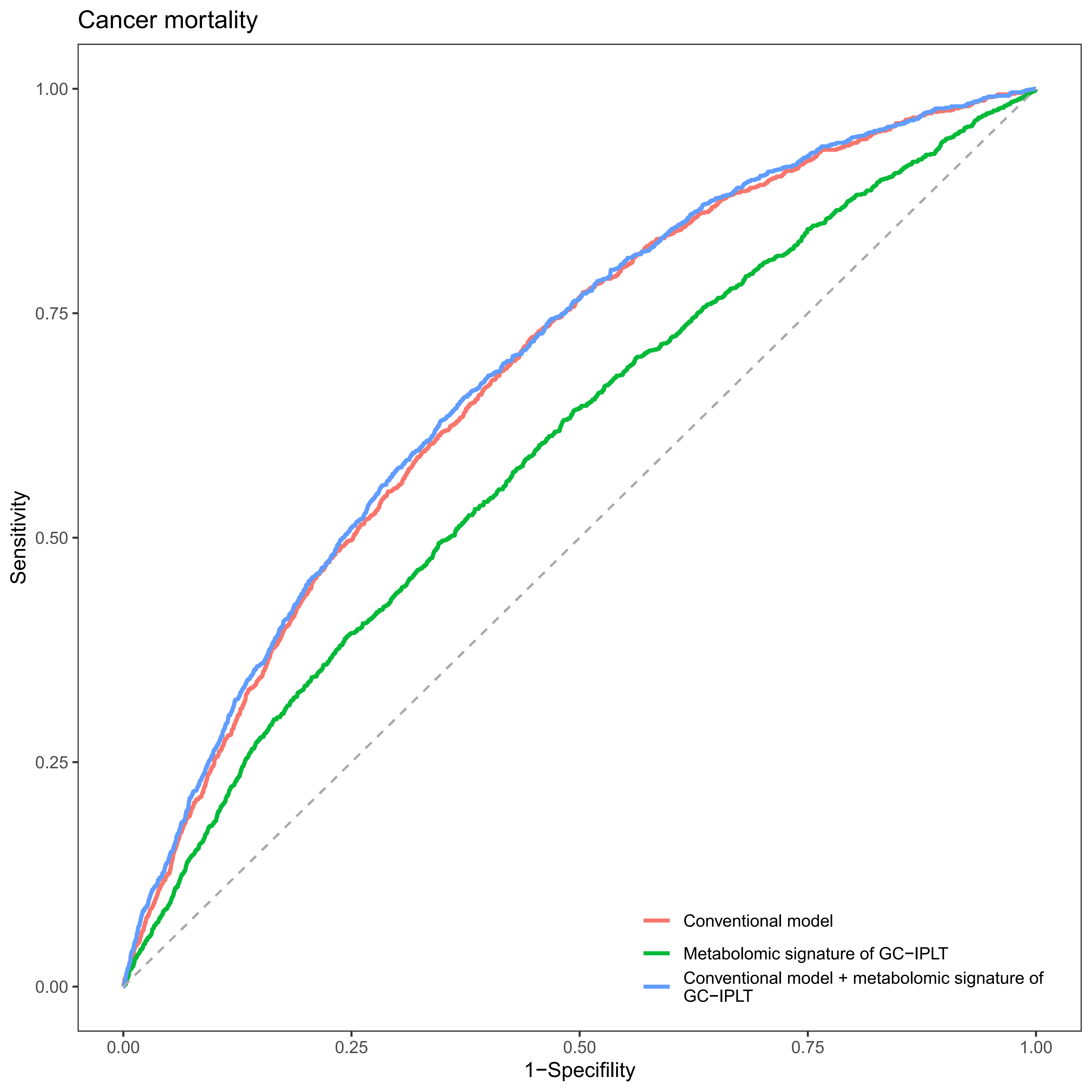

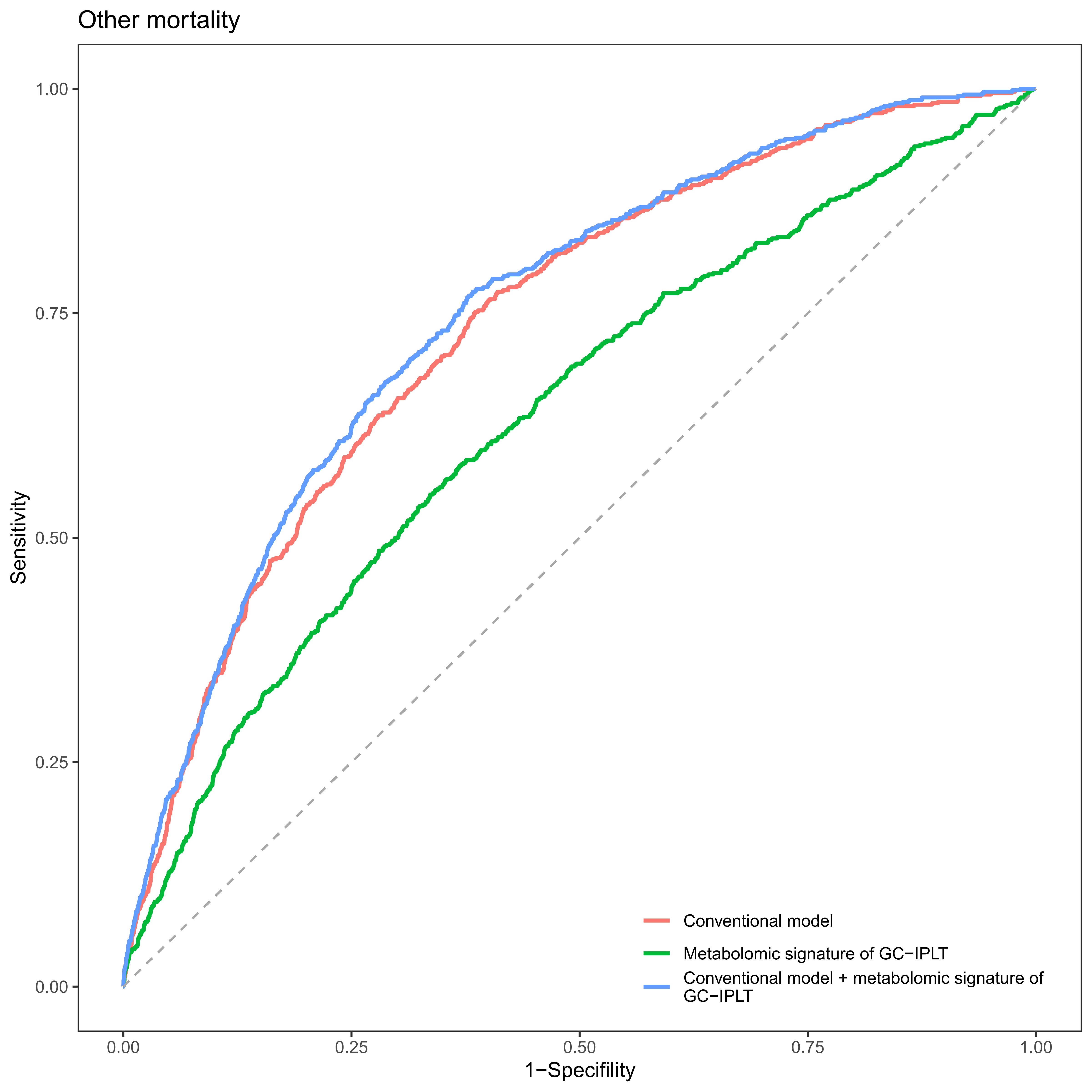

#### Supplementary Figure S12

**Receiver operating characteristic (ROC) curves of conventional model, meta-GCIPLT score, and combined model predicting systemic diseases in validation set.**

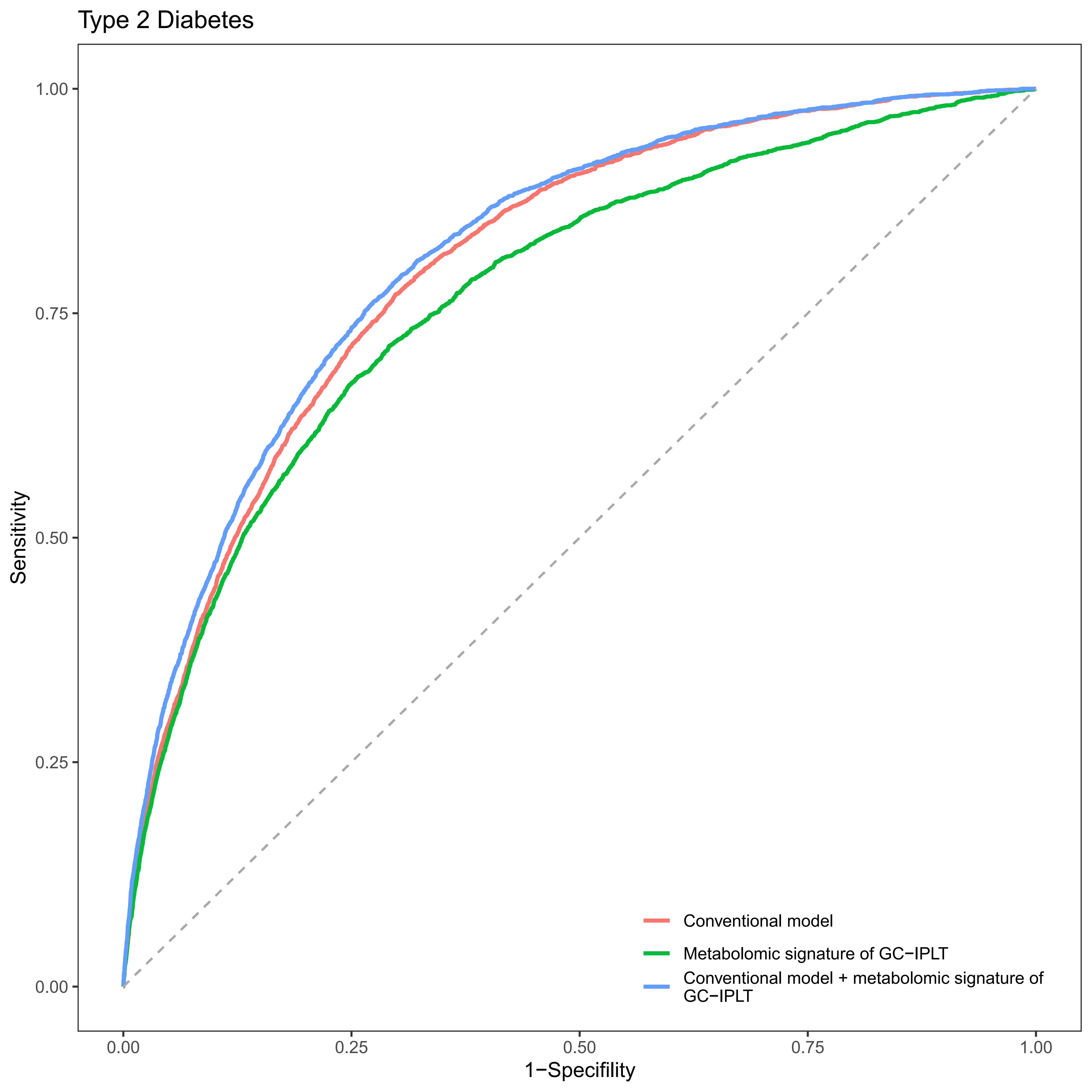

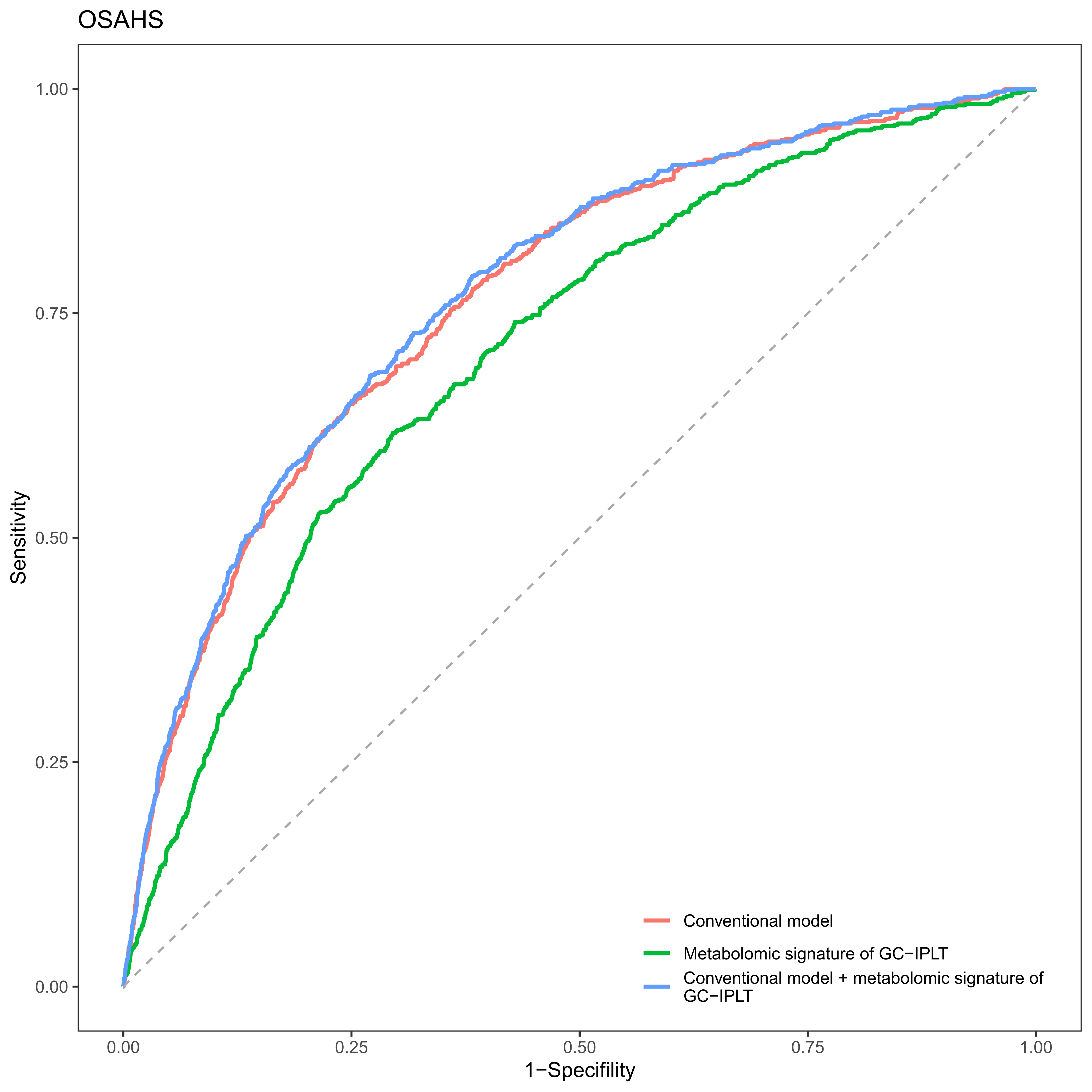

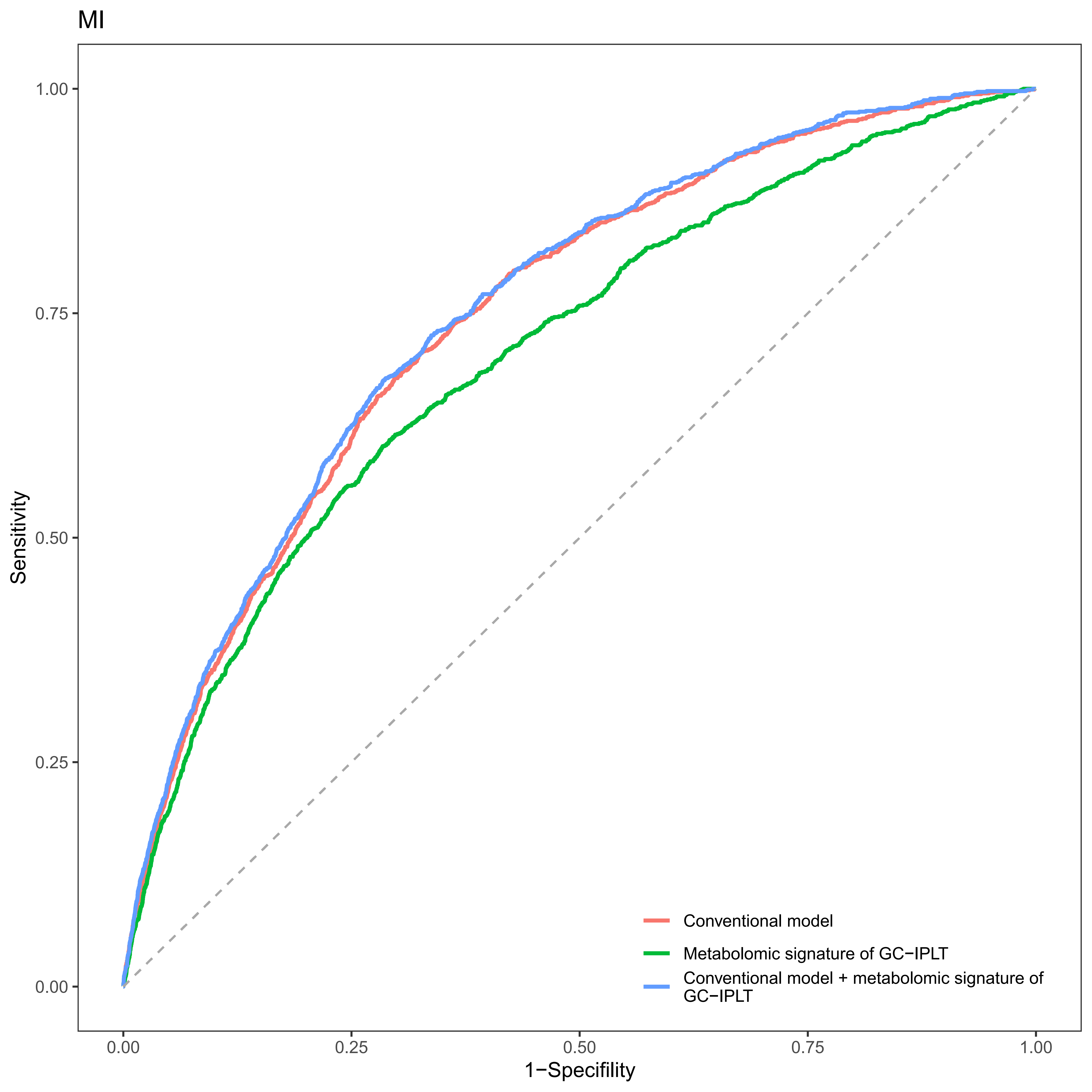

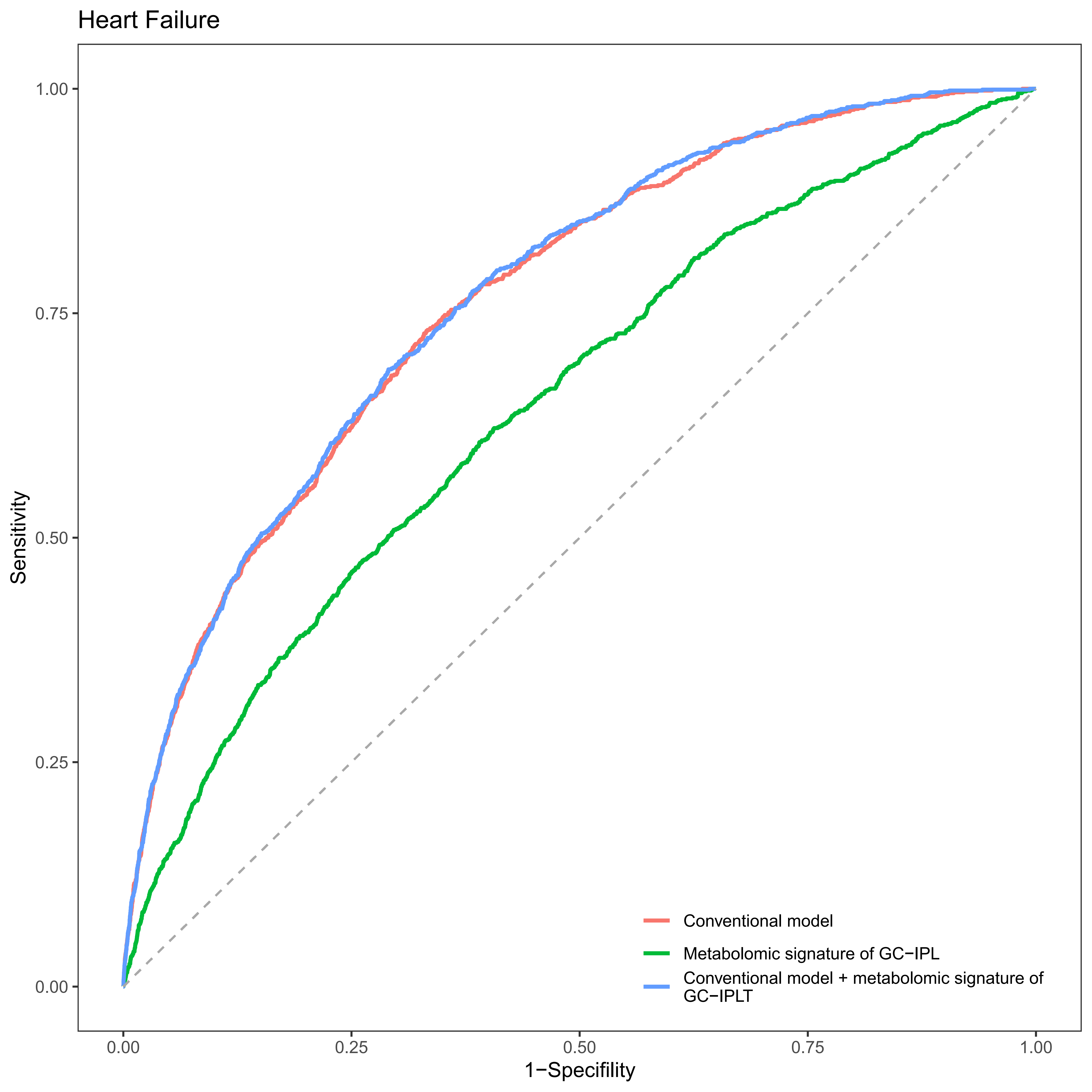

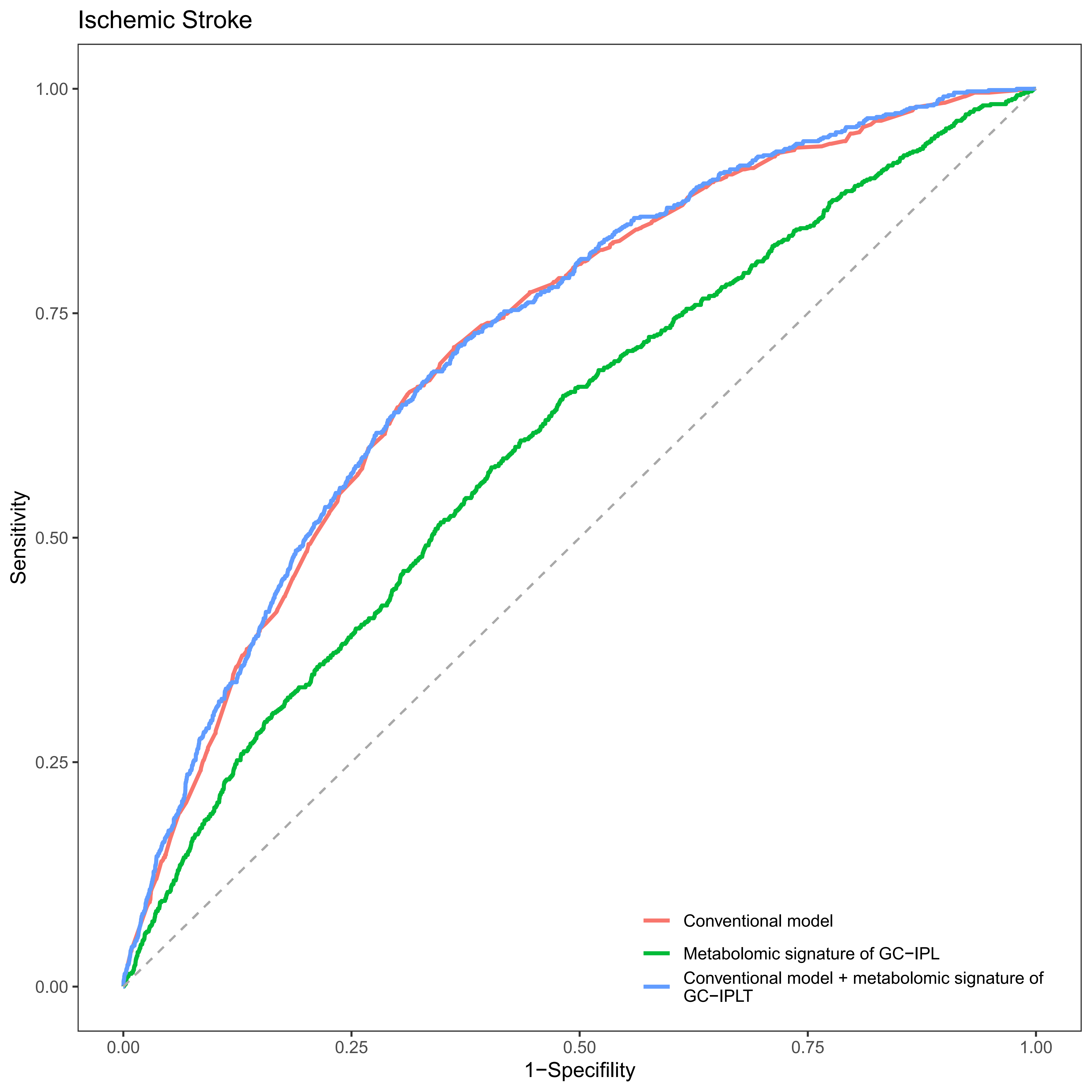

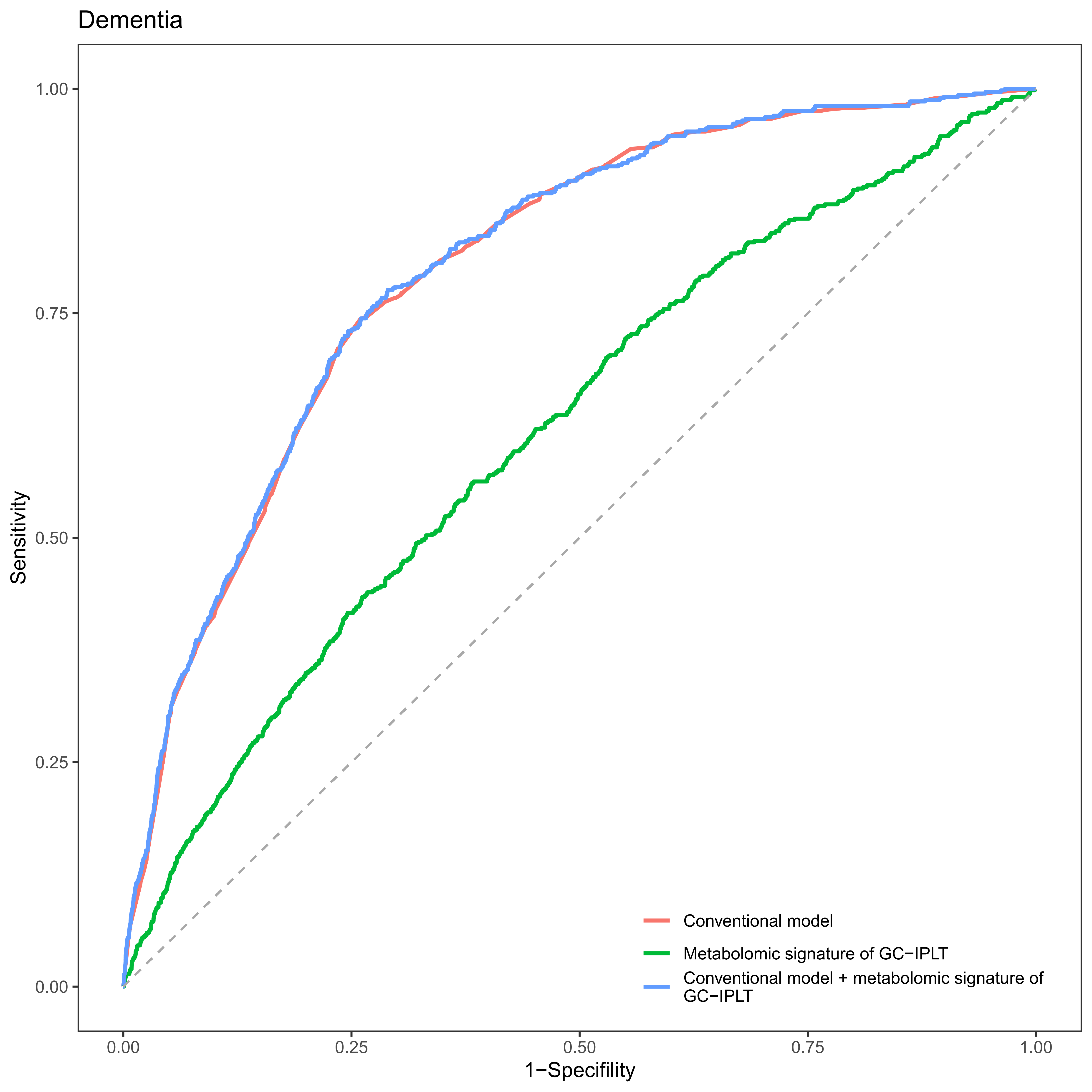

#### Supplementary Figure S13

**Receiver operating characteristic (ROC) curves of conventional model, meta-GCIPLT score, and combined model predicting mortality in validation set.**

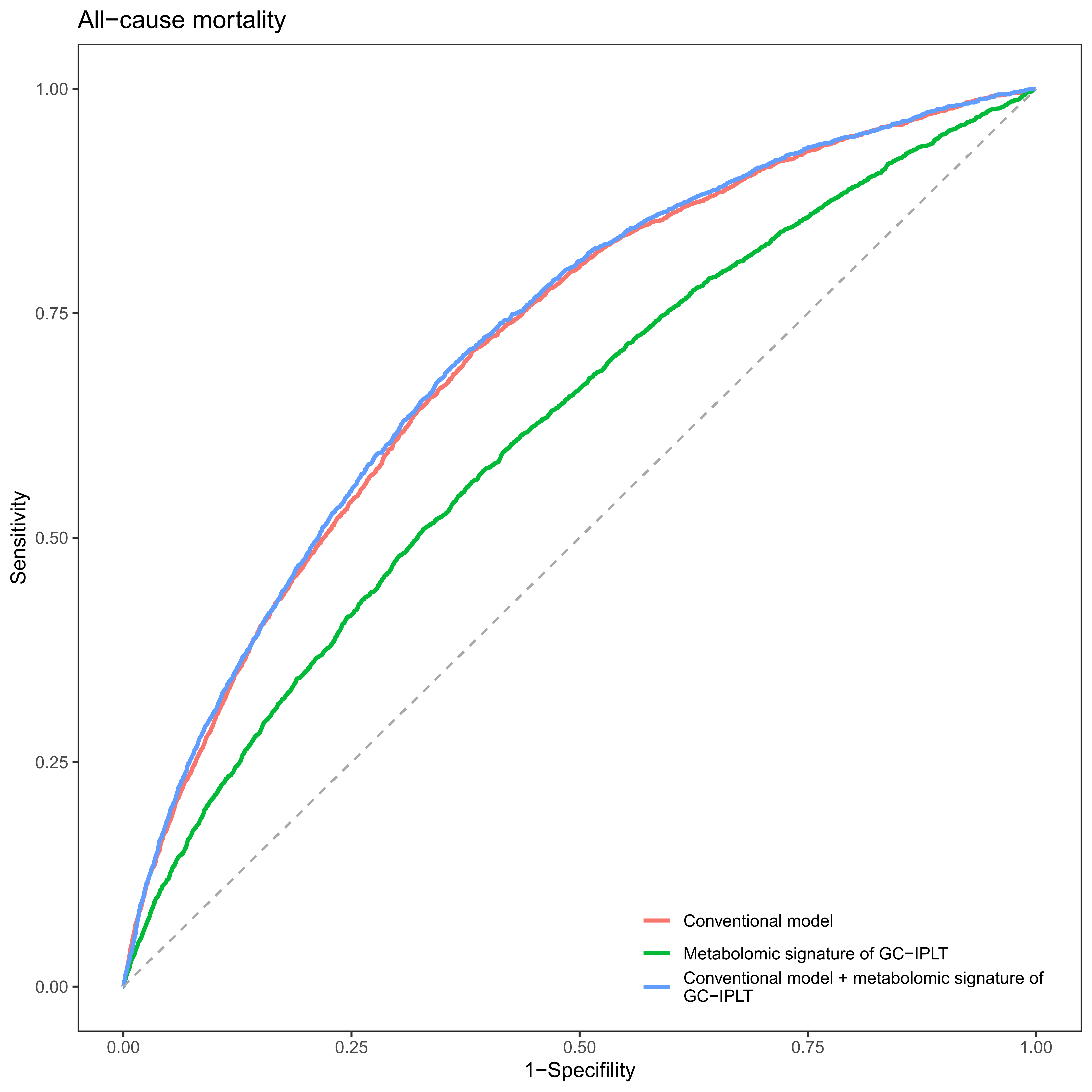

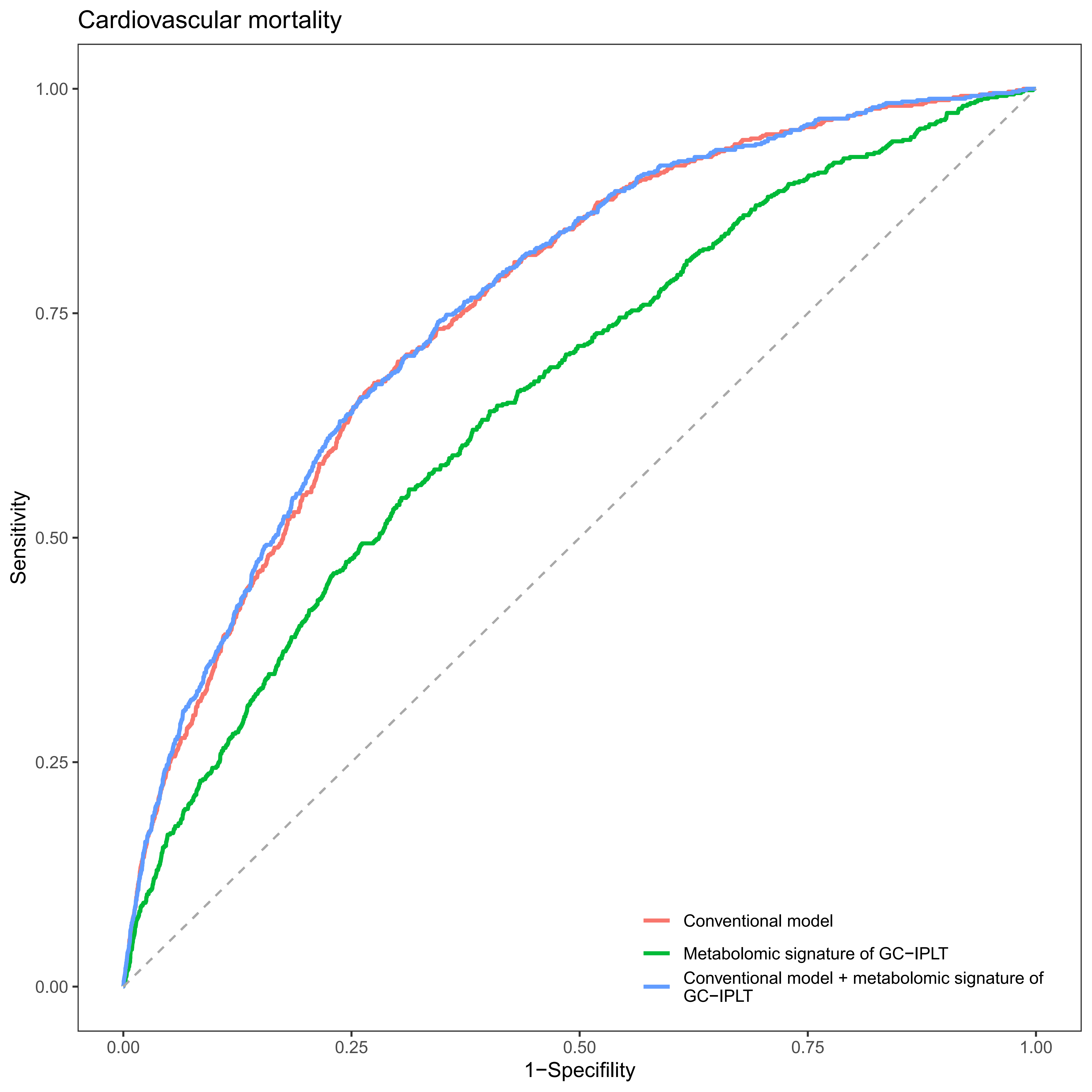

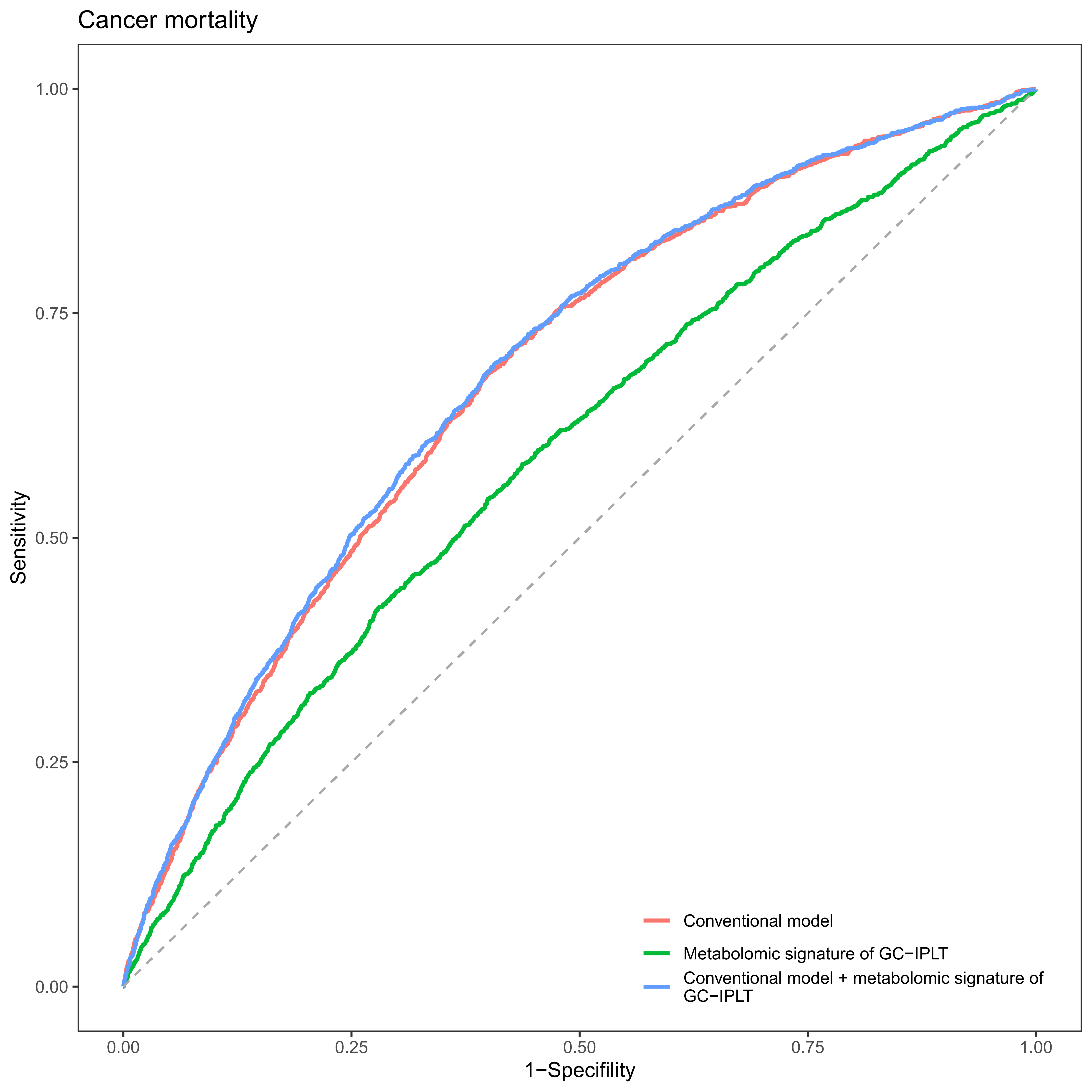

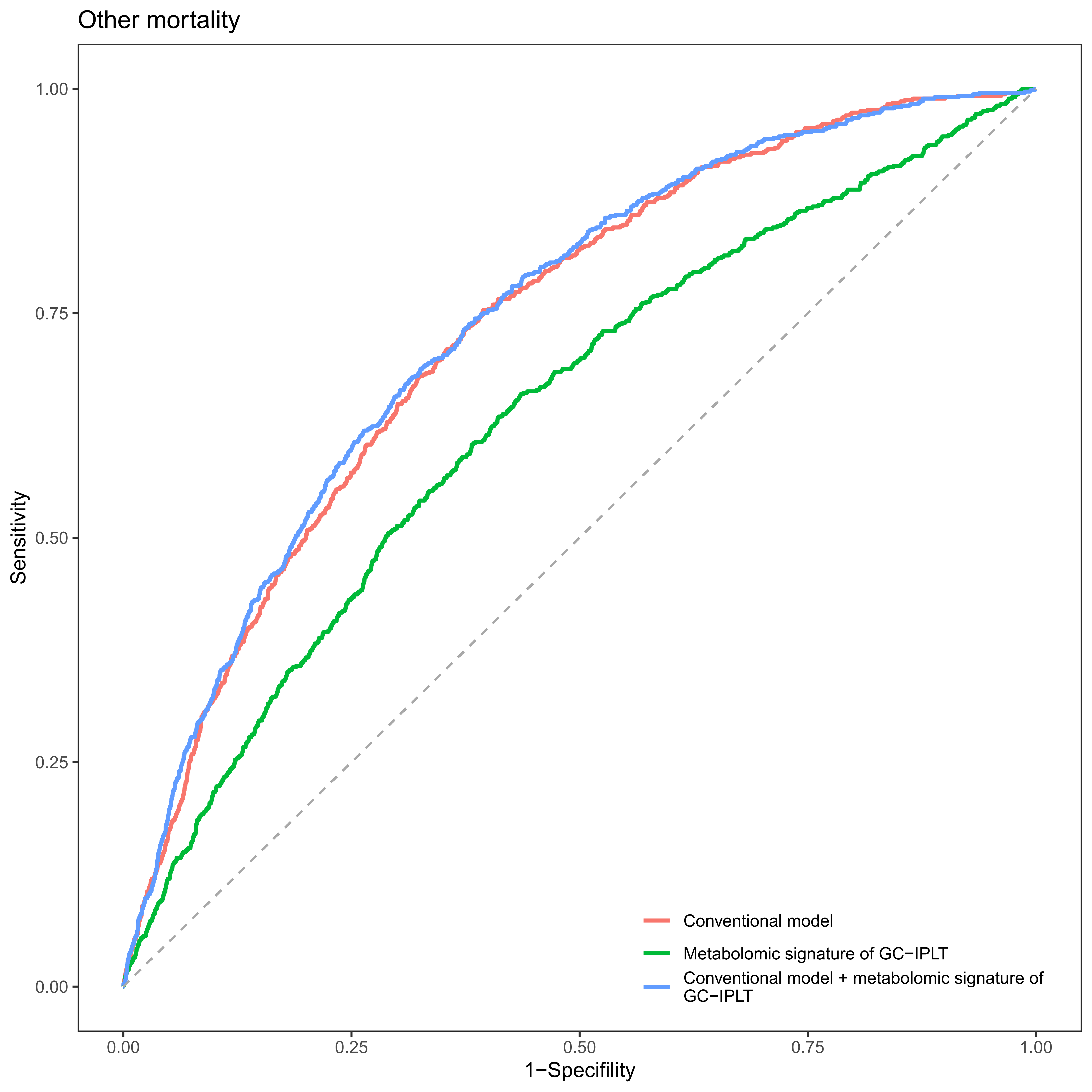

#### Supplementary Figure S14

**Reclassification in the new model for systemic diseases with the inclusion of meta-GCIPLT score in training set.**

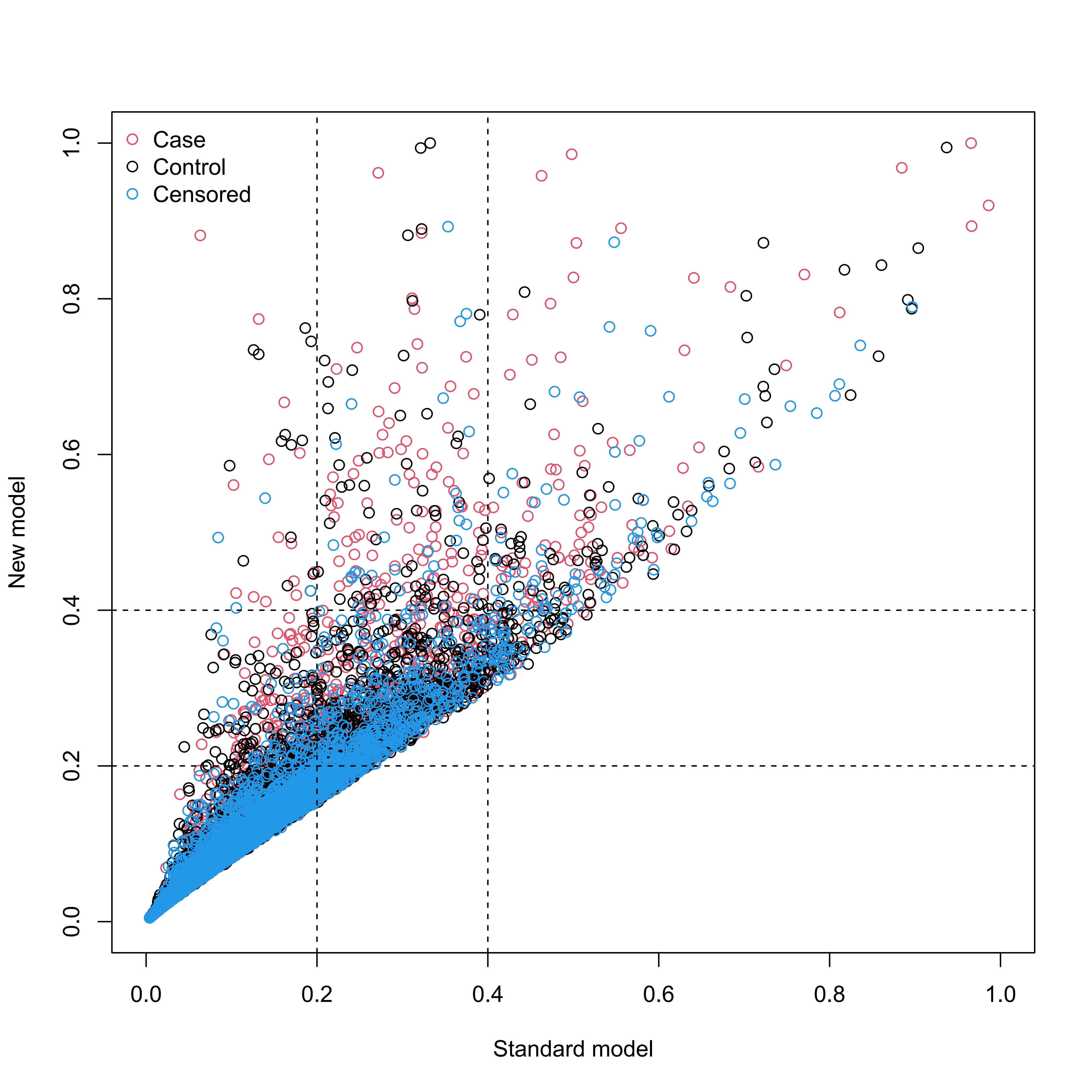

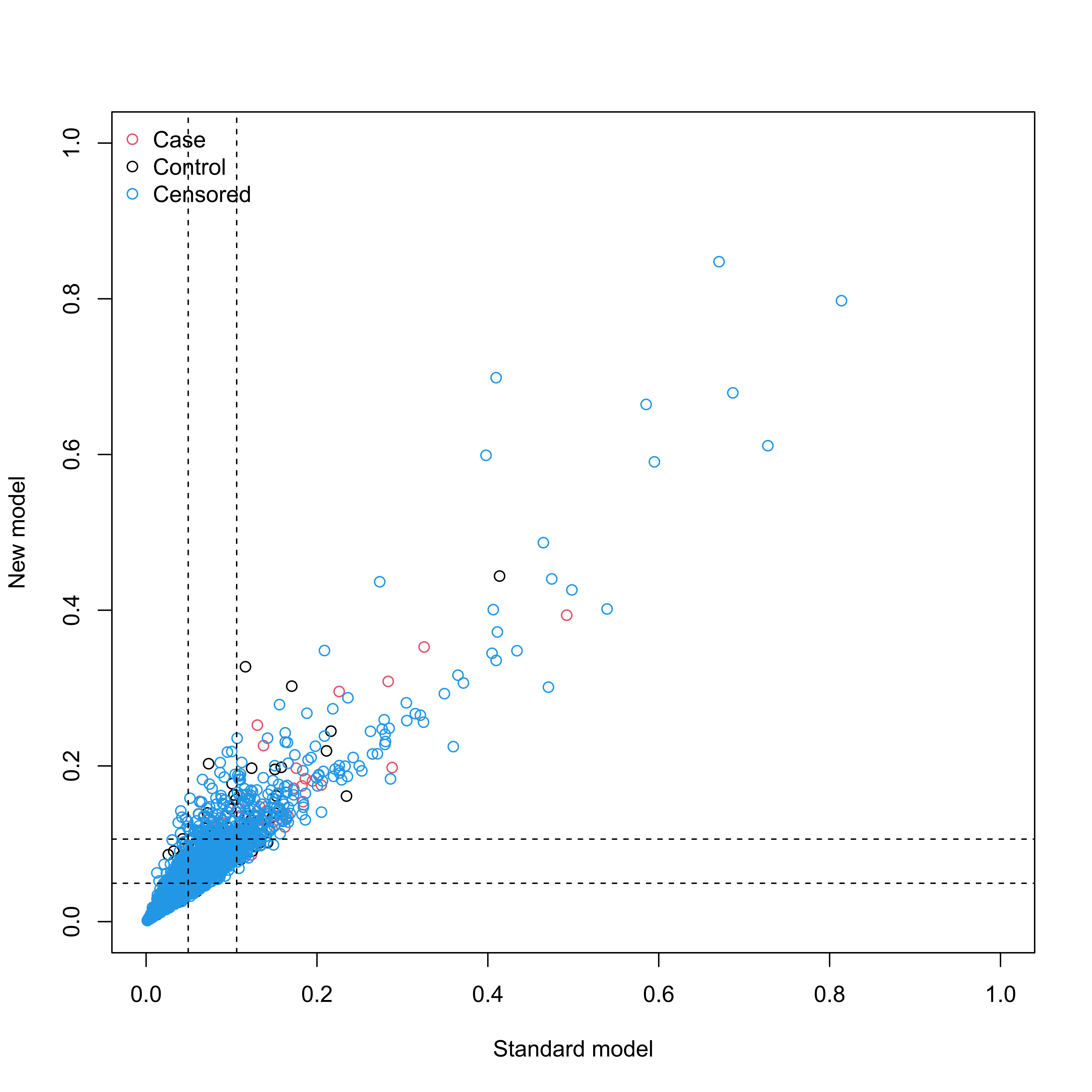

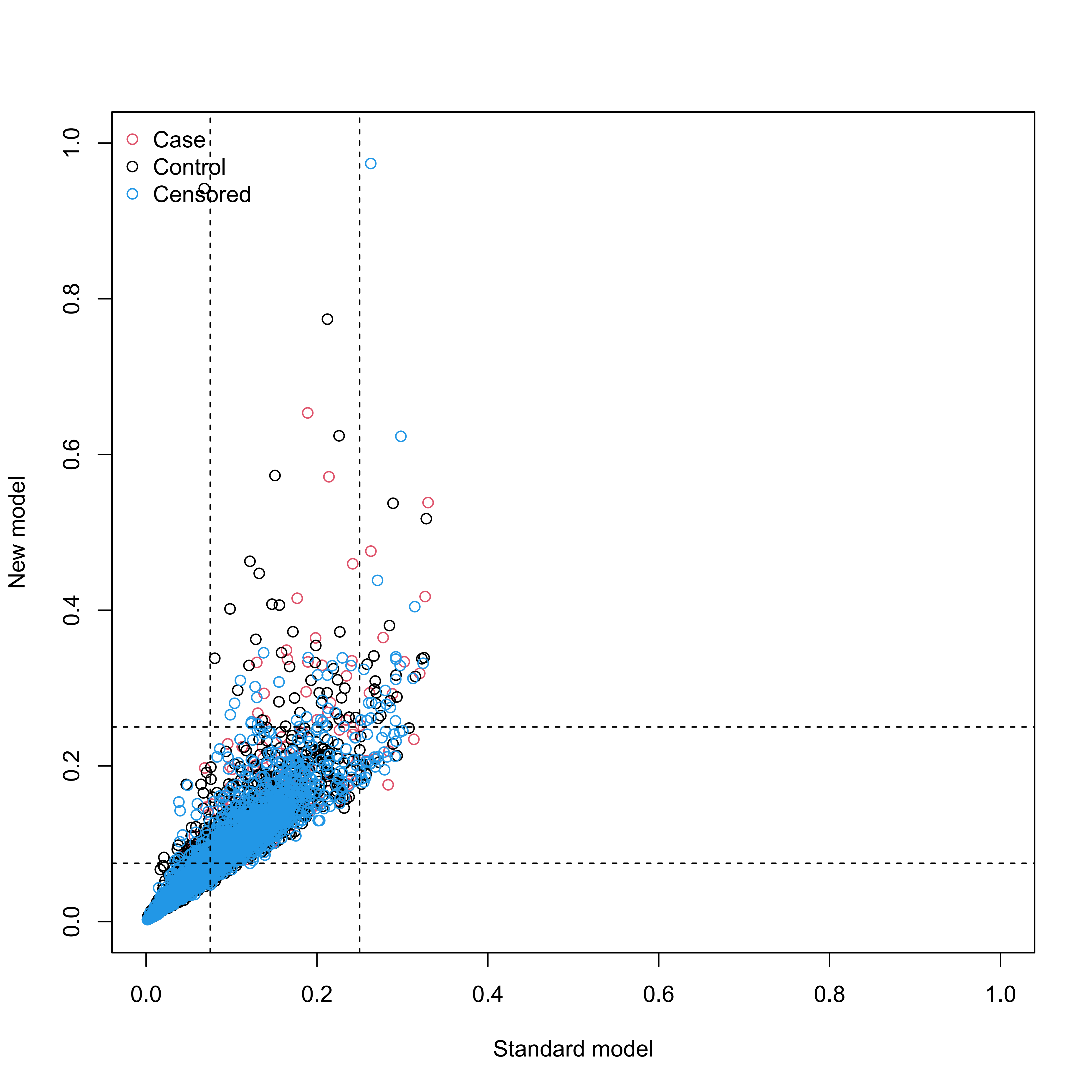

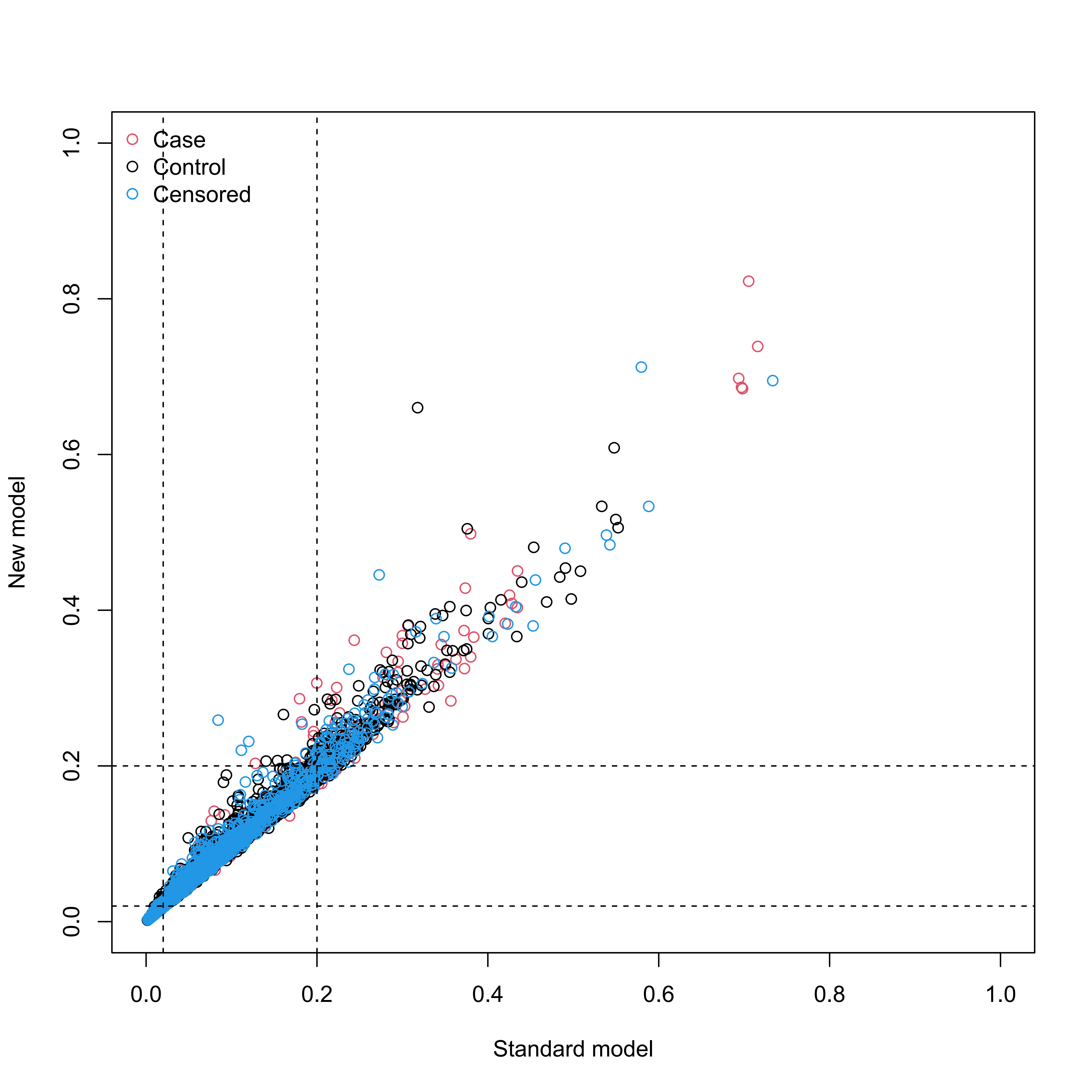

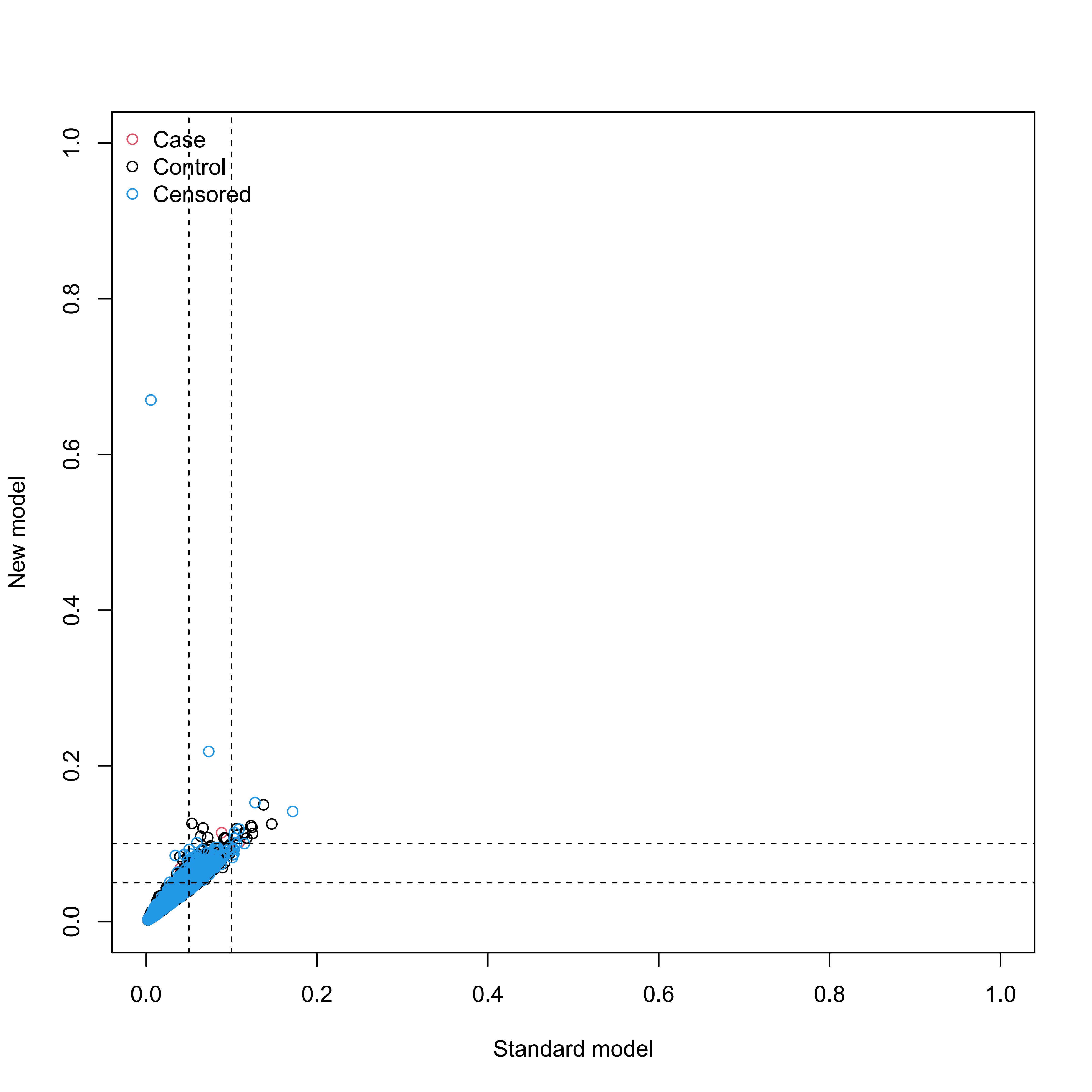

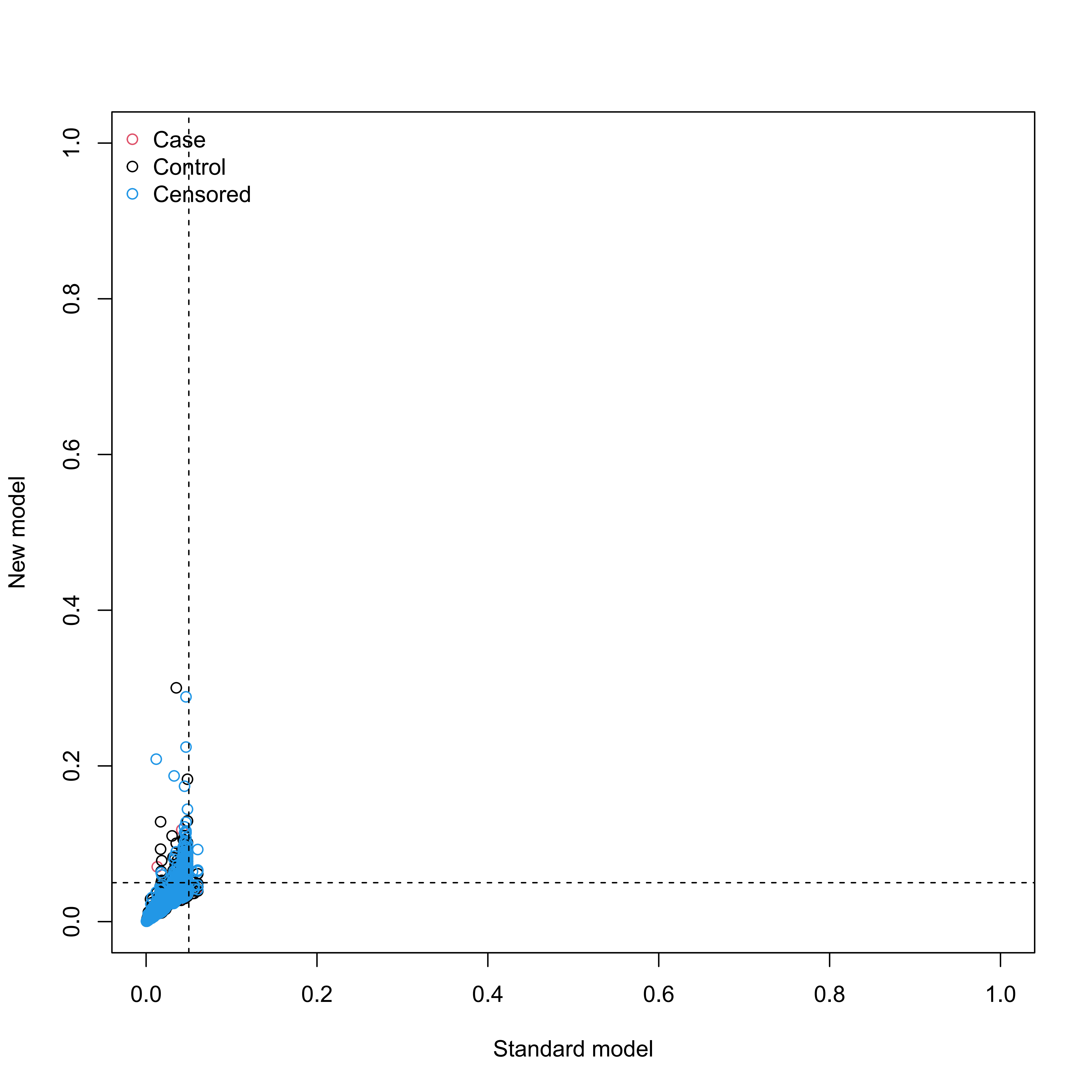

#### Supplementary Figure S15

**Reclassification in the new model for mortality with the inclusion of meta-GCIPLT score in training set.**

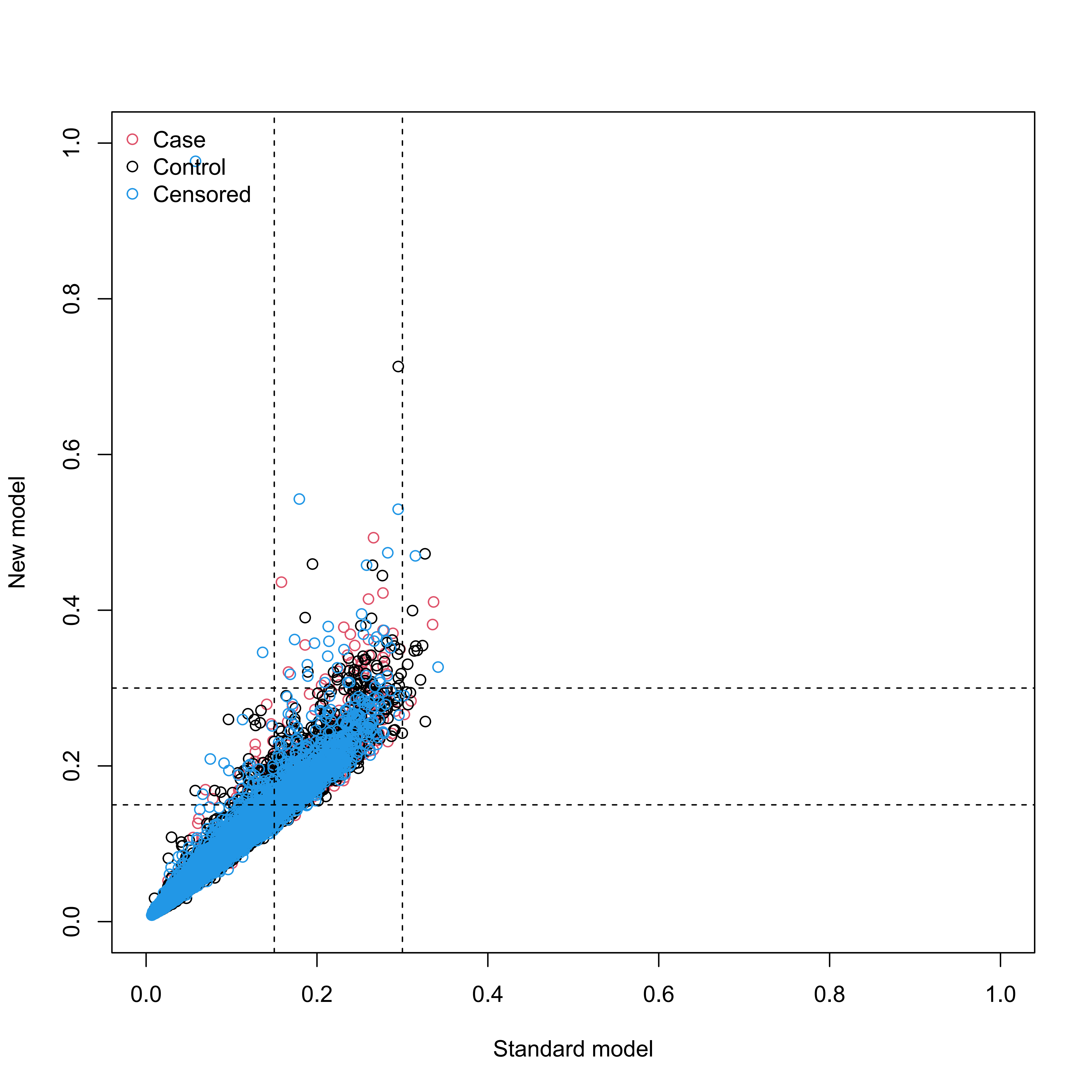

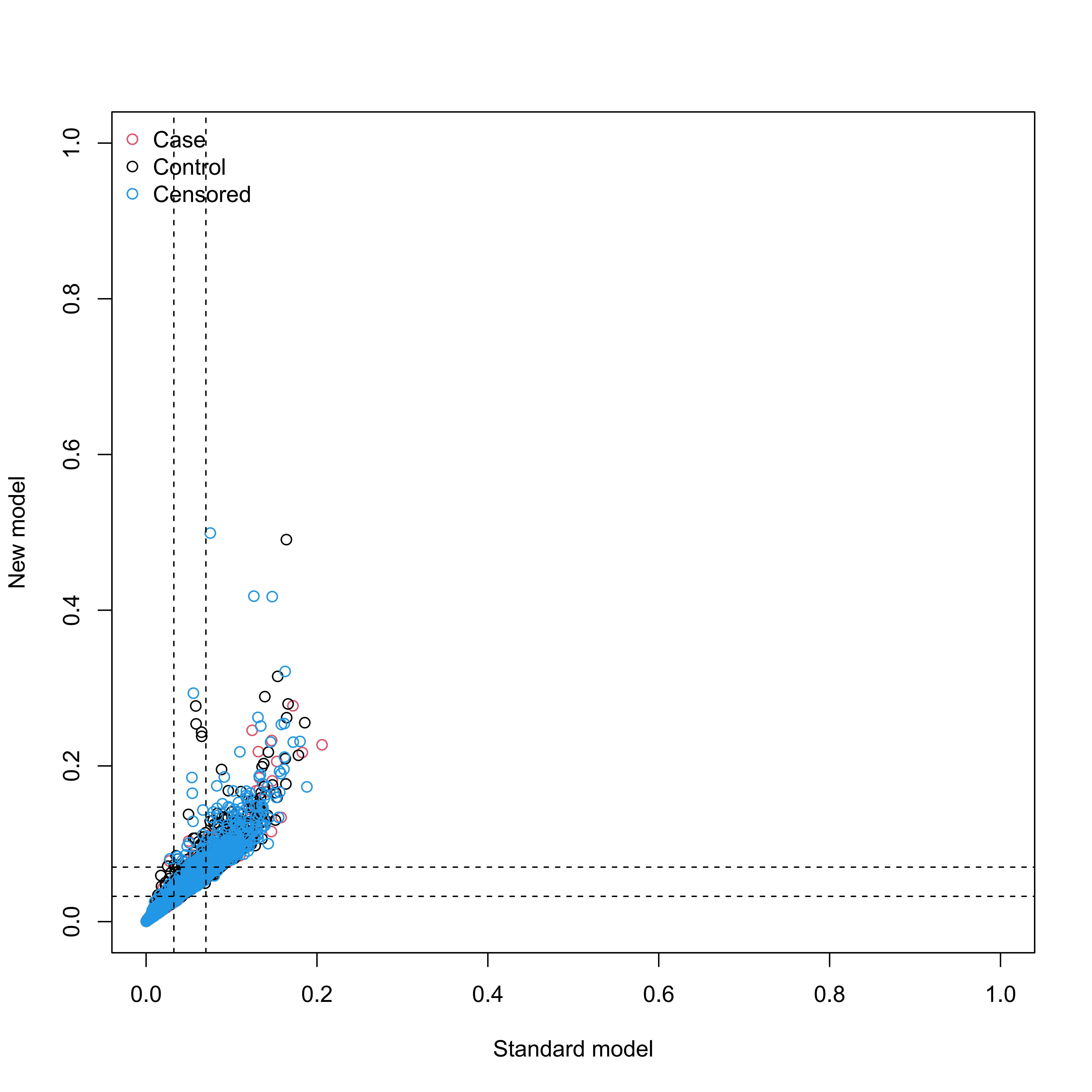

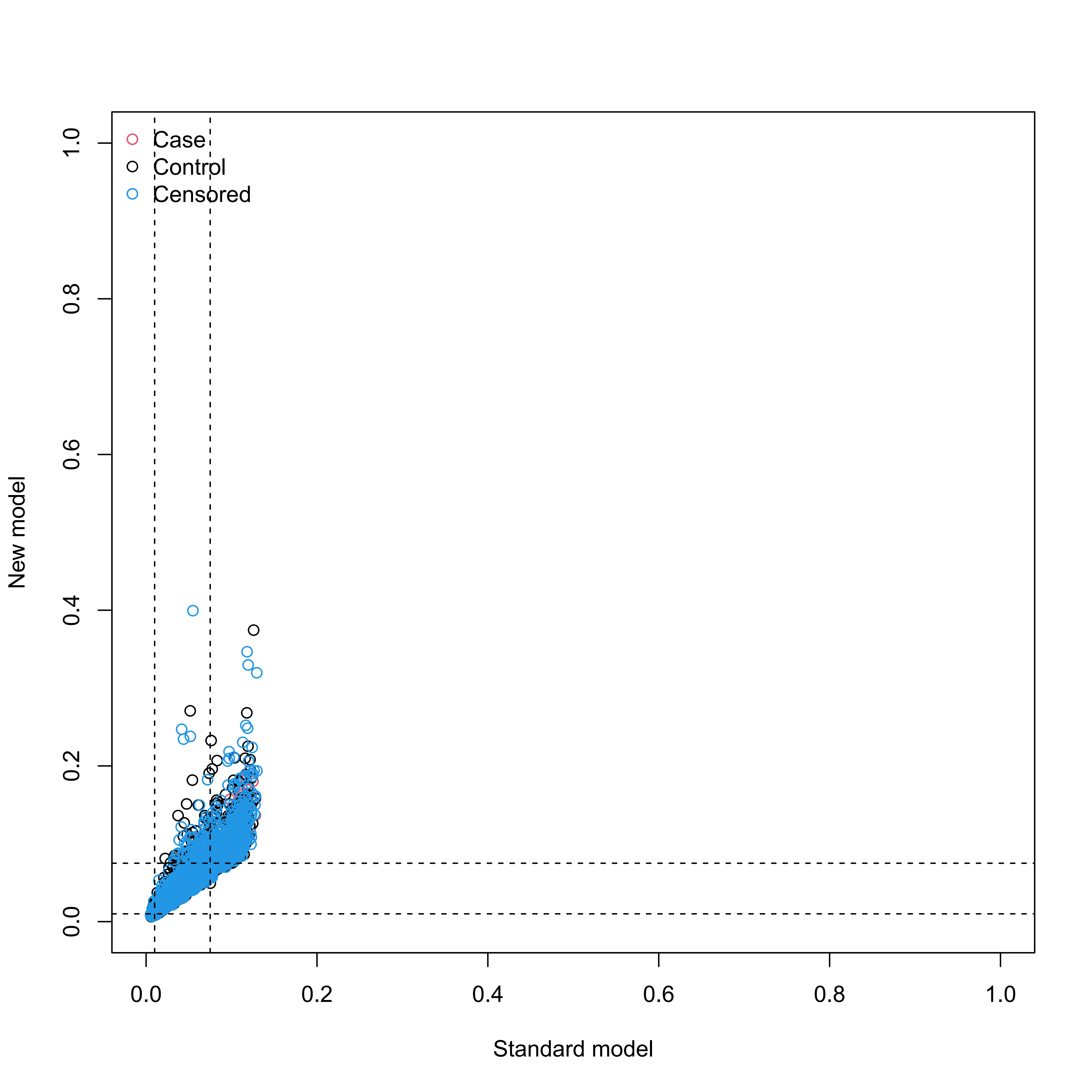

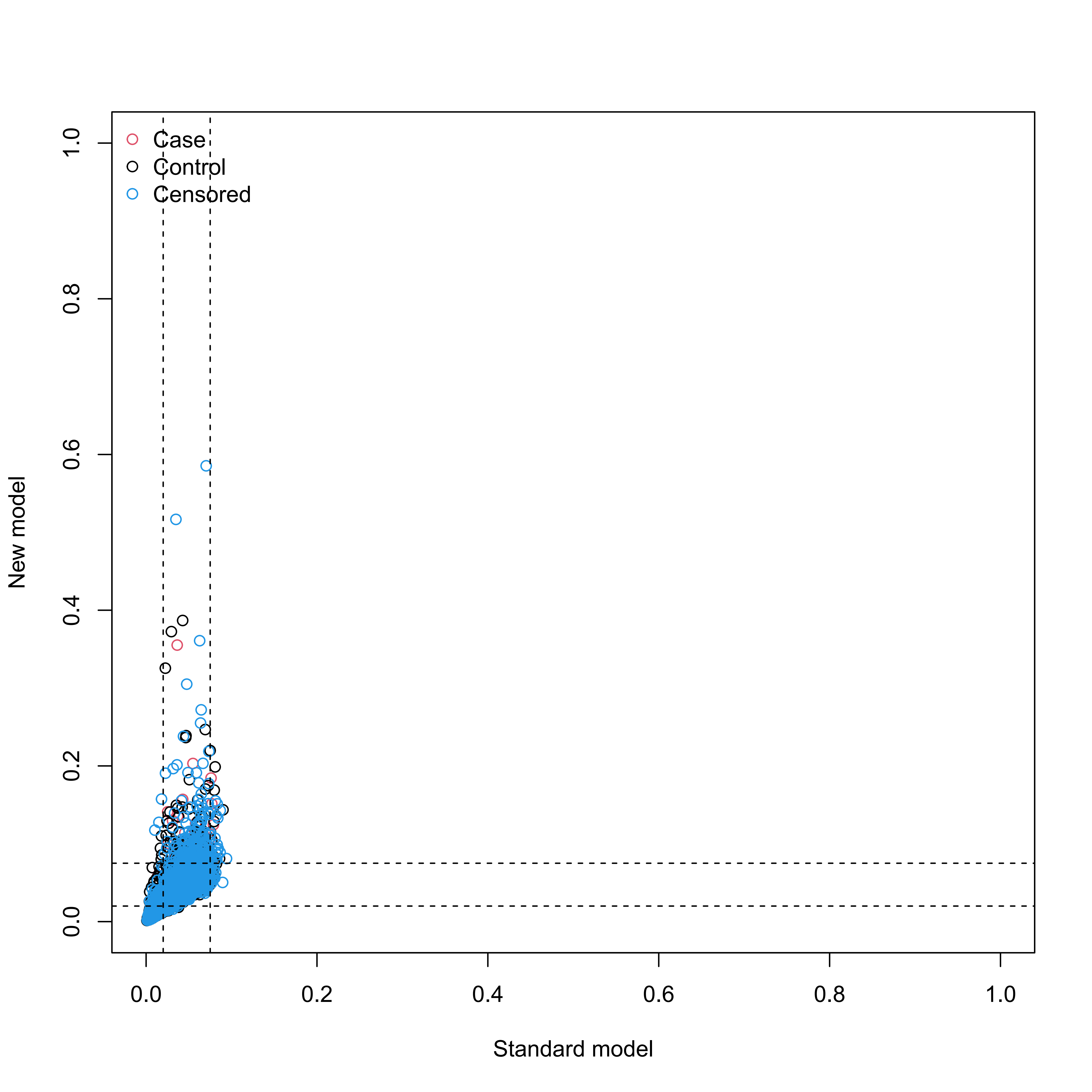

#### Supplementary Figure S16

**Reclassification in the new model for systemic diseases with the inclusion of meta-GCIPLT score in validation set.**

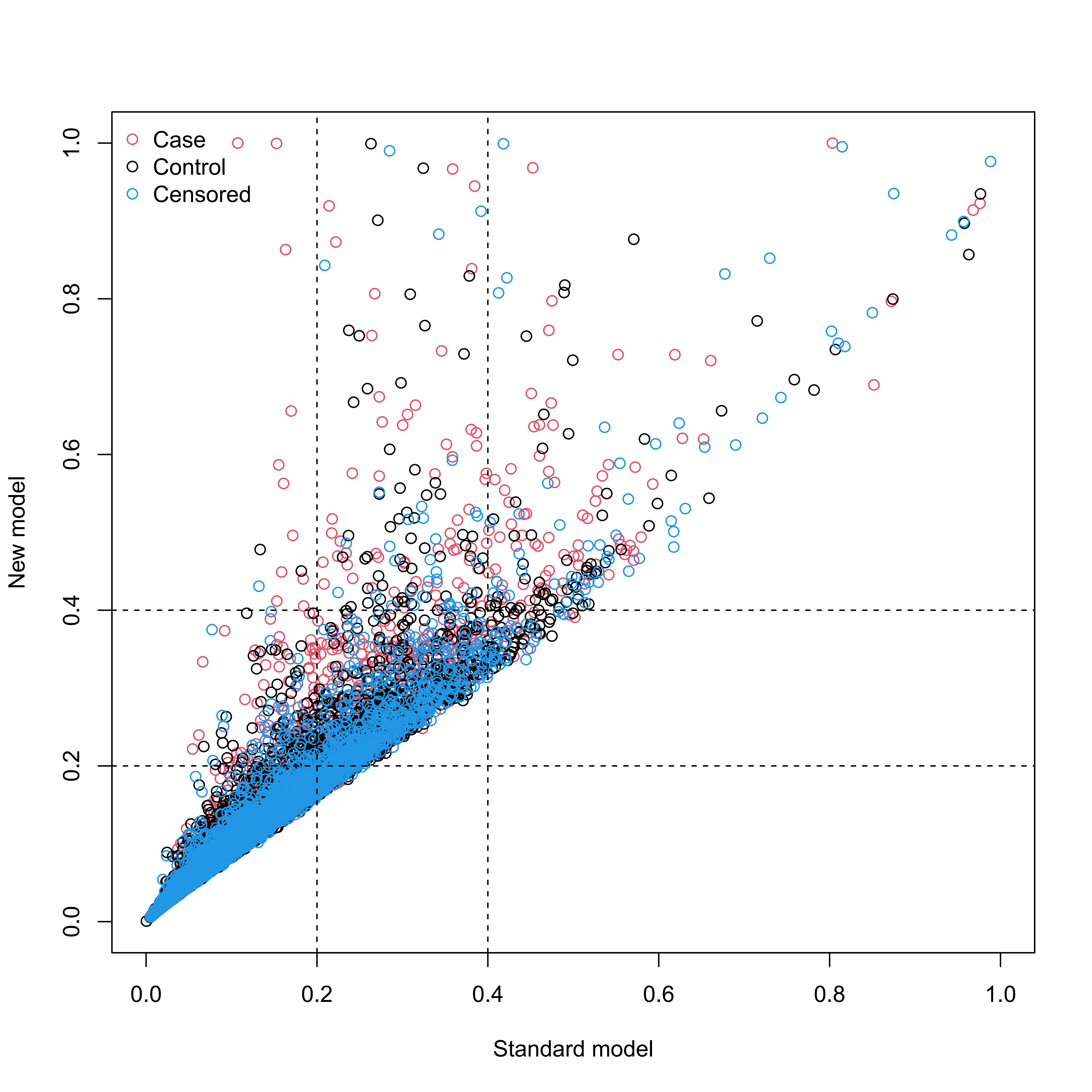

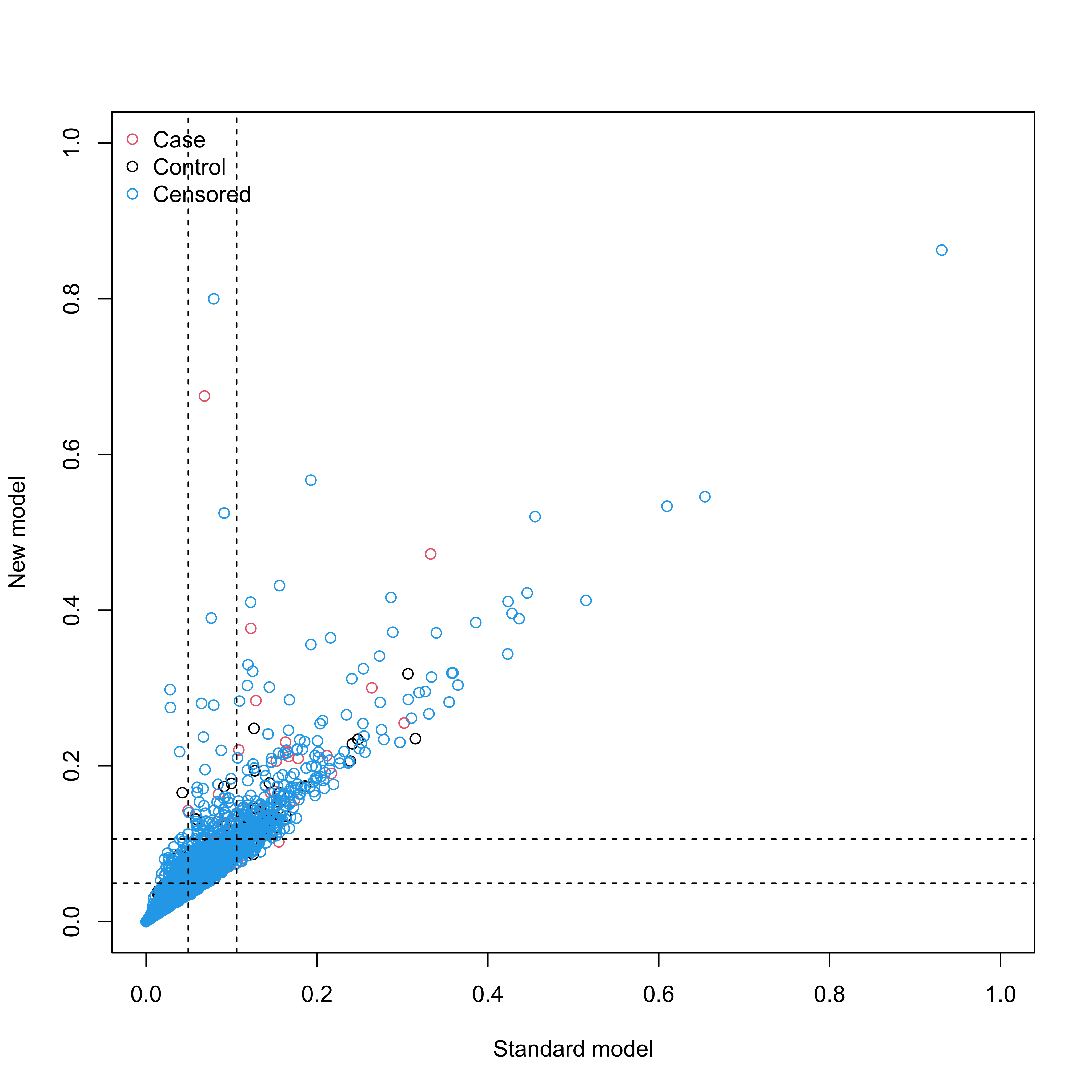

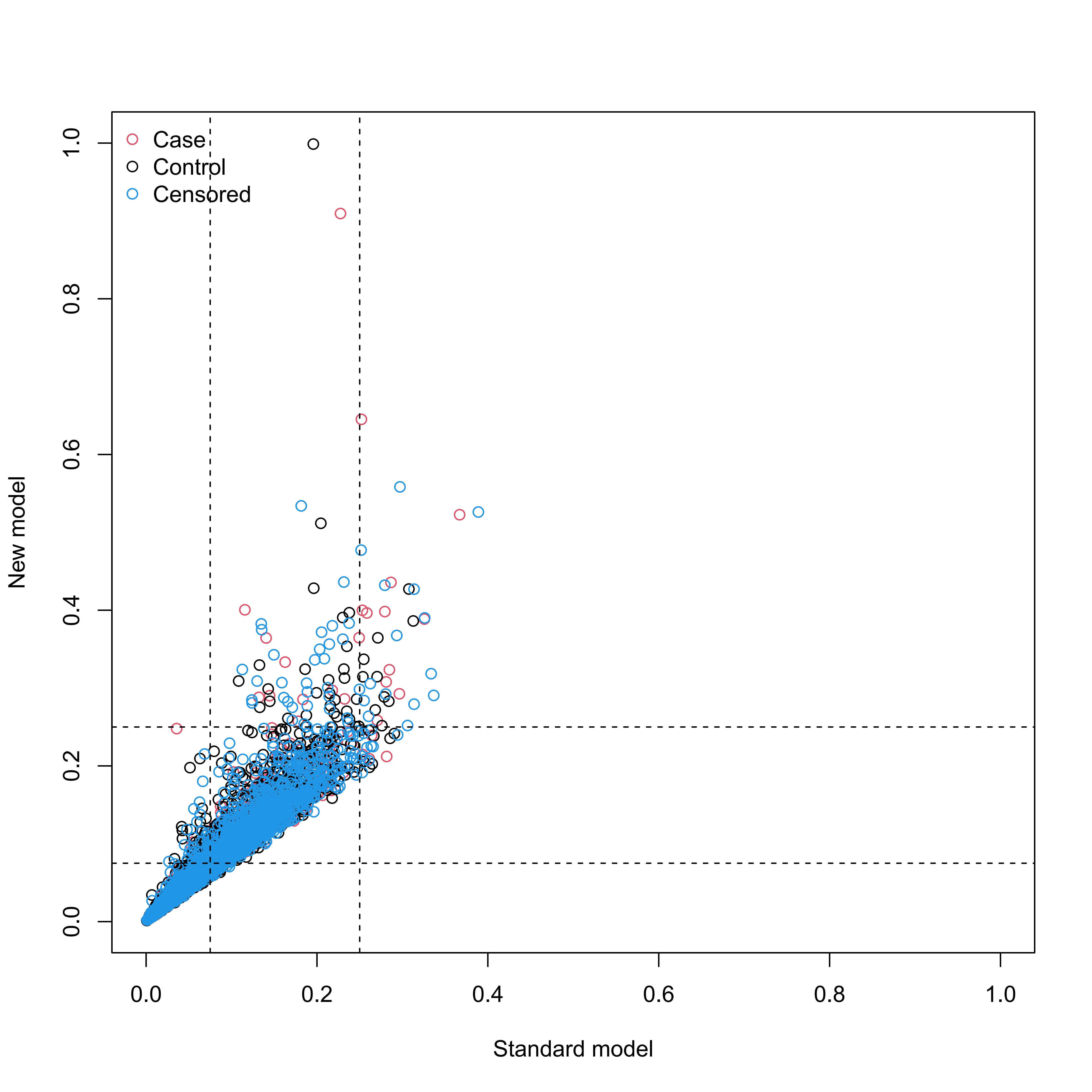

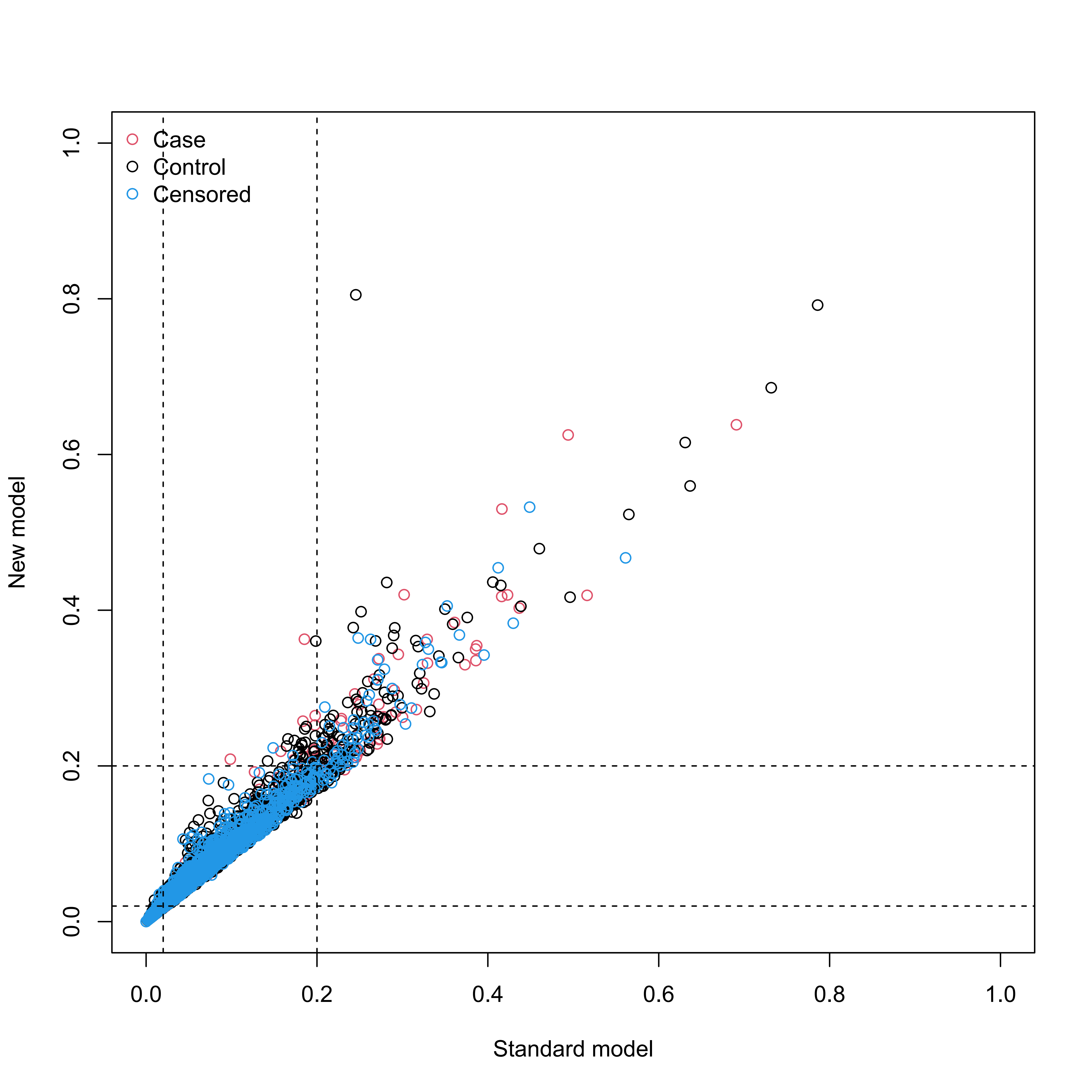

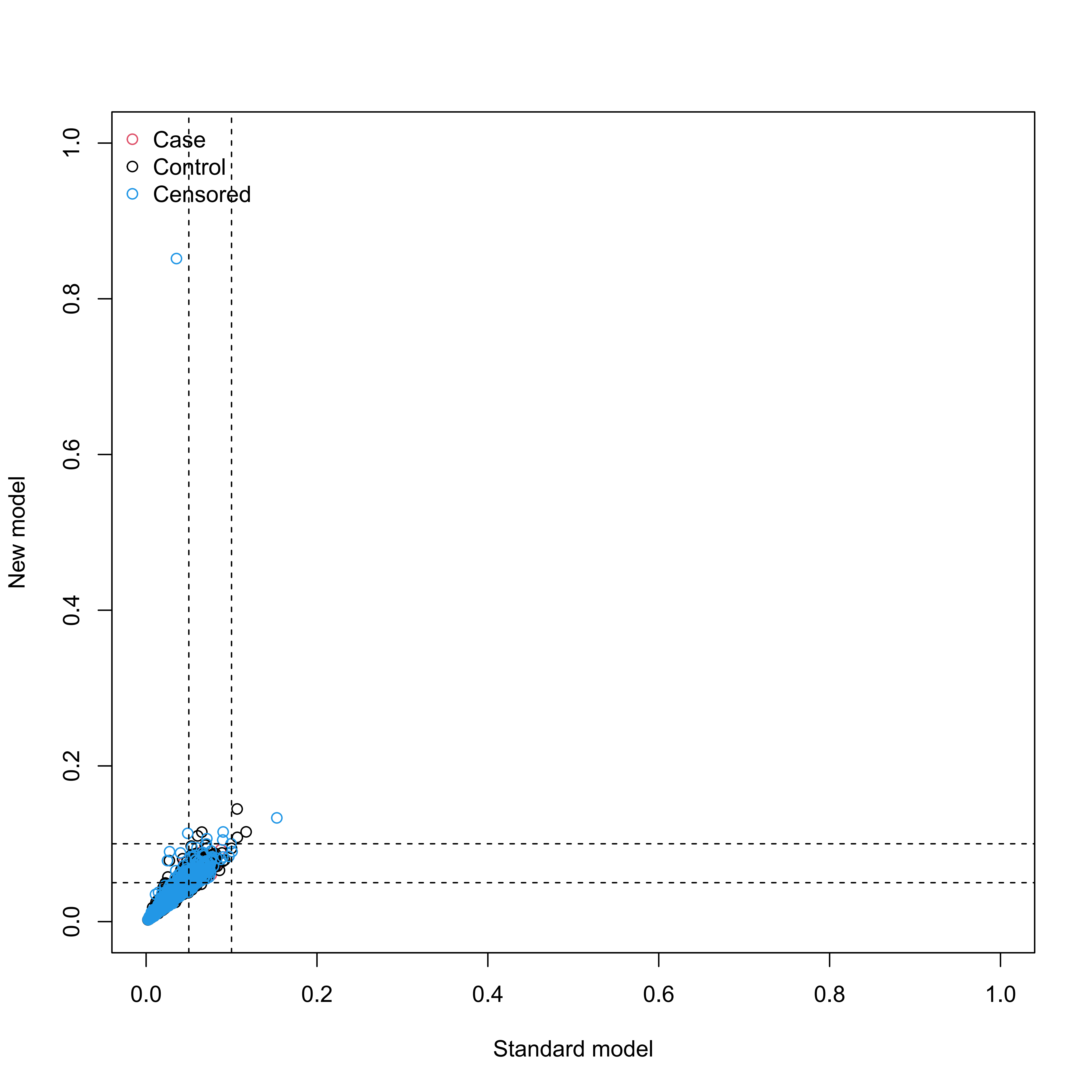

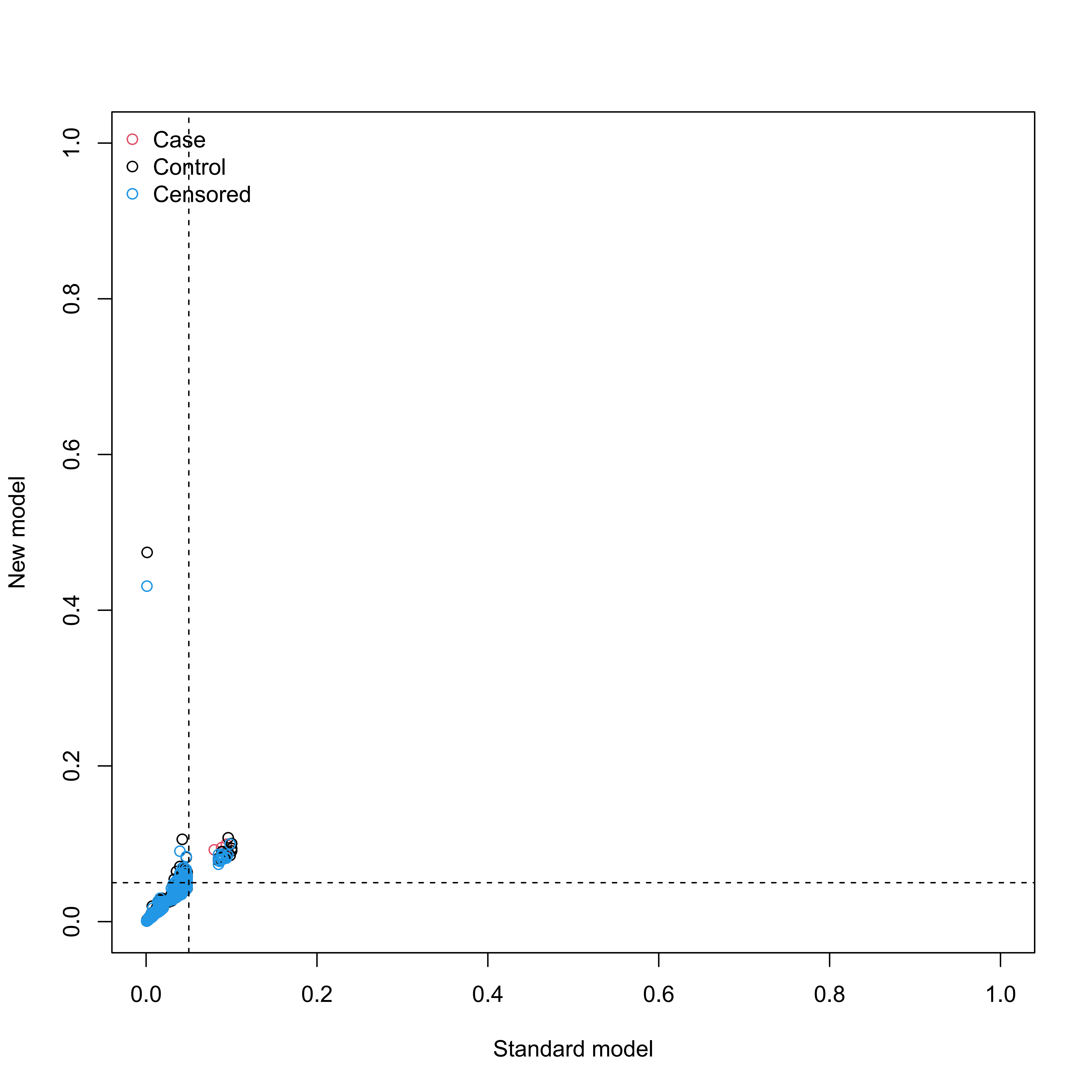

#### Supplementary Figure S17

**Reclassification in the new model for mortality with the inclusion of meta-GCIPLT score in validation set.**

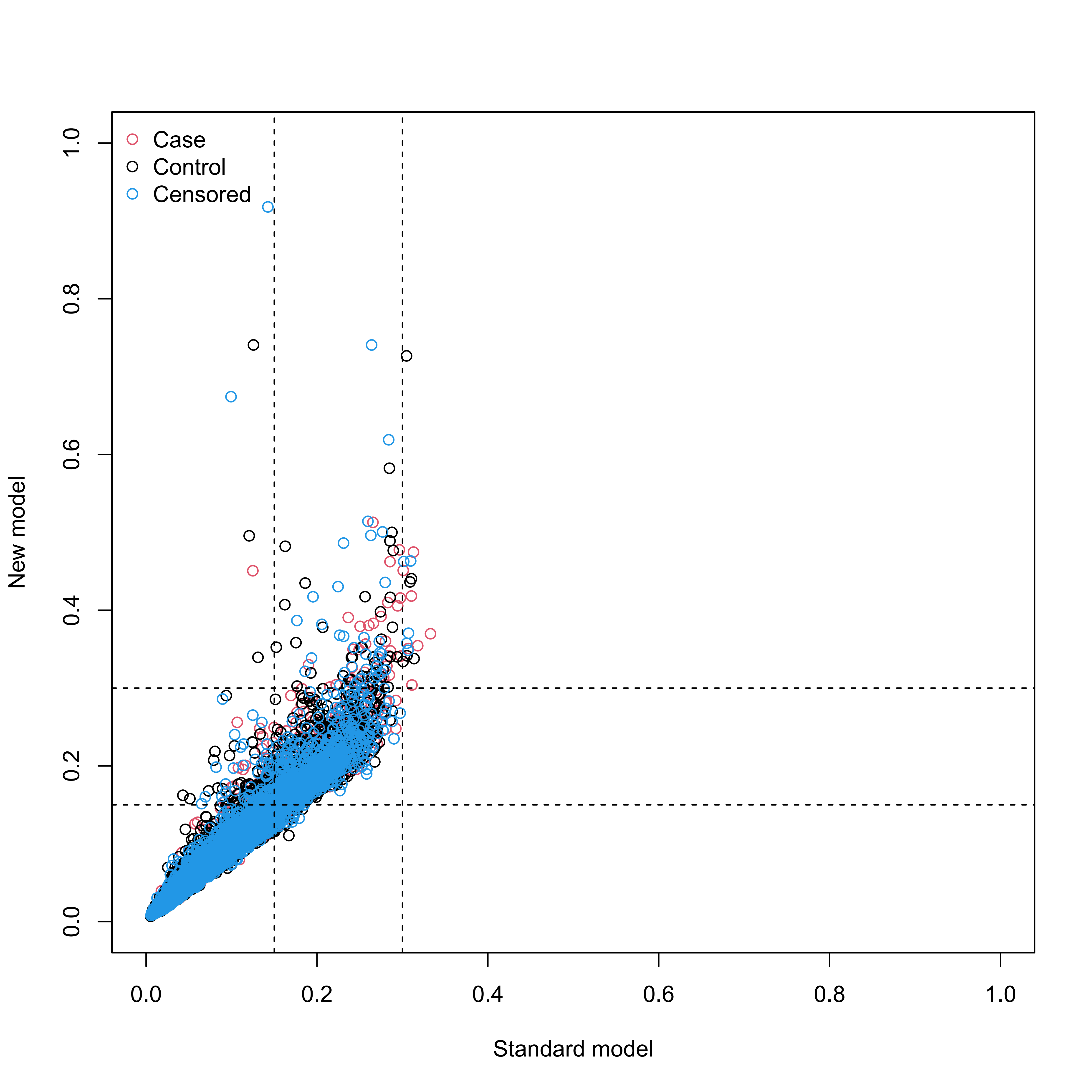

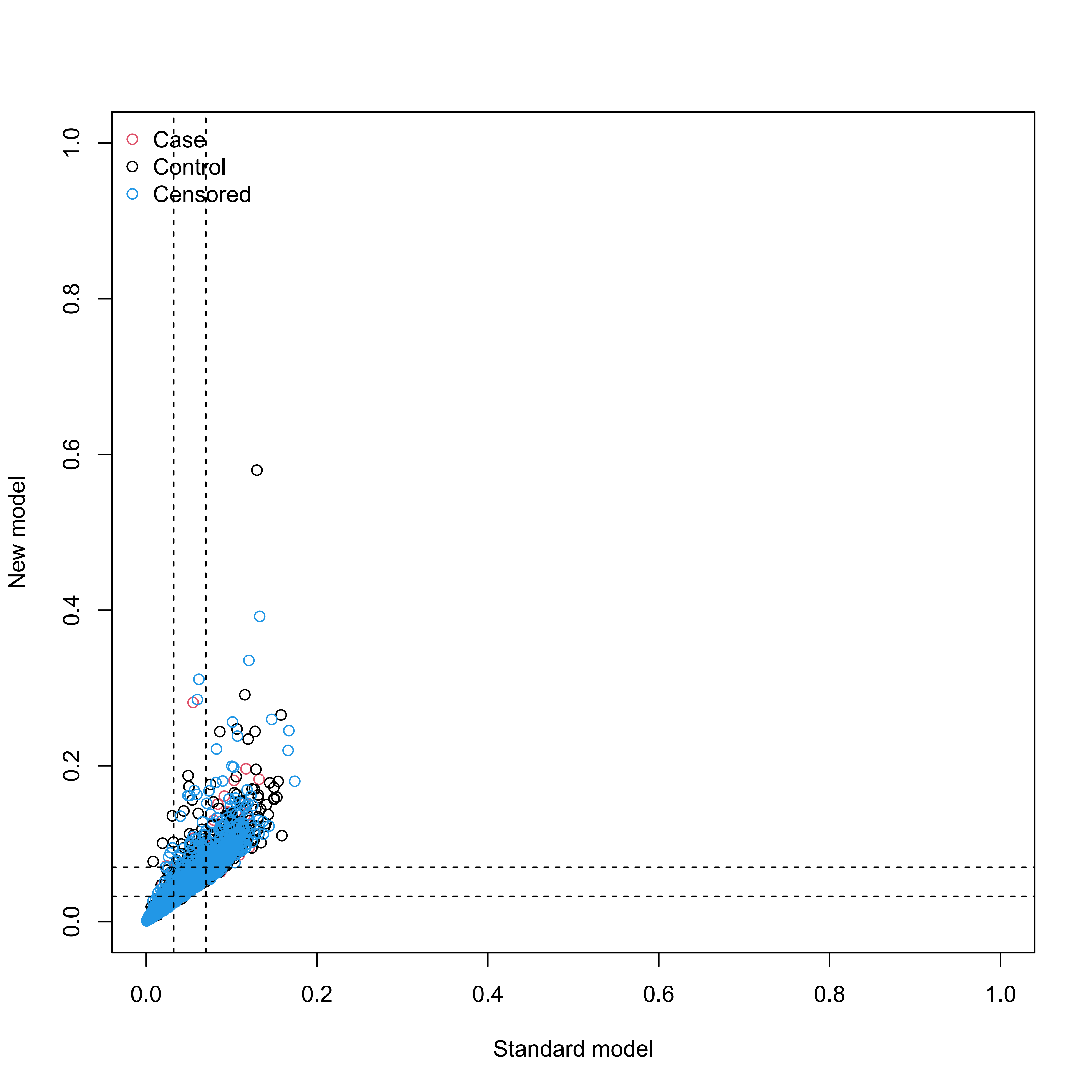

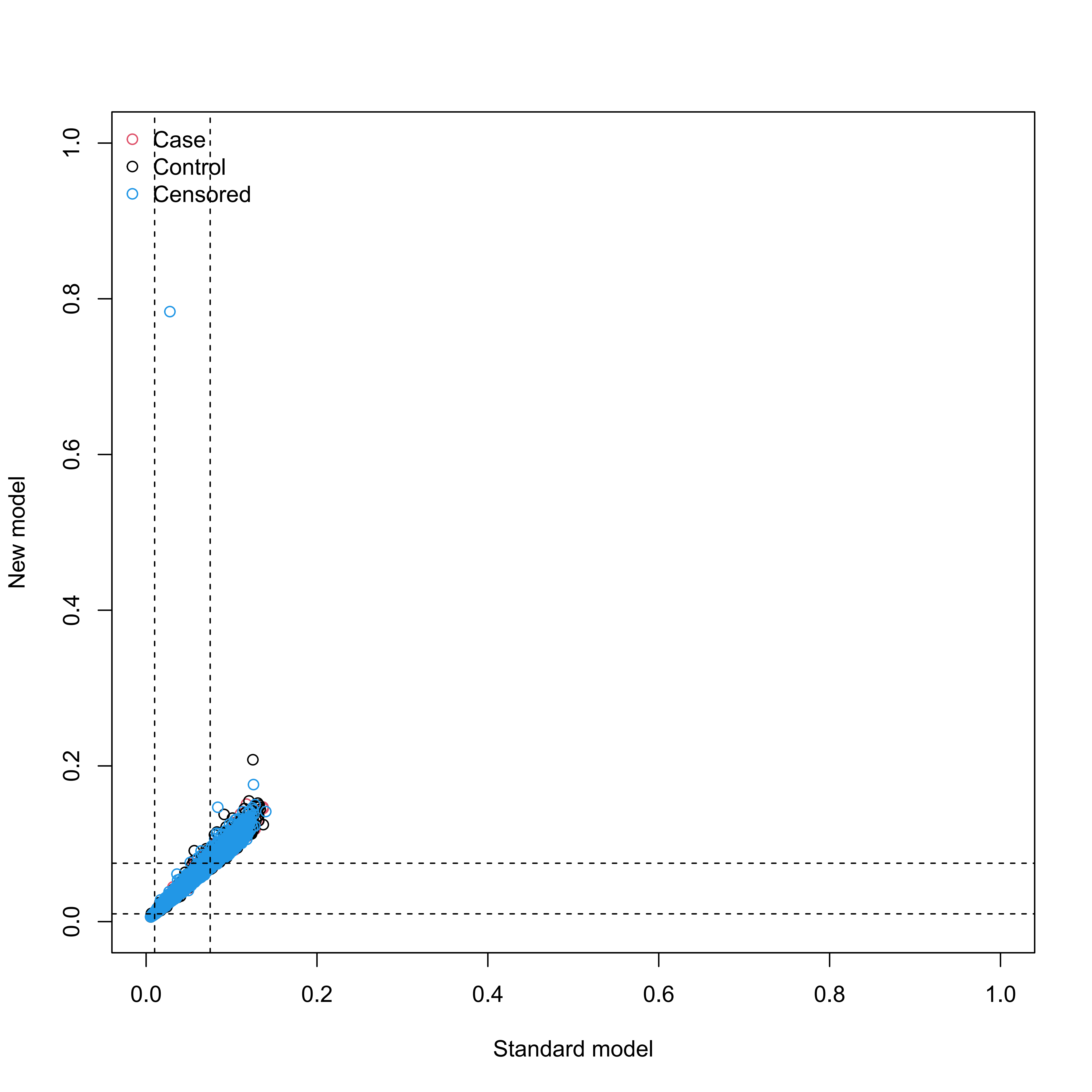

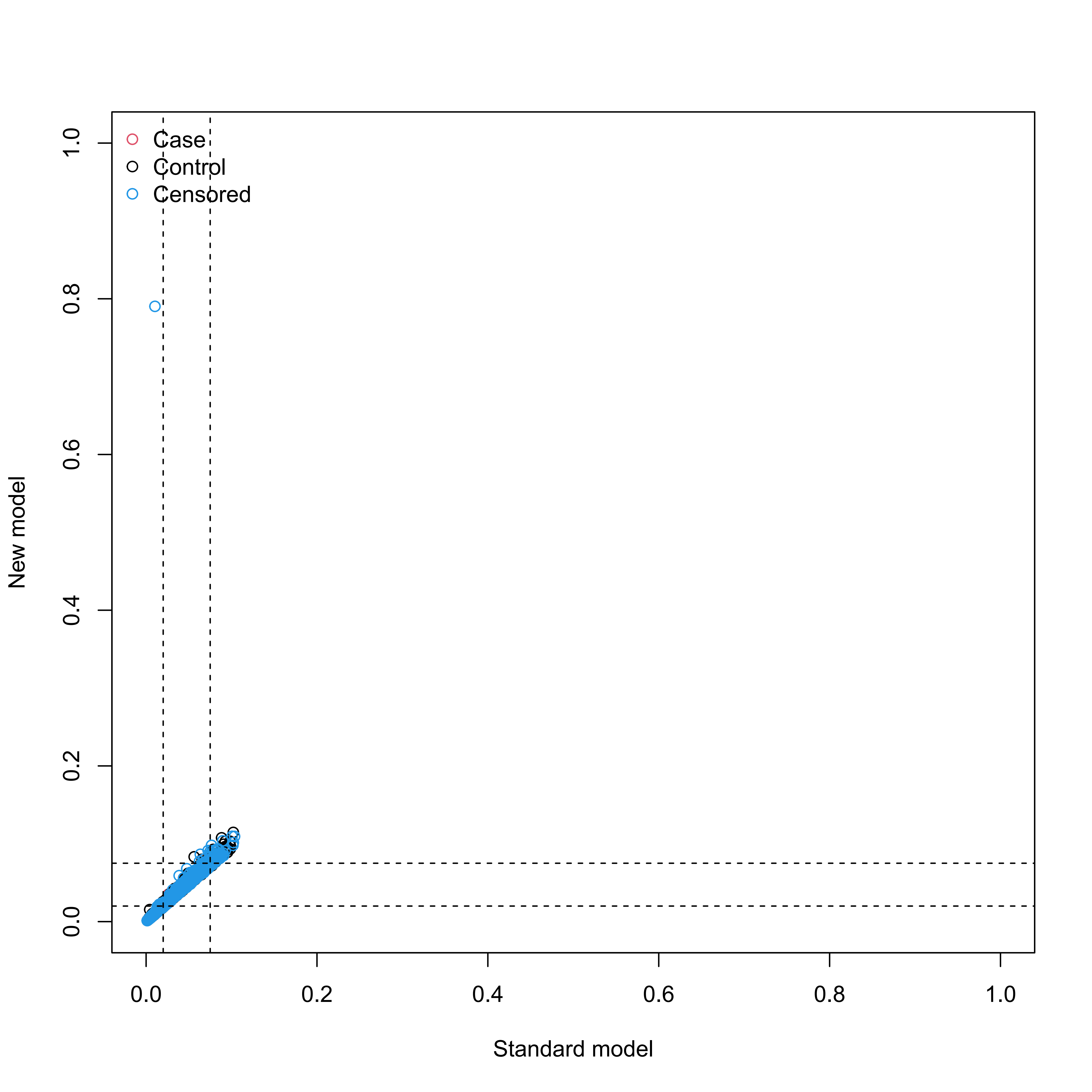

#### Supplementary Figure S18

**Calibration plots illustrating expected and observed probabilities for systemic diseases in training set.**

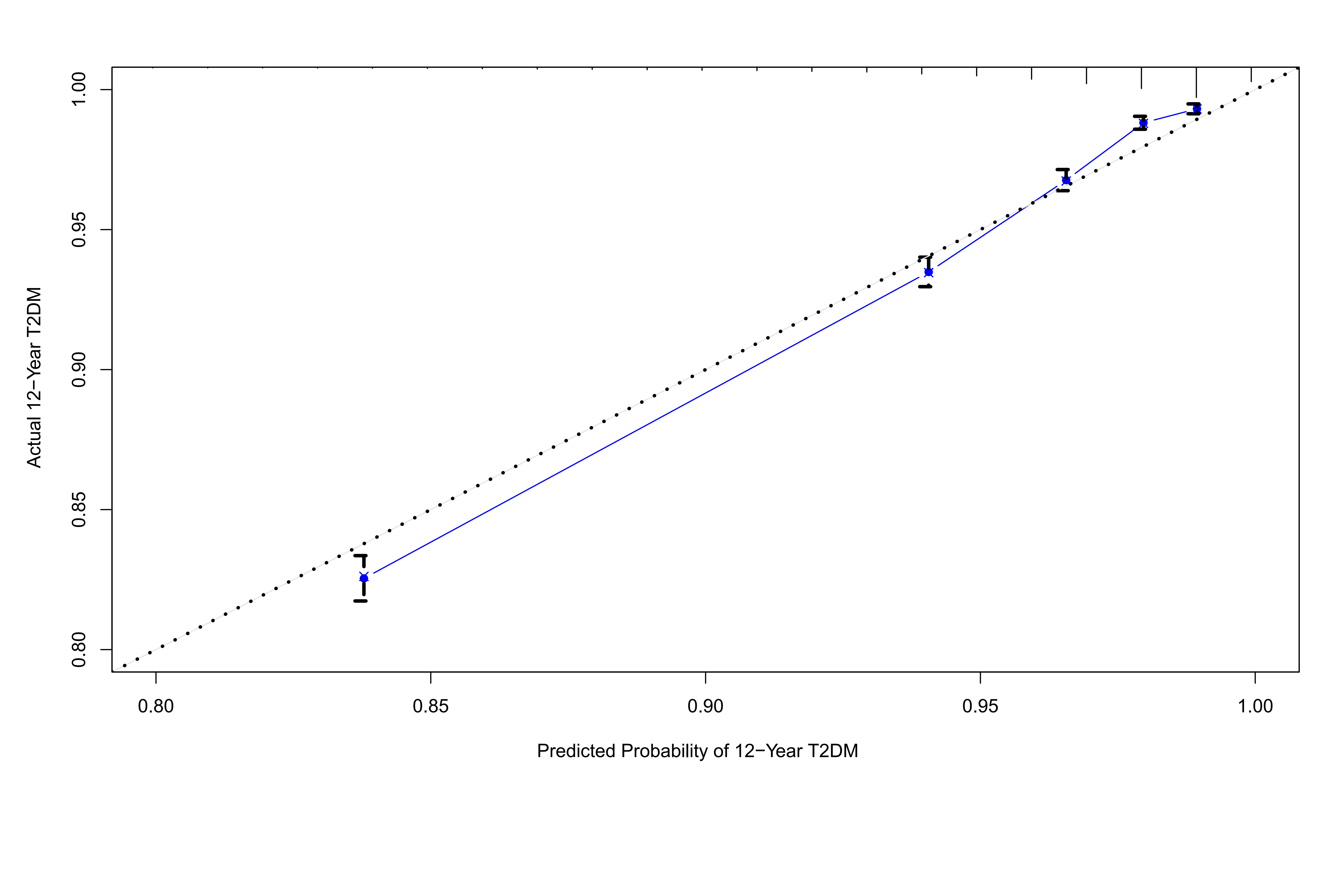

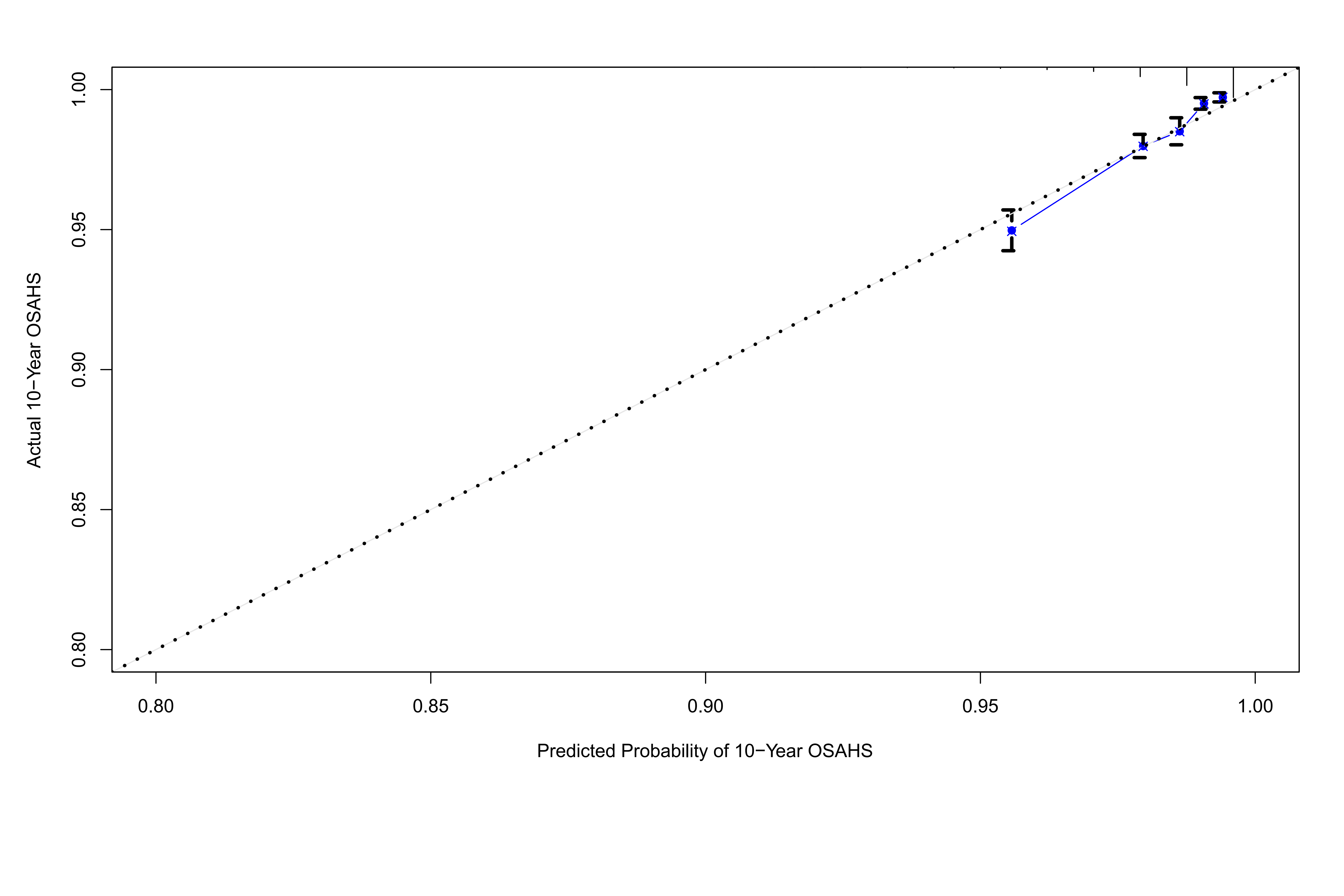

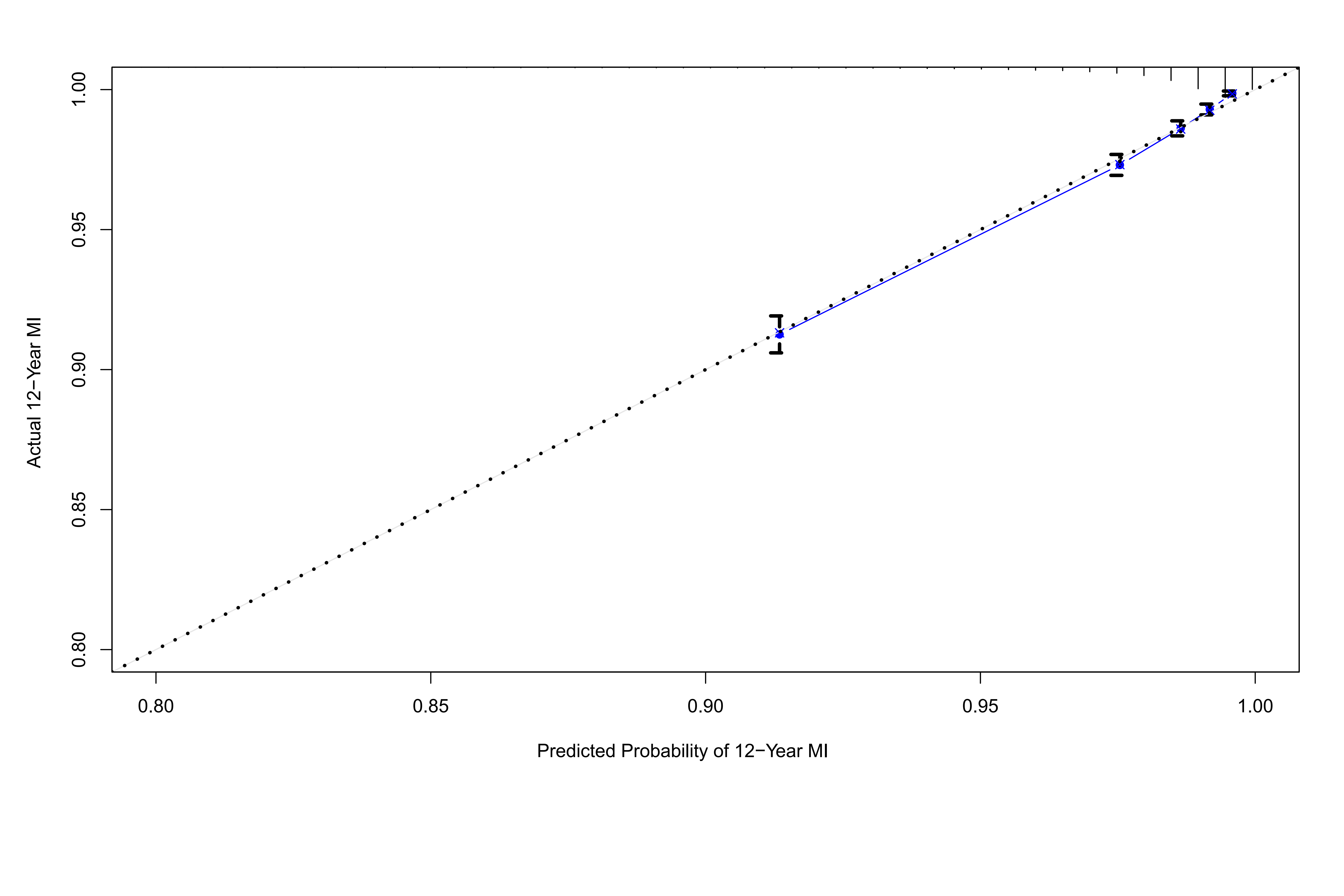

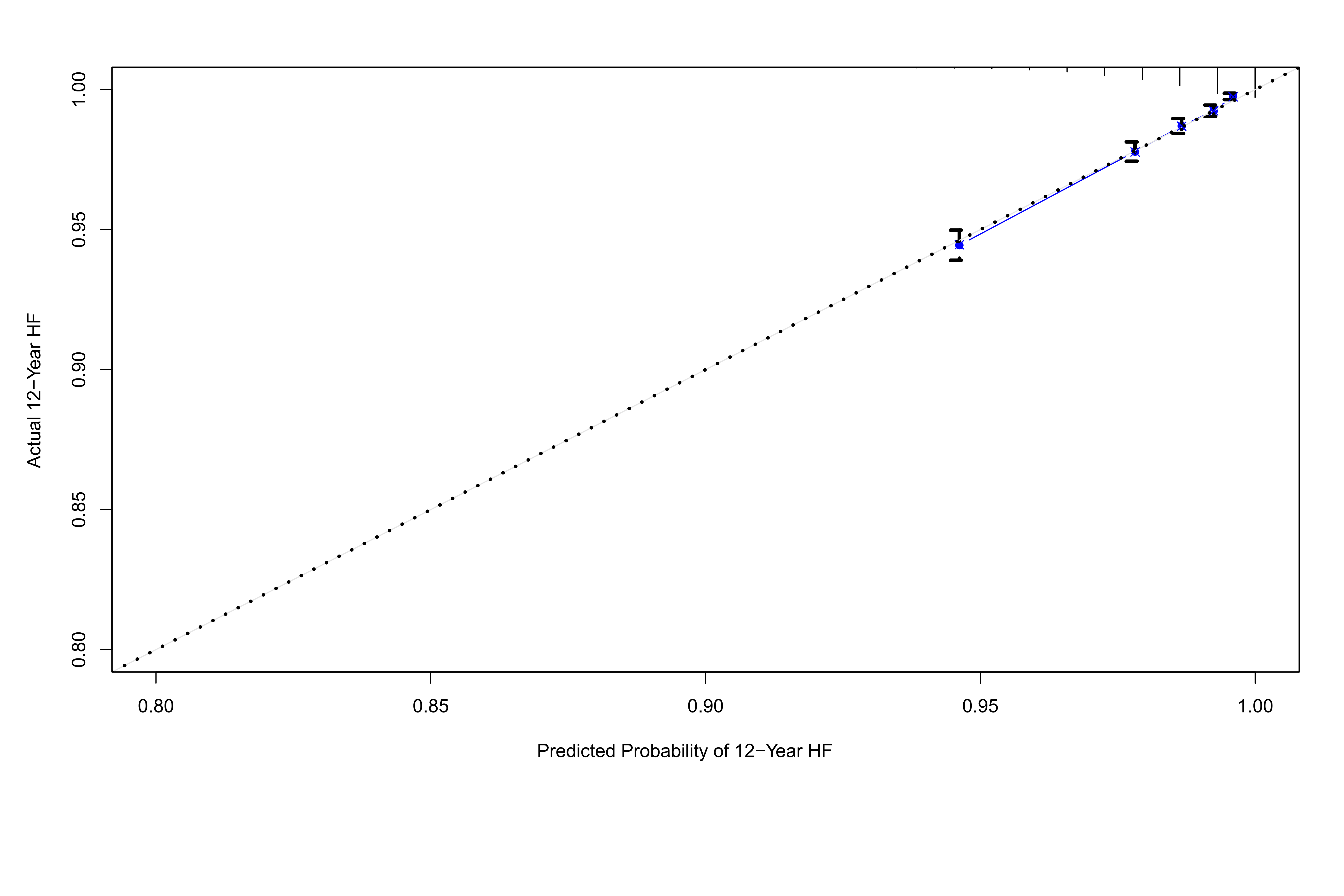

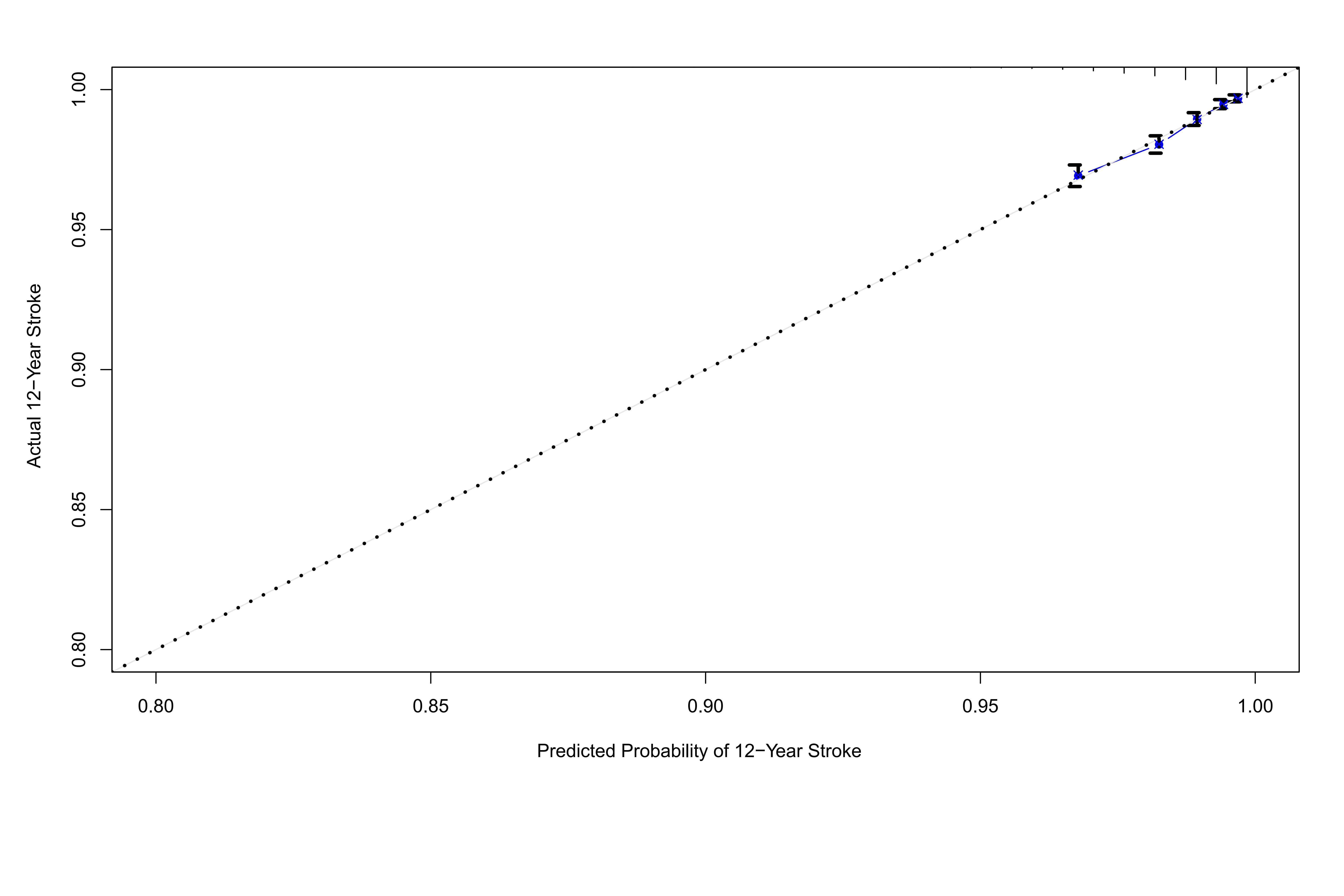

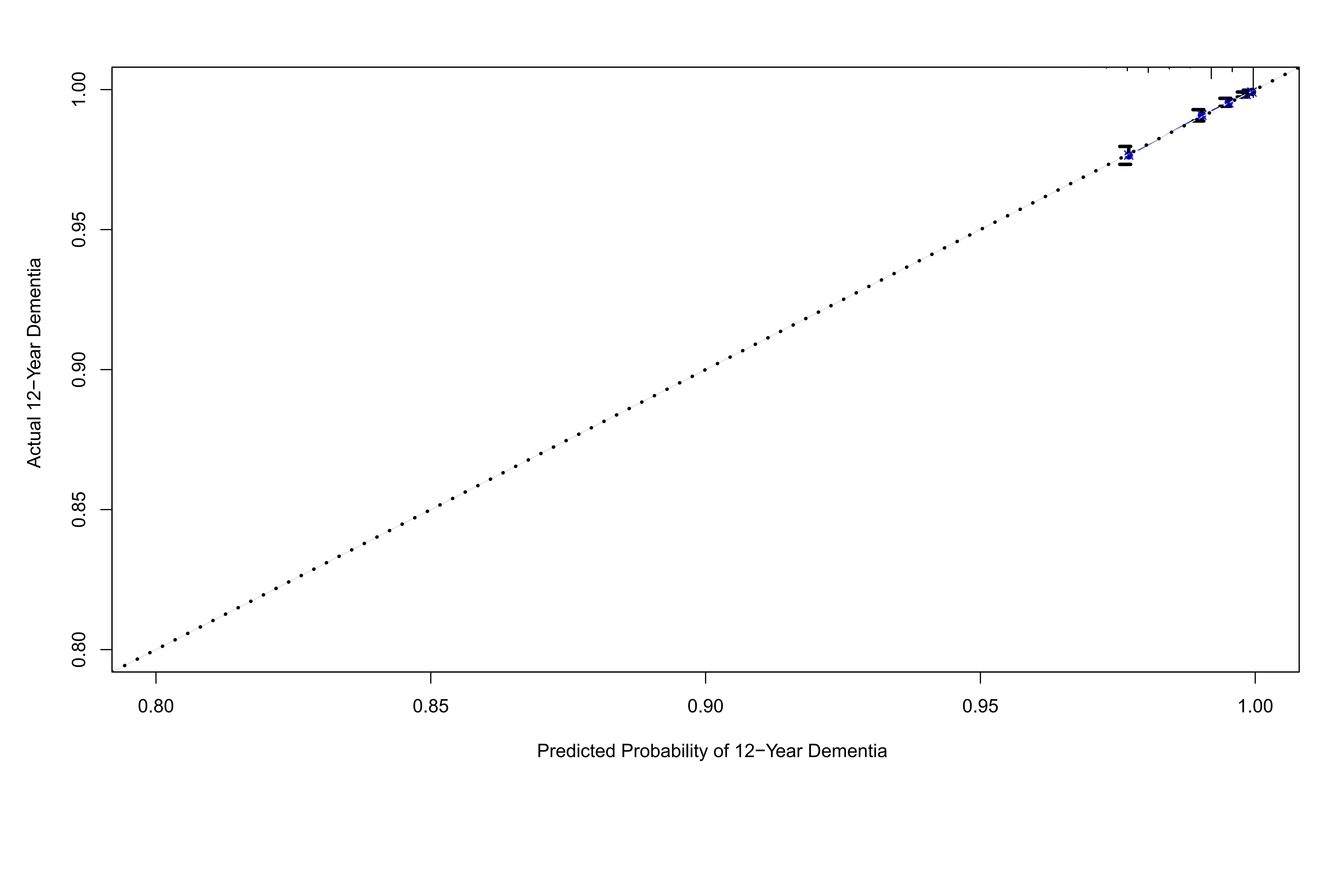

#### Supplementary Figure S19

**Calibration plots illustrating expected and observed probabilities for mortality in training set.**

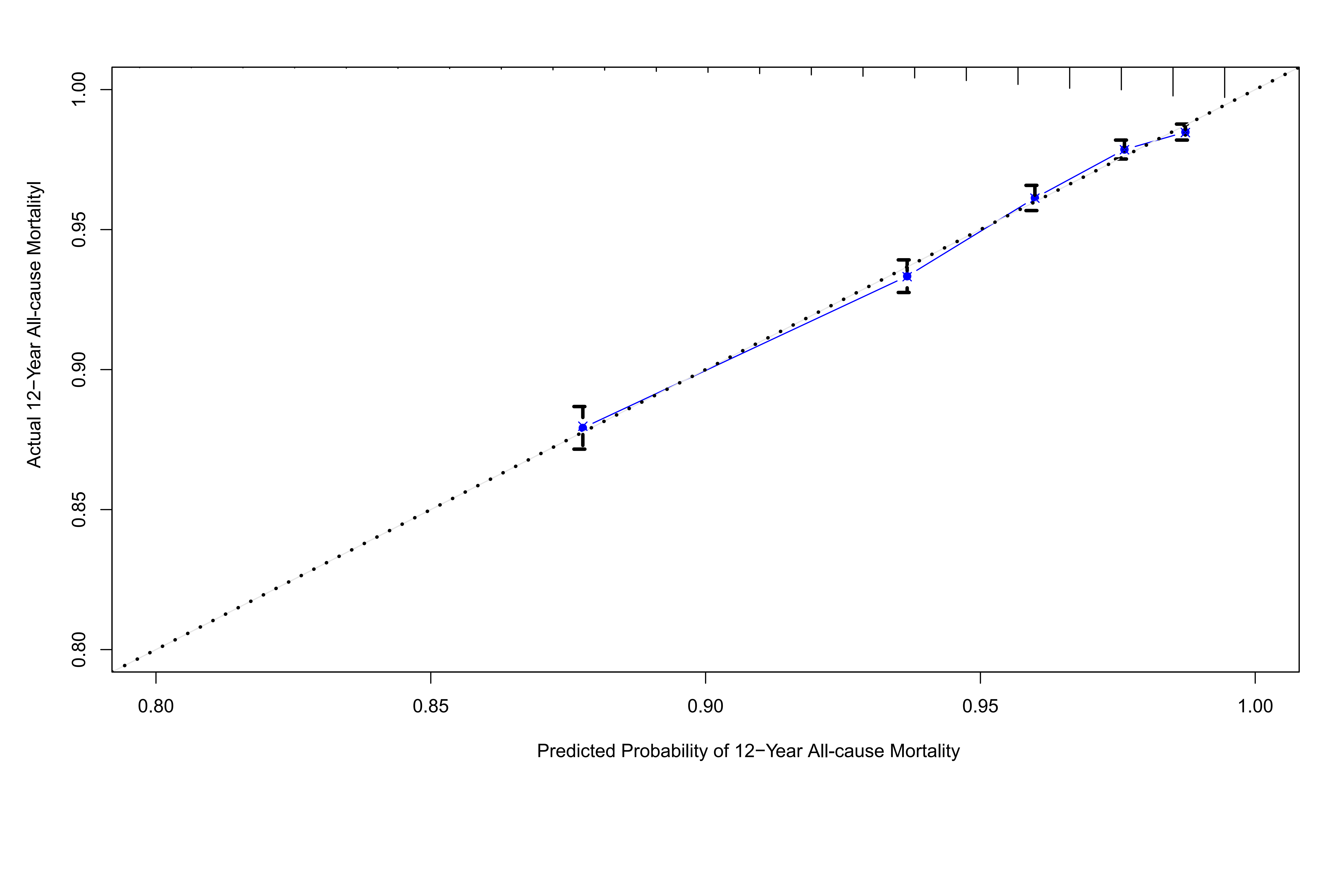

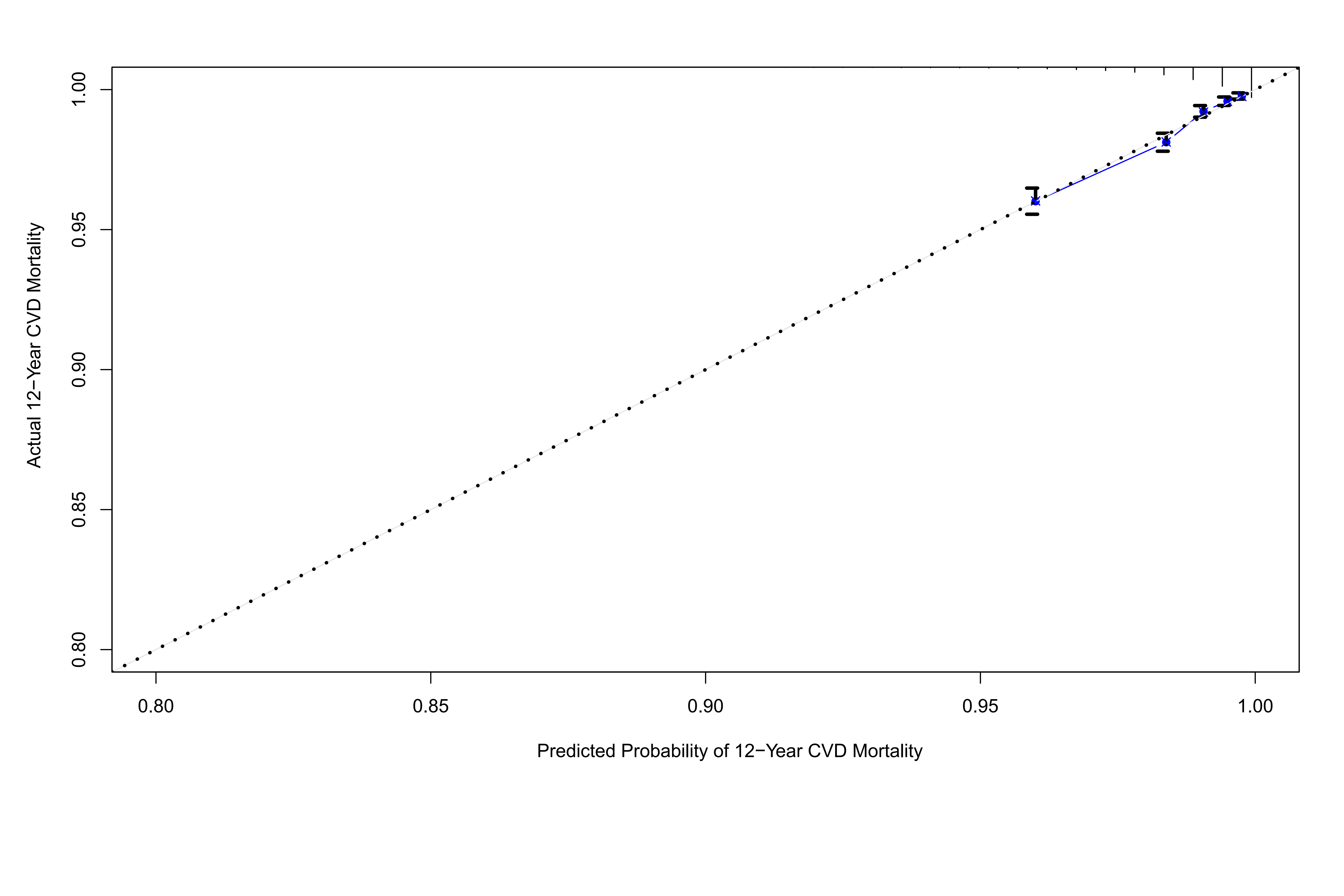

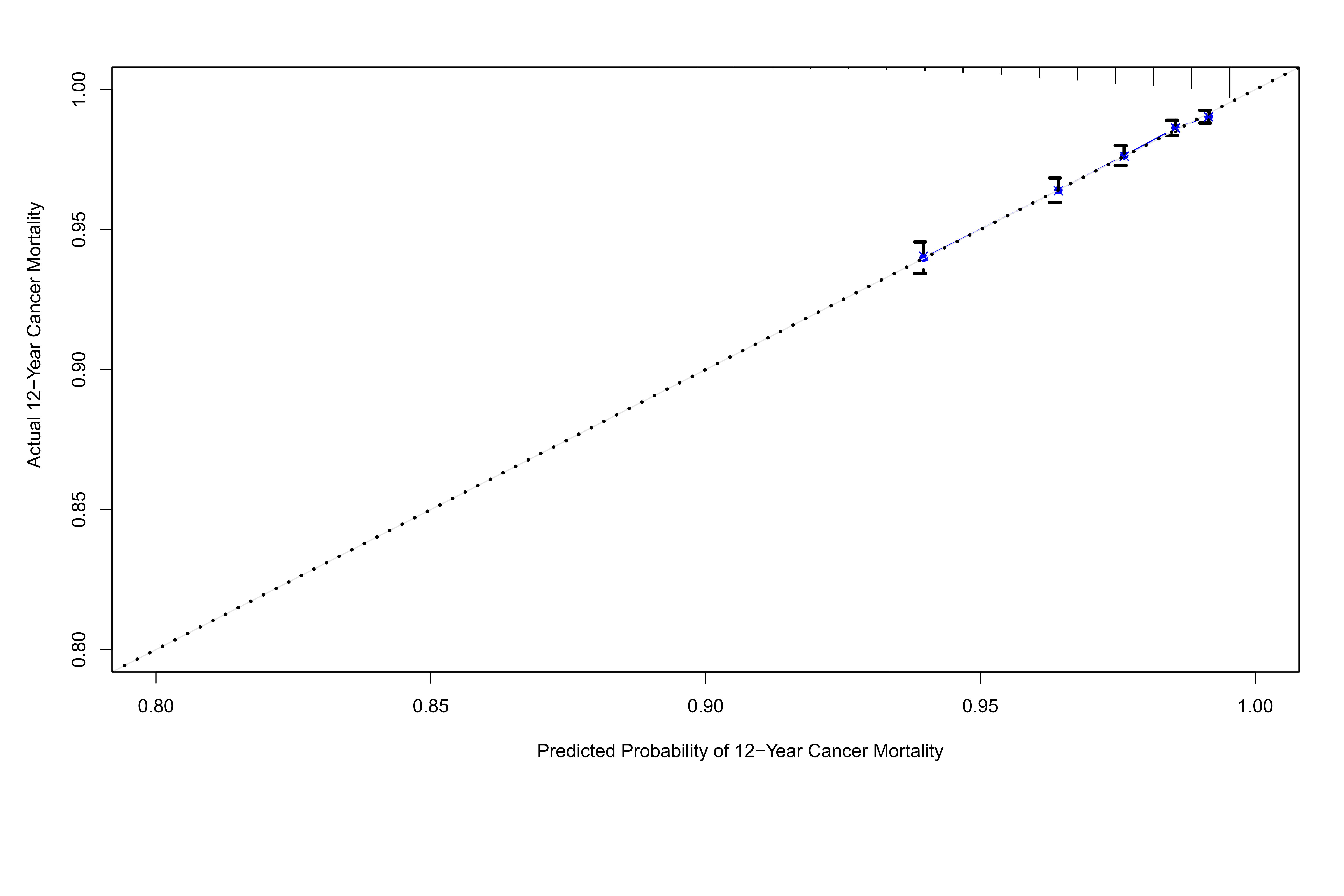

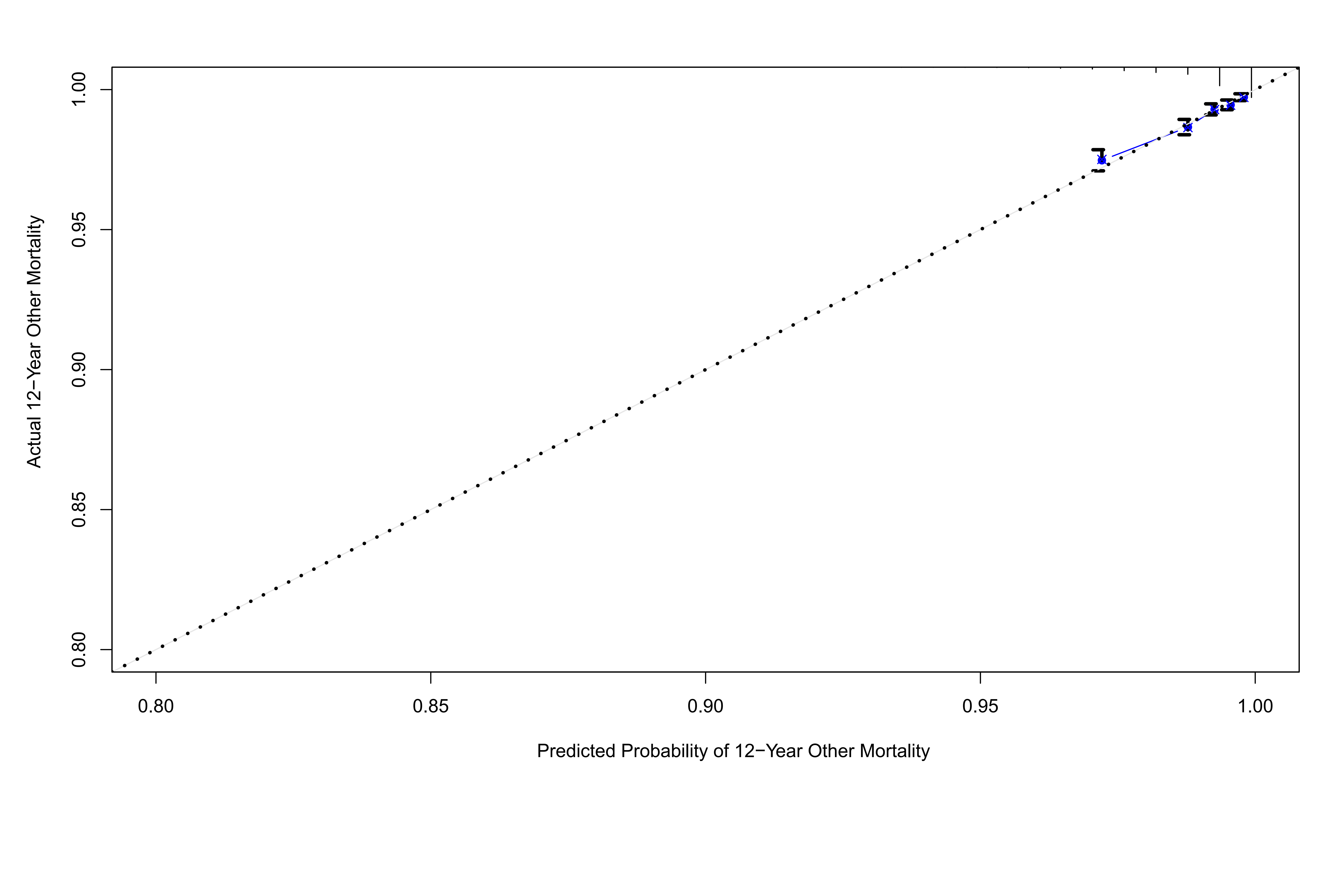

#### Supplementary Figure S20

**Calibration plots illustrating expected and observed probabilities for systemic diseases in validation set.**

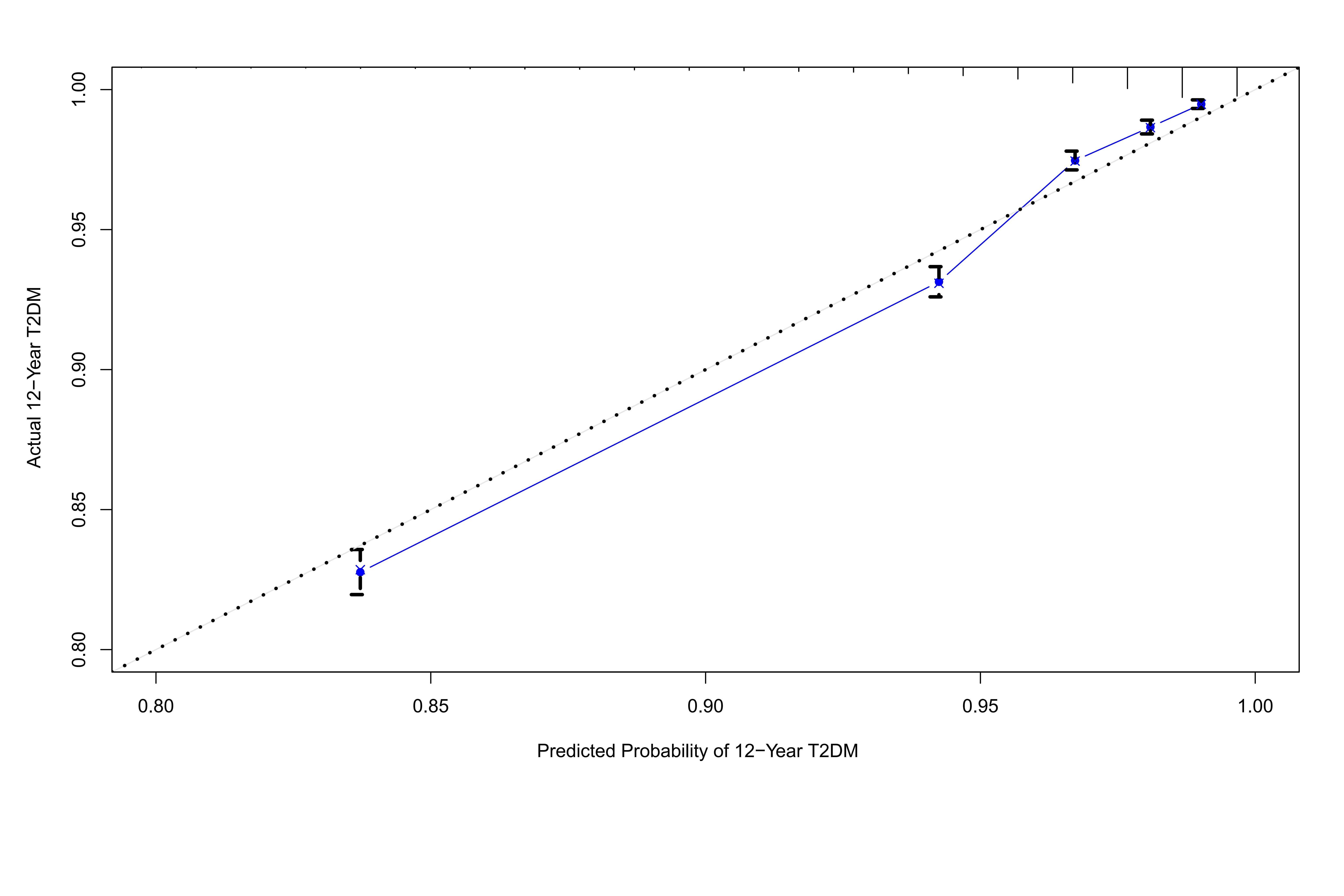

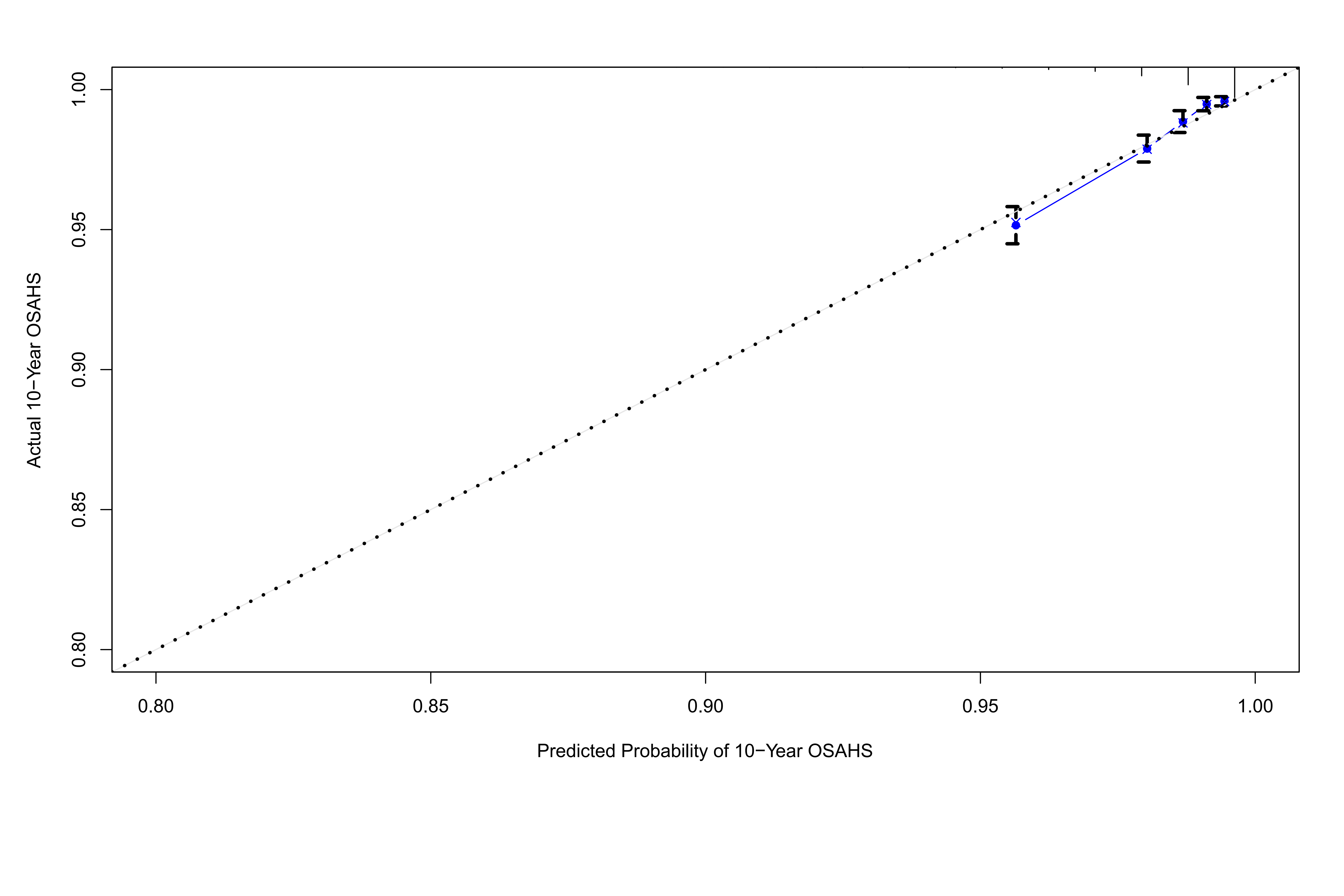

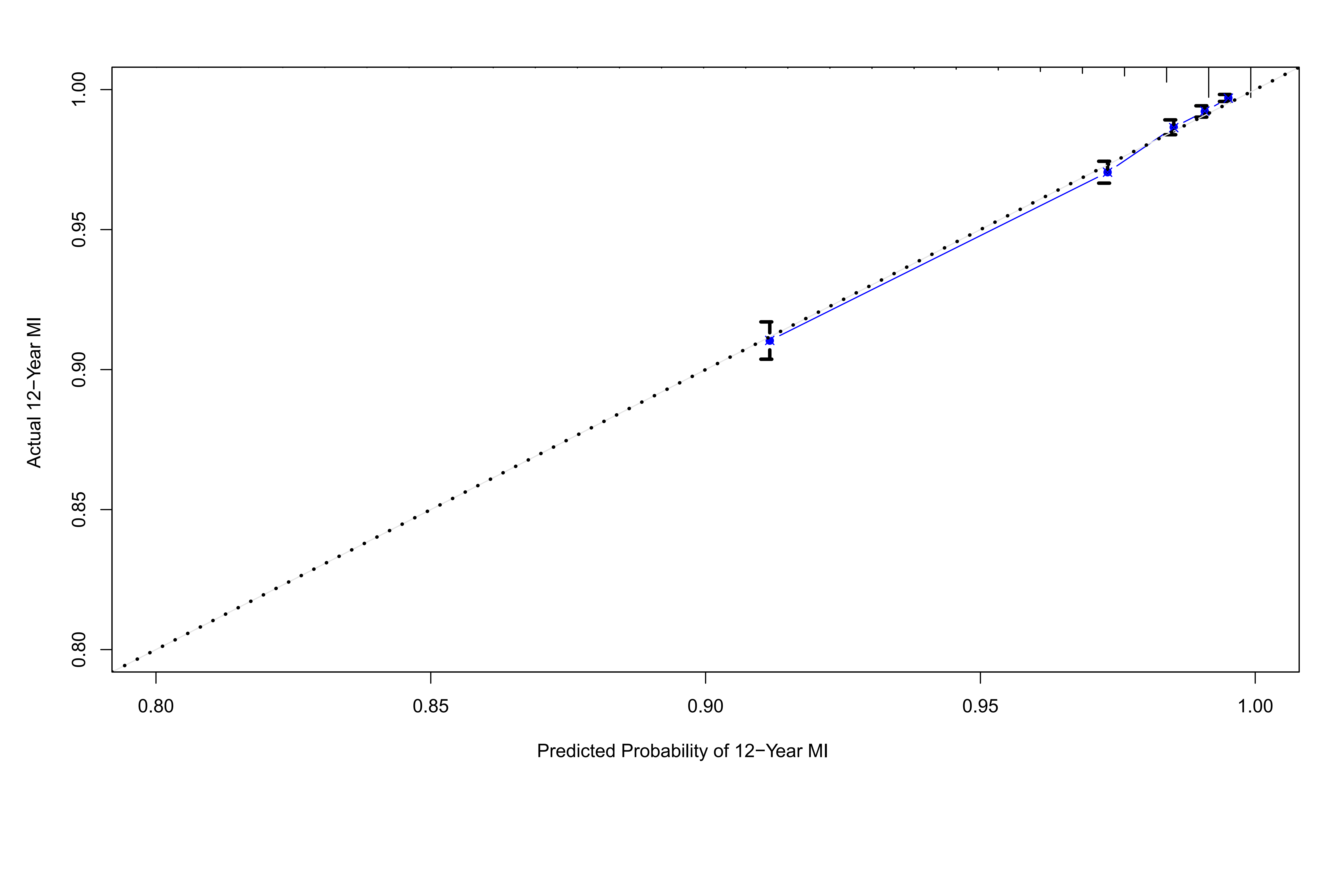

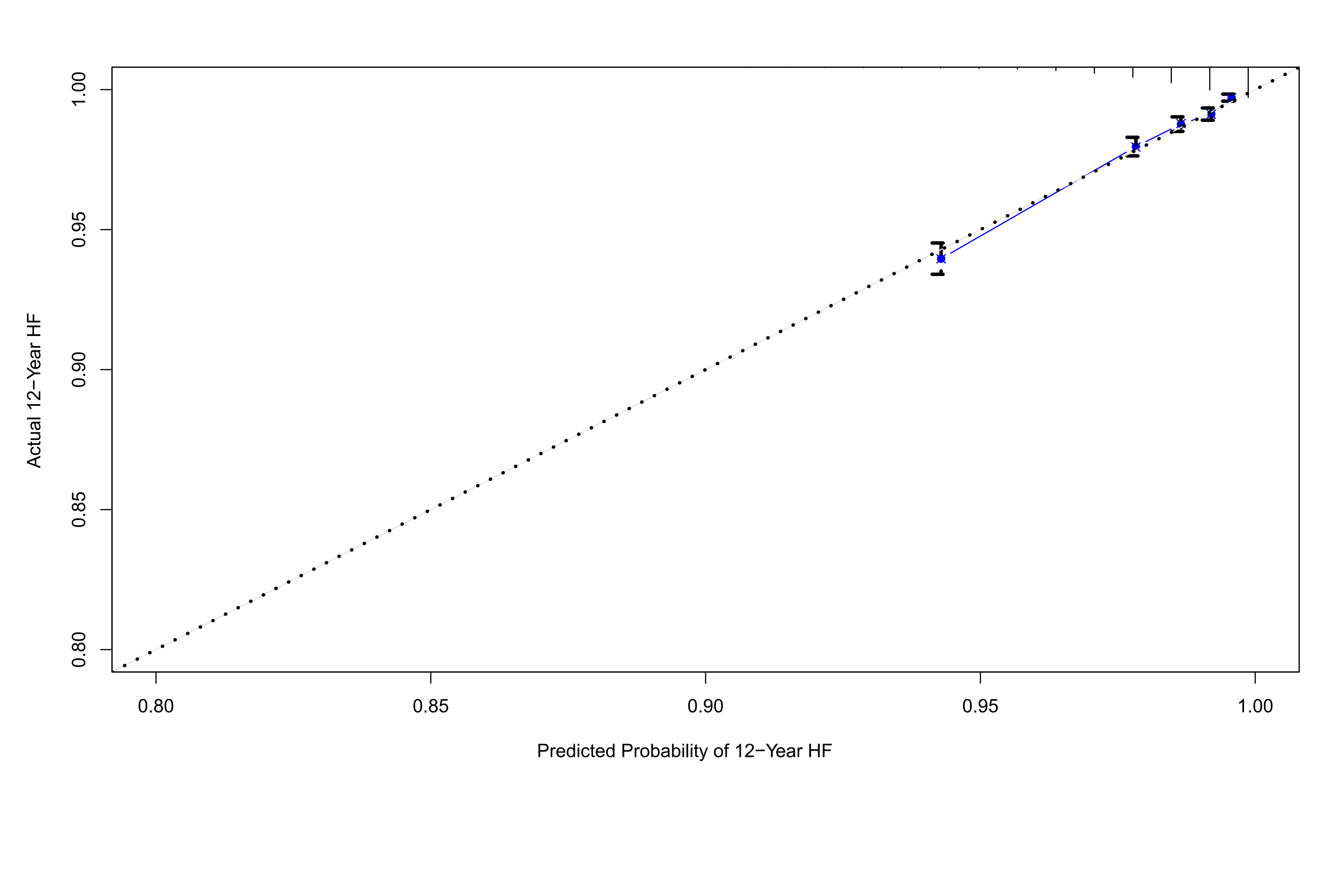

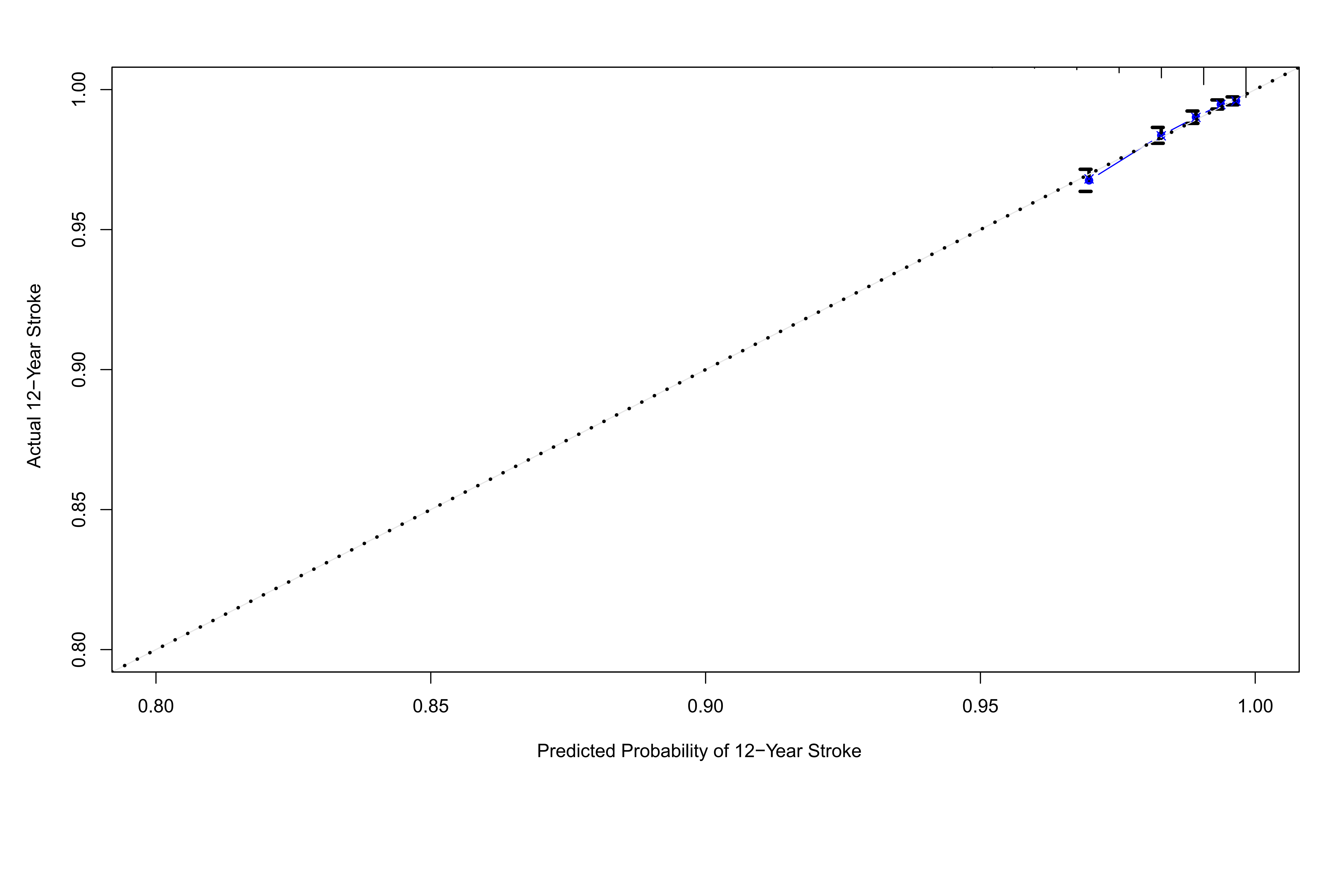

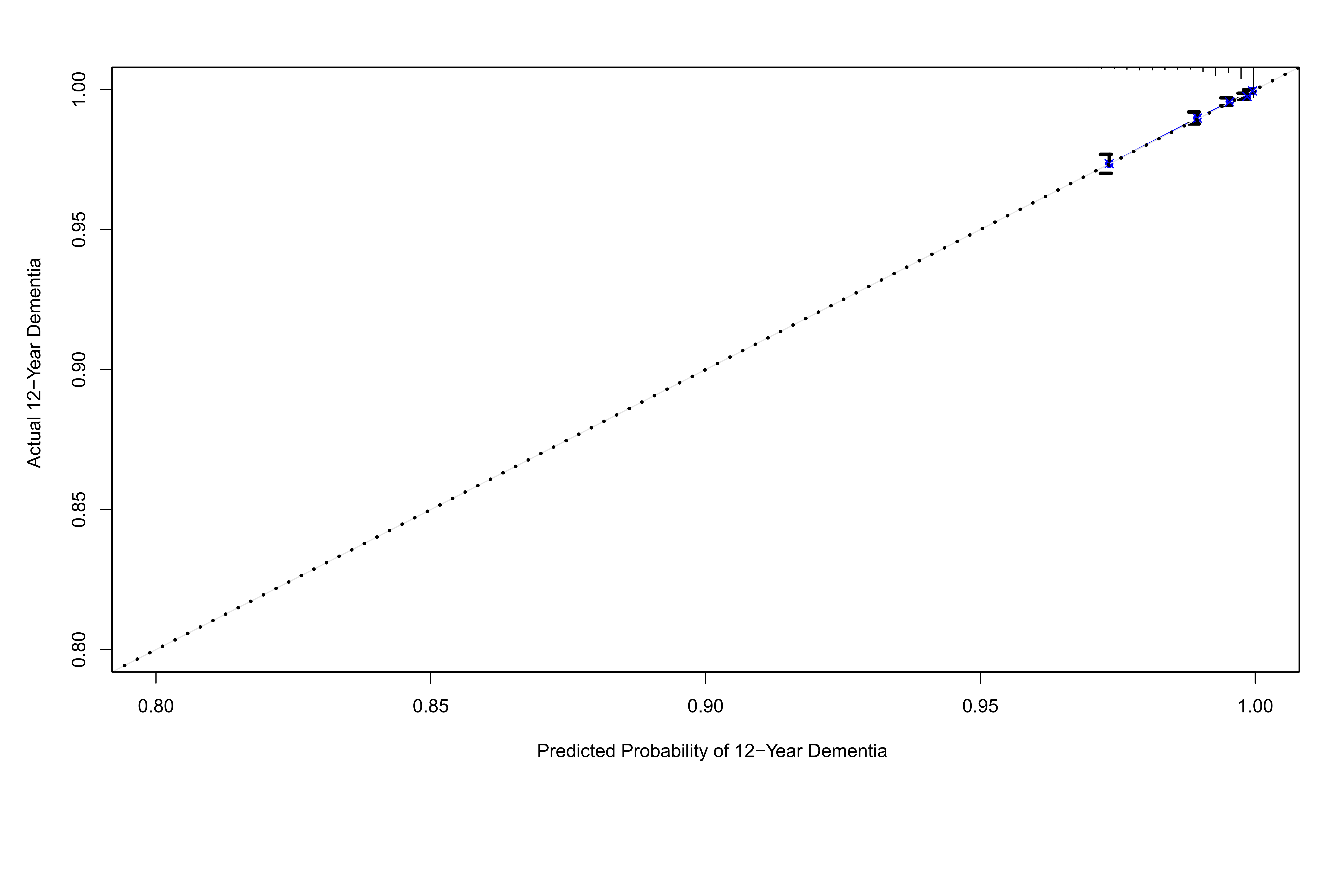

#### Supplementary Figure S21

**Calibration plots illustrating expected and observed probabilities for mortality in validation set.**

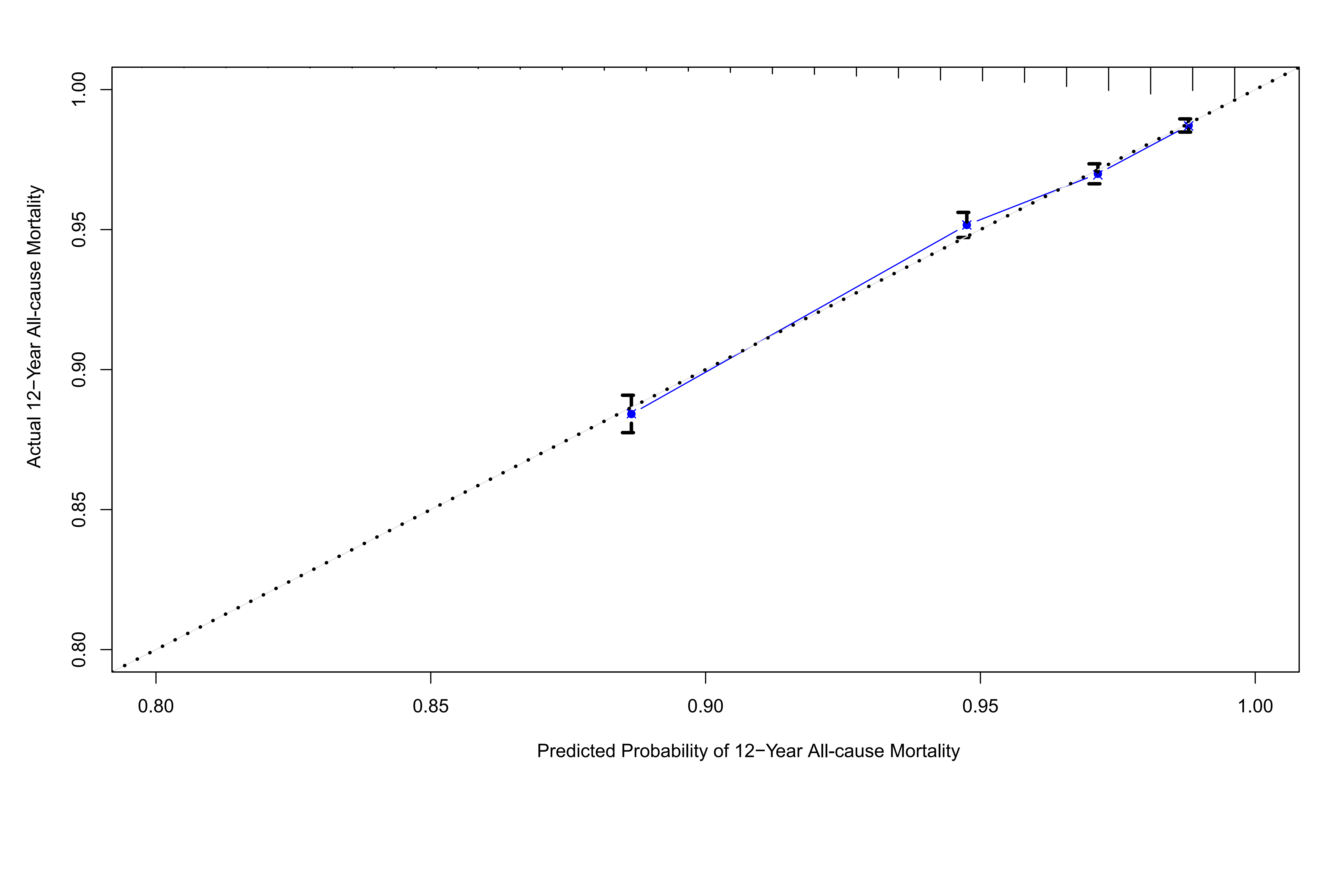

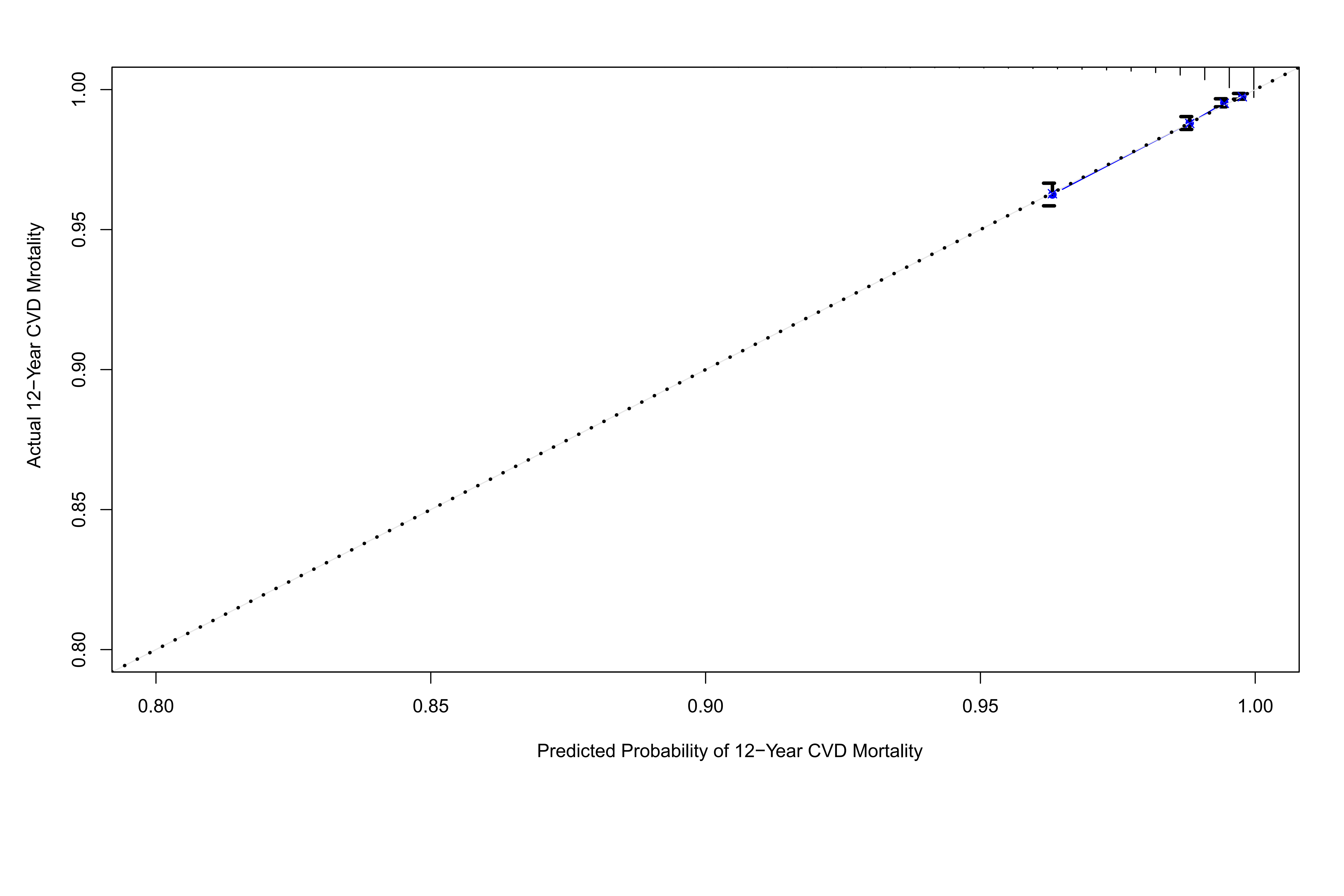

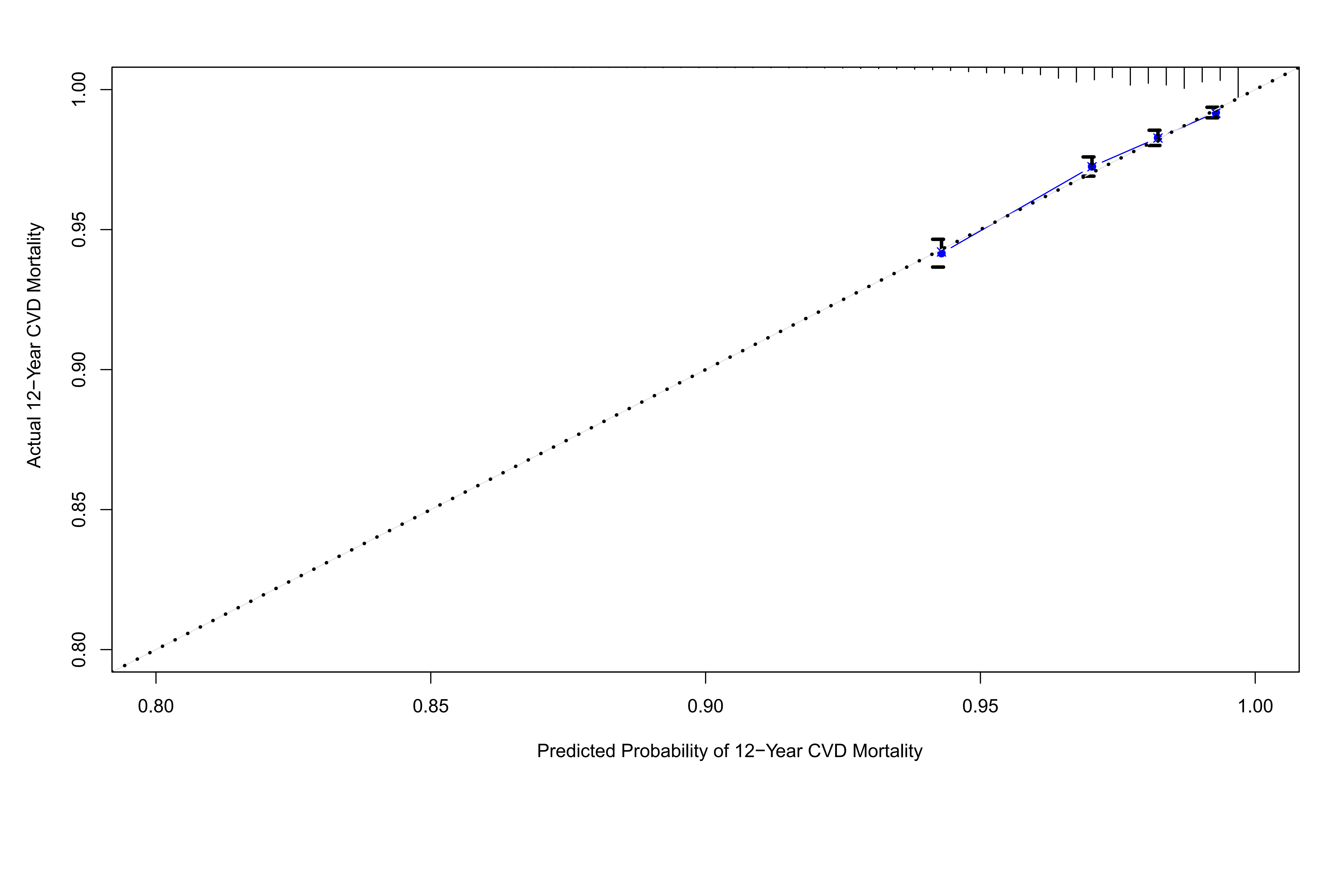

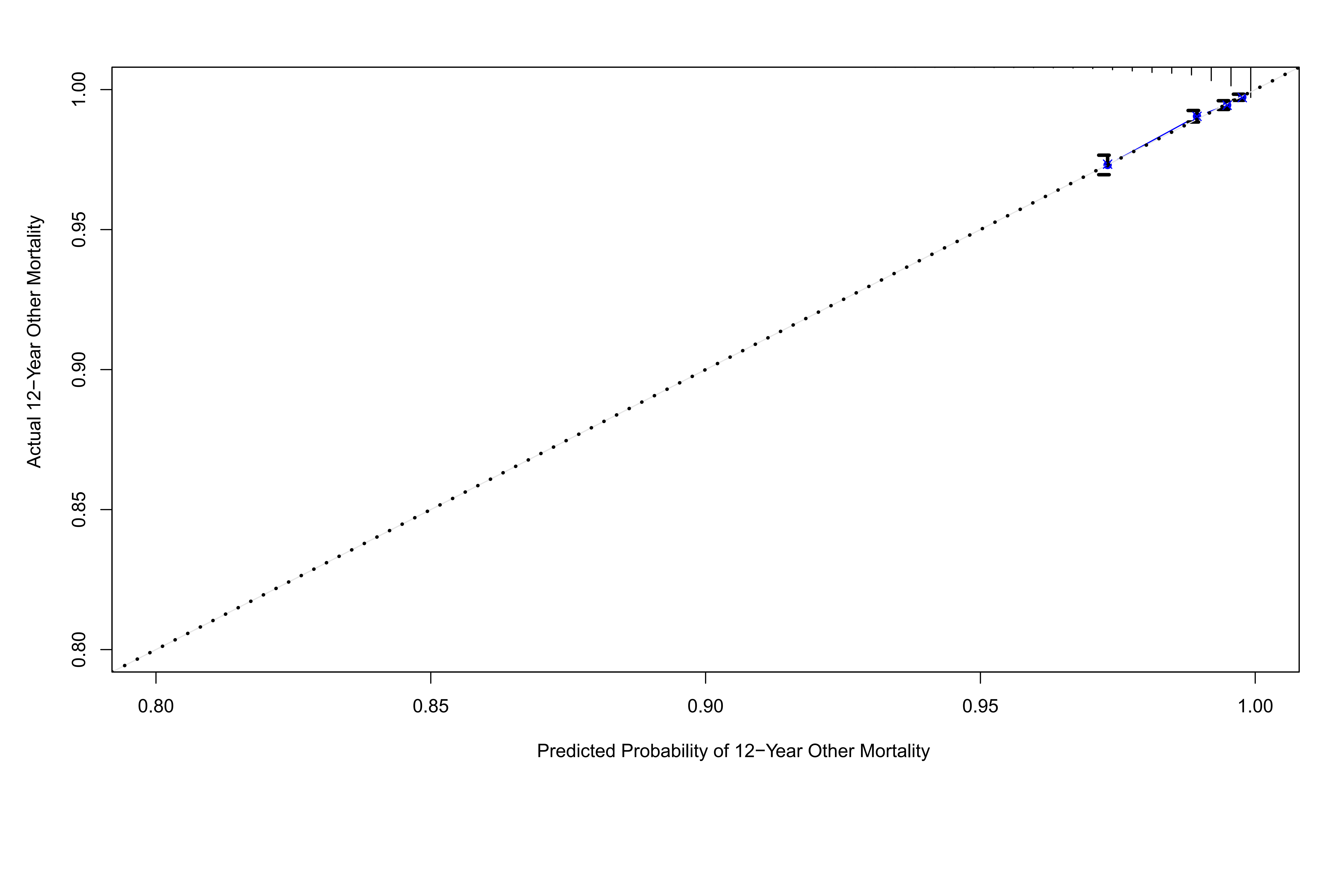

### SUPPLEMENTARY TABLES

#### Supplementary Table S1

**Definition of variables in touchscreen questionnaire, verbal interview and inpatient records of diagnosis.**

| **Variable** | **Data category** | **Data field** | **Data code** |
| --- | --- | --- | --- |
| Age at recruitment | Population characteristics | 21022 |  |
| Sex | Population characteristics | 31 |  |
| Townsend deprivation index | Population characteristics | 189 |  |
| Average total household income before tax | Touchscreen questionnaire | 738 |  |
| Smoking status | Touchscreen questionnaire | 20116 |  |
| Alcohol drinker status | Touchscreen questionnaire | 20117 |  |
| Ethnic background | Touchscreen questionnaire | 21000 |  |
| Qualifications | Touchscreen questionnaire | 6138 |  |
| BMI (Body mass index) | Physical measures | 21001 |  |
| Waist circumference | Physical measures | 48 |  |
| Hip circumference | Physical measures | 49 |  |
| HDL cholesterol | Blood assays | 30760 |  |
| LDL direct | Blood assays | 30780 |  |
| Glycated haemoglobin (HbA1c) | Blood biochemistry | 30750 |  |
| Haemoglobin concentration | Blood count | 30020 |  |
| Creatinine (enzymatic) in urine | Urine assays | 30510 |  |
| Microalbumin in urine | Urine assays | 30500 |  |
| logMAR, final (left) | Eye measures | 5208 |  |
| logMAR, final (right) | Eye measures | 5201 |  |
| Cylindrical power (left) | Eye measures | 5086 |  |
| Cylindrical power (right) | Eye measures | 5087 |  |
| Spherical power (left) | Eye measures | 5085 |  |
| Spherical power (right) | Eye measures | 5084 |  |
| Intra-ocular pressure,  corneal-compensated (left) | Eye measures | 5262 |  |
| Intra-ocular pressure,  corneal-compensated (right) | Eye measures | 5254 |  |
| Average ganglion cell-inner plexiform layer thickness (left) | Eye measures | 28504 |  |
| Average ganglion cell-inner plexiform layer thickness (right) | Eye measures | 28505 |  |
| Diabetes | Hospital inpatients (ICD-10) | 41270 | E11-E14 |
|  | Touchscreen questionnaire | 6148 | 1 |
|  | Touchscreen questionnaire | 2443 | 1 |
|  | Verbal interview | 20002 | 1276, 1220, 1222, 1223, 1468, 1521, 1607 |
| Date of diabetes first reported | Health-related outcomes | 2404 | 130706, 130708, 130710, 130712. 130714 |
| Type 2 diabetes | Hospital inpatients (ICD-10) | 41270 | E11 |
|  | Verbal interview | 20002 | 1223 |
| Date of type 2 diabetes first reported | Health-related outcomes | 2404 | 130709 |
| Cardiovascular diseases | Hospital inpatients (ICD-10) | 41270 | I20-I25, I50, I60-I64 |
|  | Touchscreen questionnaire | 6150 | 1, 2, 3 |
| Date of Cardiovascular diseases first reported | Health-related outcomes | 2409 | 131296, 131298, 131300, 131302, 131304, 131306, 131360, 131362, 131364, 131366, 131368 |
|  | Verbal interview | 20002 | 1074, 1075, 1076, 1081,  1082, 1086, 1491, 1583 |
| Hypertension | Hospital inpatients (ICD-10) | 41270 | I10, I15 |
|  | Touchscreen questionnaire | 6150 | 4 |
|  | Verbal interview | 20002 | 1065, 1072 |
| Date of hypertension first reported | Health-related outcomes | 2409 | 131286, 131290, 131292, 131294 |
| Glaucoma | Hospital inpatients (ICD-10) | 41270 | H40, H42 |
|  | Touchscreen questionnaire | 6148 | 2 |
|  | Verbal interview | 20002 | 1277 |
| Retinal diseases | Hospital inpatients (ICD-10) | 41270 | H30-H36 |
|  | Touchscreen questionnaire | 6148 | 1, 5 |
|  | Verbal interview | 20002 | 1275, 1281, 1282, 1528 |
| Multiple sclerosis | Hospital inpatients (ICD-10) | 41270 | G35 |
|  | Verbal interview | 20002 | 1261 |
| Dementia | Hospital inpatients (ICD-10) | 41270 | F00-F03, F05.1, G30, G31.0, G31.1, G31.8, I67.3, A81.0, F10.6 |
|  | Verbal interview | 20002 | 1263 |
| Date of dementia first reported | Health-related outcomes | 2406 | 131038 |
| Heart failure | Hospital inpatients (ICD-10) | 41270 | I42, I50 |
|  | Verbal interview | 20002 | 1076, 1079, 1588 |
| Date of heart failure first reported | Health-related outcomes | 2409 | 131354 |
| Myocardial infarction | Hospital inpatients (ICD-10) | 41270 | I20-I23 |
|  | Touchscreen questionnaire | 6150 | 1 |
|  | Verbal interview | 20002 | 1075 |
| Date of myocardial infarction first reported | Health-related outcomes | 2409 | 131298, 131300,131302 |
| Stroke | Hospital inpatients (ICD-10) | 41270 | I60-I64 |
|  | Touchscreen questionnaire | 6150 | 3 |
|  | Verbal interview | 20002 | 1081, 1083, 1086 |
| Date of stroke first reported | Health-related outcomes | 2409 | 131360, 131362, 131364, 131366, 131368 |
| Ischemic stroke | Hospital inpatients (ICD-10) | 41270 | I63 |
|  | Verbal interview | 20002 | 1583 |
| Date of ischemic stroke first reported | Health-related outcomes | 2409 | 131366 |
| Obstructive sleep apnea/hypoxia syndrome | Hospital inpatients (ICD-10) | 41270 | G47.3 |
|  | Verbal interview | 20002 | 1958 |
| Date of obstructive sleep apnea syndrome first reported | Health-related outcomes |  | 131060 |
| Date of death | Death register | 40000 |  |
| Death | Death register (ICD-10) | 40001 | A00-U49 |
| Cardiovascular disease deaths | Death register (ICD-10) | 40001 | I05-I89 |
| Cancer deaths | Death register (ICD-10) | 40001 | C00-D48 |
| Other deaths | Death register (ICD-10) | 40001 | A00-B99, D55-H75, J09-U49 |

#### Supplementary Table S2

**Metabolic biomarkers reaching the set significance threshold of P value (< 0.0009).**

| **Metabolic Biomarkers** | **β** | **95%CI** | | **P-value*** |
| --- | --- | --- | --- | --- |
| Ratio of saturated fatty acids to total fatty acids | -0.411 | -0.576 | -0.246 | 1.02E-06 |
| Phospholipids in Medium HDL | -0.284 | -0.410 | -0.157 | 1.09E-05 |
| Ratio of linoleic acid to total fatty acids | 0.327 | 0.176 | 0.477 | 2.03E-05 |
| Total Lipids in Medium HDL | -0.271 | -0.399 | -0.143 | 3.17E-05 |
| Phospholipids in HDL | -0.279 | -0.411 | -0.148 | 3.19E-05 |
| Phospholipids to total lipids ratio in small HDL | -0.257 | -0.380 | -0.134 | 4.16E-05 |
| Concentration of Medium HDL Particles | -0.266 | -0.394 | -0.138 | 4.78E-05 |
| Free Cholesterol in Medium HDL | -0.264 | -0.394 | -0.134 | 6.90E-05 |
| Apolipoprotein A1 | -0.253 | -0.383 | -0.124 | 1.29E-04 |
| Total Lipids in HDL | -0.260 | -0.393 | -0.127 | 1.34E-04 |
| Cholesterol in Medium HDL | -0.251 | -0.381 | -0.121 | 1.56E-04 |
| Cholesteryl Esters in Medium HDL | -0.247 | -0.377 | -0.117 | 1.90E-04 |
| Free Cholesterol in HDL | -0.240 | -0.375 | -0.105 | 4.87E-04 |
| Ratio of omega-6 fatty acids to total fatty acids | 0.259 | 0.110 | 0.408 | 6.85E-04 |
| Ratio of apolipoprotein B to apolipoprotein A1 | 0.222 | 0.093 | 0.351 | 7.43E-04 |
| Concentration of HDL Particles | -0.213 | -0.338 | -0.088 | 8.47E-04 |

*Fully adjusted for sex, age, race, Townsend Deprivation Index, household income, body mass index, smoking, alcohol, lipid-lowering medication, spherical equivalent and intraocular pressure.

GCIPL = ganglion cell-inner plexus layer; HDL = high-density lipoprotein; CI = confidence interval.

#### Supplementary Table S3

**No. of incident outcomes in total, training set and validation set.**

| **Outcome** | **Total** | **Training set** | **Validation set** |
| --- | --- | --- | --- |
| **Systemic diseases** |  |  |  |
| Type 2 diabetes | 5,714 (7.28%) | 2,918 (7.45%) | 2,796 (7.11%) |
| OSAHS | 1,366 (1.62%) | 688 (1.64%) | 678 (1.61%) |
| Dementia | 1,219 (1.44%) | 608 (1.44%) | 611 (1.44%) |
| Ischemic stroke | 1,578 (1.88%) | 796 (1.89%) | 782 (1.86%) |
| Heart failure | 2,537 (3.05%) | 1,255 (3.02%) | 1,282 (3.09%) |
| Myocardial infarction | 2,866 (3.47%) | 1,420 (3.44%) | 1,446 (3.50%) |
| **Mortality** |  |  |  |
| All-cause mortality | 6,254 (7.84%) | 3,166 (7.95%) | 3,088 (7.74%) |
| CVD mortality | 1,544 (1.94%) | 800 (2.01%) | 744 (1.86%) |
| Cancer mortality | 3,151 (3.95%) | 1,572 (3.95%) | 1,579 (3.96%) |
| Other mortality | 1,559 (1.95%) | 794 (1.99%) | 765 (1.92%) |

T2DM = type 2 diabetes mellites; OSAHS = obstructive sleep apnea/hypoxia syndrome; CVD = cardiovascular disease.

#### Supplementary Table S4

**Baseline characteristics of subjects stratified by incident type 2 diabetes.**

| **Characteristic** | **Training set** | | **Validation set** | |
| --- | --- | --- | --- | --- |
|  | **No** | **Yes** | **No** | **Yes** |
| No. of subjects | 39194 | 2918 | 39316 | 2796 |
| Age at recruitment (%) | | | | |
| ≤49 | 9193 (23.5) | 364 (12.5) | 9092 (23.1) | 317 (11.3) |
| 50-54 | 5911 (15.1) | 333 (11.4) | 5832 (14.8) | 349 (12.5) |
| 55-59 | 7031 (17.9) | 484 (16.6) | 7227 (18.4) | 502 (18.0) |
| 60-64 | 9676 (24.7) | 869 (29.8) | 9602 (24.4) | 806 (28.8) |
| ≥65 | 7383 (18.8) | 868 (29.7) | 7563 (19.2) | 822 (29.4) |
| Gender (%) | | | | |
| Female | 21996 (56.1) | 1198 (41.1) | 21979 (55.9) | 1152 (41.2) |
| Male | 17198 (43.9) | 1720 (58.9) | 17337 (44.1) | 1644 (58.8) |
| Race (%) | | | | |
| White | 37381 (95.4) | 2596 (89.0) | 37514 (95.4) | 2493 (89.2) |
| Others | 1646 (4.2) | 301 (10.3) | 1646 (4.2) | 282 (10.1) |
| Missing | 167 (0.4) | 21 (0.7) | 156 (0.4) | 21 (0.8) |
| Townsend Deprivation Index (%) | | | | |
| Quantile 1 | 10301 (26.3) | 559 (19.2) | 10099 (25.7) | 585 (20.9) |
| Quantile 2 | 9864 (25.2) | 614 (21.0) | 10103 (25.7) | 585 (20.9) |
| Quantile 3 | 9723 (24.8) | 709 (24.3) | 9654 (24.6) | 650 (23.2) |
| Quantile 4 | 9255 (23.6) | 1033 (35.4) | 9403 (23.9) | 974 (34.8) |
| Missing | 51 (0.1) | 3 (0.1) | 57 (0.1) | 2 (0.1) |
| Average total household income before tax (£) | | | | |
| < 18k | 7647 (19.5) | 891 (30.5) | 7600 (19.3) | 873 (31.2) |
| 18k~30k | 8650 (22.1) | 702 (24.1) | 8747 (22.2) | 662 (23.7) |
| 31k~51k | 8735 (22.3) | 475 (16.3) | 8800 (22.4) | 464 (16.6) |
| 52k~100k | 6678 (17.0) | 258 (8.8) | 6755 (17.2) | 271 (9.7) |
| > 100k | 1669 (4.3) | 46 (1.6) | 1681 (4.3) | 41 (1.5) |
| Missing | 5815 (14.8) | 546 (18.7) | 5733 (14.6) | 485 (17.3) |
| Body mass index (%) | | | | |
| Normal | 13418 (34.2) | 257 (8.8) | 13293 (33.8) | 261 (9.3) |
| Overweight | 16897 (43.1) | 1049 (35.9) | 17008 (43.3) | 982 (35.1) |
| Obesity | 8737 (22.3) | 1590 (54.5) | 8871 (22.6) | 1531 (54.8) |
| Missing | 142 (0.4) | 22 (0.8) | 144 (0.4) | 22 (0.8) |
| Smoking (%) | | | | |
| Never | 13464 (34.4) | 1196 (41.0) | 13718 (34.9) | 1104 (39.5) |
| Ever/Current | 4201 (10.7) | 426 (14.6) | 4106 (10.4) | 397 (14.2) |
| Missing | 21529 (54.9) | 1296 (44.4) | 21492 (54.7) | 1295 (46.3) |
| Drinking (%) | | | | |
| Never | 1353 (3.5) | 201 (6.9) | 1360 (3.5) | 171 (6.1) |
| Ever/Current | 36137 (92.2) | 2465 (84.5) | 36296 (92.3) | 2391 (85.5) |
| Missing | 1704 (4.3) | 252 (8.6) | 1660 (4.2) | 234 (8.4) |
| Lipid-lowering medication (%) | | | | |
| No | 33488 (85.4) | 1516 (52.0) | 33555 (85.3) | 1438 (51.4) |
| Yes | 5706 (14.6) | 1402 (48.0) | 5761 (14.7) | 1358 (48.6) |

#### Supplementary Table S5

**Baseline characteristics of subjects stratified by incident OSAHS.**

| **Characteristic** | **Training set** | | **Validation set** | |
| --- | --- | --- | --- | --- |
|  | **No** | **Yes** | **No** | **Yes** |
| No. of subjects | 42081 | 688 | 42101 | 678 |
| Age at recruitment (%) | | | | |
| ≤49 | 9426 (22.4) | 164 (23.8) | 9367 (22.2) | 158 (23.3) |
| 50-54 | 6200 (14.7) | 91 (13.2) | 6179 (14.7) | 110 (16.2) |
| 55-59 | 7508 (17.8) | 138 (20.1) | 7660 (18.2) | 133 (19.6) |
| 60-64 | 10512 (25.0) | 180 (26.2) | 10466 (24.9) | 164 (24.2) |
| ≥65 | 8435 (20.0) | 115 (16.7) | 8429 (20.0) | 113 (16.7) |
| Gender (%) | | | | |
| Female | 23323 (55.4) | 221 (32.1) | 23204 (55.1) | 203 (29.9) |
| Male | 18758 (44.6) | 467 (67.9) | 18897 (44.9) | 475 (70.1) |
| Race (%) | | | | |
| White | 39899 (94.8) | 643 (93.5) | 39954 (94.9) | 621 (91.6) |
| Others | 1998 (4.7) | 40 (5.8) | 1957 (4.6) | 53 (7.8) |
| Missing | 184 (0.4) | 5 (0.7) | 190 (0.5) | 4 (0.6) |
| Townsend Deprivation Index (%) | | | | |
| Quantile 1 | 10694 (25.4) | 136 (19.8) | 10791 (25.6) | 137 (20.2) |
| Quantile 2 | 10498 (24.9) | 148 (21.5) | 10631 (25.3) | 134 (19.8) |
| Quantile 3 | 10462 (24.9) | 176 (25.6) | 10275 (24.4) | 159 (23.5) |
| Quantile 4 | 10369 (24.6) | 228 (33.1) | 10349 (24.6) | 246 (36.3) |
| Missing | 58 (0.1) | 0 (0.0) | 55 (0.1) | 2 (0.3) |
| Average total household income before tax (£) | | | | |
| < 18k | 8501 (20.2) | 180 (26.2) | 8621 (20.5) | 184 (27.1) |
| 18k~30k | 9386 (22.3) | 160 (23.3) | 9335 (22.2) | 161 (23.7) |
| 31k~51k | 9223 (21.9) | 156 (22.7) | 9143 (21.7) | 131 (19.3) |
| 52k~100k | 6936 (16.5) | 80 (11.6) | 6941 (16.5) | 91 (13.4) |
| > 100k | 1679 (4.0) | 12 (1.7) | 1741 (4.1) | 14 (2.1) |
| Missing | 6356 (15.1) | 100 (14.5) | 6320 (15.0) | 97 (14.3) |
| BMI (%) | | | | |
| Normal | 13683 (32.5) | 49 (7.1) | 13638 (32.4) | 68 (10.0) |
| Overweight | 18002 (42.8) | 205 (29.8) | 17976 (42.7) | 182 (26.8) |
| Obesity | 10240 (24.3) | 431 (62.6) | 10308 (24.5) | 421 (62.1) |
| Missing | 156 (0.4) | 3 (0.4) | 179 (0.4) | 7 (1.0) |
| Smoking (%) | | | | |
| Never | 14691 (34.9) | 282 (41.0) | 14804 (35.2) | 275 (40.6) |
| Ever/Current | 4535 (10.8) | 98 (14.2) | 4560 (10.8) | 81 (11.9) |
| Missing | 22855 (54.3) | 308 (44.8) | 22737 (54.0) | 322 (47.5) |
| Drinking (%) | | | | |
| Never | 1552 (3.7) | 51 (7.4) | 1569 (3.7) | 49 (7.2) |
| Ever/Current | 38583 (91.7) | 594 (86.3) | 38548 (91.6) | 599 (88.3) |
| Missing | 1946 (4.6) | 43 (6.2) | 1984 (4.7) | 30 (4.4) |
| Lipid-lowering medication (%) | | | | |
| No | 34642 (82.3) | 446 (64.8) | 34622 (82.2) | 453 (66.8) |
| Yes | 7439 (17.7) | 242 (35.2) | 7479 (17.8) | 225 (33.2) |

#### Supplementary Table S6

**Baseline characteristics of subjects stratified characteristics by incident dementia.**

| **Characteristic** | **Training set** | | **Validation set** | |
| --- | --- | --- | --- | --- |
|  | **No** | **Yes** | **No** | **Yes** |
| No. of subjects | 42365 | 608 | 42379 | 611 |
| Age at recruitment (%) | | | | |
| ≤49 | 9604 (22.7) | 13 (2.1) | 9551 (22.5) | 10 (1.6) |
| 50-54 | 6307 (14.9) | 21 (3.5) | 6297 (14.9) | 18 (2.9) |
| 55-59 | 7642 (18.0) | 45 (7.4) | 7795 (18.4) | 45 (7.4) |
| 60-64 | 10560 (24.9) | 176 (28.9) | 10534 (24.9) | 166 (27.2) |
| ≥65 | 8252 (19.5) | 353 (58.1) | 8202 (19.4) | 372 (60.9) |
| Gender (%) | | | | |
| Female | 23275 (54.9) | 293 (48.2) | 23187 (54.7) | 265 (43.4) |
| Male | 19090 (45.1) | 315 (51.8) | 19192 (45.3) | 346 (56.6) |
| Race (%) | | | | |
| White | 40153 (94.8) | 586 (96.4) | 40188 (94.8) | 590 (96.6) |
| Others | 2028 (4.8) | 16 (2.6) | 2001 (4.7) | 17 (2.8) |
| Missing | 184 (0.4) | 6 (1.0) | 190 (0.4) | 4 (0.7) |
| Townsend Deprivation Index (%) | | | | |
| Quantile 1 | 10739 (25.3) | 126 (20.7) | 10836 (25.6) | 136 (22.3) |
| Quantile 2 | 10556 (24.9) | 151 (24.8) | 10659 (25.2) | 161 (26.4) |
| Quantile 3 | 10537 (24.9) | 149 (24.5) | 10341 (24.4) | 145 (23.7) |
| Quantile 4 | 10475 (24.7) | 182 (29.9) | 10486 (24.7) | 169 (27.7) |
| Missing | 58 (0.1) | 0 (0.0) | 57 (0.1) | 0 (0.0) |
| Average total household income before tax (£) | | | | |
| < 18k | 8505 (20.1) | 225 (37.0) | 8652 (20.4) | 205 (33.6) |
| 18k~30k | 9447 (22.3) | 139 (22.9) | 9398 (22.2) | 152 (24.9) |
| 31k~51k | 9370 (22.1) | 50 (8.2) | 9256 (21.8) | 64 (10.5) |
| 52k~100k | 7027 (16.6) | 30 (4.9) | 7019 (16.6) | 38 (6.2) |
| > 100k | 1689 (4.0) | 9 (1.5) | 1756 (4.1) | 9 (1.5) |
| Missing | 6327 (14.9) | 155 (25.5) | 6298 (14.9) | 143 (23.4) |
| BMI (%) | | | | |
| Normal | 13563 (32.0) | 177 (29.1) | 13521 (31.9) | 195 (31.9) |
| Overweight | 18011 (42.5) | 266 (43.8) | 17974 (42.4) | 247 (40.4) |
| Obesity | 10641 (25.1) | 154 (25.3) | 10706 (25.3) | 161 (26.4) |
| Missing | 150 (0.4) | 11 (1.8) | 178 (0.4) | 8 (1.3) |
| Smoking (%) | | | | |
| Never | 14830 (35.0) | 246 (40.5) | 14899 (35.2) | 263 (43.0) |
| Ever/Current | 4579 (10.8) | 72 (11.8) | 4608 (10.9) | 67 (11.0) |
| Missing | 22956 (54.2) | 290 (47.7) | 22872 (54.0) | 281 (46.0) |
| Drinking (%) | | | | |
| Never | 1563 (3.7) | 48 (7.9) | 1586 (3.7) | 46 (7.5) |
| Ever/Current | 38849 (91.7) | 516 (84.9) | 38811 (91.6) | 520 (85.1) |
| Missing | 1953 (4.6) | 44 (7.2) | 1982 (4.7) | 45 (7.4) |
| Lipid-lowering medication (%) | | | | |
| No | 34811 (82.2) | 393 (64.6) | 34806 (82.1) | 385 (63.0) |
| Yes | 7554 (17.8) | 215 (35.4) | 7573 (17.9) | 226 (37.0) |

#### Supplementary Table S7

**Baseline characteristics of subjects stratified by incident ischemic stroke.**

| **Characteristic** | **Training set** | | **Validation set** | |
| --- | --- | --- | --- | --- |
|  | **No** | **Yes** | **No** | **Yes** |
| No. of subjects | 42043 | 796 | 42067 | 782 |
| Age at recruitment (%) | | | | |
| ≤49 | 9559 (22.7) | 49 (6.2) | 9493 (22.6) | 53 (6.8) |
| 50-54 | 6264 (14.9) | 57 (7.2) | 6248 (14.9) | 55 (7.0) |
| 55-59 | 7580 (18.0) | 95 (11.9) | 7707 (18.3) | 107 (13.7) |
| 60-64 | 10461 (24.9) | 234 (29.4) | 10432 (24.8) | 237 (30.3) |
| ≥65 | 8179 (19.5) | 361 (45.4) | 8187 (19.5) | 330 (42.2) |
| Gender (%) | | | | |
| Female | 23232 (55.3) | 312 (39.2) | 23110 (54.9) | 297 (38.0) |
| Male | 18811 (44.7) | 484 (60.8) | 18957 (45.1) | 485 (62.0) |
| Race (%) | | | | |
| White | 39842 (94.8) | 767 (96.4) | 39903 (94.9) | 742 (94.9) |
| Others | 2013 (4.8) | 27 (3.4) | 1979 (4.7) | 33 (4.2) |
| Missing | 188 (0.4) | 2 (0.3) | 185 (0.4) | 7 (0.9) |
| Townsend Deprivation Index (%) | | | | |
| Quantile 1 | 10641 (25.3) | 190 (23.9) | 10779 (25.6) | 170 (21.7) |
| Quantile 2 | 10496 (25.0) | 193 (24.2) | 10581 (25.2) | 191 (24.4) |
| Quantile 3 | 10466 (24.9) | 183 (23.0) | 10272 (24.4) | 194 (24.8) |
| Quantile 4 | 10382 (24.7) | 230 (28.9) | 10378 (24.7) | 227 (29.0) |
| Missing | 58 (0.1) | 0 (0.0) | 57 (0.1) | 0 (0.0) |
| Average total household income before tax (£) | | | | |
| < 18k | 8437 (20.1) | 239 (30.0) | 8581 (20.4) | 232 (29.7) |
| 18k~30k | 9360 (22.3) | 201 (25.3) | 9329 (22.2) | 183 (23.4) |
| 31k~51k | 9273 (22.1) | 126 (15.8) | 9159 (21.8) | 140 (17.9) |
| 52k~100k | 6986 (16.6) | 60 (7.5) | 6978 (16.6) | 67 (8.6) |
| > 100k | 1684 (4.0) | 15 (1.9) | 1748 (4.2) | 18 (2.3) |
| Missing | 6303 (15.0) | 155 (19.5) | 6272 (14.9) | 142 (18.2) |
| BMI (%) | | | | |
| Normal | 13507 (32.1) | 210 (26.4) | 13483 (32.1) | 207 (26.5) |
| Overweight | 17880 (42.5) | 334 (42.0) | 17823 (42.4) | 342 (43.7) |
| Obesity | 10507 (25.0) | 246 (30.9) | 10582 (25.2) | 228 (29.2) |
| Missing | 149 (0.4) | 6 (0.8) | 179 (0.4) | 5 (0.6) |
| Smoking (%) | | | | |
| Never | 14702 (35.0) | 303 (38.1) | 14759 (35.1) | 336 (43.0) |
| Ever/Current | 4513 (10.7) | 120 (15.1) | 4518 (10.7) | 133 (17.0) |
| Missing | 22828 (54.3) | 373 (46.9) | 22790 (54.2) | 313 (40.0) |
| Drinking (%) | | | | |
| Never | 1562 (3.7) | 35 (4.4) | 1585 (3.8) | 38 (4.9) |
| Ever/Current | 38528 (91.6) | 723 (90.8) | 38516 (91.6) | 697 (89.1) |
| Missing | 1953 (4.6) | 38 (4.8) | 1966 (4.7) | 47 (6.0) |
| Lipid-lowering medication (%) | | | | |
| No | 34675 (82.5) | 521 (65.5) | 34646 (82.4) | 526 (67.3) |
| Yes | 7368 (17.5) | 275 (34.5) | 7421 (17.6) | 256 (32.7) |

#### Supplementary Table S8

**Baseline characteristics of subjects stratified by incident heart failure.**

| **Characteristic** | **Training set** | | **Validation set** | |
| --- | --- | --- | --- | --- |
|  | **No** | **Yes** | **No** | **Yes** |
| No. of subjects | 41584 | 1255 | 41553 | 1282 |
| Age at recruitment (%) | | | | |
| ≤49 | 9551 (23.0) | 58 (4.6) | 9485 (22.8) | 64 (5.0) |
| 50-54 | 6236 (15.0) | 82 (6.5) | 6206 (14.9) | 94 (7.3) |
| 55-59 | 7486 (18.0) | 181 (14.4) | 7655 (18.4) | 165 (12.9) |
| 60-64 | 10320 (24.8) | 370 (29.5) | 10249 (24.7) | 410 (32.0) |
| ≥65 | 7991 (19.2) | 564 (44.9) | 7958 (19.2) | 549 (42.8) |
| Gender (%) | | | | |
| Female | 23085 (55.5) | 467 (37.2) | 22971 (55.3) | 454 (35.4) |
| Male | 18499 (44.5) | 788 (62.8) | 18582 (44.7) | 828 (64.6) |
| Race (%) |  |  |  |  |
| White | 39421 (94.8) | 1194 (95.1) | 39406 (94.8) | 1224 (95.5) |
| Others | 1976 (4.8) | 57 (4.5) | 1960 (4.7) | 52 (4.1) |
| Missing | 187 (0.4) | 4 (0.3) | 187 (0.5) | 6 (0.5) |
| Townsend Deprivation Index (%) | | | | |
| Quantile 1 | 10568 (25.4) | 270 (21.5) | 10677 (25.7) | 263 (20.5) |
| Quantile 2 | 10396 (25.0) | 281 (22.4) | 10492 (25.2) | 297 (23.2) |
| Quantile 3 | 10354 (24.9) | 301 (24.0) | 10139 (24.4) | 305 (23.8) |
| Quantile 4 | 10209 (24.6) | 402 (32.0) | 10188 (24.5) | 417 (32.5) |
| Missing | 57 (0.1) | 1 (0.1) | 57 (0.1) | 0 (0.0) |
| Average total household income before tax (£) | | | | |
| < 18k | 8271 (19.9) | 404 (32.2) | 8355 (20.1) | 453 (35.3) |
| 18k~30k | 9263 (22.3) | 296 (23.6) | 9220 (22.2) | 288 (22.5) |
| 31k~51k | 9207 (22.1) | 191 (15.2) | 9125 (22.0) | 172 (13.4) |
| 52k~100k | 6962 (16.7) | 86 (6.9) | 6943 (16.7) | 103 (8.0) |
| > 100k | 1680 (4.0) | 17 (1.4) | 1739 (4.2) | 24 (1.9) |
| Missing | 6201 (14.9) | 261 (20.8) | 6171 (14.9) | 242 (18.9) |
| BMI (%) | | | | |
| Normal | 13461 (32.4) | 264 (21.0) | 13448 (32.4) | 237 (18.5) |
| Overweight | 17746 (42.7) | 476 (37.9) | 17689 (42.6) | 486 (37.9) |
| Obesity | 10227 (24.6) | 508 (40.5) | 10246 (24.7) | 545 (42.5) |
| Missing | 150 (0.4) | 7 (0.6) | 170 (0.4) | 14 (1.1) |
| Smoking (%) | | | | |
| Never | 14446 (34.7) | 555 (44.2) | 14489 (34.9) | 577 (45.0) |
| Ever/Current | 4419 (10.6) | 218 (17.4) | 4463 (10.7) | 195 (15.2) |
| Missing | 22719 (54.6) | 482 (38.4) | 22601 (54.4) | 510 (39.8) |
| Drinking (%) | | | | |
| Never | 1516 (3.6) | 85 (6.8) | 1531 (3.7) | 86 (6.7) |
| Ever/Current | 38165 (91.8) | 1086 (86.5) | 38065 (91.6) | 1131 (88.2) |
| Missing | 1903 (4.6) | 84 (6.7) | 1957 (4.7) | 65 (5.1) |
| Lipid-lowering medication (%) | | | | |
| No | 34479 (82.9) | 702 (55.9) | 34439 (82.9) | 718 (56.0) |
| Yes | 7105 (17.1) | 553 (44.1) | 7114 (17.1) | 564 (44.0) |

#### Supplementary Table S9

**Baseline characteristics of subjects stratified by incident myocardial infarction.**

| **Characteristic** | **Training set** | | **Validation set** | |
| --- | --- | --- | --- | --- |
|  | **No** | **Yes** | **No** | **Yes** |
| No. of subjects | 41318 | 1420 | 41331 | 1446 |
| Age at recruitment (%) | | | | |
| ≤49 | 9516 (23.0) | 100 (7.0) | 9444 (22.8) | 106 (7.3) |
| 50-54 | 6194 (15.0) | 123 (8.7) | 6160 (14.9) | 144 (10.0) |
| 55-59 | 7451 (18.0) | 200 (14.1) | 7595 (18.4) | 209 (14.5) |
| 60-64 | 10214 (24.7) | 433 (30.5) | 10177 (24.6) | 459 (31.7) |
| ≥65 | 7943 (19.2) | 564 (39.7) | 7955 (19.2) | 528 (36.5) |
| Gender (%) | | | | |
| Female | 23141 (56.0) | 414 (29.2) | 23019 (55.7) | 394 (27.2) |
| Male | 18177 (44.0) | 1006 (70.8) | 18312 (44.3) | 1052 (72.8) |
| Race (%) | | | | |
| White | 39175 (94.8) | 1345 (94.7) | 39219 (94.9) | 1363 (94.3) |
| Others | 1958 (4.7) | 69 (4.9) | 1923 (4.7) | 80 (5.5) |
| Missing | 185 (0.4) | 6 (0.4) | 189 (0.5) | 3 (0.2) |
| Townsend Deprivation Index (%) | | | | |
| Quantile 1 | 10495 (25.4) | 327 (23.0) | 10610 (25.7) | 316 (21.9) |
| Quantile 2 | 10305 (24.9) | 351 (24.7) | 10432 (25.2) | 345 (23.9) |
| Quantile 3 | 10315 (25.0) | 318 (22.4) | 10097 (24.4) | 340 (23.5) |
| Quantile 4 | 10145 (24.6) | 424 (29.9) | 10137 (24.5) | 443 (30.6) |
| Missing | 58 (0.1) | 0 (0.0) | 55 (0.1) | 2 (0.1) |
| Average total household income before tax (£) | | | | |
| < 18k | 8198 (19.8) | 449 (31.6) | 8326 (20.1) | 453 (31.3) |
| 18k~30k | 9180 (22.2) | 346 (24.4) | 9169 (22.2) | 332 (23.0) |
| 31k~51k | 9157 (22.2) | 229 (16.1) | 9054 (21.9) | 244 (16.9) |
| 52k~100k | 6936 (16.8) | 113 (8.0) | 6918 (16.7) | 125 (8.6) |
| > 100k | 1671 (4.0) | 25 (1.8) | 1733 (4.2) | 30 (2.1) |
| Missing | 6176 (14.9) | 258 (18.2) | 6131 (14.8) | 262 (18.1) |
| BMI (%) | | | | |
| Normal | 13435 (32.5) | 268 (18.9) | 13389 (32.4) | 297 (20.5) |
| Overweight | 17542 (42.5) | 640 (45.1) | 17537 (42.4) | 602 (41.6) |
| Obesity | 10194 (24.7) | 505 (35.6) | 10228 (24.7) | 539 (37.3) |
| Missing | 147 (0.4) | 7 (0.5) | 177 (0.4) | 8 (0.6) |
| Smoking (%) | | | | |
| Never | 14292 (34.6) | 660 (46.5) | 14374 (34.8) | 665 (46.0) |
| Ever/Current | 4387 (10.6) | 229 (16.1) | 4407 (10.7) | 230 (15.9) |
| Missing | 22639 (54.8) | 531 (37.4) | 22550 (54.6) | 551 (38.1) |
| Drinking (%) | | | | |
| Never | 1522 (3.7) | 74 (5.2) | 1532 (3.7) | 90 (6.2) |
| Ever/Current | 37912 (91.8) | 1255 (88.4) | 37863 (91.6) | 1279 (88.5) |
| Missing | 1884 (4.6) | 91 (6.4) | 1936 (4.7) | 77 (5.3) |
| Lipid-lowering medication (%) | | | | |
| No | 34581 (83.7) | 620 (43.7) | 34527 (83.5) | 660 (45.6) |
| Yes | 6737 (16.3) | 800 (56.3) | 6804 (16.5) | 786 (54.4) |

#### Supplementary Table S10

**Baseline characteristics of subjects stratified by all-cause mortality.**

| **Characteristic** | **Training set** | | **Validation set** | |
| --- | --- | --- | --- | --- |
|  | **No** | **Yes** | **No** | **Yes** |
| No. of subjects | 39841 | 3166 | 39919 | 3088 |
| Age at recruitment (%) | | | | |
| ≤49 | 9409 (23.6) | 217 (6.9) | 9382 (23.5) | 182 (5.9) |
| 50-54 | 6112 (15.3) | 222 (7.0) | 6071 (15.2) | 247 (8.0) |
| 55-59 | 7231 (18.1) | 463 (14.6) | 7414 (18.6) | 429 (13.9) |
| 60-64 | 9806 (24.6) | 936 (29.6) | 9748 (24.4) | 956 (31.0) |
| ≥65 | 7283 (18.3) | 1328 (41.9) | 7304 (18.3) | 1274 (41.3) |
| Gender (%) | | | | |
| Female | 22333 (56.1) | 1263 (39.9) | 22235 (55.7) | 1229 (39.8) |
| Male | 17508 (43.9) | 1903 (60.1) | 17684 (44.3) | 1859 (60.2) |
| Race (%) | | | | |
| White | 37725 (94.7) | 3046 (96.2) | 37810 (94.7) | 2985 (96.7) |
| Others | 1940 (4.9) | 105 (3.3) | 1935 (4.8) | 83 (2.7) |
| Missing | 176 (0.4) | 15 (0.5) | 174 (0.4) | 20 (0.6) |
| Townsend Deprivation Index (%) | | | | |
| Quantile 1 | 10209 (25.6) | 662 (20.9) | 10273 (25.7) | 705 (22.8) |
| Quantile 2 | 9945 (25.0) | 768 (24.3) | 10127 (25.4) | 693 (22.4) |
| Quantile 3 | 9948 (25.0) | 741 (23.4) | 9754 (24.4) | 736 (23.8) |
| Quantile 4 | 9682 (24.3) | 994 (31.4) | 9709 (24.3) | 953 (30.9) |
| Missing | 57 (0.1) | 1 (0.0) | 56 (0.1) | 1 (0.0) |
| Average total household income before tax (£) | | | | |
| < 18k | 7665 (19.2) | 1077 (34.0) | 7849 (19.7) | 1015 (32.9) |
| 18k~30k | 8832 (22.2) | 761 (24.0) | 8862 (22.2) | 688 (22.3) |
| 31k~51k | 9012 (22.6) | 413 (13.0) | 8839 (22.1) | 484 (15.7) |
| 52k~100k | 6817 (17.1) | 241 (7.6) | 6791 (17.0) | 269 (8.7) |
| > 100k | 1644 (4.1) | 56 (1.8) | 1717 (4.3) | 49 (1.6) |
| Missing | 5871 (14.7) | 618 (19.5) | 5861 (14.7) | 583 (18.9) |
| BMI (%) | | | | |
| Normal | 12898 (32.4) | 853 (26.9) | 12842 (32.2) | 877 (28.4) |
| Overweight | 16962 (42.6) | 1328 (41.9) | 16950 (42.5) | 1281 (41.5) |
| Obesity | 9853 (24.7) | 952 (30.1) | 9967 (25.0) | 903 (29.2) |
| Missing | 128 (0.3) | 33 (1.0) | 160 (0.4) | 27 (0.9) |
| Smoking (%) | | | | |
| Never | 13765 (34.5) | 1320 (41.7) | 13878 (34.8) | 1289 (41.7) |
| Ever/Current | 4087 (10.3) | 574 (18.1) | 4028 (10.1) | 650 (21.0) |
| Missing | 21989 (55.2) | 1272 (40.2) | 22013 (55.1) | 1149 (37.2) |
| Drinking (%) | | | | |
| Never | 1387 (3.5) | 228 (7.2) | 1409 (3.5) | 225 (7.3) |
| Ever/Current | 36635 (92.0) | 2758 (87.1) | 36631 (91.8) | 2715 (87.9) |
| Missing | 1819 (4.6) | 180 (5.7) | 1879 (4.7) | 148 (4.8) |
| Lipid-lowering medication (%) | | | | |
| No | 33085 (83.0) | 2146 (67.8) | 33074 (82.9) | 2131 (69.0) |
| Yes | 6756 (17.0) | 1020 (32.2) | 6845 (17.1) | 957 (31.0) |

#### Supplementary Table S11

**Associations of GCIPLT metabolomic signature and incident T2DM.**

| **Metabolic Biomarkers** | **Training set** | | | | | **Validation set** | | | | |
| --- | --- | --- | --- | --- | --- | --- | --- | --- | --- | --- |
|  | **HR** | **95% CI** | | **SE** | **P value*** | **HR** | **95% CI** | | **SE** | **P value*** |
| Ratio of saturated fatty acids to total fatty acids | 1.047 | 1.039 | 1.055 | 0.004 | 2.00E-32 | 1.037 | 1.025 | 1.049 | 0.006 | 9.10E-10 |
| Phospholipids in Medium HDL | 0.909 | 0.885 | 0.934 | 0.014 | 3.09E-12 | 0.905 | 0.871 | 0.941 | 0.020 | 5.81E-07 |
| Ratio of linoleic acid to total fatty acids | 0.693 | 0.675 | 0.712 | 0.014 | 0.00E+00 | 0.700 | 0.673 | 0.728 | 0.020 | 0.00E+00 |
| Total Lipids in Medium HDL | 0.862 | 0.839 | 0.886 | 0.014 | 1.31E-26 | 0.861 | 0.827 | 0.896 | 0.020 | 1.22E-13 |
| Phospholipids in HDL | 0.833 | 0.809 | 0.857 | 0.015 | 4.50E-35 | 0.833 | 0.799 | 0.869 | 0.021 | 2.27E-17 |
| Phospholipids to total lipids ratio in small HDL | 1.107 | 1.079 | 1.135 | 0.013 | 4.47E-15 | 1.097 | 1.057 | 1.138 | 0.019 | 9.32E-07 |
| Concentration of Medium HDL Particles | 0.820 | 0.796 | 0.844 | 0.015 | 0.00E+00 | 0.821 | 0.787 | 0.857 | 0.022 | 8.44E-20 |
| Free Cholesterol in Medium HDL | 0.804 | 0.780 | 0.829 | 0.015 | 0.00E+00 | 0.806 | 0.772 | 0.842 | 0.022 | 4.33E-22 |
| Apolipoprotein A1 | 0.809 | 0.787 | 0.833 | 0.015 | 0.00E+00 | 0.811 | 0.778 | 0.845 | 0.021 | 3.65E-23 |
| Total Lipids in HDL | 0.783 | 0.761 | 0.807 | 0.015 | 0.00E+00 | 0.786 | 0.753 | 0.821 | 0.022 | 4.98E-28 |
| Cholesterol in Medium HDL | 0.773 | 0.751 | 0.796 | 0.015 | 0.00E+00 | 0.777 | 0.745 | 0.810 | 0.022 | 1.01E-31 |
| Cholesteryl Esters in Medium HDL | 0.766 | 0.745 | 0.789 | 0.015 | 0.00E+00 | 0.771 | 0.739 | 0.804 | 0.022 | 1.26E-33 |
| Free Cholesterol in HDL | 0.737 | 0.713 | 0.762 | 0.017 | 0.00E+00 | 0.742 | 0.707 | 0.779 | 0.025 | 1.27E-33 |
| Ratio of omega-6 fatty acids to total fatty acids | 0.712 | 0.695 | 0.729 | 0.012 | 0.00E+00 | 0.719 | 0.694 | 0.745 | 0.018 | 0.00E+00 |
| Ratio of apolipoprotein B to apolipoprotein A1 | 1.000 | 0.973 | 1.028 | 0.014 | 9.99E-01 | 0.993 | 0.954 | 1.033 | 0.020 | 7.10E-01 |
| Concentration of HDL Particles | 0.807 | 0.785 | 0.830 | 0.014 | 0.00E+00 | 0.809 | 0.776 | 0.842 | 0.021 | 1.39E-24 |

*Fully adjusted for sex, age, race, Townsend Deprivation Index, household income, BMI, smoking, alcohol, and lipid-lowering medication.

GCIPLT = ganglion cell-inner plexus layer thickness; T2DM = type 2 diabetes mellitus; HDL = high-density lipoprotein; HR = hazard ratio; CI = confidence interval; SE = standard error.

#### Supplementary Table S12

**Associations of GCIPLT metabolomic signature and incident OSAHS.**

| **Metabolic Biomarkers** | **Training set** | | | | | **Validation set** | | | | |
| --- | --- | --- | --- | --- | --- | --- | --- | --- | --- | --- |
|  | **HR** | **95% CI** | | **SE** | **P-value*** | **HR** | **95% CI** | | **SE** | **P-value*** |
| Ratio of saturated fatty acids to total fatty acids | 1.020 | 0.984 | 1.057 | 0.018 | 2.71E-01 | 1.017 | 0.965 | 1.071 | 0.027 | 5.32E-01 |
| Phospholipids in Medium HDL | 0.895 | 0.846 | 0.947 | 0.029 | 1.12E-04 | 0.871 | 0.802 | 0.947 | 0.042 | 1.18E-03 |
| Ratio of linoleic acid to total fatty acids | 0.909 | 0.858 | 0.962 | 0.029 | 1.06E-03 | 0.912 | 0.837 | 0.994 | 0.044 | 3.50E-02 |
| Total Lipids in Medium HDL | 0.875 | 0.826 | 0.927 | 0.029 | 5.62E-06 | 0.851 | 0.781 | 0.926 | 0.043 | 1.88E-04 |
| Phospholipids in HDL | 0.873 | 0.822 | 0.928 | 0.031 | 1.32E-05 | 0.850 | 0.777 | 0.930 | 0.046 | 4.00E-04 |
| Phospholipids to total lipids ratio in small HDL | 1.053 | 1.000 | 1.109 | 0.026 | 5.04E-02 | 1.041 | 0.965 | 1.123 | 0.039 | 2.99E-01 |
| Concentration of Medium HDL Particles | 0.851 | 0.799 | 0.905 | 0.032 | 3.68E-07 | 0.827 | 0.754 | 0.907 | 0.047 | 5.47E-05 |
| Free Cholesterol in Medium HDL | 0.845 | 0.793 | 0.901 | 0.033 | 2.41E-07 | 0.820 | 0.746 | 0.902 | 0.048 | 4.04E-05 |
| Apolipoprotein A1 | 0.847 | 0.797 | 0.900 | 0.031 | 7.75E-08 | 0.825 | 0.754 | 0.903 | 0.046 | 2.76E-05 |
| Total Lipids in HDL | 0.852 | 0.800 | 0.906 | 0.032 | 4.41E-07 | 0.827 | 0.755 | 0.907 | 0.047 | 5.67E-05 |
| Cholesterol in Medium HDL | 0.840 | 0.790 | 0.894 | 0.031 | 3.22E-08 | 0.811 | 0.740 | 0.888 | 0.047 | 7.21E-06 |
| Cholesteryl Esters in Medium HDL | 0.839 | 0.788 | 0.892 | 0.032 | 2.41E-08 | 0.808 | 0.737 | 0.886 | 0.047 | 5.83E-06 |
| Free Cholesterol in HDL | 0.826 | 0.770 | 0.886 | 0.036 | 1.06E-07 | 0.804 | 0.724 | 0.892 | 0.053 | 4.05E-05 |
| Ratio of omega-6 fatty acids to total fatty acids | 0.937 | 0.888 | 0.987 | 0.027 | 1.52E-02 | 0.932 | 0.861 | 1.009 | 0.040 | 8.25E-02 |
| Ratio of apolipoprotein B to apolipoprotein A1 | 1.006 | 0.952 | 1.064 | 0.029 | 8.21E-01 | 1.052 | 0.969 | 1.141 | 0.042 | 2.28E-01 |
| Concentration of HDL Particles | 0.833 | 0.785 | 0.885 | 0.031 | 2.67E-09 | 0.818 | 0.748 | 0.894 | 0.046 | 1.03E-05 |

*Fully adjusted for sex, age, race, Townsend Deprivation Index, household income, BMI, smoking, alcohol, and lipid-lowering medication.

GCIPLT = ganglion cell-inner plexus layer thickness; T2DM = type 2 diabetes mellitus; HDL = high-density lipoprotein; HR = hazard ratio; CI = confidence interval; SE = standard error.

#### Supplementary Table S13

**Associations of GCIPLT metabolomic signature and incident dementia.**

| **Metabolic Biomarkers** | **Training set** | | | | | **Validation set** | | | | |
| --- | --- | --- | --- | --- | --- | --- | --- | --- | --- | --- |
|  | **HR** | **95% CI** | | **SE** | **P-value*** | **HR** | **95% CI** | | **SE** | **P-value*** |
| Ratio of saturated fatty acids to total fatty acids | 1.026 | 0.984 | 1.070 | 0.021 | 2.29E-01 | 1.037 | 0.991 | 1.084 | 0.023 | 1.13E-01 |
| Phospholipids in Medium HDL | 0.905 | 0.851 | 0.962 | 0.031 | 1.46E-03 | 0.884 | 0.809 | 0.966 | 0.045 | 6.21E-03 |
| Ratio of linoleic acid to total fatty acids | 0.925 | 0.867 | 0.987 | 0.033 | 1.81E-02 | 0.902 | 0.823 | 0.989 | 0.047 | 2.83E-02 |
| Total Lipids in Medium HDL | 0.905 | 0.851 | 0.962 | 0.031 | 1.39E-03 | 0.887 | 0.812 | 0.968 | 0.045 | 7.46E-03 |
| Phospholipids in HDL | 0.924 | 0.867 | 0.984 | 0.032 | 1.46E-02 | 0.894 | 0.816 | 0.980 | 0.047 | 1.69E-02 |
| Phospholipids to total lipids ratio in small HDL | 0.972 | 0.916 | 1.031 | 0.030 | 3.39E-01 | 0.926 | 0.851 | 1.008 | 0.043 | 7.51E-02 |
| Concentration of Medium HDL Particles | 0.909 | 0.853 | 0.968 | 0.032 | 3.04E-03 | 0.887 | 0.810 | 0.972 | 0.047 | 1.03E-02 |
| Free Cholesterol in Medium HDL | 0.918 | 0.861 | 0.979 | 0.033 | 8.98E-03 | 0.893 | 0.814 | 0.980 | 0.047 | 1.65E-02 |
| Apolipoprotein A1 | 0.909 | 0.854 | 0.968 | 0.032 | 2.75E-03 | 0.898 | 0.821 | 0.982 | 0.046 | 1.87E-02 |
| Total Lipids in HDL | 0.926 | 0.869 | 0.987 | 0.033 | 1.86E-02 | 0.899 | 0.820 | 0.986 | 0.047 | 2.41E-02 |
| Cholesterol in Medium HDL | 0.916 | 0.861 | 0.976 | 0.032 | 6.27E-03 | 0.892 | 0.815 | 0.976 | 0.046 | 1.30E-02 |
| Cholesteryl Esters in Medium HDL | 0.918 | 0.862 | 0.977 | 0.032 | 6.89E-03 | 0.892 | 0.816 | 0.977 | 0.046 | 1.33E-02 |
| Free Cholesterol in HDL | 0.951 | 0.891 | 1.016 | 0.034 | 1.37E-01 | 0.919 | 0.835 | 1.011 | 0.049 | 8.34E-02 |
| Ratio of omega-6 fatty acids to total fatty acids | 0.974 | 0.915 | 1.037 | 0.032 | 4.12E-01 | 0.933 | 0.853 | 1.021 | 0.046 | 1.34E-01 |
| Ratio of apolipoprotein B to apolipoprotein A1 | 1.038 | 0.977 | 1.104 | 0.031 | 2.30E-01 | 1.091 | 1.000 | 1.190 | 0.044 | 4.93E-02 |
| Concentration of HDL Particles | 0.900 | 0.847 | 0.957 | 0.031 | 8.17E-04 | 0.904 | 0.828 | 0.988 | 0.045 | 2.52E-02 |

*Fully adjusted for sex, age, race, Townsend Deprivation Index, household income, BMI, smoking, alcohol, and lipid-lowering medication.

GCIPLT = ganglion cell-inner plexus layer thickness; T2DM = type 2 diabetes mellitus; HDL = high-density lipoprotein; HR = hazard ratio; CI = confidence interval; SE = standard error.

#### Supplementary Table S14

**Associations of GCIPLT metabolomic signature and incident ischemic stroke.**

| **Metabolic Biomarkers** | **Training set** | | | | | **Validation set** | | | | |
| --- | --- | --- | --- | --- | --- | --- | --- | --- | --- | --- |
|  | **HR** | **95% CI** | | **SE** | **P-value*** | **HR** | **95% CI** | | **SE** | **P-value*** |
| Ratio of saturated fatty acids to total fatty acids | 1.027 | 0.996 | 1.060 | 0.016 | 9.25E-02 | 1.029 | 0.984 | 1.076 | 0.023 | 2.16E-01 |
| Phospholipids in Medium HDL | 0.928 | 0.881 | 0.979 | 0.027 | 5.73E-03 | 0.917 | 0.849 | 0.990 | 0.039 | 2.63E-02 |
| Ratio of linoleic acid to total fatty acids | 0.907 | 0.858 | 0.960 | 0.028 | 6.51E-04 | 0.888 | 0.820 | 0.963 | 0.041 | 3.91E-03 |
| Total Lipids in Medium HDL | 0.921 | 0.873 | 0.971 | 0.027 | 2.26E-03 | 0.904 | 0.837 | 0.977 | 0.039 | 1.07E-02 |
| Phospholipids in HDL | 0.931 | 0.881 | 0.984 | 0.028 | 1.19E-02 | 0.908 | 0.838 | 0.985 | 0.041 | 1.94E-02 |
| Phospholipids to total lipids ratio in small HDL | 0.984 | 0.936 | 1.036 | 0.026 | 5.45E-01 | 0.999 | 0.929 | 1.076 | 0.037 | 9.86E-01 |
| Concentration of Medium HDL Particles | 0.915 | 0.867 | 0.967 | 0.028 | 1.61E-03 | 0.894 | 0.825 | 0.969 | 0.041 | 6.19E-03 |
| Free Cholesterol in Medium HDL | 0.926 | 0.876 | 0.979 | 0.028 | 6.82E-03 | 0.899 | 0.828 | 0.975 | 0.042 | 1.07E-02 |
| Apolipoprotein A1 | 0.922 | 0.873 | 0.973 | 0.028 | 3.27E-03 | 0.895 | 0.827 | 0.969 | 0.040 | 6.18E-03 |
| Total Lipids in HDL | 0.927 | 0.876 | 0.980 | 0.028 | 7.42E-03 | 0.899 | 0.828 | 0.975 | 0.042 | 1.02E-02 |
| Cholesterol in Medium HDL | 0.906 | 0.858 | 0.957 | 0.028 | 3.86E-04 | 0.883 | 0.815 | 0.956 | 0.041 | 2.19E-03 |
| Cholesteryl Esters in Medium HDL | 0.902 | 0.854 | 0.952 | 0.028 | 2.03E-04 | 0.879 | 0.812 | 0.952 | 0.041 | 1.57E-03 |
| Free Cholesterol in HDL | 0.947 | 0.893 | 1.004 | 0.030 | 6.77E-02 | 0.906 | 0.831 | 0.988 | 0.044 | 2.51E-02 |
| Ratio of omega-6 fatty acids to total fatty acids | 0.911 | 0.863 | 0.961 | 0.027 | 6.00E-04 | 0.893 | 0.827 | 0.965 | 0.040 | 4.29E-03 |
| Ratio of apolipoprotein B to apolipoprotein A1 | 1.100 | 1.044 | 1.159 | 0.027 | 3.75E-04 | 1.097 | 1.016 | 1.183 | 0.039 | 1.72E-02 |
| Concentration of HDL Particles | 0.918 | 0.871 | 0.968 | 0.027 | 1.66E-03 | 0.894 | 0.827 | 0.967 | 0.040 | 4.93E-03 |

*Fully adjusted for sex, age, race, Townsend Deprivation Index, household income, BMI, smoking, alcohol, and lipid-lowering medication.

GCIPLT = ganglion cell-inner plexus layer thickness; T2DM = type 2 diabetes mellitus; HDL = high-density lipoprotein; HR = hazard ratio; CI = confidence interval; SE = standard error.

#### Supplementary Table S15

**Associations of GCIPLT metabolomic signature and incident heart failure.**

| **Metabolic Biomarkers** | **Training set** | | | | | **Validation set** | | | | |
| --- | --- | --- | --- | --- | --- | --- | --- | --- | --- | --- |
|  | **HR** | **95% CI** | | **SE** | **P-value*** | **HR** | **95% CI** | | **SE** | **P-value*** |
| Ratio of saturated fatty acids to total fatty acids | 1.022 | 0.994 | 1.050 | 0.014 | 1.24E-01 | 1.002 | 0.956 | 1.051 | 0.024 | 9.23E-01 |
| Phospholipids in Medium HDL | 0.895 | 0.858 | 0.933 | 0.021 | 2.23E-07 | 0.900 | 0.848 | 0.956 | 0.031 | 6.63E-04 |
| Ratio of linoleic acid to total fatty acids | 0.958 | 0.918 | 1.000 | 0.022 | 5.18E-02 | 0.936 | 0.879 | 0.996 | 0.032 | 3.63E-02 |
| Total Lipids in Medium HDL | 0.891 | 0.854 | 0.929 | 0.021 | 7.51E-08 | 0.896 | 0.843 | 0.952 | 0.031 | 3.74E-04 |
| Phospholipids in HDL | 0.912 | 0.872 | 0.953 | 0.023 | 4.31E-05 | 0.912 | 0.856 | 0.972 | 0.033 | 4.69E-03 |
| Phospholipids to total lipids ratio in small HDL | 1.051 | 1.009 | 1.093 | 0.020 | 1.56E-02 | 1.062 | 1.003 | 1.124 | 0.029 | 4.05E-02 |
| Concentration of Medium HDL Particles | 0.891 | 0.853 | 0.932 | 0.023 | 3.29E-07 | 0.896 | 0.841 | 0.955 | 0.032 | 7.31E-04 |
| Free Cholesterol in Medium HDL | 0.897 | 0.857 | 0.938 | 0.023 | 2.09E-06 | 0.896 | 0.839 | 0.956 | 0.033 | 9.25E-04 |
| Apolipoprotein A1 | 0.887 | 0.849 | 0.926 | 0.022 | 5.64E-08 | 0.887 | 0.833 | 0.945 | 0.032 | 1.88E-04 |
| Total Lipids in HDL | 0.909 | 0.869 | 0.950 | 0.023 | 2.54E-05 | 0.907 | 0.851 | 0.968 | 0.033 | 3.14E-03 |
| Cholesterol in Medium HDL | 0.893 | 0.855 | 0.933 | 0.022 | 3.59E-07 | 0.898 | 0.843 | 0.956 | 0.032 | 8.23E-04 |
| Cholesteryl Esters in Medium HDL | 0.894 | 0.856 | 0.933 | 0.022 | 3.99E-07 | 0.900 | 0.845 | 0.958 | 0.032 | 1.01E-03 |
| Free Cholesterol in HDL | 0.931 | 0.888 | 0.977 | 0.024 | 3.46E-03 | 0.921 | 0.860 | 0.987 | 0.035 | 2.02E-02 |
| Ratio of omega-6 fatty acids to total fatty acids | 1.017 | 0.982 | 1.054 | 0.018 | 3.39E-01 | 1.005 | 0.956 | 1.058 | 0.026 | 8.37E-01 |
| Ratio of apolipoprotein B to apolipoprotein A1 | 0.986 | 0.945 | 1.029 | 0.022 | 5.11E-01 | 0.957 | 0.900 | 1.018 | 0.031 | 1.67E-01 |
| Concentration of HDL Particles | 0.857 | 0.821 | 0.895 | 0.022 | 1.79E-12 | 0.858 | 0.806 | 0.912 | 0.032 | 1.11E-06 |

*Fully adjusted for sex, age, race, Townsend Deprivation Index, household income, BMI, smoking, alcohol, and lipid-lowering medication.

GCIPLT = ganglion cell-inner plexus layer thickness; T2DM = type 2 diabetes mellitus; HDL = high-density lipoprotein; HR = hazard ratio; CI = confidence interval; SE = standard error.

#### Supplementary Table S16

**Associations of GCIPLT metabolomic signature and incident myocardial infarction.**

| **Metabolic Biomarkers** | **Training set** | | | | | **Validation set** | | | | |
| --- | --- | --- | --- | --- | --- | --- | --- | --- | --- | --- |
|  | **HR** | **95% CI** | | **SE** | **P-value*** | **HR** | **95% CI** | | **SE** | **P-value*** |
| Ratio of saturated fatty acids to total fatty acids | 1.008 | 0.976 | 1.041 | 0.016 | 6.23E-01 | 1.023 | 0.991 | 1.055 | 0.016 | 1.61E-01 |
| Phospholipids in Medium HDL | 0.800 | 0.768 | 0.832 | 0.020 | 4.46E-28 | 0.813 | 0.767 | 0.861 | 0.029 | 1.78E-12 |
| Ratio of linoleic acid to total fatty acids | 0.919 | 0.882 | 0.957 | 0.021 | 4.56E-05 | 0.910 | 0.858 | 0.965 | 0.030 | 1.66E-03 |
| Total Lipids in Medium HDL | 0.785 | 0.754 | 0.817 | 0.020 | 1.26E-32 | 0.795 | 0.750 | 0.842 | 0.029 | 6.45E-15 |
| Phospholipids in HDL | 0.783 | 0.751 | 0.817 | 0.022 | 8.96E-30 | 0.791 | 0.744 | 0.840 | 0.031 | 4.64E-14 |
| Phospholipids to total lipids ratio in small HDL | 0.948 | 0.913 | 0.985 | 0.019 | 5.93E-03 | 0.975 | 0.923 | 1.029 | 0.028 | 3.57E-01 |
| Concentration of Medium HDL Particles | 0.767 | 0.735 | 0.801 | 0.022 | 1.05E-33 | 0.779 | 0.732 | 0.828 | 0.032 | 2.84E-15 |
| Free Cholesterol in Medium HDL | 0.769 | 0.736 | 0.803 | 0.022 | 6.73E-32 | 0.775 | 0.727 | 0.825 | 0.032 | 3.42E-15 |
| Apolipoprotein A1 | 0.774 | 0.743 | 0.807 | 0.021 | 5.75E-34 | 0.778 | 0.733 | 0.826 | 0.031 | 2.46E-16 |
| Total Lipids in HDL | 0.772 | 0.739 | 0.805 | 0.022 | 6.33E-33 | 0.775 | 0.729 | 0.824 | 0.031 | 4.68E-16 |
| Cholesterol in Medium HDL | 0.757 | 0.726 | 0.789 | 0.021 | 1.11E-38 | 0.765 | 0.720 | 0.813 | 0.031 | 6.43E-18 |
| Cholesteryl Esters in Medium HDL | 0.754 | 0.723 | 0.787 | 0.021 | 0.00E+00 | 0.764 | 0.719 | 0.812 | 0.031 | 4.05E-18 |
| Free Cholesterol in HDL | 0.773 | 0.738 | 0.811 | 0.024 | 1.79E-26 | 0.771 | 0.720 | 0.826 | 0.035 | 1.11E-13 |
| Ratio of omega-6 fatty acids to total fatty acids | 0.946 | 0.911 | 0.983 | 0.019 | 4.12E-03 | 0.956 | 0.906 | 1.010 | 0.028 | 1.09E-01 |
| Ratio of apolipoprotein B to apolipoprotein A1 | 1.171 | 1.126 | 1.218 | 0.020 | 2.36E-15 | 1.132 | 1.070 | 1.198 | 0.029 | 1.74E-05 |
| Concentration of HDL Particles | 0.779 | 0.747 | 0.811 | 0.021 | 3.08E-33 | 0.781 | 0.736 | 0.828 | 0.030 | 2.74E-16 |

*Fully adjusted for sex, age, race, Townsend Deprivation Index, household income, BMI, smoking, alcohol, and lipid-lowering medication.

GCIPLT = ganglion cell-inner plexus layer thickness; T2DM = type 2 diabetes mellitus; HDL = high-density lipoprotein; HR = hazard ratio; CI = confidence interval; SE = standard error.

#### Supplementary Table S17

**Associations of GCIPLT metabolomic signature and all-cause mortality.**

| **Metabolic Biomarkers** | **Training set** | | | | | **Validation set** | | | | |
| --- | --- | --- | --- | --- | --- | --- | --- | --- | --- | --- |
|  | **HR** | **95% CI** | | **SE** | **P-value*** | **HR** | **95% CI** | | **SE** | **P-value*** |
| Ratio of saturated fatty acids to total fatty acids | 1.037 | 1.025 | 1.050 | 0.006 | 6.03E-09 | 1.039 | 1.020 | 1.058 | 0.009 | 4.89E-05 |
| Phospholipids in Medium HDL | 0.950 | 0.926 | 0.976 | 0.014 | 1.69E-04 | 0.939 | 0.904 | 0.976 | 0.020 | 1.40E-03 |
| Ratio of linoleic acid to total fatty acids | 0.872 | 0.848 | 0.897 | 0.014 | 9.95E-22 | 0.862 | 0.828 | 0.898 | 0.021 | 7.38E-13 |
| Total Lipids in Medium HDL | 0.939 | 0.915 | 0.965 | 0.014 | 4.65E-06 | 0.928 | 0.893 | 0.965 | 0.020 | 1.55E-04 |
| Phospholipids in HDL | 0.967 | 0.940 | 0.994 | 0.014 | 1.66E-02 | 0.952 | 0.914 | 0.991 | 0.021 | 1.72E-02 |
| Phospholipids to total lipids ratio in small HDL | 1.115 | 1.087 | 1.143 | 0.013 | 4.07E-17 | 1.117 | 1.077 | 1.159 | 0.019 | 2.93E-09 |
| Concentration of Medium HDL Particles | 0.942 | 0.916 | 0.968 | 0.014 | 1.85E-05 | 0.927 | 0.890 | 0.965 | 0.020 | 2.06E-04 |
| Free Cholesterol in Medium HDL | 0.947 | 0.921 | 0.974 | 0.014 | 1.42E-04 | 0.931 | 0.894 | 0.970 | 0.021 | 6.25E-04 |
| Apolipoprotein A1 | 0.933 | 0.908 | 0.959 | 0.014 | 7.40E-07 | 0.917 | 0.881 | 0.954 | 0.020 | 2.01E-05 |
| Total Lipids in HDL | 0.956 | 0.929 | 0.983 | 0.014 | 1.60E-03 | 0.940 | 0.903 | 0.979 | 0.021 | 3.15E-03 |
| Cholesterol in Medium HDL | 0.930 | 0.905 | 0.956 | 0.014 | 2.18E-07 | 0.918 | 0.882 | 0.955 | 0.020 | 2.68E-05 |
| Cholesteryl Esters in Medium HDL | 0.928 | 0.903 | 0.954 | 0.014 | 1.07E-07 | 0.917 | 0.881 | 0.954 | 0.020 | 2.04E-05 |
| Free Cholesterol in HDL | 0.979 | 0.951 | 1.008 | 0.015 | 1.54E-01 | 0.959 | 0.919 | 1.001 | 0.022 | 5.75E-02 |
| Ratio of omega-6 fatty acids to total fatty acids | 0.917 | 0.893 | 0.942 | 0.014 | 3.39E-10 | 0.923 | 0.888 | 0.960 | 0.020 | 7.16E-05 |
| Ratio of apolipoprotein B to apolipoprotein A1 | 0.956 | 0.931 | 0.982 | 0.014 | 1.06E-03 | 0.946 | 0.909 | 0.983 | 0.020 | 4.95E-03 |
| Concentration of HDL Particles | 0.894 | 0.870 | 0.918 | 0.014 | 5.06E-16 | 0.877 | 0.843 | 0.912 | 0.020 | 7.85E-11 |

*Fully adjusted for sex, age, race, Townsend Deprivation Index, household income, BMI, smoking, alcohol, and lipid-lowering medication.

GCIPLT = ganglion cell-inner plexus layer thickness; T2DM = type 2 diabetes mellitus; HDL = high-density lipoprotein; HR = hazard ratio; CI = confidence interval; SE = standard error.

#### Supplementary Table S18

**Associations of GCIPLT metabolomic signature and CVD mortality.**

| **Metabolic Biomarkers** | **Training set** | | | | | **Validation set** | | | | |
| --- | --- | --- | --- | --- | --- | --- | --- | --- | --- | --- |
|  | **HR** | **95% CI** | | **SE** | **P-value*** | **HR** | **95% CI** | | **SE** | **P-value*** |
| Ratio of saturated fatty acids to total fatty acids | 1.029 | 0.998 | 1.061 | 0.016 | 6.75E-02 | 1.031 | 0.990 | 1.074 | 0.021 | 1.42E-01 |
| Phospholipids in Medium HDL | 0.871 | 0.825 | 0.919 | 0.028 | 5.20E-07 | 0.870 | 0.804 | 0.942 | 0.040 | 5.78E-04 |
| Ratio of linoleic acid to total fatty acids | 0.860 | 0.813 | 0.909 | 0.028 | 9.68E-08 | 0.828 | 0.764 | 0.898 | 0.041 | 5.09E-06 |
| Total Lipids in Medium HDL | 0.857 | 0.812 | 0.905 | 0.028 | 2.75E-08 | 0.858 | 0.793 | 0.929 | 0.040 | 1.55E-04 |
| Phospholipids in HDL | 0.879 | 0.830 | 0.930 | 0.029 | 9.21E-06 | 0.871 | 0.802 | 0.947 | 0.043 | 1.22E-03 |
| Phospholipids to total lipids ratio in small HDL | 1.059 | 1.006 | 1.115 | 0.026 | 2.86E-02 | 1.051 | 0.975 | 1.134 | 0.038 | 1.91E-01 |
| Concentration of Medium HDL Particles | 0.852 | 0.805 | 0.903 | 0.029 | 5.12E-08 | 0.849 | 0.781 | 0.924 | 0.043 | 1.48E-04 |
| Free Cholesterol in Medium HDL | 0.856 | 0.807 | 0.908 | 0.030 | 2.09E-07 | 0.850 | 0.780 | 0.927 | 0.044 | 2.20E-04 |
| Apolipoprotein A1 | 0.849 | 0.803 | 0.898 | 0.029 | 1.20E-08 | 0.842 | 0.776 | 0.914 | 0.042 | 3.91E-05 |
| Total Lipids in HDL | 0.869 | 0.820 | 0.920 | 0.029 | 1.72E-06 | 0.861 | 0.791 | 0.936 | 0.043 | 4.79E-04 |
| Cholesterol in Medium HDL | 0.838 | 0.792 | 0.887 | 0.029 | 9.67E-10 | 0.844 | 0.777 | 0.917 | 0.042 | 6.14E-05 |
| Cholesteryl Esters in Medium HDL | 0.836 | 0.790 | 0.885 | 0.029 | 5.45E-10 | 0.845 | 0.778 | 0.917 | 0.042 | 6.21E-05 |
| Free Cholesterol in HDL | 0.891 | 0.837 | 0.948 | 0.032 | 2.72E-04 | 0.872 | 0.796 | 0.956 | 0.047 | 3.52E-03 |
| Ratio of omega-6 fatty acids to total fatty acids | 0.897 | 0.851 | 0.945 | 0.027 | 4.81E-05 | 0.904 | 0.837 | 0.976 | 0.039 | 1.01E-02 |
| Ratio of apolipoprotein B to apolipoprotein A1 | 1.019 | 0.965 | 1.076 | 0.028 | 5.06E-01 | 0.980 | 0.904 | 1.062 | 0.041 | 6.22E-01 |
| Concentration of HDL Particles | 0.815 | 0.771 | 0.862 | 0.028 | 6.06E-13 | 0.812 | 0.748 | 0.881 | 0.042 | 5.40E-07 |

*Fully adjusted for sex, age, race, Townsend Deprivation Index, household income, BMI, smoking, alcohol, and lipid-lowering medication.

GCIPLT = ganglion cell-inner plexus layer thickness; T2DM = type 2 diabetes mellitus; HDL = high-density lipoprotein; HR = hazard ratio; CI = confidence interval; SE = standard error.

#### Supplementary Table S19

**Associations of GCIPLT metabolomic signature and cancer mortality.**

| **Metabolic Biomarkers** | **Training set** | | | | | **Validation set** | | | | |
| --- | --- | --- | --- | --- | --- | --- | --- | --- | --- | --- |
|  | **HR** | **95% CI** | | **SE** | **P-value*** | **HR** | **95% CI** | | **SE** | **P-value*** |
| Ratio of saturated fatty acids to total fatty acids | 1.033 | 1.013 | 1.053 | 0.010 | 8.94E-04 | 1.033 | 1.003 | 1.065 | 0.015 | 3.35E-02 |
| Phospholipids in Medium HDL | 0.972 | 0.936 | 1.008 | 0.019 | 1.28E-01 | 0.950 | 0.901 | 1.003 | 0.027 | 6.30E-02 |
| Ratio of linoleic acid to total fatty acids | 0.890 | 0.855 | 0.926 | 0.020 | 9.74E-09 | 0.894 | 0.844 | 0.947 | 0.029 | 1.36E-04 |
| Total Lipids in Medium HDL | 0.959 | 0.924 | 0.996 | 0.019 | 2.88E-02 | 0.937 | 0.888 | 0.989 | 0.028 | 1.91E-02 |
| Phospholipids in HDL | 0.973 | 0.936 | 1.012 | 0.020 | 1.70E-01 | 0.953 | 0.901 | 1.009 | 0.029 | 9.61E-02 |
| Phospholipids to total lipids ratio in small HDL | 1.104 | 1.065 | 1.144 | 0.018 | 5.77E-08 | 1.107 | 1.052 | 1.165 | 0.026 | 1.01E-04 |
| Concentration of Medium HDL Particles | 0.960 | 0.924 | 0.997 | 0.020 | 3.67E-02 | 0.938 | 0.887 | 0.991 | 0.028 | 2.35E-02 |
| Free Cholesterol in Medium HDL | 0.960 | 0.923 | 0.998 | 0.020 | 3.90E-02 | 0.938 | 0.886 | 0.993 | 0.029 | 2.69E-02 |
| Apolipoprotein A1 | 0.948 | 0.912 | 0.985 | 0.020 | 6.83E-03 | 0.925 | 0.874 | 0.978 | 0.029 | 5.91E-03 |
| Total Lipids in HDL | 0.959 | 0.922 | 0.998 | 0.020 | 3.80E-02 | 0.938 | 0.886 | 0.994 | 0.029 | 2.95E-02 |
| Cholesterol in Medium HDL | 0.946 | 0.911 | 0.983 | 0.020 | 4.74E-03 | 0.924 | 0.874 | 0.977 | 0.028 | 5.59E-03 |
| Cholesteryl Esters in Medium HDL | 0.945 | 0.910 | 0.982 | 0.020 | 4.07E-03 | 0.924 | 0.874 | 0.976 | 0.028 | 5.10E-03 |
| Free Cholesterol in HDL | 0.970 | 0.931 | 1.010 | 0.021 | 1.42E-01 | 0.951 | 0.896 | 1.010 | 0.030 | 1.01E-01 |
| Ratio of omega-6 fatty acids to total fatty acids | 0.925 | 0.890 | 0.962 | 0.020 | 9.00E-05 | 0.940 | 0.889 | 0.994 | 0.029 | 3.09E-02 |
| Ratio of apolipoprotein B to apolipoprotein A1 | 0.964 | 0.929 | 1.001 | 0.019 | 5.77E-02 | 0.964 | 0.914 | 1.018 | 0.028 | 1.89E-01 |
| Concentration of HDL Particles | 0.924 | 0.890 | 0.960 | 0.019 | 4.29E-05 | 0.898 | 0.850 | 0.948 | 0.028 | 1.19E-04 |

*Fully adjusted for sex, age, race, Townsend Deprivation Index, household income, BMI, smoking, alcohol, and lipid-lowering medication.

GCIPLT = ganglion cell-inner plexus layer thickness; T2DM = type 2 diabetes mellitus; HDL = high-density lipoprotein; HR = hazard ratio; CI = confidence interval; SE = standard error.

#### Supplementary Table S20

**Associations of GCIPLT metabolomic signature and other mortality.**

| **Metabolic Biomarkers** | **Training set** | | | | | **Validation set** | | | | |
| --- | --- | --- | --- | --- | --- | --- | --- | --- | --- | --- |
|  | **HR** | **95% CI** | | **SE** | **P-value*** | **HR** | **95% CI** | | **SE** | **P-value*** |
| Ratio of saturated fatty acids to total fatty acids | 1.050 | 1.031 | 1.070 | 0.010 | 3.14E-07 | 1.052 | 1.024 | 1.081 | 0.014 | 2.10E-04 |
| Phospholipids in Medium HDL | 0.991 | 0.940 | 1.045 | 0.027 | 7.34E-01 | 0.989 | 0.916 | 1.069 | 0.039 | 7.86E-01 |
| Ratio of linoleic acid to total fatty acids | 0.848 | 0.802 | 0.897 | 0.028 | 7.43E-09 | 0.832 | 0.767 | 0.901 | 0.041 | 7.64E-06 |
| Total Lipids in Medium HDL | 0.986 | 0.935 | 1.040 | 0.027 | 6.13E-01 | 0.983 | 0.910 | 1.063 | 0.040 | 6.69E-01 |
| Phospholipids in HDL | 1.044 | 0.988 | 1.103 | 0.028 | 1.27E-01 | 1.034 | 0.954 | 1.120 | 0.041 | 4.19E-01 |
| Phospholipids to total lipids ratio in small HDL | 1.193 | 1.134 | 1.254 | 0.026 | 6.59E-12 | 1.205 | 1.120 | 1.296 | 0.037 | 5.84E-07 |
| Concentration of Medium HDL Particles | 0.994 | 0.941 | 1.050 | 0.028 | 8.21E-01 | 0.982 | 0.907 | 1.064 | 0.041 | 6.58E-01 |
| Free Cholesterol in Medium HDL | 1.012 | 0.957 | 1.069 | 0.028 | 6.80E-01 | 0.998 | 0.920 | 1.082 | 0.041 | 9.57E-01 |
| Apolipoprotein A1 | 0.989 | 0.936 | 1.045 | 0.028 | 6.89E-01 | 0.980 | 0.905 | 1.061 | 0.041 | 6.12E-01 |
| Total Lipids in HDL | 1.039 | 0.983 | 1.099 | 0.028 | 1.78E-01 | 1.027 | 0.947 | 1.114 | 0.041 | 5.16E-01 |
| Cholesterol in Medium HDL | 0.991 | 0.938 | 1.047 | 0.028 | 7.52E-01 | 0.981 | 0.906 | 1.062 | 0.041 | 6.29E-01 |
| Cholesteryl Esters in Medium HDL | 0.988 | 0.936 | 1.044 | 0.028 | 6.72E-01 | 0.977 | 0.903 | 1.058 | 0.041 | 5.69E-01 |
| Free Cholesterol in HDL | 1.082 | 1.022 | 1.145 | 0.029 | 6.79E-03 | 1.058 | 0.973 | 1.150 | 0.043 | 1.85E-01 |
| Ratio of omega-6 fatty acids to total fatty acids | 0.919 | 0.871 | 0.971 | 0.028 | 2.43E-03 | 0.907 | 0.838 | 0.982 | 0.040 | 1.60E-02 |
| Ratio of apolipoprotein B to apolipoprotein A1 | 0.886 | 0.839 | 0.935 | 0.028 | 1.23E-05 | 0.880 | 0.813 | 0.952 | 0.040 | 1.55E-03 |
| Concentration of HDL Particles | 0.914 | 0.866 | 0.965 | 0.028 | 1.18E-03 | 0.903 | 0.834 | 0.978 | 0.040 | 1.18E-02 |

*Fully adjusted for sex, age, race, Townsend Deprivation Index, household income, BMI, smoking, alcohol, and lipid-lowering medication.

GCIPLT = ganglion cell-inner plexus layer thickness; T2DM = type 2 diabetes mellitus; HDL = high-density lipoprotein; HR = hazard ratio; CI = confidence interval; SE = standard error.
